# Evaluation of statistical methods used to meta-analyse results from interrupted time series studies: a simulation study

**DOI:** 10.1101/2022.10.17.22281160

**Authors:** Elizabeth Korevaar, Simon L Turner, Andrew B Forbes, Amalia Karahalios, Monica Taljaard, Joanne E McKenzie

## Abstract

**Background:** Interrupted time series (ITS) are often meta-analysed to inform public health and policy decisions but examination of the statistical methods for ITS analysis and meta-analysis in this context is limited.

**Methods:** We simulated meta-analyses of ITS studies with continuous outcome data, analysed the studies using segmented linear regression with two estimation methods [ordinary least squares (OLS) and restricted maximum likelihood (REML)], and meta-analysed the immediate level- and slope-change effect estimates using fixed-effect and (multiple) random-effects meta-analysis methods. Simulation design parameters included varying series length; magnitude of lag-1 autocorrelation; magnitude of level- and slope-changes; number of included studies; and, effect size heterogeneity.

**Results:** All meta-analysis methods yielded unbiased estimates of the interruption effects. All random effects meta-analysis methods yielded coverage close to the nominal level, irrespective of the ITS analysis method used and other design parameters. However, heterogeneity was frequently overestimated in scenarios where the ITS study standard errors were underestimated, which occurred for short series or when the ITS analysis method did not appropriately account for autocorrelation.

**Conclusions:** The performance of meta-analysis methods depends on the design and analysis of the included ITS studies. Although all random effects methods performed well in terms of coverage, irrespective of the ITS analysis method, we recommend the use of effect estimates calculated from ITS methods that adjust for autocorrelation when possible. Doing so will likely to lead to more accurate estimates of the heterogeneity variance.

## 1 Introduction

Healthcare policy decision making is often informed by systematic reviews examining the impact of policy or public health interventions, or the impact from exposures such as natural disasters (both referred to as ‘interruptions’ hereafter). These reviews may need to consider evidence beyond randomised trials, as it is not always possible to randomise interventions targeted at populations (e.g., when evaluating the impact of a media campaign broadcast to an entire country)^1, 2^. A quasi-experimental non-randomised design that is often used to evaluate the impact of interruptions targeted at populations is the interrupted time series (ITS) design^3-5^. This design is immune to common threats to internal validity compared with other non-randomised designs (e.g., uncontrolled before-after design), and as such, is often included in systematic reviews^6-8^. The results from multiple ITS studies within systematic reviews may be statistically combined using meta-analysis methods; the findings of which underpin review conclusions^9, 10^.

Before proceeding with meta-analysis of ITS studies, there are a range of issues for analysis of a *single* ITS study that require consideration. In a single ITS study, data are often collected continuously over time pre and post an interruption. Commonly the data are aggregated using summary statistics (such as means or proportions) over regular time intervals (e.g., weekly or monthly) for analysis^11^. A commonly fitted model structure is a segmented linear model^12-14^, which allows estimation of separate underlying time trends in the pre-interruption period and the post-interruption period. The estimated time trend in the pre-interruption period can be used to predict what would have occurred in the absence of the interruption, thus providing a counterfactual for comparison with what was observed, using the estimated post-interruption time trend. Several effect metrics can then be calculated to quantify the impact of the interruption; commonly these include an immediate level change, and a change in slope from pre-interruption period to post-interruption period^15^ (see Figure 1(A) for examples). Researchers aiming to include ITS in a meta-analysis may need to re-analyse the original data (which is often possible when data are presented in figures in primary publications^12, 16^) to calculate interruption effects using desired effect metrics, and appropriate statistical methods^14, 17^.

**Figure 1.**
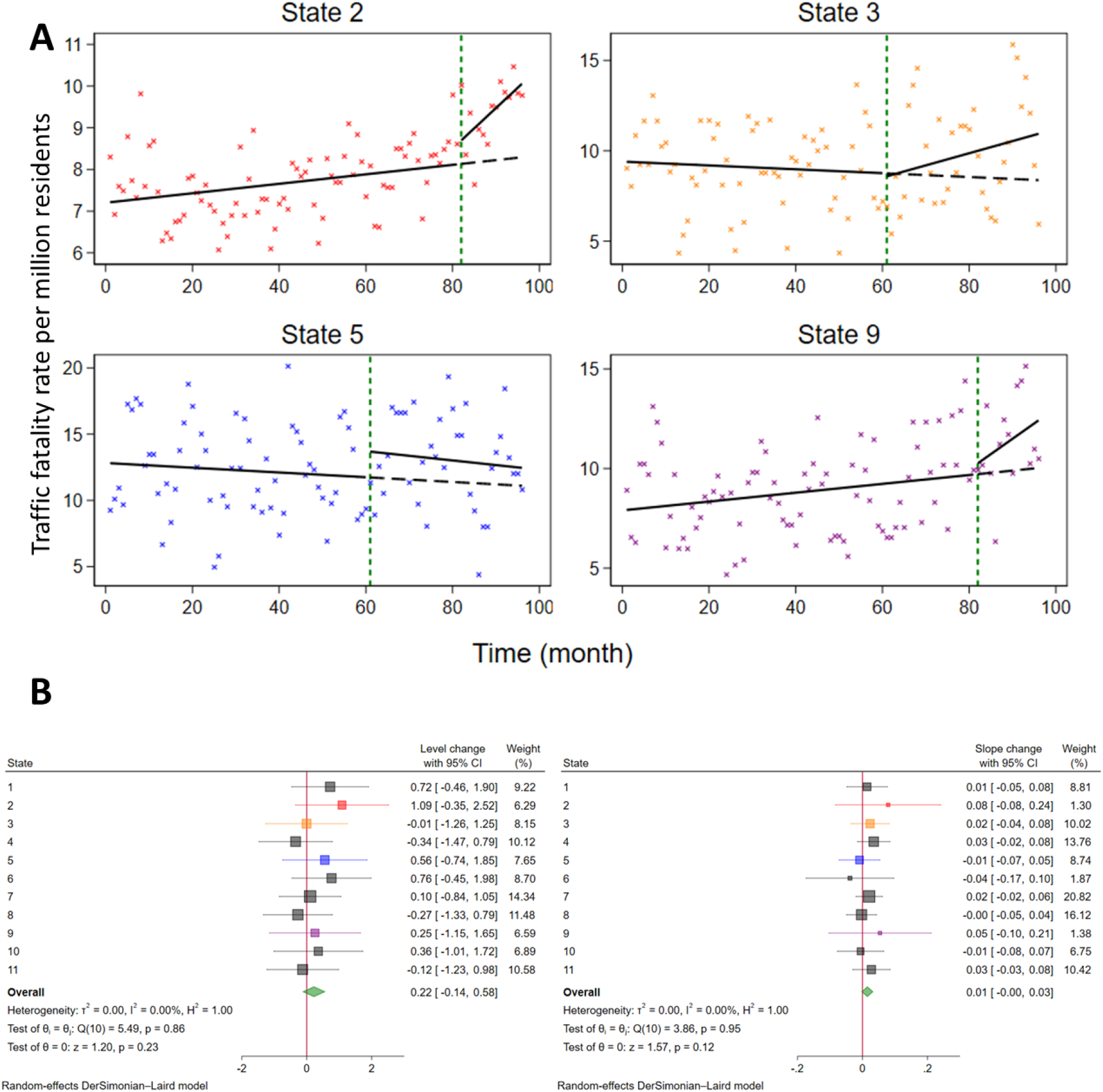
(A) Plots of interrupted time series (ITS) data examining the effect of state laws to legalise recreational cannabis sales on the Traffic fatality rate (per million residents)^38^. The crosses represent data points, the solid lines represent the pre- and post-interruption trend lines and the dashed line represents the counterfactual trend line. The green dashed line indicates the time of the interruption. The four states’ datapoints are coloured red (State 2), orange (State 3), blue (State 5) and purple (State 9) for matching with their respective level-change and slope-change effect estimates in B. See Appendix Figure S1 for an ITS graph annotated with the effect measures of interest and plots of all eleven states’ ITS data. (B) Forest plots depicting state-level and meta-analysis estimates of immediate level-change (left) and slope-change (right).

A range of statistical methods are available for estimating the regression parameters and effect estimates from a segmented linear model^12, 18, 19^. While Ordinary Least Squares (OLS) is often used, it fails to account for the potential correlation of consecutive time points (known as autocorrelation or serial correlation), which is a key characteristic of time series data^20, 21^. Failing to account for autocorrelation may lead to incorrect estimates of the standard errors of the regression parameters^22, 23^. Several methods that attempt to account for potential autocorrelation include, for example, generalised least squares methods (e.g., Prais-Winsten (PW)^24^), and Restricted Maximum Likelihood (REML)^25^. A numerical simulation study has provided insight on the performance of these statistical methods (and others) for analysing single ITS studies with continuous outcomes using segmented linear models^17^. The authors found that performance differed across the methods, but that REML was often preferable to the other methods; however, its performance was dependent on the length of the series and the underlying magnitude of autocorrelation.

Meta-analysis may be used to estimate a combined effect across ITS studies^26, 27^ (see Figure 1(B) for an example). Commonly a two-stage meta-analysis approach is used^8^, whereby the interruption effect estimates (e.g., level-change or slope-change) are calculated for each ITS study, and then statistically combined^27, 28^. The effects are commonly combined assuming the fixed (or common) effect model, or the random effects model^8, 29, 30^. The fixed-effect approach requires estimates of the interruption effect and its standard error only, while the random-effects approach additionally requires the estimation of the between-study variance ^10, 30^. Numerous between-study variance estimators exist (e.g., the DerSimonian and Laird (DL) and REML estimators)^31^ and, in addition, numerous methods are available for calculating the confidence interval of the combined effect (e.g., Wald-type (WT) and Hartung-Knapp / Sidik-Jonkman (HKSJ) methods)^32^.

The performance of the between-study variance estimators and the confidence interval methods have been reviewed and compared in numerical simulation studies and empirically using real-world data^31-33^. The DL estimator, commonly used to estimate the between-study variance, is well known to have suboptimal statistical properties in circumstances where there are few studies and the underlying statistical heterogeneity is large^21, 34^. The REML estimator has been proposed as an alternative because it has been shown to yield less biased estimates of the between-study variance compared to DL^12^. The WT method, a commonly used method for calculating confidence intervals, has been shown to yield less than nominal coverage levels when there are few studies or when the underlying between-study variance is large, or both^32^. The HKSJ method has been shown to yield wider confidence intervals than the WT, although may yield narrower confidence intervals when the number of included studies is small or true between-study variance is small^35^.

While ITS analysis methods have been evaluated at the individual study level^17, 22^, and the meta-analysis methods have been evaluated generally^31, 32, 36, 37^, neither has been evaluated in the context of multiple ITS studies. This context necessitates consideration of both individual study (re-)analysis and meta-analysis simultaneously. Hence, in this simulation study, we aimed to examine the performance of different meta-analysis methods to combine results from ITS studies, and how characteristics of the meta-analysis, ITS design, and ITS analysis methods, modify the performance. Specifically, we examined how the performance was altered when the magnitude of AR(1) autocorrelation, series length, degree of heterogeneity in the interruption effects and number of included ITS studies were varied. We limited the meta-analyses examined to those that included ITS studies with continuous outcomes, a fixed number of data points, an equal number of data points pre- and post-interruption, and the same pre-interruption level and slope. We did not consider scenarios or statistical methods that include control series.

## 2 Methods

This simulation study was designed according to the “ADEMP” structure proposed by Morris et al^39^. The background and **A**ims were described above, while in subsequent sections we outline the **D**ata generation mechanisms (Section 2.1 and 2.2.1), the **E**stimands (and their estimation procedures of interest, Section 2.2.2), **M**ethods and **P**erformance measures (Section 2.2.3-2.2.6).

### 2.1 Statistical models

#### 2.1.1 Statistical model for an ITS study

An ITS with a single interruption is commonly modelled using segmented linear regression as follows^1^:

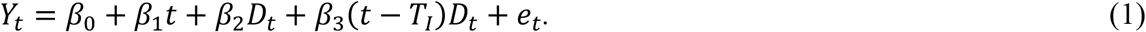

*Y*_*t*_ is a continuous outcome at time *t* (*t* = 1, …, *T*) and the interruption time is indicated by *T*_*I*_. *D*_*t*_ is an indicator variable that represents the post-interruption period (*D*_*t*_ = 1_(*t*≥*TI*_)). *β*_0_ represents the intercept in the pre-interruption period, *β*_1_ the pre-interruption slope, and *μ*_2_ and *μ*_3_ represent the interruption effects; respectively, change in level and change in slope. The error term, *e*_*t*_, is constructed from two components (*ρe*_*t*−1_ + *w*_*t*_). The first, *ρ* (−1 ≤ *ρ* ≤ 1), represents the degree of the correlation between the error at time *t* and the error of the previous time point (*t* − 1), and the second represents ‘white noise’, which is assumed to be normally distributed (*w*_*t*_∼*N*(0,1)). Here, the error term accommodates lag-1 (AR(1)) autocorrelation, but can be extended to accommodate longer lags.

#### 2.1.2 Estimation methods for ITS analysis

There are several estimation methods that can be used to estimate the parameters of the segmented linear regression model. Here, we focus on three estimation methods – ordinary least squares (OLS), Prais-Winsten (PW) and Restricted Maximum Likelihood (REML).

##### Ordinary least squares (OLS)

estimators, commonly used in practice^8, 12^, can be used to estimate the regression parameters and their standard errors^21^. A key assumption of OLS is that the model errors are uncorrelated between observations, which may be violated with time series data. In the presence of autocorrelation, estimates of the regression parameters will be unbiased, however, their standard errors may be biased. In the presence of (likely^22^) positive autocorrelation, they will be too small^18, 40^.

##### Prais-Winsten (PW)

a generalised least-squares approach, provides an extension of OLS that allows for lag-1 autocorrelation (AR(1))^41^. In brief, the estimation procedure involves first fitting the segmented linear regression model (Equation 1) using OLS, from which an estimate of autocorrelation is calculated from the residuals. The data are then transformed using the estimated autocorrelation, aiming to remove the autocorrelation from the errors. The regression parameters are then re-estimated using OLS. Further iteration of these steps may be required until the estimated autocorrelation converges^42^.

##### Restricted Maximum Likelihood (REML)

estimators can be used to estimate the regression parameters and their standard errors. REML is a form of maximum likelihood estimation in which the log-likelihood is partitioned into two terms. The first term, comprised of only variance components, is first maximised to obtain estimates of the error variance and correlation parameters, accounting for the appropriate degrees of freedom. The second term, comprised of both regression and error variance parameters, is then maximised using estimates from the first term. Maximum likelihood variance estimators do not appropriately account for the loss in degrees of freedom that result from estimating the regression parameters, which leads to negatively biased variance components for small samples^25^.

#### 2.3.1 Statistical models for meta-analysis

Meta-analysis may be used to estimate a combined effect from at least two ITS studies^9, 10^. Here, we focus on the two-stage meta-analysis approach. The two most common meta-analysis models include the fixed-effect (also known as common effect) and random-effects models.

In a fixed-effect meta-analysis model, it is assumed that the included ITS studies estimate a single true (common) interruption effect, and any variability in the observed effects is only due to sampling variability. The model can be specified by:

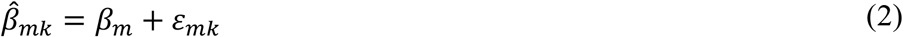

where *β*_*m*_ represents the underlying true interruption effect of the *m*^*th*^ regression parameter from Equation 1, and of interest here is *β*_2_ (immediate level-change) and *β*_3_ (slope-change); 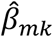is an estimate of the *m*^*th*^ regression parameter from the *k*^*th*^ ITS study (*k* = 1, …, *K* and *K* ≥ 2), and the error in estimating a particular ITS study *k*’s true effect from a sample of participants, assumed to be normally distributed, is represented by 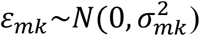.

In a random-effects meta-analysis model, it is assumed that the true interruption effects follow a (conventionally assumed normal) distribution, and the observed ITS study effects are a random sample from this distribution^10^. The random-effects model can be specified by:

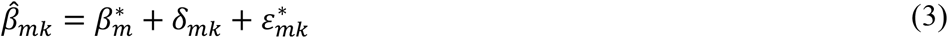

where 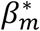 represents the mean of the distribution of true interruption effects (where, as above, *m* represents the regression parameter (and effect measure) of interest); *δ*_*mk*_ represents a random effect that allows a separate interruption effect in the *k*^*th*^ ITS study, where these effects are assumed to be normally distributed about the average interruption effect 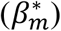, with between-study variance 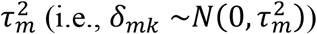; and within-study error 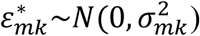.

#### 2.1.4 Estimation methods for meta-analysis

For a given effect measure, *m*, the meta-analytic effect is estimated as the weighted average of the *K* ITS study effect estimates. For a fixed-effect model, the estimator for the meta-analytic effect is 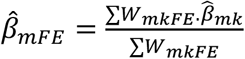(with a variance of 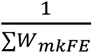), where the weight given to the *k*^*th*^ ITS study is simply the reciprocal of the within-study variance, 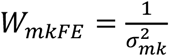. For a random-effects model, the same estimator is used, but the weights are modified to incorporate the additional source of between-study variation, 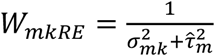. A common assumption is that the within-study variances are known, when in practice they are estimated from the observed study data. For large studies, this assumption is generally reasonable, however, for small studies, this can bias the model parameters^31^. Different between-study variance estimators are available^31^, as well as methods to calculate the confidence interval for the meta-analytic effect^32^. Here we consider two between-study variance estimators and confidence interval methods.

##### Between-study variance estimators

DerSimonian and Laird (DL) is a moment-based between-study variance estimator derived from Cochrane’s Q-statistic^43^, chosen for inclusion in this study as it is commonly used^8^ and is implemented as the default estimator in many software packages (e.g., RevMan^44^, *metan* in Stata^45^). The estimator is given by:

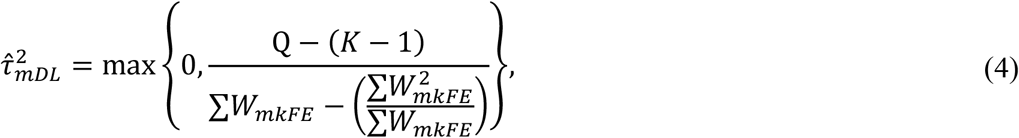

where the weights are from a fixed-effect meta-analysis model, and Q is calculated based on the fixed-effect meta-analysis estimate,

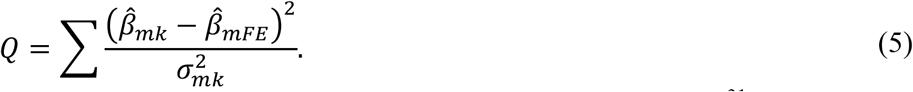

An alternative between-study variance estimator can be derived using REML^31^, chosen for inclusion in this study as it has been recommended as a preferable estimator^31, 46, 47^. The estimator is given by:

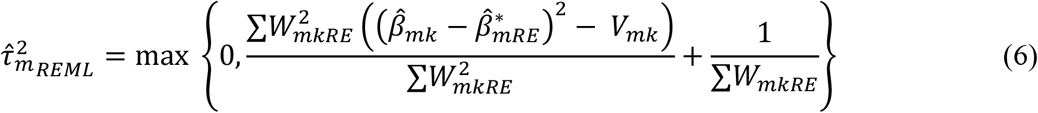

The estimate of the between-study variance is calculated through a process of iteration, whereby the initial value of 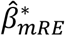 is the maximum likelihood estimate, from which an initial 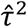 is computed, then 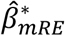 is updated and the process repeated until convergence. However, the algorithm can occasionally fail to converge.

##### Confidence interval calculation

A range of confidence interval methods for the meta-analytic (summary) estimate are available^32^. The two outlined here can be used with both the DL and REML between-study variance estimators.

The method chosen for inclusion in this study for its wide use in practice^8, 34, 43^, the Wald-type normal distribution (WT) confidence interval^48^, is calculated as:

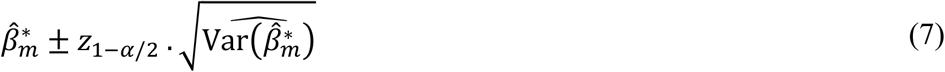

where *z*_1−*α*/2_ is the 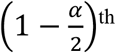 quantile of the standard normal distribution (note that 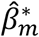 is replaced with 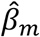 for a fixed-effect meta-analysis). This method assumes 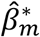 is normally distributed, despite the within-study and between-study variances not being known and estimated^32, 49^.

Hartung and Knapp, and independently, Sidik and Jonkman, (henceforth referred to as the Hartung-Knapp / Sidik-Jonkman (HKSJ))^50^ derived an alternative confidence interval method in an attempt to deal with meta-analyses with few studies, selected here for its better performance when there are few included studies^33, 51-53^. Rather than assuming normality of 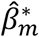, the method assumes the t-distribution (with K-1 degrees of freedom), and includes a small sample standard error adjustment, q, and is calculated as:

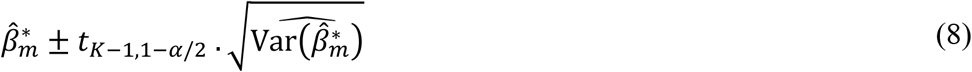

where

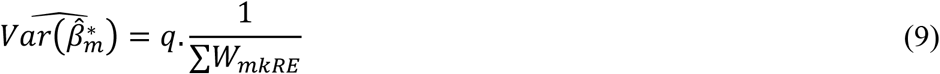

and

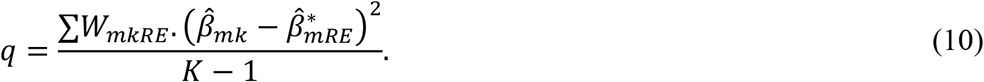

### 2.2 Simulation methods

Before providing full details, we briefly outline our simulation approach. We generated ITS studies, analysed these using segmented linear regression using two estimation methods (Section 2.1.2) and meta-analysed the resulting level-change and slope-change effect estimates using a fixed-effect and multiple random-effects meta-analysis methods (Section 2.1.4). The ITS studies and meta-analyses were generated using a range of design parameters (e.g., varying levels of autocorrelation, varying number of studies per meta-analysis). These design parameters were combined using a fully factorial approach (1620 simulation scenarios), with 1000 replicate meta-analyses generated per scenario. Various criteria (e.g., bias, 95% confidence interval coverage, Appendix 2) were used to assess the performance of the meta-analysis methods.

#### 2.1.1 Data generation

The design parameters, which were informed by reviews of ITS studies^12, 17, 22^ and of meta-analyses of ITS studies^8^, are provided in Table 1. For each combination of these parameters, the ITS studies were generated by randomly sampling from the model in Equation 1, with an equal number of points pre- and post-interruption. Models were constructed assuming level-changes (*β*_2_) of 0 and 1, and slope-changes (*β*_3_) of 0 and 0.1. We limited the number of level- and slope-changes investigated based on findings of a simulation study that^17^ demonstrate that the magnitude of these parameters did not impact the performance (across a range of metrics, excluding power) for the ITS estimation methods considered (Section 2.3.3). Lag-1 autocorrelation was varied between fixed values of *ρ* = 0 and 0.6 in increments of 0.2, and values drawn from a distribution *ρ*^*^∼*N*(0.4,0.15^2^). Sampling autocorrelation from a distribution aimed to reflect the likely scenario that autocorrelation will vary across the ITS studies. Three different series lengths were generated (12, 48 and 100 datapoints).

**Table 1.**
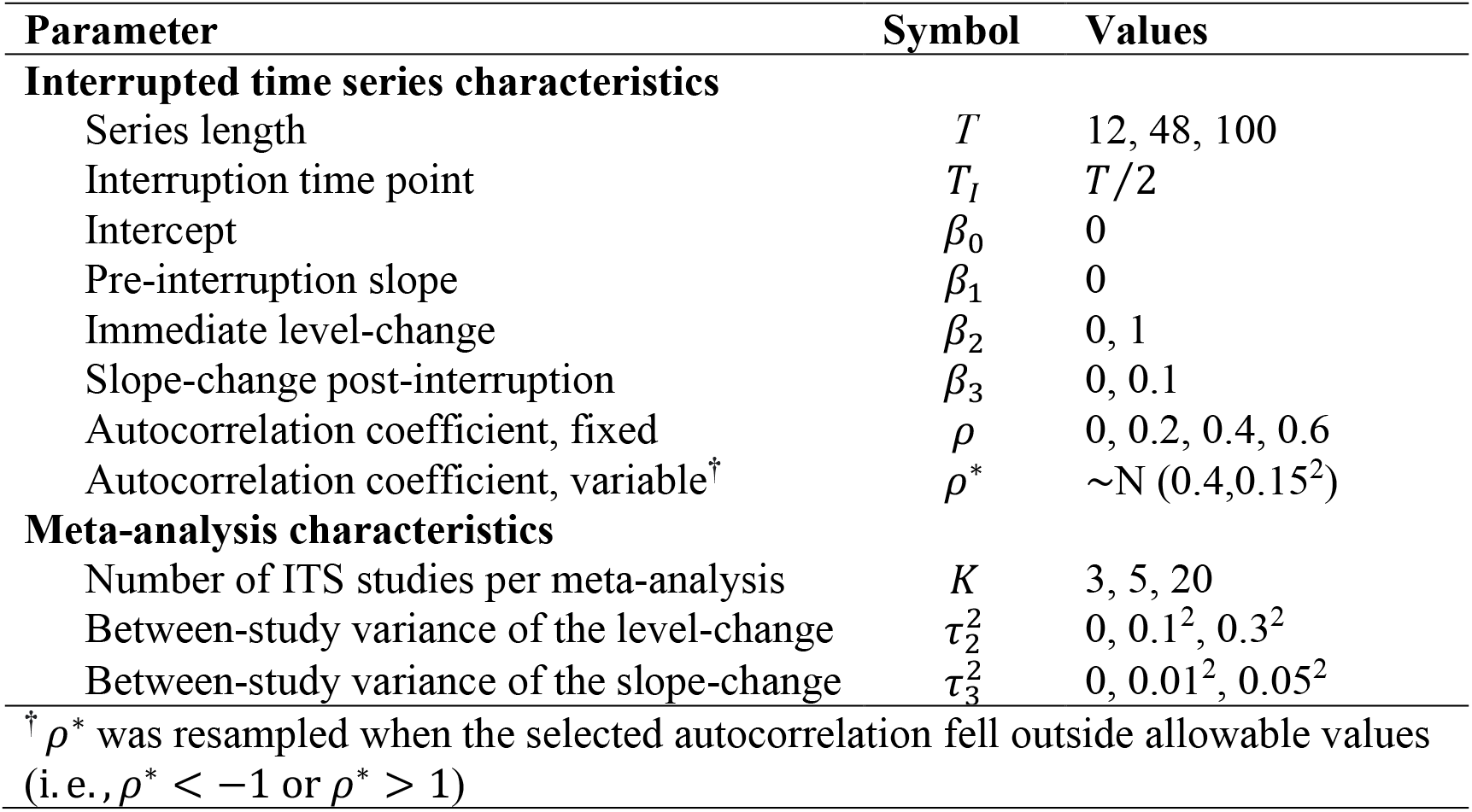
Design parameters used in the simulation study.

| Parameter | Symbol | Values |
| --- | --- | --- |
| **Interrupted time series characteristics** |  |  |
| Series length | $T$ | 12, 48, 100 |
| Interruption time point | $T_I$ | $T/2$ |
| Intercept | $\beta_0$ | 0 |
| Pre-interruption slope | $\beta_1$ | 0 |
| Immediate level-change | $\beta_2$ | 0, 1 |
| Slope-change post-interruption | $\beta_3$ | 0, 0.1 |
| Autocorrelation coefficient, fixed | $\rho$ | 0, 0.2, 0.4, 0.6 |
| Autocorrelation coefficient, variable<sup>†</sup> | $\rho^*$ | $\sim N(0.4, 0.15^2)$ |
| **Meta-analysis characteristics** |  |  |
| Number of ITS studies per meta-analysis | $K$ | 3, 5, 20 |
| Between-study variance of the level-change | $\tau_2^2$ | 0, $0.1^2$, $0.3^2$ |
| Between-study variance of the slope-change | $\tau_3^2$ | 0, $0.01^2$, $0.05^2$ |
| <sup>†</sup> $\rho^*$ was resampled when the selected autocorrelation fell outside allowable values (i. e., $\rho^* < -1$ or $\rho^* > 1$) | | |

Meta-analyses were generated with the number of ITS studies per meta-analysis being fixed at 3, 5 and 20 studies^54^. Furthermore, they were generated assuming a between-study variance of 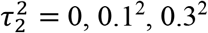 for level-change heterogeneity, and 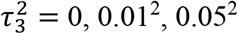 for slope-change heterogeneity^8, 17^.

#### 2.2.2 Estimands and other targets

The estimands of interest in this simulation study were the meta-analytic effects for the immediate level-change and slope-change parameters from Equation 1 (fixed-effect *β*_2_ and *β*_3_, and random-effects 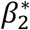 and 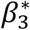), and the between-study variance for both parameters 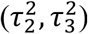

#### 2.2.3 Statistical methods to analyse ITS studies

The ITS studies were analysed with both OLS and REML (Figure 2). When REML failed to converge, we used PW, and if this failed, we used OLS. The choice of ITS estimation methods was based on methods that are commonly used and those shown to have better performance^8, 12, 17, 22^.

**Figure 2.**
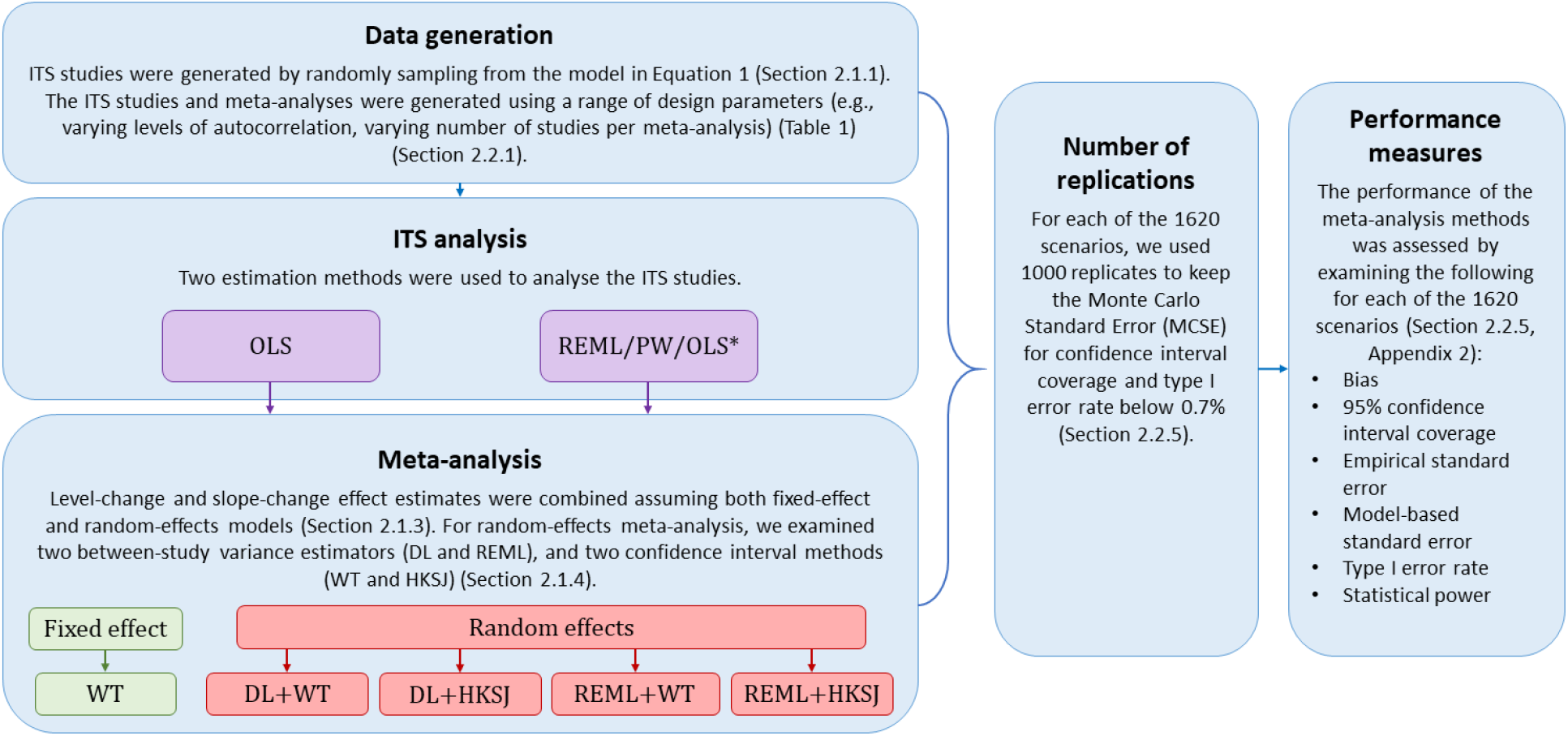
Simulation procedure and analysis methods. DL, DerSimonian and Laird. HKSJ, Hartung-Knapp / Sidik-Jonkman. ITS, interrupted time series. OLS, ordinary least squares. PW, Prais-Winsten. REML, restricted maximum likelihood. WT, Wald-type. *The estimation methods for ITS analysis are listed in order of preference, i.e., REML is used whenever it converges, while PW followed by OLS are used in the case of non-convergence.

#### 2.2.4 Statistical methods for meta-analysis

Level-change and slope-change effect estimates were combined assuming both fixed-effect and random-effects models (Figure 2). For random-effects meta-analysis, we examined two between-study variance estimators (DL and REML) and two confidence interval methods (WT and HKSJ) (Section 2.1.4). All meta-analyses were implemented using *meta* in Stata version 16.1^55^.

#### 2.2.5 Performance measures

The performance of the meta-analysis methods was assessed by examining *bias, 95% confidence interval coverage, empirical standard error, model-based standard error, type I error rate*, and *statistical power* (see Appendix 2 – Table S1 for definitions). For each of the 1620 combinations of design parameters (scenarios), we used 1000 replicates to keep the Monte Carlo Standard Error (MCSE) for confidence interval coverage and type I error rate below 0.7%^39^. The non-convergence rate of the REML and PW estimation methods were tabulated.

#### 2.2.6 Simulation procedures

Prior to running the simulations, data generation mechanisms were checked by initially simulating series with 100,000 datapoints and meta-analyses with 50 included studies, to ensure the estimated level-change, slope-change, autocorrelation and estimates of heterogeneity matched their input values, that is, that these estimators were all consistent in the statistical sense in large samples. Scatter and box plots were used to visualise the performance for all metrics.

The simulation was conducted using Stata version 16.1^55^ and results were visualised using R version 4.1.0 (*dplyr*^*56*^, *foreign*^*57*^, *ggplot2*^*58*^). All code for generating and analysing the simulated data are available in the Monash University repository known as Bridges^59^.

## 3 Results

For simplicity of presentation, we restrict the descriptions of our findings to a limited number of the simulation scenarios, meta-analysis methods and ITS effect measures. We focus on scenarios in which the data were generated with an immediate level-change (*β*_2_) of 1, a slope-change (*β*_3_) of 0.1, and where autocorrelation was fixed at specific magnitudes; autocorrelation variability had no impact on performance (Appendix 2.8). The simulation performance measures (aside from power) were not impacted by combinations of level- and slope-changes. Furthermore, given minimal difference in performance between the random-effects methods with DL and REML between-study variance estimators, we restrict presentation of findings to i) DL between-study variance estimator with WT confidence intervals (DL+WT) and ii) REML between-study variance estimator with HKSJ confidence intervals (REML+HKSJ), with the former representing the most common method combination. Results from all random-effects methods are presented in Appendix 2. Finally, we focus on the performance of the meta-analysis methods for combining immediate level-change estimates, given the patterns for slope-change were similar across the scenarios (see Appendix 4 for all findings).

### 3.1 Bias of meta-analytic level-change

All meta-analysis method and ITS analysis combinations yielded approximately unbiased estimates of meta-analytic level-change across all simulated scenarios [Appendix 3.2].

### 3.2 Confidence interval coverage of meta-analytic level-change

When data were generated under a fixed-effect model (i.e., no underlying level-change heterogeneity), fixed-effect meta-analysis of level-change effects estimated from OLS ITS yielded coverage less than the nominal 95% level, except when autocorrelation was zero and the number of data points was 48 or greater (Figure 3(A)). The less than nominal coverage decreased further with increasing autocorrelation. When the number of ITS datapoints was 12 (Figure 3(B)), fixed-effect meta-analysis of level-change effects estimated from REML ITS yielded coverage less than the nominal 95% level, and was importantly less than when the ITS were analysed using OLS. However, when the number of datapoints was greater than 12, fixed-effect meta-analysis of REML ITS level-change estimates reached coverage close to the nominal 95% level. Random-effects meta-analysis (irrespective of the method) of level-change effects estimated from OLS ITS or REML ITS yielded coverage close to the nominal 95% level. Exceptions to this were when the number of included ITS studies was small (i.e., ≤ 5) and meta-analysed with DL+WT meta-analysis; in this circumstance, coverage, for some scenarios, was less than the nominal 95% level (but still at least 83%).

**Figure 3.**
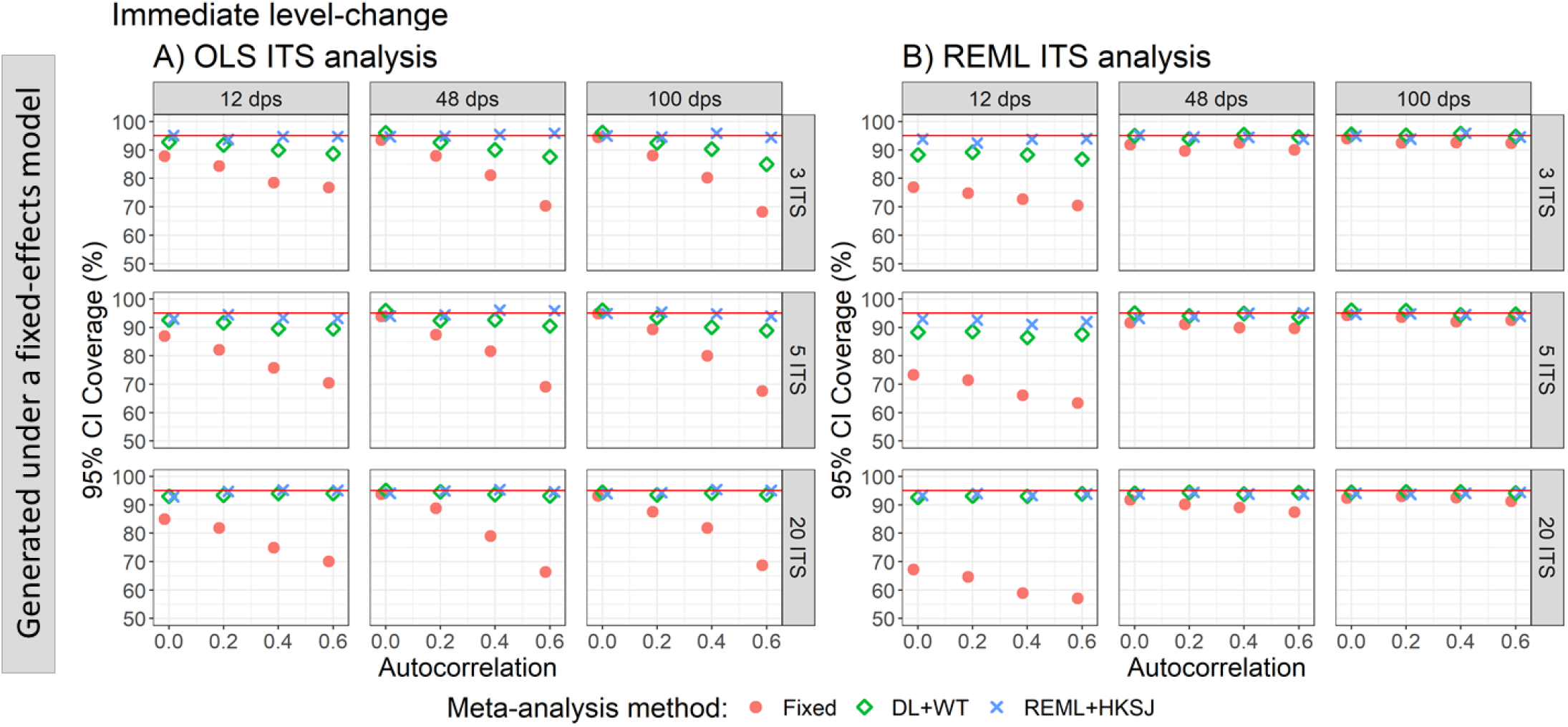
Plots of 95% confidence interval coverage of immediate level-change (y-axis) when the data was generated under a fixed-effect model and the ITS studies were analysed with OLS (A) and REML (B) using fixed-effect (red circles), DL+WT (green diamonds) and REML+ HKSJ (blue crosses) meta-analysis methods versus autocorrelation (x-axis). Plots are presented separately by combinations of the number of included studies (rows) and the number of datapoints (columns). The solid red line depicts the nominal 95% coverage level. Simulation scenarios presented include a level-change of 1, level-change between-study heterogeneity of 0, slope-change of 0, slope-change between-study heterogeneity of 0, and autocorrelation constant across included ITS studies. CI, confidence interval. DL, DerSimonian and Laird. dps, datapoints. HKSJ, Hartung-Knapp / Sidik-Jonkman. ITS, interrupted time series. OLS, ordinary least squares. REML, restricted maximum likelihood. WT, Wald-type.

When data were generated under a random-effects model (Figure 4), random-effects meta-analysis of level-change effects estimated from OLS ITS or REML ITS yielded coverage close to the nominal 95% level, irrespective of the meta-analysis method used and the magnitude of heterogeneity.

**Figure 4.**
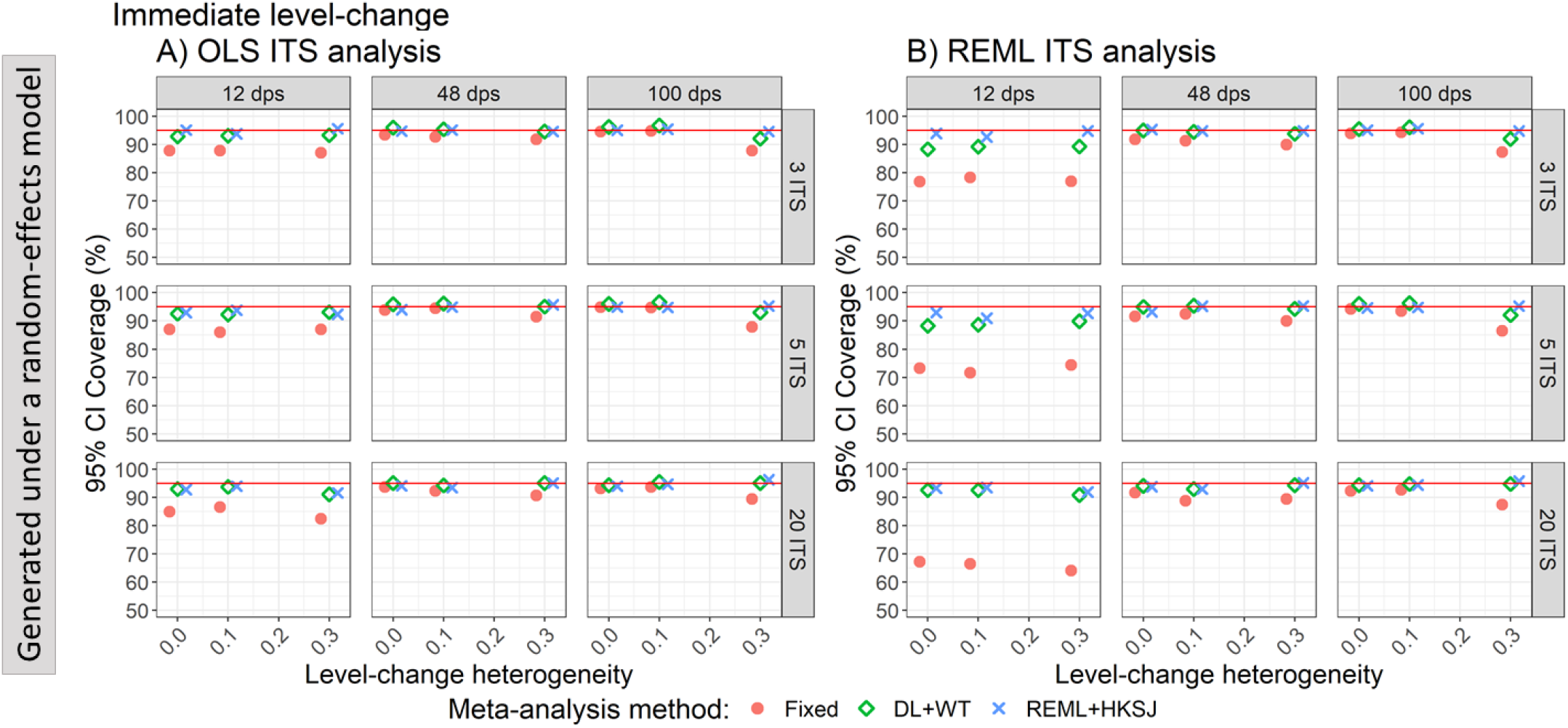
Plots of 95% confidence interval coverage of immediate level-change (y-axis) when the ITS studies were analysed with OLS (A) and REML (B) using fixed-effect (red circles), DL+WT (green diamonds) and REML+HKSJ (blue crosses) meta-analysis methods versus level-change heterogeneity (x-axis). Plots are presented separately by combinations of the number of included studies (rows) and number of datapoints (columns). The solid red line depicts the nominal 95% coverage level. Simulation scenarios presented include a level-change of 1, slope-change of 0, slope-change between-study heterogeneity of 0, and autocorrelation of 0. CI, confidence interval. DL, DerSimonian and Laird. dps, datapoints. HKSJ, Hartung-Knapp / Sidik-Jonkman. ITS, interrupted time series. OLS, ordinary least squares. REML, restricted maximum likelihood. WT, Wald-type.

### 3.3 Standard errors of meta-analytic level-change

When data were generated under a fixed-effect model, fixed-effect meta-analysis of level-change effects estimated from OLS ITS yielded model-based standard errors that were smaller than empirical standard errors (i.e., the ratio was less than one), except when autocorrelation was zero and the number of datapoints was 48 or greater (Figure 5(A)). The underestimation was exacerbated by increasing autocorrelation. Fixed-effect meta-analysis of level-change effects estimated from REML ITS yielded model-based standard errors that were smaller than the empirical standard errors when the number of ITS datapoints was 12 (Figure 5(B)), and the ratio was importantly less than when the ITS were analysed using OLS. However, when the number of datapoints was greater than 12, fixed-effect meta-analysis of REML ITS level-change estimates yielded model-based standard errors that were similar to the empirical standard errors. Random-effects meta-analysis (irrespective of the method) of level-change effects estimated from OLS ITS or REML ITS yielded model-based standard errors similar to the empirical standard errors (i.e., all ratios were close to one).

**Figure 5.**
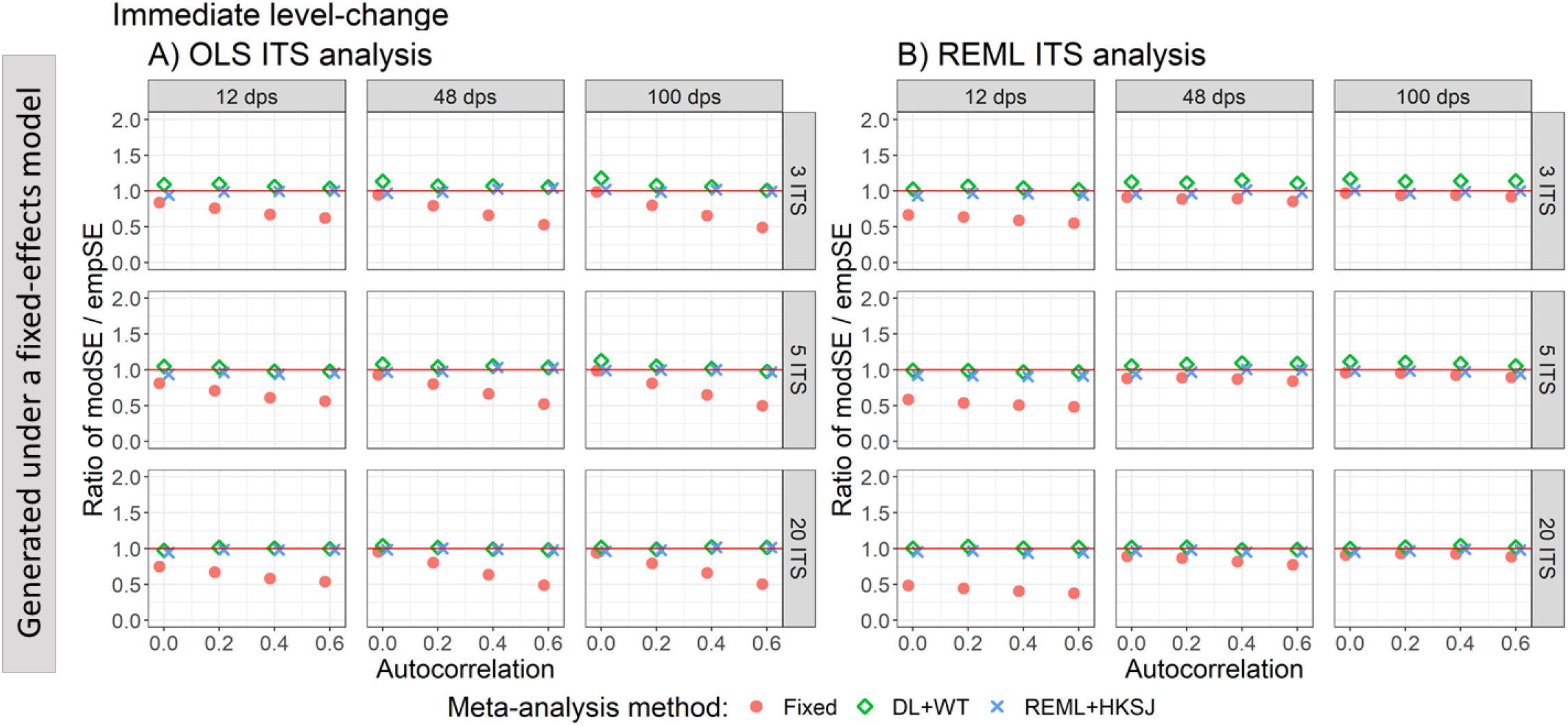
Plots of the ratio of model-based standard error (modSE) to the empirical standard error (empSE) of immediate level-change (y-axis) when the data was generated under a fixed-effect model and the ITS studies were analysed with OLS (A) and REML (B) using fixed-effect (red circles), DL+WT (green diamonds) and REML+HKSJ (blue crosses) meta-analysis methods versus autocorrelation (x-axis). Plots are presented separately by the number of included studies (rows) and number of datapoints (columns). The solid red line depicts a ratio of one, where the model-based standard error and empirical standard error are equal and thus that the model-based standard error accurately estimates the true standard error. Simulation scenarios include a level-change of 1, level-change heterogeneity of 0, slope-change of 0.1, slope-change heterogeneity of 0, and fixed autocorrelation. CI, confidence interval. DL, DerSimonian and Laird. dps, datapoints. empSE, empirical standard error. HKSJ, Hartung-Knapp / Sidik-Jonkman. ITS, interrupted time series. modSE, model-based standard error. OLS, ordinary least squares. REML, restricted maximum likelihood. WT, Wald-type.

When data were generated under a random-effects model (Figure 6), random-effects meta-analysis (irrespective of the method) of level-change effects estimated from OLS ITS or REML ITS yielded ratios close to one (i.e., appropriate estimation of standard errors in the presence of heterogeneity).

**Figure 6.**
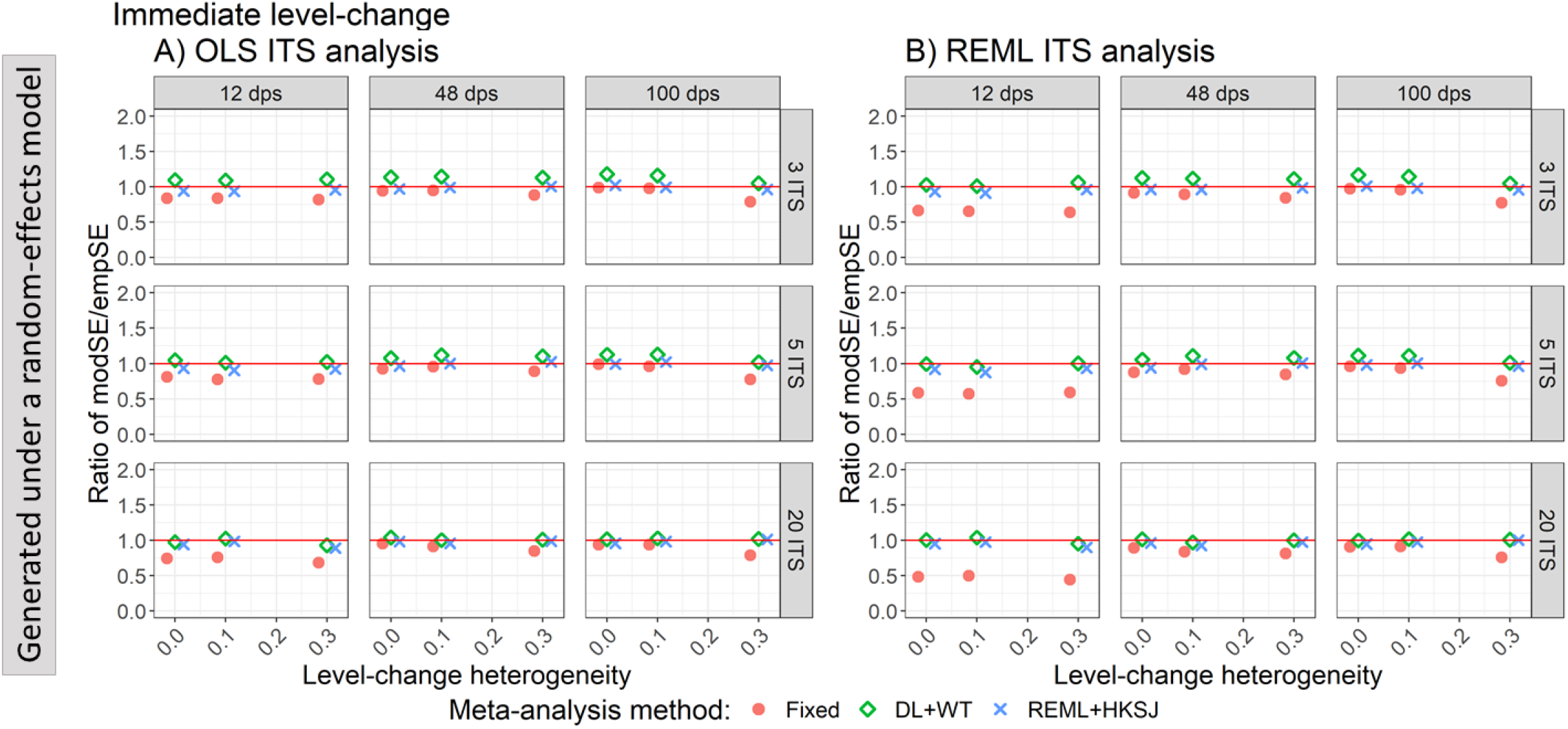
Plots of the ratio of model based standard error (modSE) to the empirical standard error (empSE) of immediate level-change (y-axis) when the ITS studies were analysed with OLS (A) and REML (B) using fixed-effect (red circles), DL+WT (green diamonds) and REML+HKSJ (blue crosses) meta-analysis methods versus level-change heterogeneity (x-axis). Plots are presented separately by combinations of the number of included studies (rows) and number of datapoints (columns). The solid red line depicts a ratio of one, where the model-based standard error and empirical standard error are equal and thus that the model-based standard error accurately estimates the true standard error. Simulation scenarios include a level-change of 1, slope-change of 0, slope-change heterogeneity of 0, and autocorrelation of 0. CI, confidence interval. DL, DerSimonian and Laird. dps, datapoints. empSE, empirical standard error. HKSJ, Hartung-Knapp / Sidik-Jonkman. ITS, interrupted time series. modSE, model-based standard error. OLS, ordinary least squares. REML, restricted maximum likelihood. WT, Wald-type.

### 3.4 Statistical power to detect a level-change

To avoid misleading interpretations of statistical power, we limit presentation of results to only scenarios in which coverage was at least 90%; acknowledging that with this threshold, there will be some artificial inflation of power when coverage is less than the nominal 95% level (due to the inflated type I error rate, i.e., 100-coverage%). When the number of ITS was large (i.e., 20), power was reasonable, irrespective of the number of time points, ITS analysis method, or meta-analysis method (Figure 7). When there were few ITS studies per meta-analysis (i.e., 5 or fewer), power importantly reduced with a decreasing number of datapoints and with increasing autocorrelation. Furthermore, power was affected by the meta-analysis method used, with REML+HKSJ yielding less power than DL+WT.

**Figure 7.**
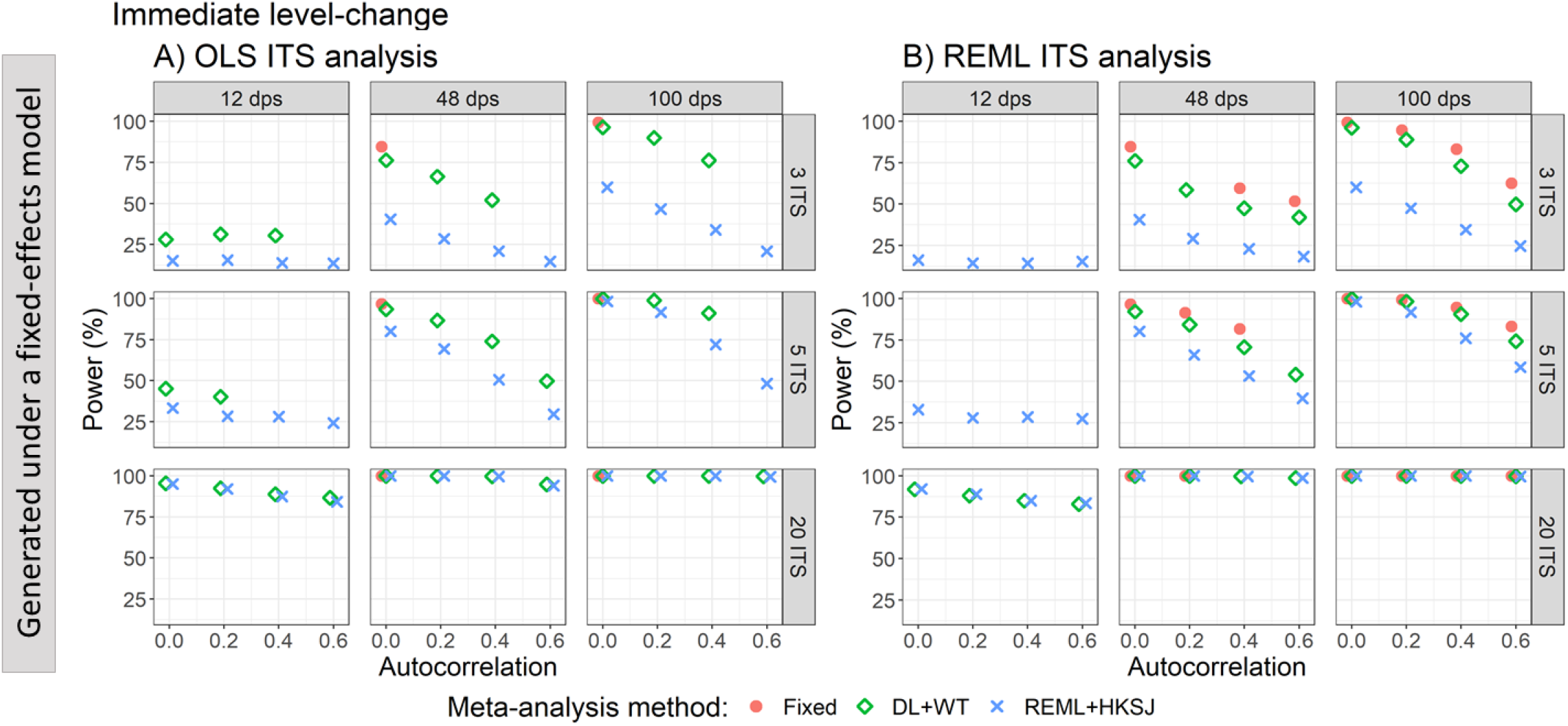
Plots of statistical power (the percentage of simulations that have a 95% confidence interval that did not include zero) of immediate level-change (y-axis) when the data was generated under a fixed-effect model and the ITS studies were analysed with OLS (A) and REML (B) using fixed-effect (red circles), DL+WT (green diamonds) and REML+HKSJ (blue crosses) meta-analysis methods versus autocorrelation (x-axis). Plots are presented separately by the number of included studies (rows) and number of datapoints (columns). Simulation scenarios include a level-change of 1, level-change heterogeneity of 0, slope-change of 0, slope-change heterogeneity of 0, and fixed autocorrelation. Only scenarios with a confidence interval coverage greater than 90% have been plotted. CI, confidence interval. DL, DerSimonian and Laird. dps, datapoints. HKSJ, Hartung-Knapp / Sidik-Jonkman. ITS, interrupted time series. OLS, ordinary least squares. REML, restricted maximum likelihood. WT, Wald-type

### 3.5 Convergence of estimation methods

When analysing the ITS datasets using REML, if the analysis failed to converge, we used PW, followed by OLS. Of the 15,120,000 ITS studies analysed using REML, 6% (899,970) failed to converge. When analysing these 899,970 ITS studies using PW, all converged, precluding the need to use OLS. Non-convergence of REML was more common for short series (17.21% for 12 datapoints, 0.57% for 48 datapoints, and 0.08% for 100 datapoints). However, the performance across all measures when comparing ITS studies analysed using REML with those analysed with PW were similar (results shown for some simulation scenarios for coverage, Appendix 3.9). Among the 3,240,000 meta-analyses of level-change performed using REML, 2,161 (0.0006%) failed to converge.

### 3.6 Estimation of heterogeneity

The magnitude of heterogeneity was overestimated in most scenarios by both REML and DL estimators. The DL estimates of heterogeneity were comparable with those from REML (Appendix 3.8). In scenarios where the ITS study standard errors were underestimated (i.e., when there were a small number of datapoints (i.e., 12 datapoints), or when autocorrelation was present but not accounted for in the analysis (i.e., OLS)), the between-study variance was overestimated (Figure 8). As autocorrelation increased, the overestimation of heterogeneity when analysed with OLS increased, and this relationship was not modified by the number of included ITS studies. When there was no underlying heterogeneity, often the estimated between-study variance was greater than zero (in 25,850/45,000, 57%, in scenarios with 3 studies, 31,055/45,000, 69%, with 5 studies and 39,859/45,000, 89%, with 20 studies, when analysed with OLS).

**Figure 8.**
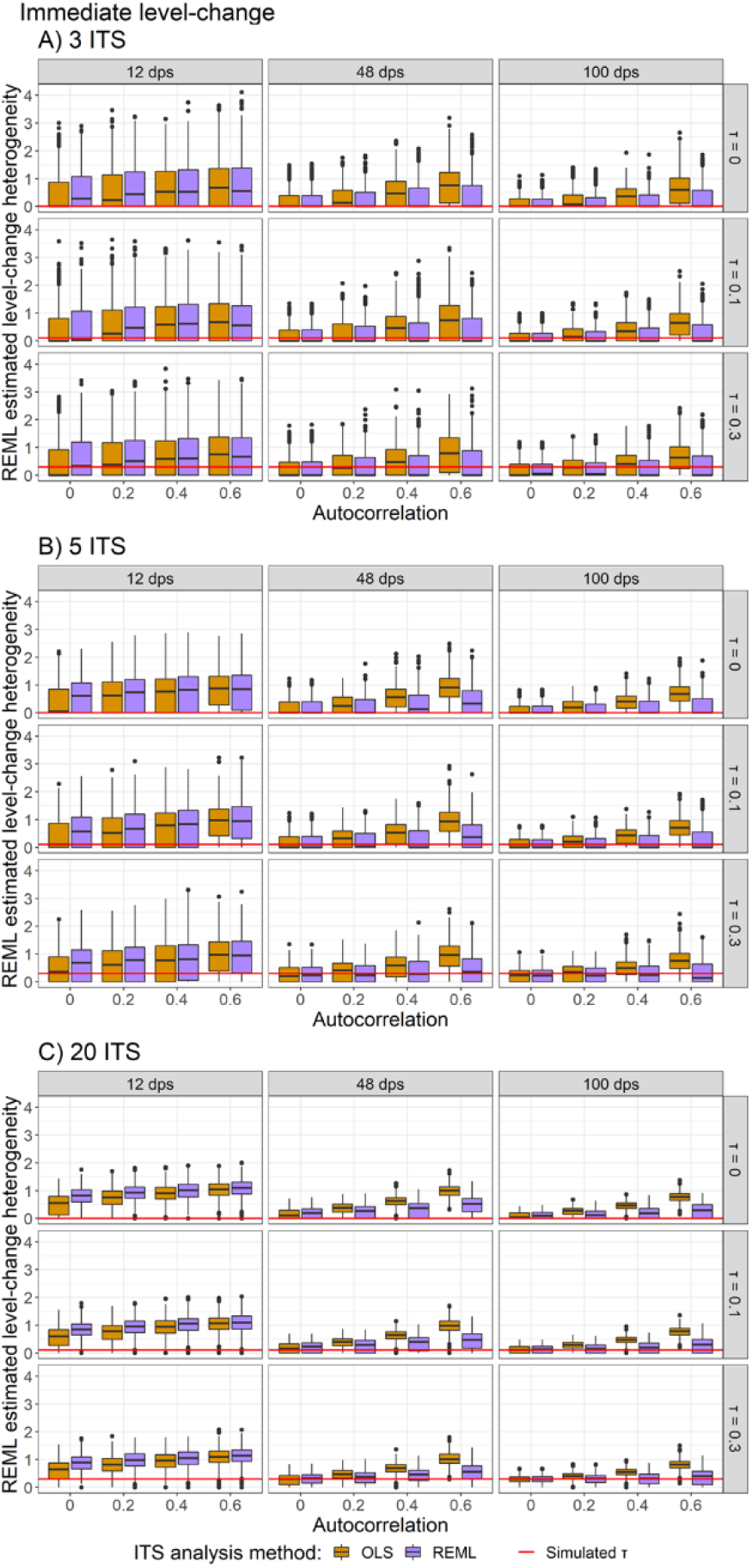
Plots of level-change heterogeneity estimated using random-effects meta-analysis with the REML between-study variance estimator (y-axis) when the A) 3 ITS, B) 5 ITS and C) 20 ITS studies were analysed with OLS (orange) and REML (purple) ITS analysis methods. Plots are presented separately by combinations of the true level-change heterogeneity (rows) and the number of datapoints (columns). The solid red lines indicate the true level-change heterogeneity. Simulation scenarios presented include a level-change of 1, slope-change of 0, slope-change heterogeneity of 0, and fixed levels of autocorrelation. CI, confidence interval. DL, DerSimonian and Laird. dps, datapoints. HKSJ, Hartung-Knapp / Sidik-Jonkman. ITS, interrupted time series. OLS, ordinary least squares. REML, restricted maximum likelihood. WT, Wald-type.

### 3.7 Performance in estimating the meta-analytic slope-change

All meta-analysis and ITS analysis method combinations yielded approximately unbiased estimates of meta-analytic slope-change in all simulated scenarios. The patterns observed for slope-change reflected those of level-change, although the specific performance values differed for coverage, empirical standard error, and power (Appendix 3). Furthermore, the patterns of between-study variance overestimation were also observed when estimating between-study variance in meta-analyses of slope-change (Appendix 2.8).

## 4 Discussion

### 4.1 Summary and discussion of key findings

Systematic reviews including meta-analyses of results from ITS studies are important for examining the effects of population-level interventions^8^. To date, there has been limited evaluation of the performance of meta-analysis methods when combining results from ITS studies, which often have characteristics that might compromise performance (e.g., short series^8, 12, 18, 60^). Our simulation study provides insight on the performance of meta-analysis methods in conjunction with ITS analysis methods, and factors that impact their performance, (e.g., series length, magnitude of autocorrelation and between-study variance). The statistical estimation methods that we examined (both the ITS analysis and meta-analysis methods) are those that are commonly used in practice and those that have been recommended for their improved performance^8, 32-34, 43, 46, 47, 51-53^.

All meta-analysis methods yielded unbiased estimates of level-change and slope-change effects. This was unsurprising given the ITS analysis methods we examined have all been shown to yield unbiased estimates of level- and slope-change^17^. However, the choice of meta-analysis method did impact the 95% confidence interval coverage, standard error, and power. We discuss these findings firstly in the context of scenarios with no underlying heterogeneity (generated under a fixed-effect model), followed by scenarios in which heterogeneity was present (generated under a random-effects model).

In scenarios *with no true underlying heterogeneity*, fixed effect meta-analysis (of level- and slope-change estimates) yielded coverage below the nominal 95% level for short series (i.e., 12 data points) or when the ITS method did not account for autocorrelation (when autocorrelation was present). In a numerical simulation study examining the performance of statistical methods for *single* ITS’, Turner et al.^17^ found that both OLS and REML analysis methods underestimated the effect estimate standard errors for short series (i.e., 12 data points). As the number of data points increased, REML ITS analysis yielded estimates of standard errors closer to the true values, even in the presence of autocorrelation. However, as expected, improvements in the estimates of standard errors were not observed when an OLS ITS analysis was used. Using OLS in the presence of autocorrelation yielded greater underestimation of the standard errors as the number of data points increased. Given that the standard error of a meta-analytic effect estimate in a fixed-effect model is a function of the within-study standard errors only, the patterns observed in the present simulation study directly reflect the patterns of Turner et al^17^.

In the same scenarios as above, with no true underlying heterogeneity, random-effects meta-analysis generally yielded coverage that was close to the nominal 95% level. An artefact of underestimating the within-study standard errors was that this induced observed between-study heterogeneity, even in the presence of no true underlying heterogeneity. Greater underestimation of the within-study standard errors led to greater overestimation of the between-study variance. For example, *greater underestimation* occurs when there are many, long series (e.g., 20 ITS studies with 100 datapoints) and autocorrelation is present (0.6) but unaccounted for in the ITS analysis (i.e., OLS is used), while *no underestimation* of within-study standard errors occurs when there are many, long series and autocorrelation is present but accounted for (i.e., REML is used for ITS analysis). Fortuitously, this under and over estimation of variances counterbalance one another when combined in the calculation of the meta-analytic effect estimate standard errors in a random-effects meta-analysis, yielding generally unbiased standard errors.

In scenarios where *true underlying heterogeneity was present*, random-effects meta-analysis generally yielded coverage close to the nominal 95% level, irrespective of the ITS analysis method used. This occurred for the same reason as above; scenarios in which the within-study standard errors were underestimated, the between-study variance was overestimated, which resulted in generally unbiased standard errors of the meta-analytic effect estimates. Because the resulting meta-analysis standard errors were generally unbiased, even with few studies (i.e., ≤ 5 ITS), the HKSJ confidence interval method offered no (or limited) advantage compared with the WT method. The HKSJ method was developed to improve coverage in scenarios in which there are few studies^32, 61^ by attempting to account for the uncertainty in estimating the between-study variance parameter, and has been shown to yield better coverage than the WT confidence interval in other simulation studies^31, 33^. Our findings may have differed because of the degree to which the between-study variance was overestimated in the present study compared with the previous studies^33, 46, 47^.

While the parameter of interest when fitting a random-effects meta-analysis is often the average interruption effect (i.e., the meta-analytic level-or slope-change), reporting the average alone provides an incomplete and potentially misleading summary of the impact of the interruption^29^. Understanding the consistency of the interruption effects (e.g., through the calculation of a prediction interval), and the factors that may explain observed heterogeneity should be of equal importance^62^. Crucially, however, this relies on accurate estimation of the between-study variance, which was most accurately estimated when REML was used for the ITS analysis, as opposed to OLS.

### 4.2 Strengths and limitations

Strengths of this numerical simulation study include the large number of design parameters (and their factorial combination) examined, and the use of a wide range of performance metrics. This allowed us to understand how the design parameters and their interactions affected key parameters required for interpreting meta-analysis results. Our design parameters were informed by those observed in practice^8, 12, 18, 22, 40^ in an attempt to create scenarios reflective of practice. We also included scenarios that would test the meta-analysis methods when the underlying assumptions were unlikely to be met.

While our simulation scenarios were extensive, there are many other design parameters and their combinations that could be investigated. One particular example, pertinent to simulation studies of meta-analysis methods, is to vary the design parameters across the individual ITS’ within the meta-analyses^36^. For example, by assuming a varying number of datapoints, pre-interruption levels and/or slopes, and methods used for ITS analysis across the ITS studies. Although we caution against generalising our findings beyond the configurations examined in the present study, our study provides a broad understanding of the factors that affect performance, which may be helpful for informing the choice of statistical methods in scenarios beyond the configurations examined here.

### 4.3 Implications for practice

For meta-analysts, our findings suggest that fitting a random-effects model generally yields coverage close to the nominal 95% level, in scenarios with and without underlying heterogeneity, and irrespective of the number of ITS studies or the method used for their analysis. Random-effects models (as opposed to fixed-effect models) may be a more appropriate model choice in the context of systematic reviews including ITS studies, as these study designs are likely to have more diversity in their characteristics (compared to randomised trials), potentially inducing statistical heterogeneity. While use of random-effects meta-analysis may mitigate some of the consequences of suboptimal ITS analysis methods in the estimation of the average effect, wherever possible, we recommend meta-analysts use effect estimates calculated from ITS methods that attempt to adjust for autocorrelation (e.g., REML). As noted above, this will lead to more accurate estimation of the between-study variance (see Section 4.1). However, caution is required in relying on the estimated heterogeneity when there are few ITS studies and the ITS have few datapoints.

For researchers undertaking the analysis of primary ITS studies, our results suggest the length of the time series and method used to analyse ITS studies have important implications for meta-analysis. To facilitate inclusion of eligible ITS studies in potential future systematic reviews, it is critical that their design and analysis methods are completely and accurately reported. Reporting should include a clear description of the ITS design (e.g., number of datapoints in the series), the model and statistical estimation method, including any adjustments made for autocorrelation, the interruption effect measure (e.g., immediate level-change), and the estimate and measure of precision^8^. Further, provision of the aggregate-level time series data (e.g., in tables or figures) would be beneficial as it would allow systematic reviewers to re-analyse time series data across the studies using a consistent and optimal method of analysis and to calculate the impact of the interruption using the desired effect measure^14, 16, 22, 60, 63, 64^.

### 4.4 Implications for future research

We examined the performance of meta-analysis methods using a two-stage meta-analysis approach; however, in certain circumstances it is possible to fit a single model that includes all the ITS to estimate the parameters in Equation (1), known as the one-stage approach^26^. Gebski et al.^26^ demonstrated this with a single model fit to ITS from three hospital units and allowing the level- and slope-changes to vary via the addition of fixed effect interaction terms between the interruption effects and the hospital units. Other one-stage models could incorporate random effects for level- and slope-change, to parallel the two-stage random-effects approach. Examination of whether there are scenarios in which the one-stage approach may offer improved efficiency would be of value^26^. Further avenues for research include examining, the impact of the different analysis methods on prediction intervals, as well as more complex scenarios such as where the ITS analysis methods differed between studies in the meta-analysis, the included ITS studies have lags of greater than 1, or exhibit seasonal patterns.

### 4.5 Conclusions

Systematic reviews including meta-analyses of results from ITS studies are important for informing public health policy. Our simulation study provides evidence on the performance of meta-analysis methods when combining results from ITS studies. We found that all meta-analysis methods yielded unbiased estimates of the interruption effects. All random effects meta-analysis methods yielded coverage close to the nominal level, irrespective of the ITS analysis method used. However, the between-study heterogeneity variance was overestimated in scenarios where the ITS study standard errors were underestimated. Therefore, meta-analysts should strive to use effect estimates and standard errors that have been calculated from ITS methods that attempt to adjust for autocorrelation (such as REML).

## Highlights

### What is already known

An interrupted time series (ITS) study is a non-randomised design in which data are collected repeatedly over time before and after an interruption (such as the introduction of a bicycle helmet law). The results from multiple ITS studies may be statistically combined using meta-analysis methods; the findings of which underpin conclusions informing public health or policy decisions.

The performance of the statistical methods for analysing single ITS studies has been shown to depend on the length of the series and the underlying correlation between consecutive data points (i.e., autocorrelation). As well, the performance of meta-analysis methods is known to depend on the number of included studies and the underlying variability in the study intervention effects.

### What is new

We undertook a numerical simulation study to examine the performance of meta-analysis methods in the context of multiple ITS studies. We found that all meta-analysis methods yielded unbiased estimates of the interruption effects. Furthermore, we found that all random effects methods yielded coverage close to the nominal level, irrespective of the ITS analysis method used and other design features (e.g. the magnitude of heterogeneity). However, heterogeneity was frequently overestimated in scenarios where the ITS study standard errors were underestimated, which is more likely to arise when ITS analysis methods do not appropriately account for autocorrelation. We therefore recommend that meta-analysts should strive to use effect estimates and standard errors that have been calculated from ITS methods that adjust for autocorrelation.

### Potential impact for RSM readers outside the authors’ field

ITS studies and systematic reviews of ITS studies are used across disciplines and topics (e.g., public health, crime, economics, war and psychology) to investigate the impact of interruptions. Our findings and recommendations are therefore likely to apply across disciplines.

## Data Availability

The datasets generated and/or analysed during the current study, in addition to the code to replicate the simulation study in its entirety, are available in the Monash University repository known as Bridges, https://doi.org/10.26180/20999185.v1

## List of abbreviations

### Abbreviation Definition

ITS: Interrupted Time Series
OLS: Ordinary Least Squares
REML: REstricted Maximum Likelihood
PW: Prais-Winsten
DL: DerSimonian and Laird between-study variance estimator
WT: Wald-type confidence interval method
HKSJ: Hartung-Knapp/Sidik-Jonkman confidence

## Declarations

### Ethics approval and consent to participate

Not applicable

### Consent for publication

Not applicable

## Author contributions

J.E.M. conceived the study, and all authors contributed to its design.

E.K., wrote the code, conducted and analysed the simulations, and wrote the first draft of the manuscript, with contributions from J.E.M.

A.K., A.B.F., S.L.T., M.T. and J.E.M. contributed to revisions of the manuscript.

## Supplementary files

Supplementary file 1:

Appendix 1 – Example meta-analysis, with annotated ITS graphs

Appendix 2 – Performance measure formulae

Appendix 3 – Additional results for scenarios with a level-change of 1 and slope-change of 1 Appendix 4 – Results for alternative scenarios; other level- and slope-change combinations

Supplementary file 2:

Appendix 5 – Code and data

