## Supplementary file 1 for "Evaluation of statistical methods used to meta-analyse results from interrupted time series studies: a simulation study"

### Appendix 1 – Example meta-analysis, with annotated ITS graphs

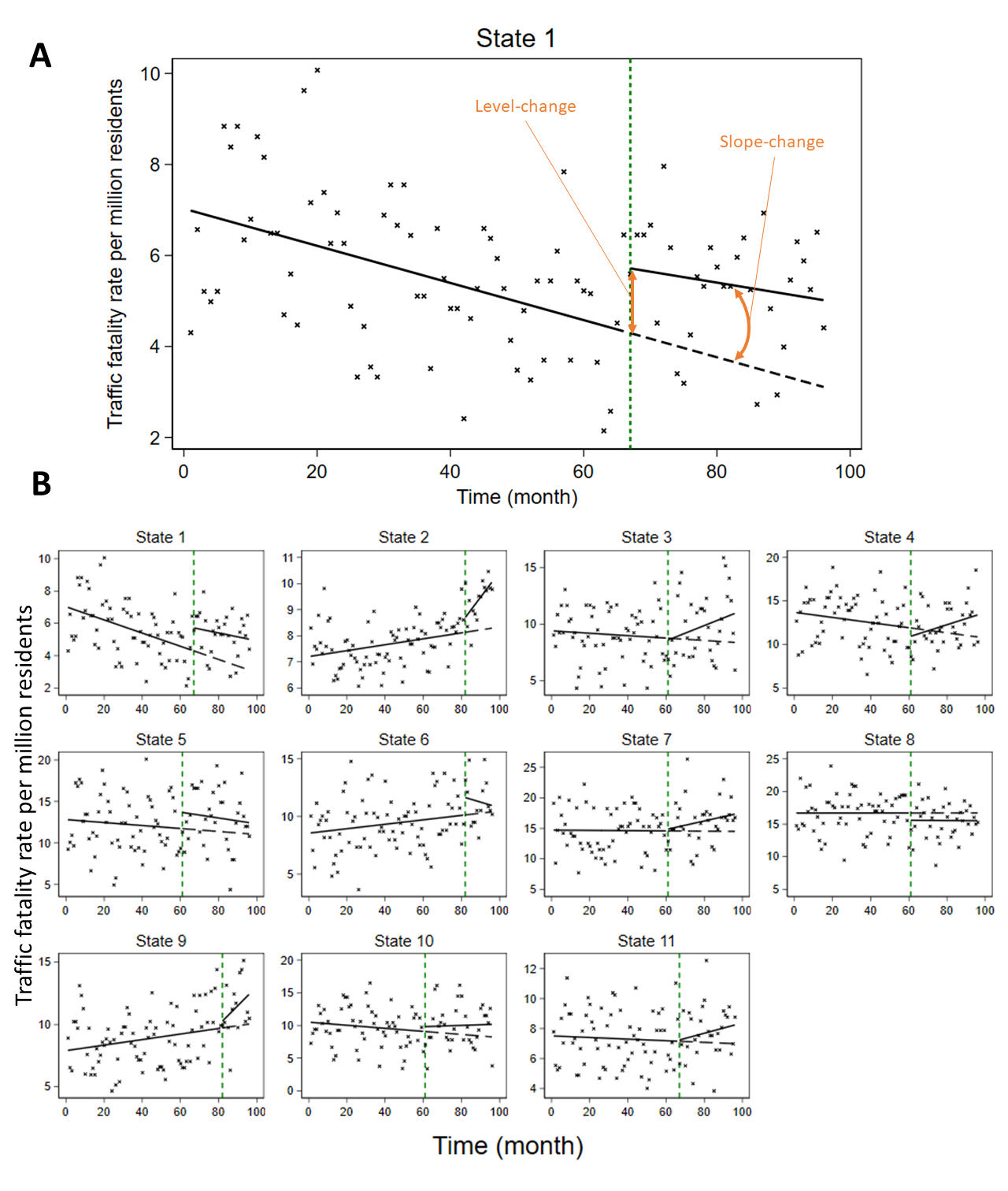

Appendix Figure S1. Examples of interrupted time series (ITS) data. A) Plot of the ITS data for State 1; shows the traffic fatality rate (per million residents) over time (months) before (left of the green dashed line) and after (right) the legalisation of recreational cannabis sales{Lane, 2019 #234}. The crosses represent data points, the solid lines represent the pre- and post-interruption trend lines and the dashed line represents the counterfactual trend line. B) Plots of the ITS data presented separately for eleven States included in meta-analysis. See Figure 1B for forest plots of the level-change and slope-change effects.

### Appendix 2 – Performance measure formulae

Table S1. Performance measure formulae

| Performance measure | Definition | Estimate | Monte Carlo standard error |
| --- | --- | --- | --- |
| Bias | $E\left[ \hat{\beta} \right]-\beta$ | $\frac{1}{n_{\mathrm{sim}}}\sum_{i=1}^{n_{\mathrm{sim}}} \hat{\beta}_{i}-\beta$ | $\sqrt{\frac{1}{n_{\mathrm{sim}}\left( n_{\mathrm{sim}}-1 \right)}\sum_{i=1}^{n_{sim}} \left( \hat{\beta}_{i}-\beta\right)^{2}}$ |
| Coverage | $Pr\left( \hat{\beta}_{low}\leq\beta\leq\hat{\beta}_{upp} \right)$ | $\frac{1}{n_{\mathrm{sim}}}\sum_{i=1}^{n_{\mathrm{sim}}} 1\left( \hat{\beta}_{low,i}\leq\beta\leq\hat{\beta}_{upp,i} \right)$ | $\sqrt{\frac{\hat{\mathrm{coverage}} \left( 1-\hat{\mathrm{coverage}} \right)}{n_{\mathrm{sim}}}}$ |
| Power / Type I error | $Pr\left( p_{i}\leq\alpha\right)$ | $\frac{1}{n_{\mathrm{sim}}}\sum_{i=1}^{n_{\mathrm{sim}}} 1\left( p_{i}\leq\alpha\right)$ | $\sqrt{\frac{\hat{\mathrm{power}} \left( 1-\hat{\mathrm{power}} \right)}{n_{\mathrm{sim}}}}$ |
| Empirical standard error | $\sqrt{\mathrm{Var}\left( \hat{\beta} \right)}$ | $\sqrt{\frac{1}{n_{\mathrm{sim}}-1}\sum_{i=1}^{n_{\mathrm{sim}}} \left( \hat{\beta}_{i}-\bar{\beta} \right)^{2}}$ | $\frac{\hat{\mathrm{EmpSE}}}{\sqrt{2\left( n_{\mathrm{sim}}-1 \right)}}$ |
| Average model-based standard error | $\sqrt{E\left[ \hat{\mathrm{Var}}\left( \hat{\beta} \right) \right]}$ | $\sqrt{\frac{1}{n_{\mathrm{sim}}}\sum_{i=1}^{n_{\mathrm{sim}}} \hat{\mathrm{Var}}\left( \hat{\beta}_{i} \right)}$ | $\sqrt{\frac{\hat{\mathrm{Var}}\left[ \hat{\mathrm{Var}}\left( \hat{\beta} \right) \right]}{4.n_{\mathrm{sim}}\hat{.\mathrm{modSE}^{2}}}}$ |
| Mean square error | $E\left[ \left( \hat{\beta}_{i}-\beta\right)^{2} \right]$ | $\frac{1}{n_{\mathrm{sim}}}\sum_{i=1}^{n_{\mathrm{sim}}} \left( \hat{\beta}_{i}-\beta\right)^{2}$ | $\sqrt{\frac{\sum_{i=1}^{n_{\mathrm{sim}}} \left( \left( \hat{\beta}_{i}-\beta\right)^{2}-\hat{\mathrm{MSE}} \right)^{2}}{n_{\mathrm{sim}}\left( n_{\mathrm{sim}}-1 \right)}}$ |
| Performance measure definitions, estimates and Monte Carlo standard error of estimate^1^.  $\beta$, the parameter of interest (i.e., meta-analytic level-change or slope-change)  $\hat{\beta}$, the estimate of the parameter of interest  $n_{sim}$, the number of simulations (i.e., 1000)  $p_{i}$, the p-value of the estimate from simulation $i$  $\alpha$, the significance level (5) | | | |

### Appendix 3 – Additional results for scenarios with a level-change of 1 and slope-change of 0.1

#### 3.1 DL+HKSJ and REML+WT

Performance results were the same for alternate combinations of between-study variance estimators and confidence interval methods (i.e., DL+HKSJ and REML+WT as opposed to DL+WT and REML+HKSJ presented in the main manuscript). There is negligible difference between the performance when between-study variance was estimated using the DL estimator versus the REML estimator. The figures in the following sections include points for all four random-effect meta-analysis methods (DL+WT, DL+HKSJ, REML+WT and REML+HKSJ).

#### 3.2 Bias

##### 3.2.1 Estimation of level-change

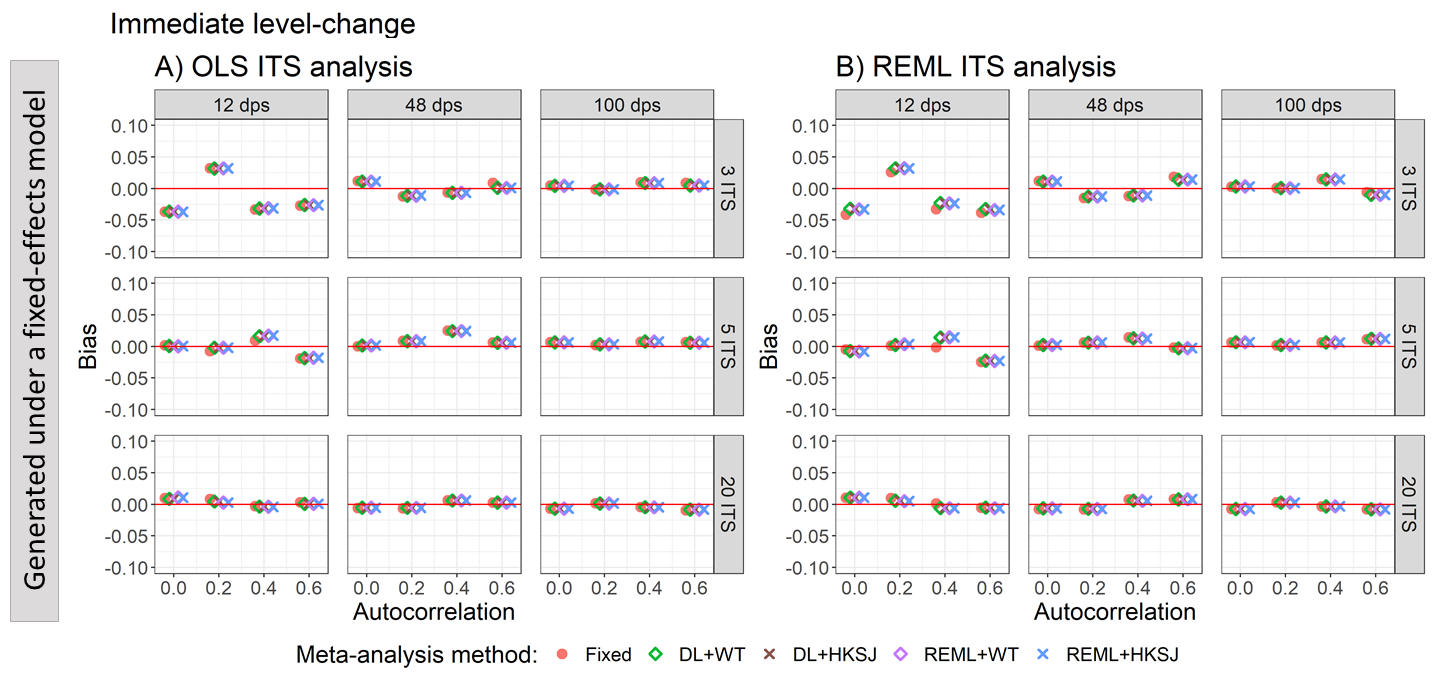

Appendix Figure S2. Plots of the bias of the immediate level-change (y-axis) when the data was generated under a fixed-effect model and the ITS studies were analysed with OLS (A) and REML (B) using fixed-effect (red circles), DL+WT (green diamonds) and DL+HKSJ (brown crosses), REML+WT (purple diamonds) and REML+HKSJ (blue crosses) meta-analysis methods versus autocorrelation (x-axis). Plots are presented separately by combinations of the number of included studies (rows) and the number of datapoints (columns). Simulation scenarios include a level-change of 1, level-change heterogeneity of 0, slope-change of 0, slope-change heterogeneity of 0, and fixed levels of autocorrelation.

DL, DerSimonian and Laird. dps, datapoints. HKSJ, Hartung-Knapp / Sidik-Jonkman. ITS, interrupted time series. OLS, ordinary least squares. REML, restricted maximum likelihood. WT, Wald-type.

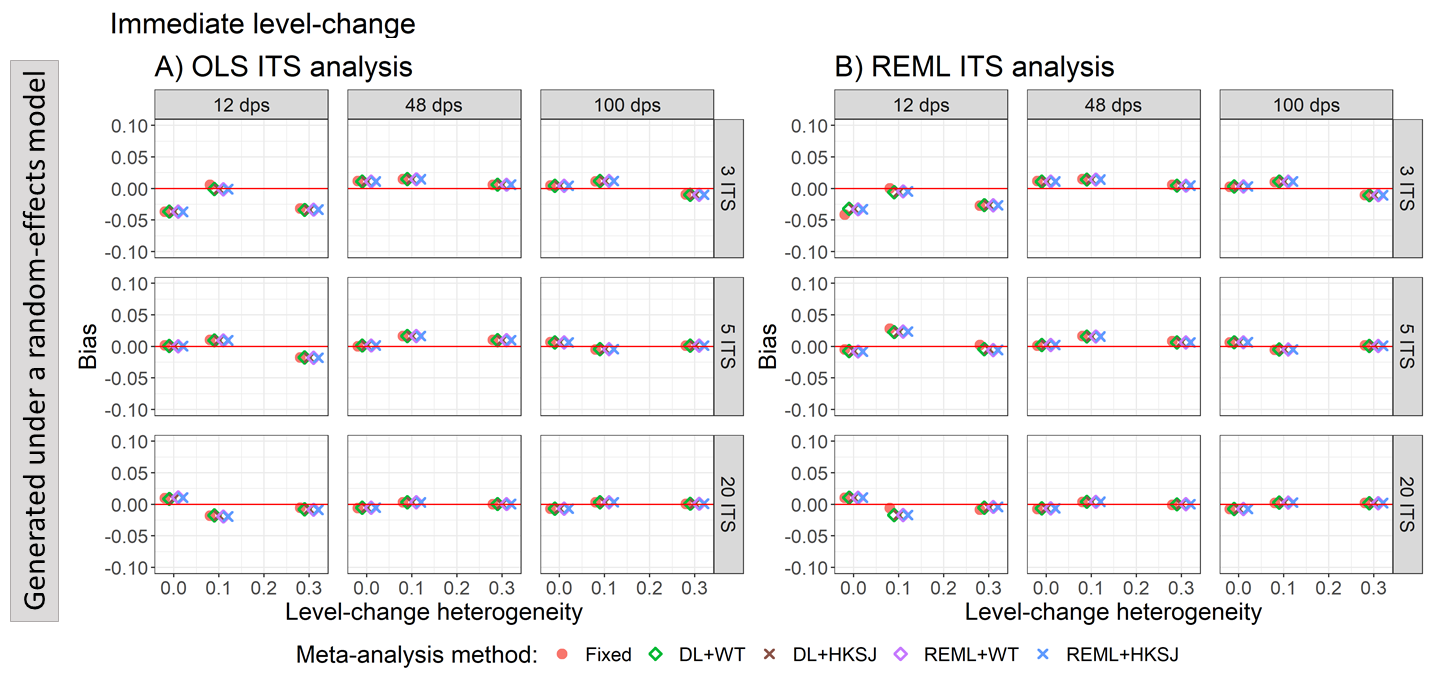

Appendix Figure S3. Plots of the bias of the immediate level-change (y-axis) when the ITS are analysed with OLS (A) and REML (B) using fixed-effect (red circles), DL+WT (green diamonds) and DL+HKSJ (brown crosses), REML+WT (purple diamonds) and REML+HKSJ (blue crosses) meta-analysis methods versus level-change heterogeneity (x-axis). Plots are presented separately by combinations of the number of included studies (rows) and the number of datapoints (columns). Simulation scenarios include a level-change of 1, slope-change of 0.1, slope-change heterogeneity of 0, and autocorrelation of zero.

DL, DerSimonian and Laird. dps, datapoints. HKSJ, Hartung-Knapp / Sidik-Jonkman. ITS, interrupted time series. OLS, ordinary least squares. REML, restricted maximum likelihood. WT, Wald-type.

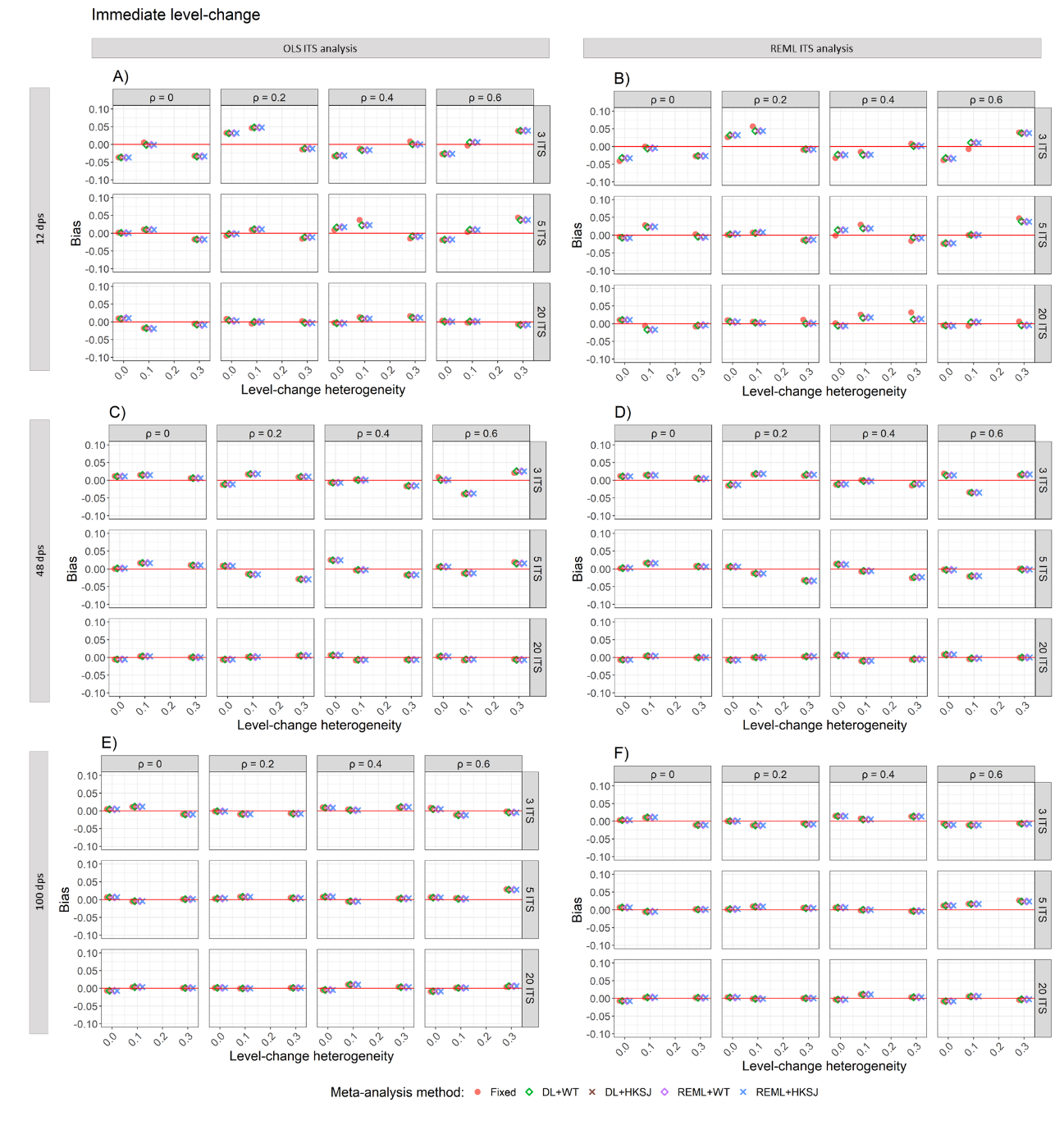

Appendix Figure S4. Plots of bias of immediate level-change (y-axis) versus level-change heterogeneity (x-axis), when the ITS studies were analysed with OLS (A, C, E) and REML (B, D, F) using fixed-effect (red circles), DL+WT (green diamonds) and DL+HKSJ (brown crosses), REML+WT (purple diamonds) and REML+HKSJ (blue crosses) meta-analysis methods. Plots are presented separately by combinations of: the series length, 12 datapoints (A, B), 48 datapoints (C, D) or 100 datapoints (E, F); the number of included studies (rows) and the level of autocorrelation (columns). The solid red line depicts the nominal 95% coverage level. Simulation settings presented include a level-change of 1, slope-change of 0.1, slope-change heterogeneity of 0, and fixed levels of autocorrelation.

DL, DerSimonian and Laird. dps, datapoints. HKSJ, Hartung-Knapp / Sidik-Jonkman. ITS, interrupted time series. OLS, ordinary least squares. REML, restricted maximum likelihood. WT, Wald-type.

##### 3.2.2 Estimation of slope-change

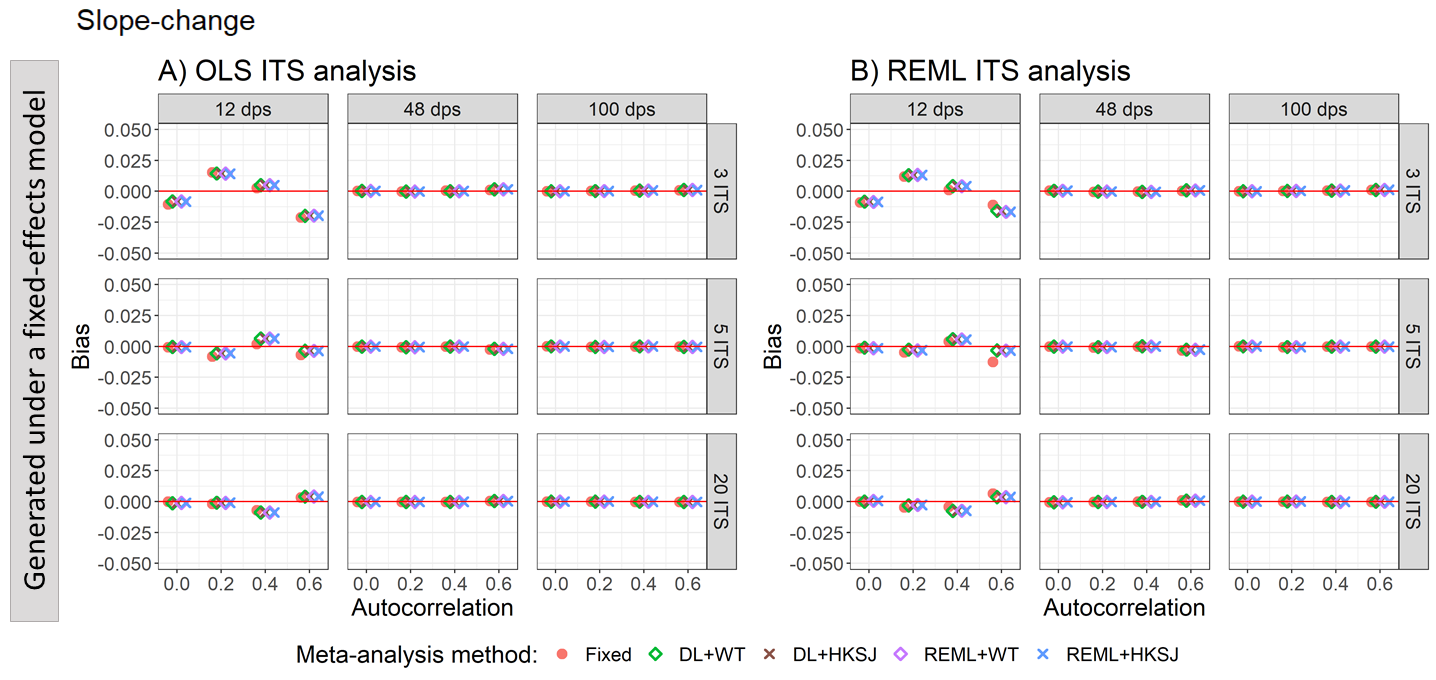

Appendix Figure S5. Plots of the bias of the slope-change (y-axis) when the data was generated under a fixed-effect model and the ITS studies were analysed with OLS (A) and REML (B) using fixed-effect (red circles), DL+WT (green diamonds) and DL+HKSJ (brown crosses), REML+WT (purple diamonds) and REML+HKSJ (blue crosses) meta-analysis methods versus autocorrelation (x-axis). Plots are presented separately by combinations of the number of included studies (rows) and the number of datapoints (columns). Simulation scenarios include a level-change of 1, level-change heterogeneity of 0, slope-change of 0.1, slope-change heterogeneity of 0, and fixed levels of autocorrelation.

DL, DerSimonian and Laird. dps, datapoints. HKSJ, Hartung-Knapp / Sidik-Jonkman. ITS, interrupted time series. OLS, ordinary least squares. REML, restricted maximum likelihood. WT, Wald-type.

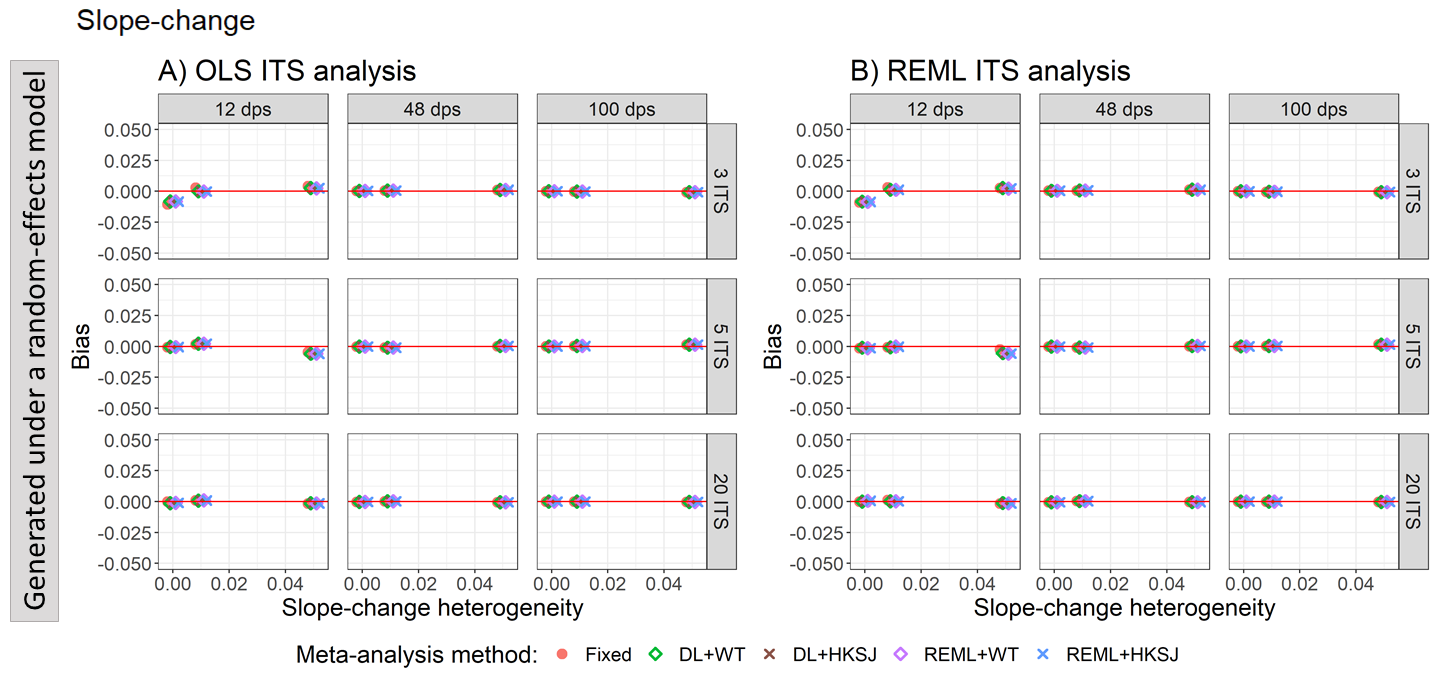

Appendix Figure S6. Plots of the bias of the slope-change (y-axis) when the ITS are analysed with OLS (A) and REML (B) using fixed-effect (red circles), DL+WT (green diamonds) and DL+HKSJ (brown crosses), REML+WT (purple diamonds) and REML+HKSJ (blue crosses) meta-analysis methods versus slope-change heterogeneity (x-axis). Plots are presented separately by combinations of the number of included studies (rows) and the number of datapoints (columns). Simulation scenarios include a level-change of 1, slope-change of 0.1, level-change heterogeneity of 0, and autocorrelation of zero.

DL, DerSimonian and Laird. dps, datapoints. HKSJ, Hartung-Knapp / Sidik-Jonkman. ITS, interrupted time series. OLS, ordinary least squares. REML, restricted maximum likelihood. WT, Wald-type.

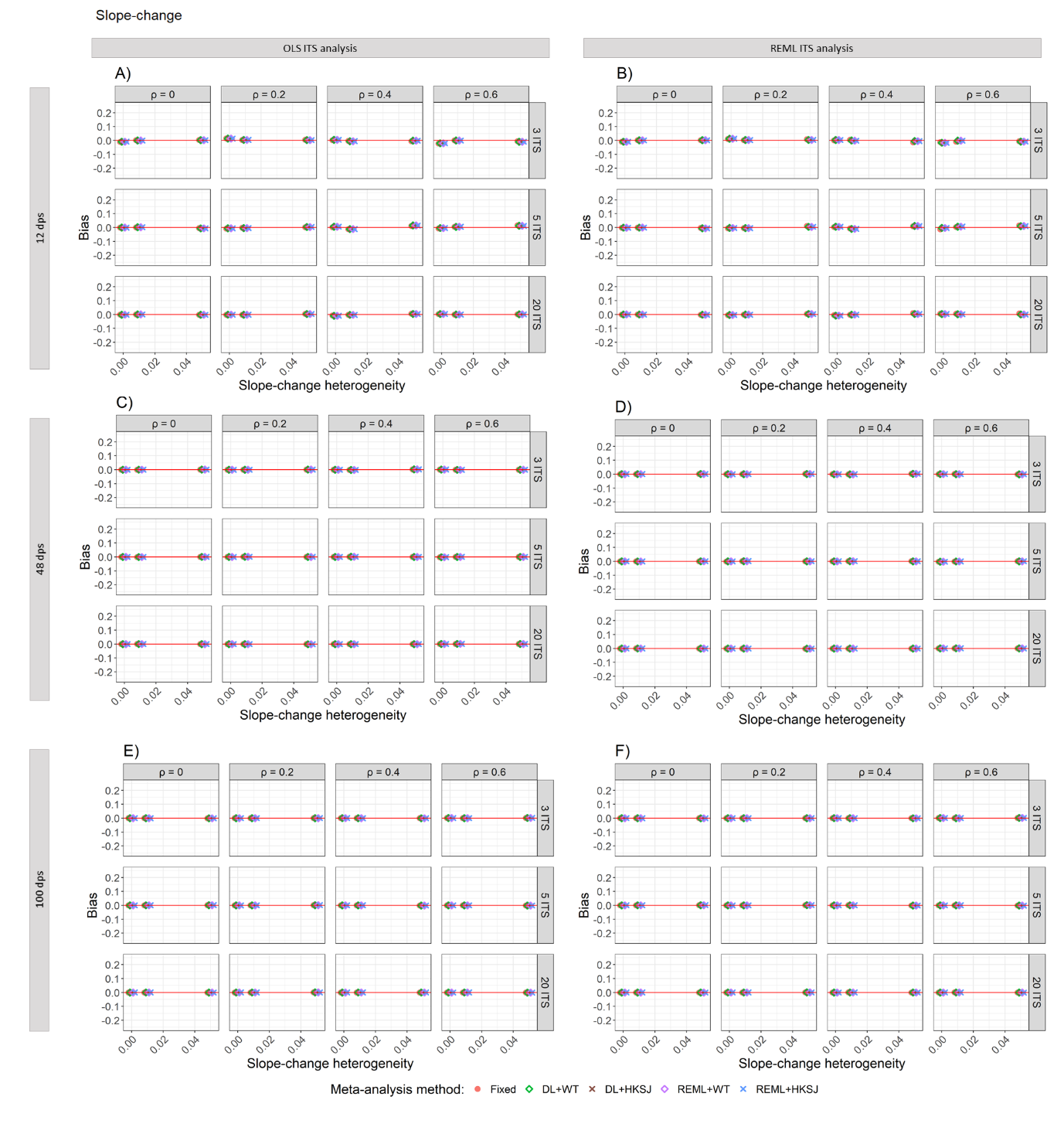

Appendix Figure S7. Plots of bias of slope-change (y-axis) versus slope-change heterogeneity (x-axis), when the ITS studies were analysed with OLS (A, C, E) and REML (B, D, F) using fixed-effect (red circles), DL+WT (green diamonds) and DL+HKSJ (brown crosses), REML+WT (purple diamonds) and REML+HKSJ (blue crosses) meta-analysis methods. Plots are presented separately by combinations of: the series length, 12 datapoints (A, B), 48 datapoints (C, D) or 100 datapoints (E, F); the number of included studies (rows) and the level of autocorrelation (columns). The solid red line depicts the nominal 95% coverage level. Simulation settings presented include a level-change of 1, slope-change of 0.1, level-change heterogeneity of 0, and fixed levels of autocorrelation.

DL, DerSimonian and Laird. dps, datapoints. HKSJ, Hartung-Knapp / Sidik-Jonkman. ITS, interrupted time series. OLS, ordinary least squares. REML, restricted maximum likelihood. WT, Wald-type.

#### 3.3 Coverage

##### 3.3.1 Estimation of level-change

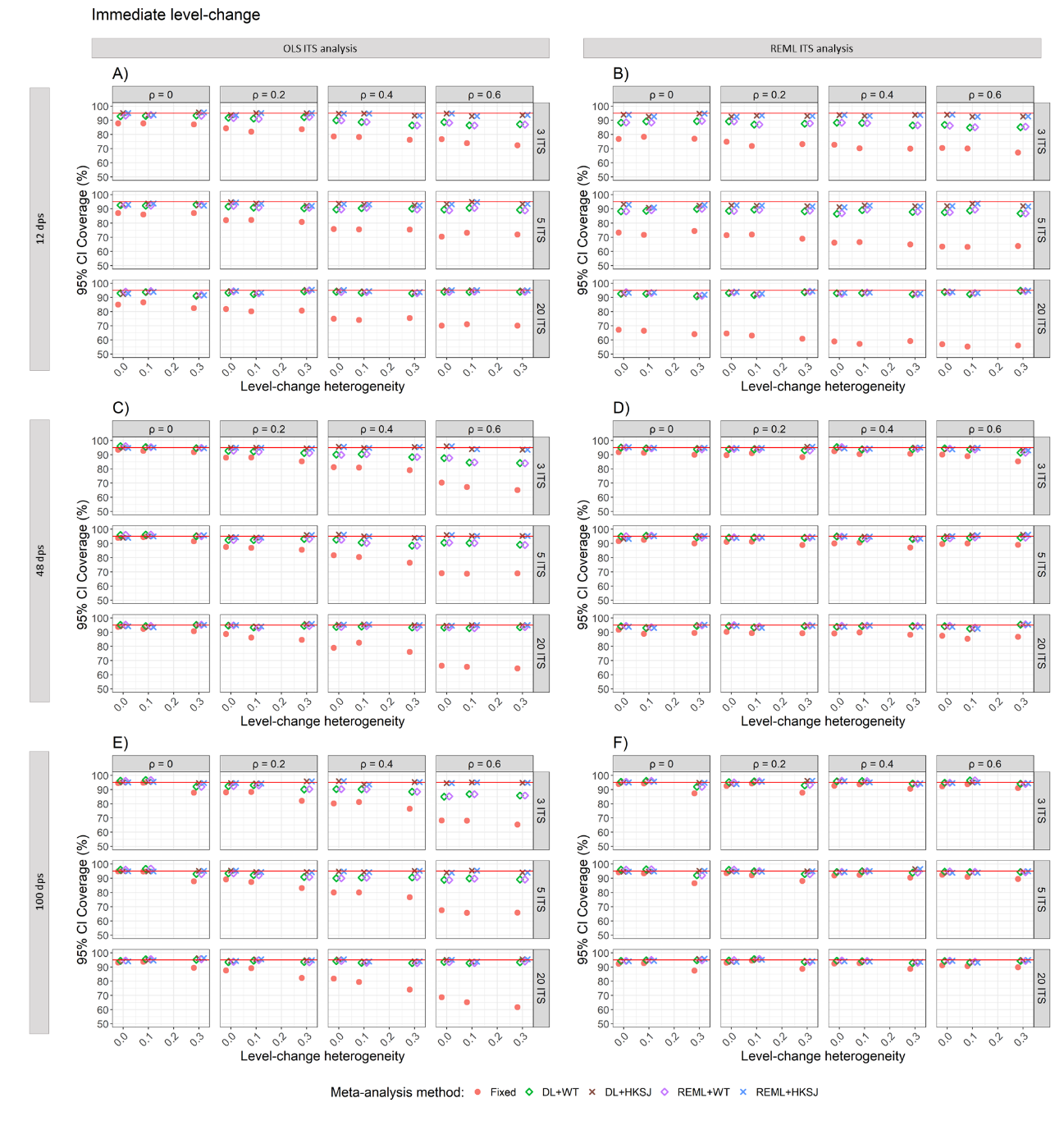

Appendix Figure S8. Plots of 95% confidence interval coverage of immediate level-change (y-axis) versus level-change heterogeneity (x-axis), when the ITS studies were analysed with OLS (A, C, E) and REML (B, D, F) using fixed-effect (red circles), DL+WT (green diamonds) and DL+HKSJ (brown crosses), REML+WT (purple diamonds) and REML+HKSJ (blue crosses) meta-analysis methods. Plots are presented separately by combinations of: the series length, 12 datapoints (A, B), 48 datapoints (C, D) or 100 datapoints (E, F); the number of included studies (rows) and the level of autocorrelation (columns). The solid red line depicts the nominal 95% coverage level. Simulation settings presented include a level-change of 1, slope-change of 0.1, slope-change heterogeneity of 0, and fixed levels of autocorrelation.

For example, to examine coverage with respect to level-change heterogeneity when the ITS are analysed using OLS, there are 20 ITS studies with 48 datapoints, an autocorrelation of 0.6, we would look to Appendix Figure S7d, at the third row and fourth column.

DL, DerSimonian and Laird. dps, datapoints. HKSJ, Hartung-Knapp / Sidik-Jonkman. ITS, interrupted time series. OLS, ordinary least squares. REML, restricted maximum likelihood. WT, Wald-type.

##### 3.3.2 Estimation of slope-change

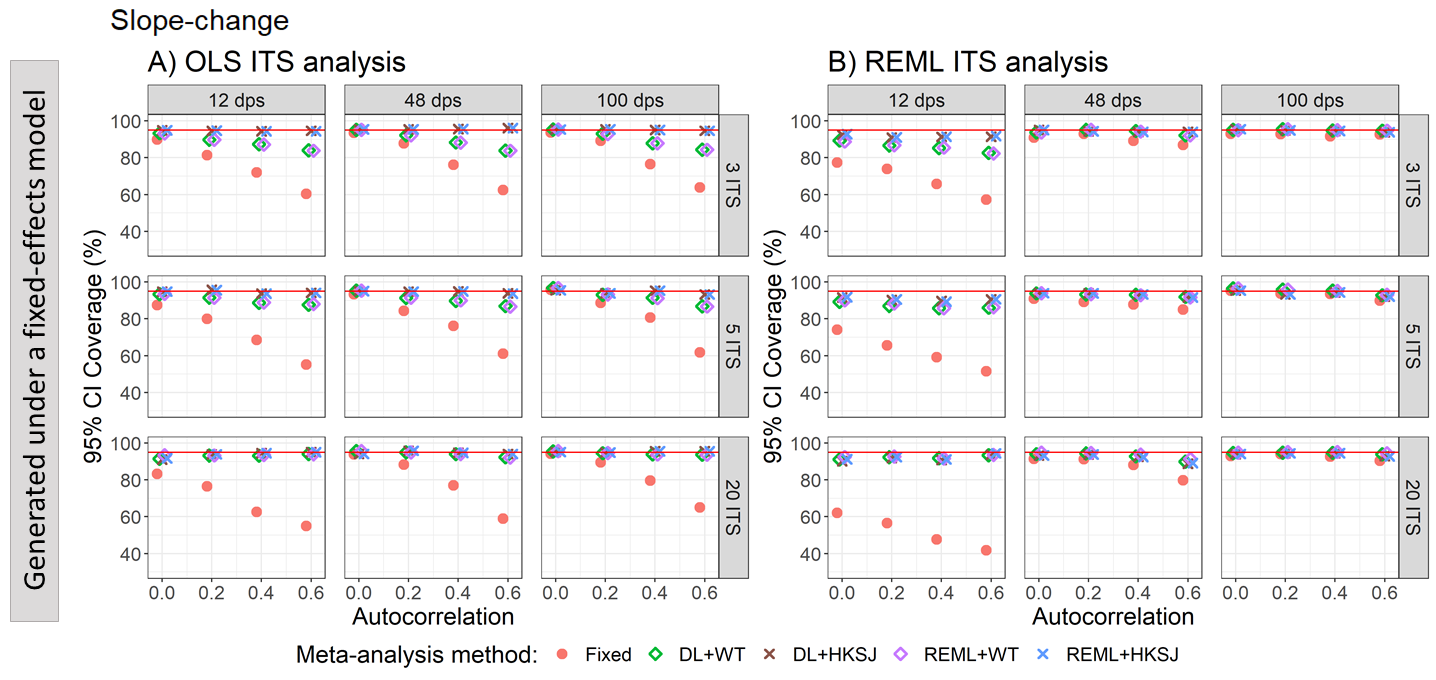

Appendix Figure S9. Plots of 95% confidence interval coverage of slope-change (y-axis) when the data was generated under a fixed-effect model and the ITS studies were analysed with OLS (A) and REML (B) using fixed-effect (red circles), DL+WT (green diamonds) and DL+HKSJ (brown crosses), REML+WT (purple diamonds) and REML+HKSJ (blue crosses) meta-analysis methods versus autocorrelation (x-axis). Plots are presented separately by combinations of the number of included studies (rows) and the number of datapoints (columns). The solid red line depicts the nominal 95% coverage level. Simulation scenarios presented include a level-change of 1, level-change heterogeneity of 0, slope-change of 0.1, slope-change heterogeneity of 0, and fixed levels of autocorrelation.

DL, DerSimonian and Laird. dps, datapoints. HKSJ, Hartung-Knapp / Sidik-Jonkman. ITS, interrupted time series. OLS, ordinary least squares. REML, restricted maximum likelihood. WT, Wald-type.

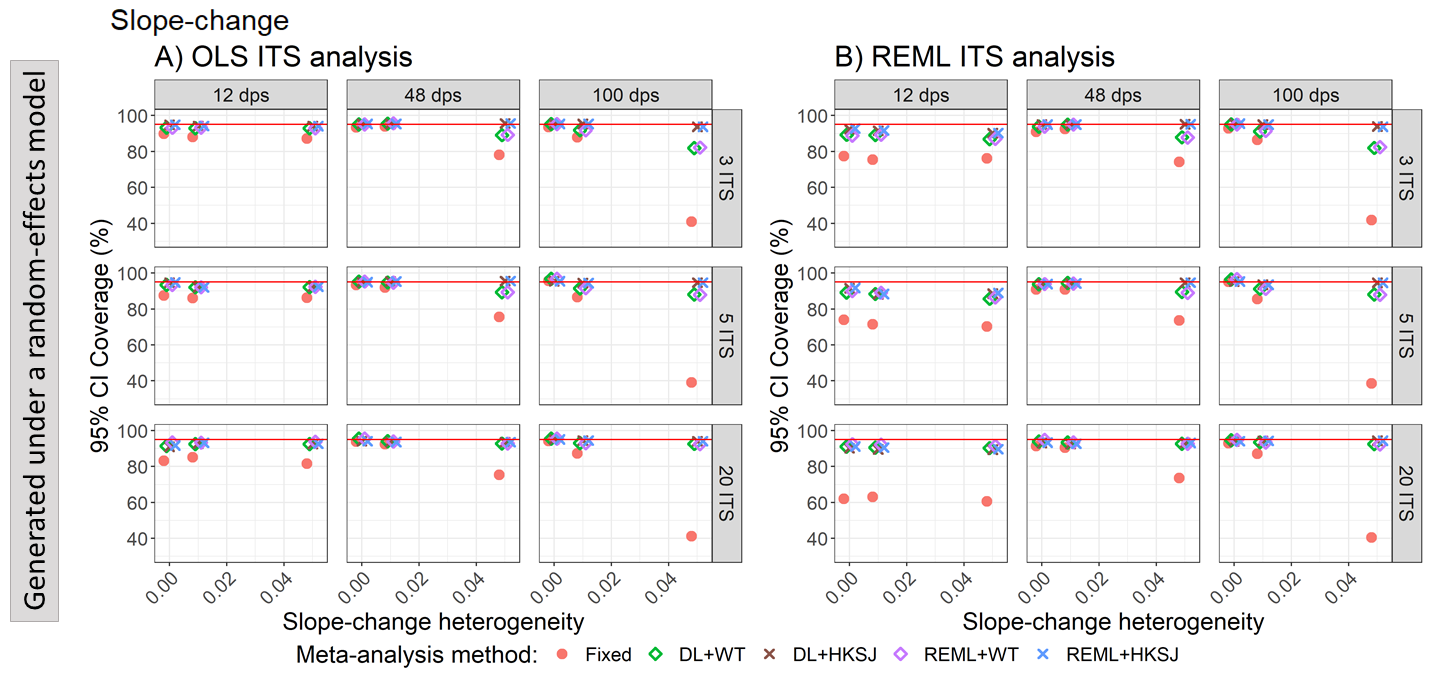

Appendix Figure S10. Plots of 95% confidence interval coverage of slope-change (y-axis) when the ITS studies were analysed with OLS (A) and REML (B) using fixed-effect (red circles), DL+WT (green diamonds) and DL+HKSJ (brown crosses), REML+WT (purple diamonds) and REML+HKSJ (blue crosses) meta-analysis methods versus slope-change heterogeneity (x-axis). Plots are presented separately by combinations of the number of included studies (rows) and number of datapoints (columns). The solid red line depicts the nominal 95% coverage level. Simulation scenarios include a level-change of 1, level-change heterogeneity of 0, slope-change of 0.1, and autocorrelation of 0.

DL, DerSimonian and Laird. dps, datapoints. HKSJ, Hartung-Knapp / Sidik-Jonkman. ITS, interrupted time series. OLS, ordinary least squares. REML, restricted maximum likelihood. WT, Wald-type.

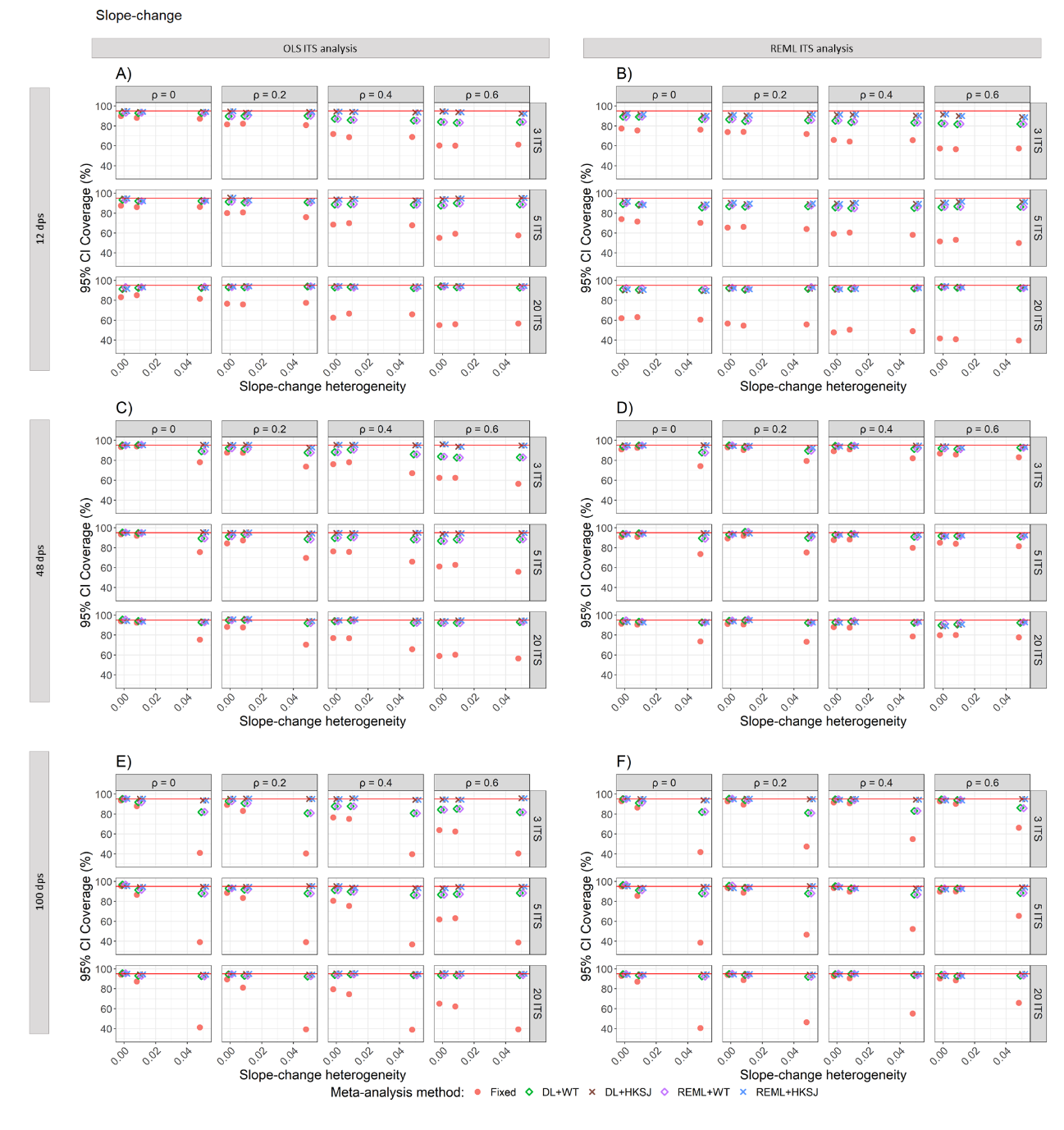

Appendix Figure S11. Plots of 95% confidence interval coverage of slope-change (y-axis) versus slope-change heterogeneity (x-axis), when the ITS studies were analysed with OLS (A, C, E) and REML (B, D, F) using fixed-effect (red circles), DL+WT (green diamonds) and DL+HKSJ (brown crosses), REML+WT (purple diamonds) and REML+HKSJ (blue crosses) meta-analysis methods. Plots are presented separately by combinations of: the series length, 12 datapoints (A, B), 48 datapoints (C, D) or 100 datapoints (E, F); the number of included studies (rows) and the level of autocorrelation (columns). The solid red line depicts the nominal 95% coverage level. Simulation settings presented include a level-change of 1, level-change heterogeneity of 0, slope-change of 0.1 and fixed levels of autocorrelation.

DL, DerSimonian and Laird. dps, datapoints. HKSJ, Hartung-Knapp / Sidik-Jonkman. ITS, interrupted time series. OLS, ordinary least squares. REML, restricted maximum likelihood. WT, Wald-type.

#### 3.4 Empirical standard errors

##### 3.4.1 Estimation of level-change

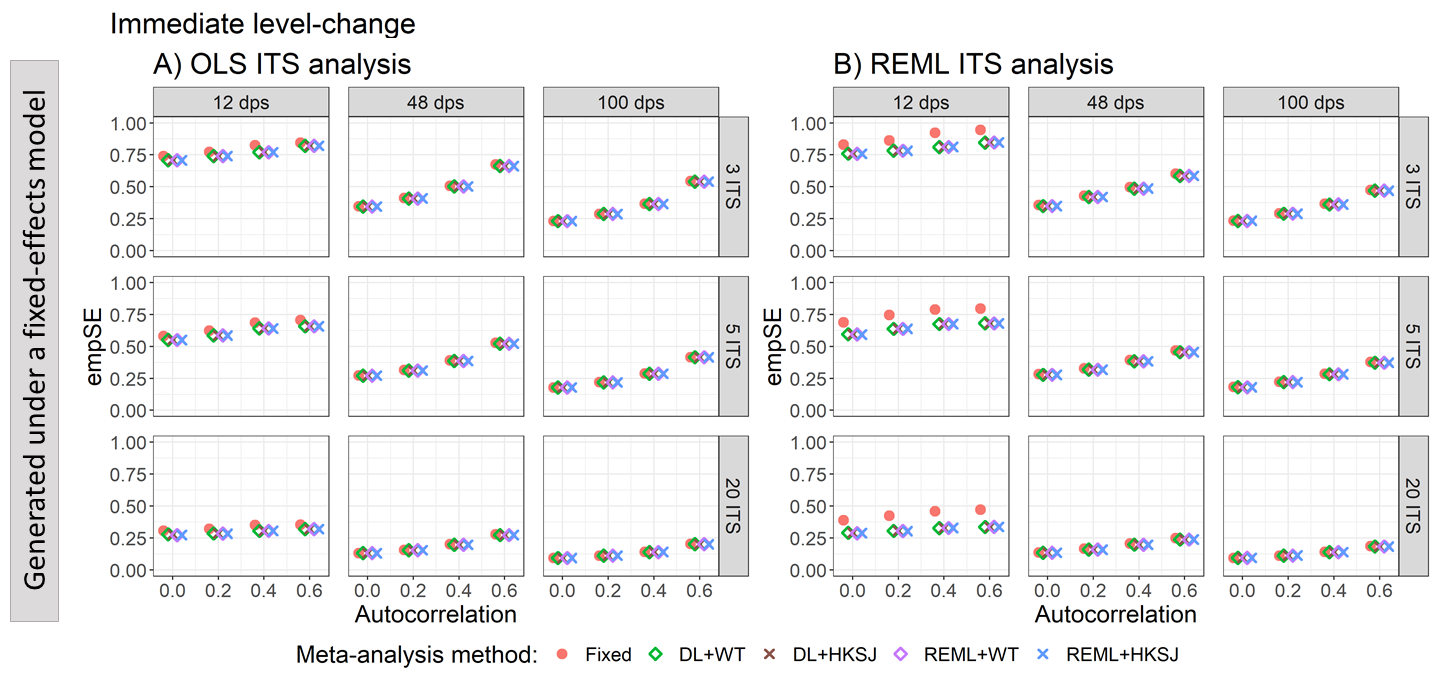

Appendix Figure S12. Plots of empirical standard error (empSE) of the immediate level-change (y-axis) when the data was generated under a fixed-effect model and the ITS studies were analysed with OLS (A) and REML (B) using fixed-effect (red circles), DL+WT (green diamonds) and DL+HKSJ (brown crosses), REML+WT (purple diamonds) and REML+HKSJ (blue crosses) meta-analysis methods versus autocorrelation (x-axis). Plots are presented separately by combinations of the number of included studies (rows) and the number of datapoints (columns). Simulation scenarios include a level-change of 1, level-change heterogeneity of 0, slope-change of 0.1, slope-change heterogeneity of 0, and fixed levels of autocorrelation.

DL, DerSimonian and Laird. dps, datapoints. HKSJ, Hartung-Knapp / Sidik-Jonkman. ITS, interrupted time series. OLS, ordinary least squares. REML, restricted maximum likelihood. WT, Wald-type.

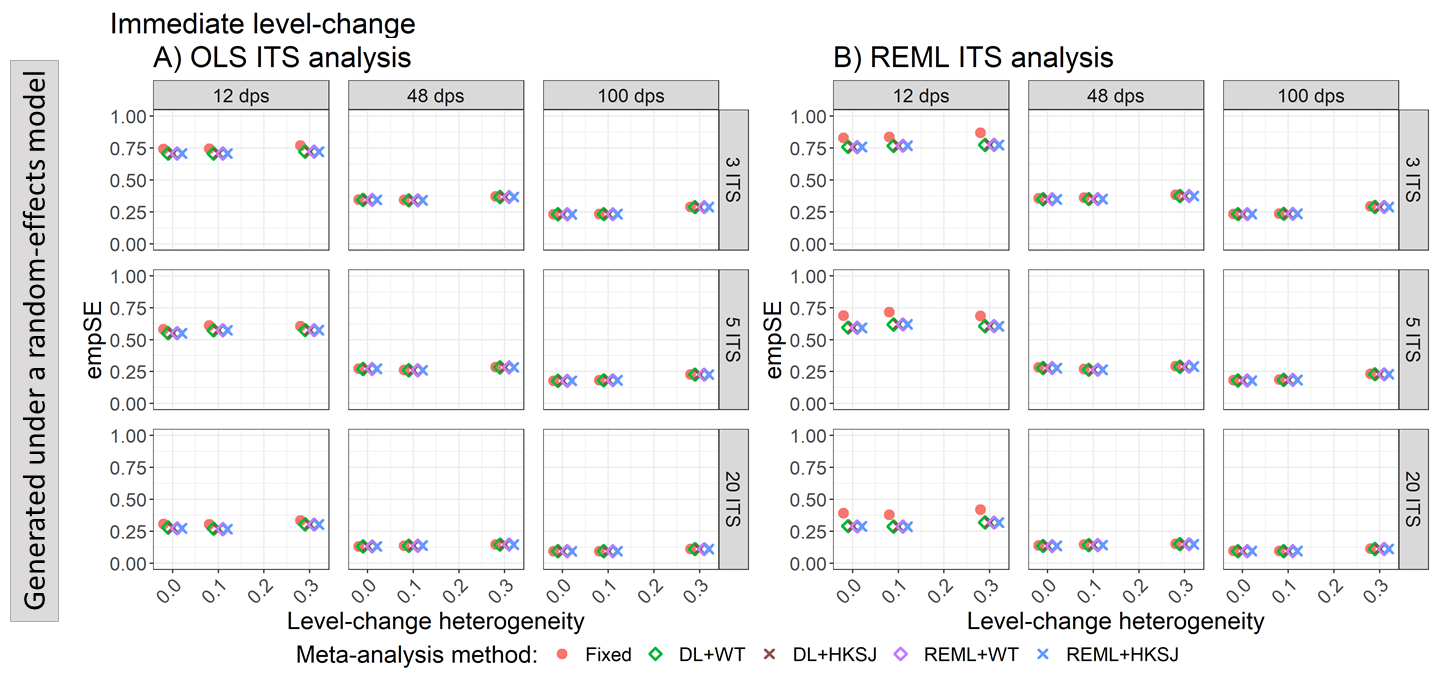

Appendix Figure S13. Plots of empirical standard error (empSE) of the immediate level-change (y-axis) when the ITS are analysed with OLS (A) and REML (B) using fixed-effect (red circles), DL+WT (green diamonds) and DL+HKSJ (brown crosses), REML+WT (purple diamonds) and REML+HKSJ (blue crosses) meta-analysis methods versus level-change heterogeneity (x-axis). Plots are presented separately by combinations of the number of included studies (rows) and the number of datapoints (columns). Simulation scenarios include a level-change of 1, slope-change of 0.1, slope-change heterogeneity of 0, and autocorrelation of 0.

DL, DerSimonian and Laird. dps, datapoints. HKSJ, Hartung-Knapp / Sidik-Jonkman. ITS, interrupted time series. OLS, ordinary least squares. REML, restricted maximum likelihood. WT, Wald-type.

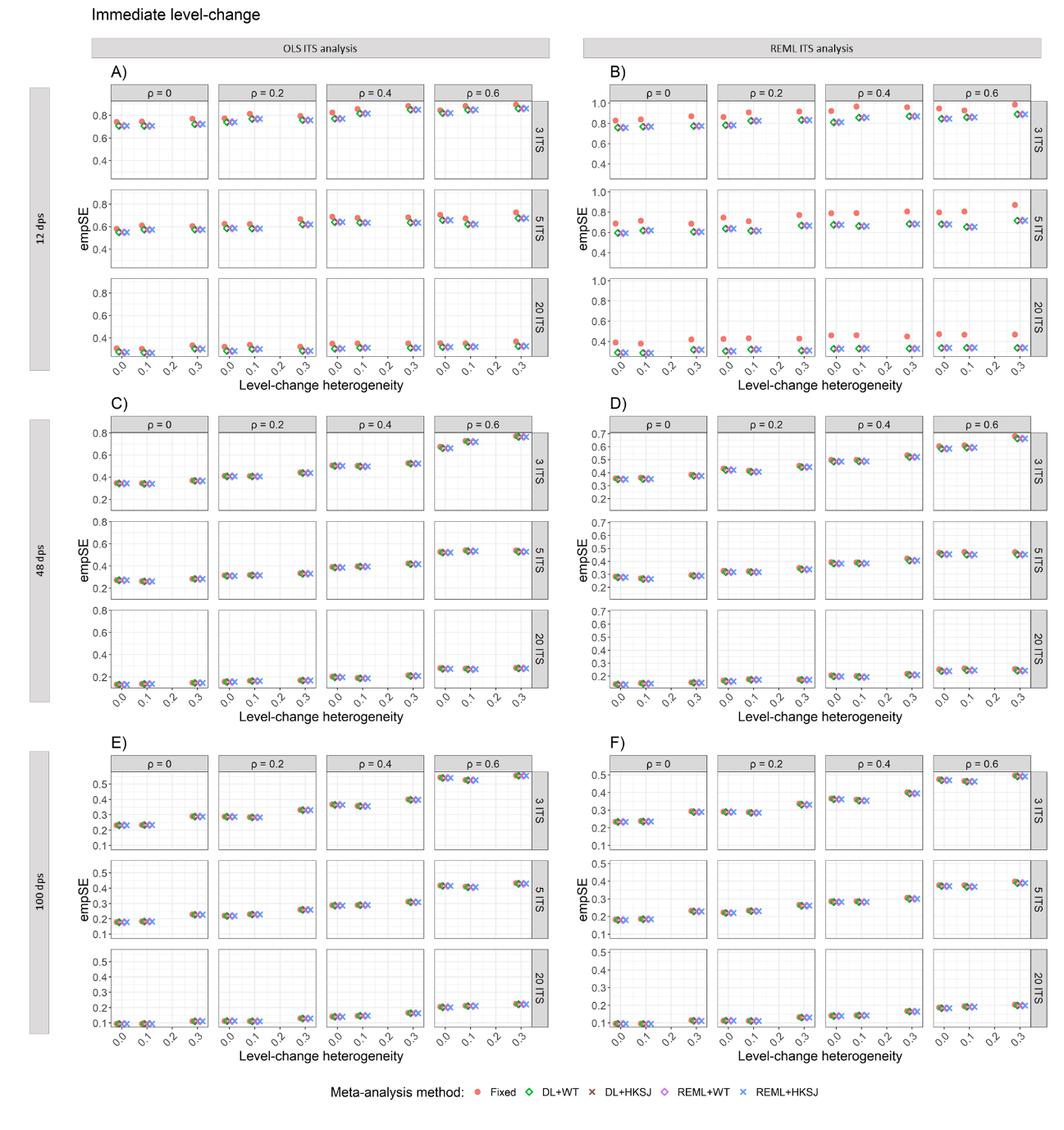

Appendix Figure S14. Plots of empirical standard error of immediate level-change (y-axis) versus level-change heterogeneity (x-axis), when the ITS studies were analysed with OLS (A, C, E) and REML (B, D, F) using fixed-effect (red circles), DL+WT (green diamonds) and DL+HKSJ (brown crosses), REML+WT (purple diamonds) and REML+HKSJ (blue crosses) meta-analysis methods. Plots are presented separately by combinations of: the series length, 12 datapoints (A, B), 48 datapoints (C, D) or 100 datapoints (E, F); the number of included studies (rows) and the level of autocorrelation (columns). The solid red line depicts the nominal 95% coverage level. Simulation settings presented include a level-change of 1, slope-change of 0.1, slope-change heterogeneity of 0, and fixed levels of autocorrelation.

DL, DerSimonian and Laird. dps, datapoints. HKSJ, Hartung-Knapp / Sidik-Jonkman. ITS, interrupted time series. OLS, ordinary least squares. REML, restricted maximum likelihood. WT, Wald-type.

##### 3.4.2 Estimation of slope-change

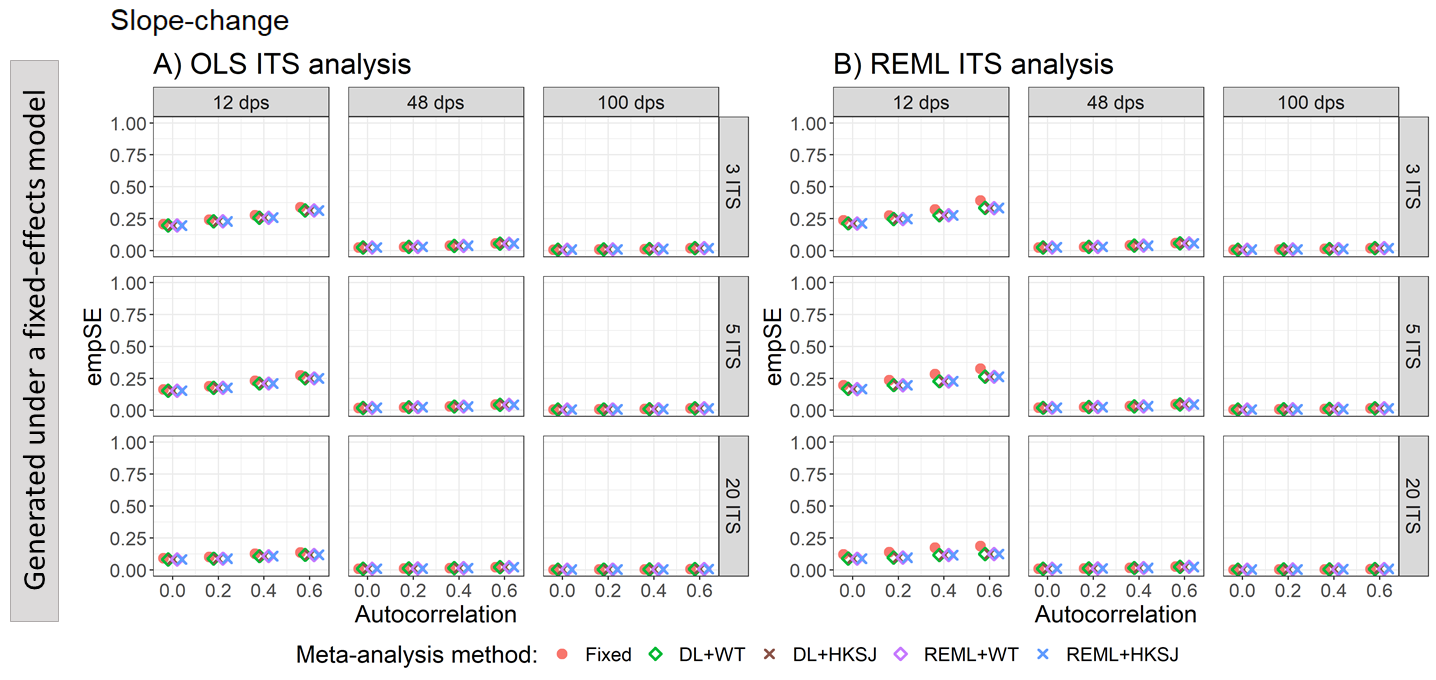

Appendix Figure S15. Plots of empirical standard error (empSE) of the immediate level-change (y-axis) when the data was generated under a fixed-effect model and the ITS studies were analysed with OLS (A) and REML (B) using fixed-effect (red circles), DL+WT (green diamonds) and DL+HKSJ (brown crosses), REML+WT (purple diamonds) and REML+HKSJ (blue crosses) meta-analysis methods versus autocorrelation (x-axis). Plots are presented separately by combinations of the number of included studies (rows) and the number of datapoints (columns). Simulation scenarios include a level-change of 1, level-change heterogeneity of 0, slope-change of 0.1, slope-change heterogeneity of 0, and fixed levels of autocorrelation.

DL, DerSimonian and Laird. dps, datapoints. HKSJ, Hartung-Knapp / Sidik-Jonkman. ITS, interrupted time series. OLS, ordinary least squares. REML, restricted maximum likelihood. WT, Wald-type.

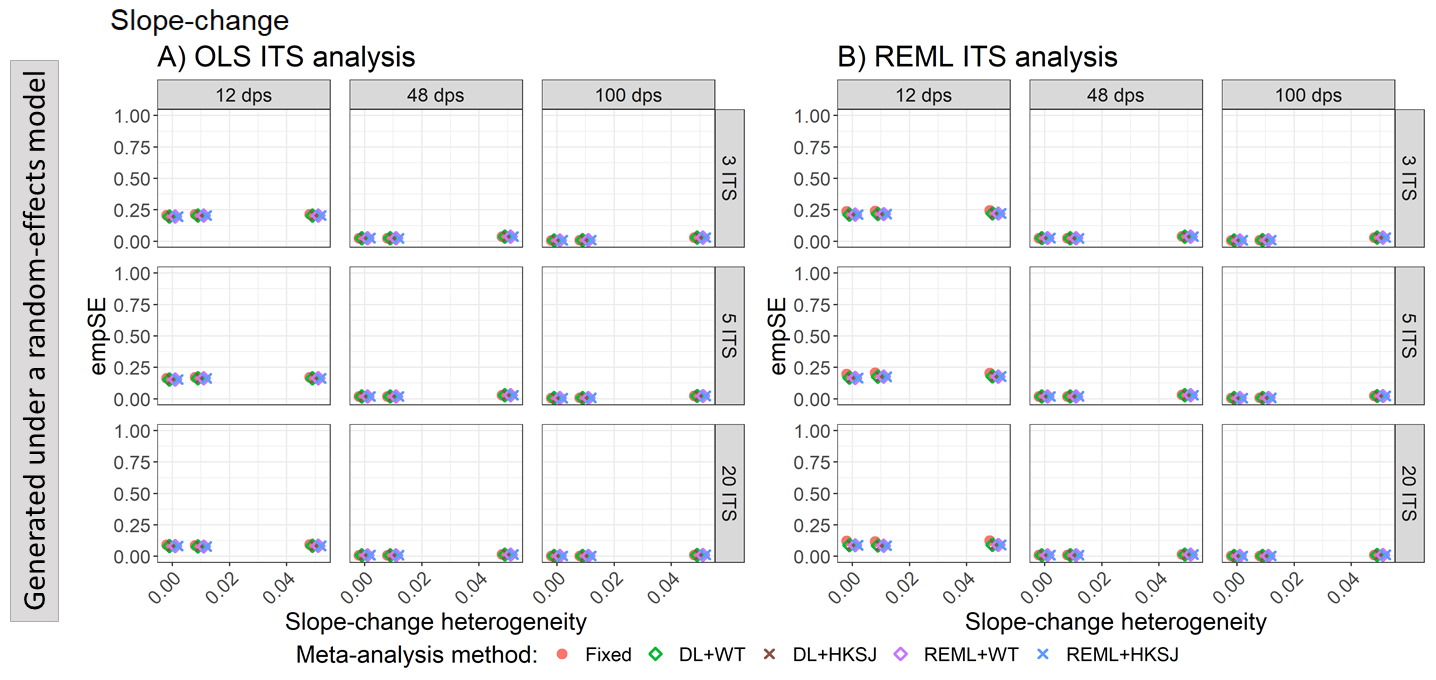

Appendix Figure S16. Plots of empirical standard error (empSE) of the immediate slope-change (y-axis) when the ITS are analysed with OLS (A) and REML (B) using fixed-effect (red circles), DL+WT (green diamonds) and DL+HKSJ (brown crosses), REML+WT (purple diamonds) and REML+HKSJ (blue crosses) meta-analysis methods versus slope-change heterogeneity (x-axis). Plots are presented separately by combinations of the number of included studies (rows) and the number of datapoints (columns). Simulation scenarios include a level-change of 1, level-change heterogeneity of 0, slope-change of 0.1, and fixed levels of autocorrelation.

DL, DerSimonian and Laird. dps, datapoints. HKSJ, Hartung-Knapp / Sidik-Jonkman. ITS, interrupted time series. OLS, ordinary least squares. REML, restricted maximum likelihood. WT, Wald-type.

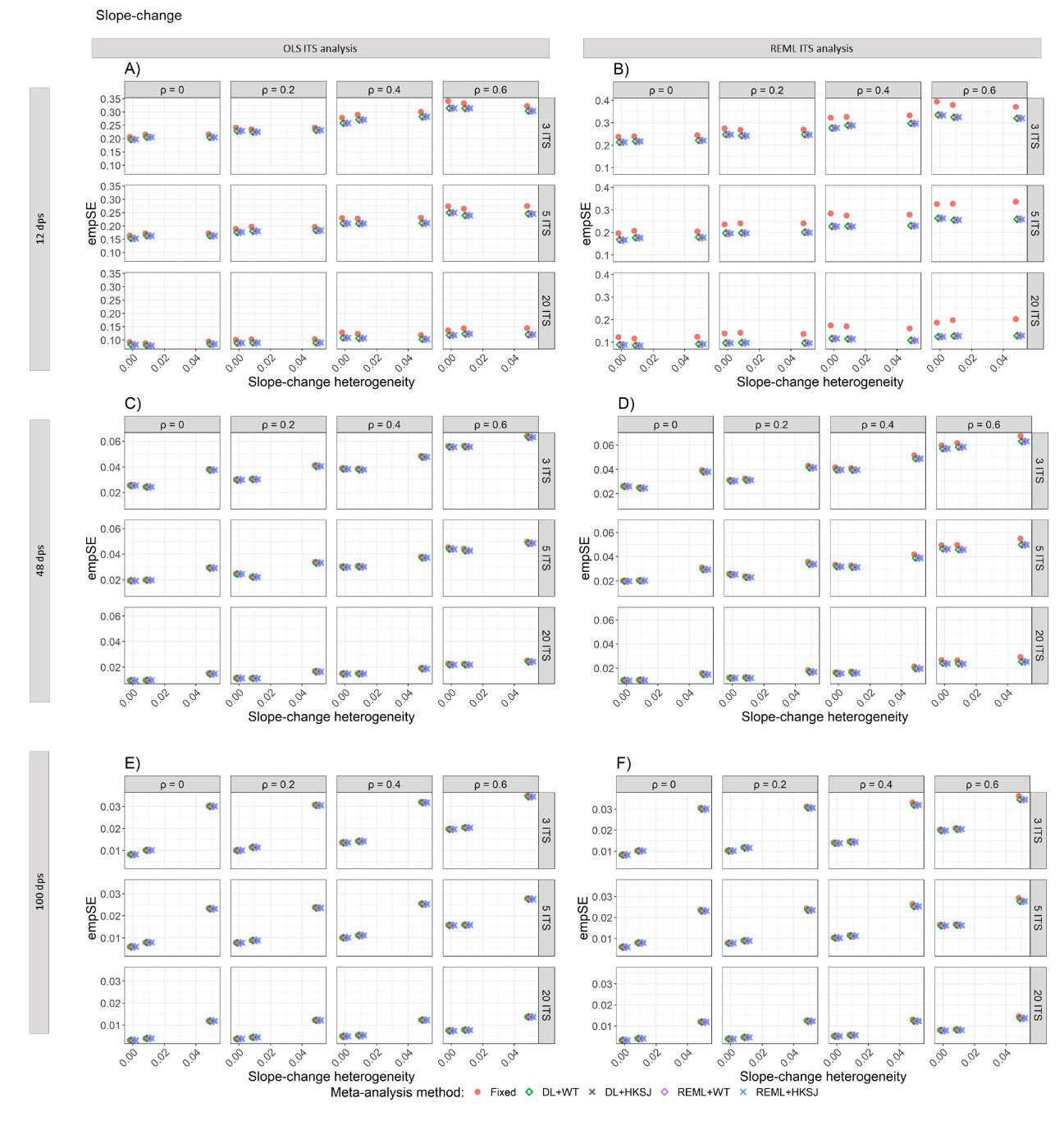

Appendix Figure S17. Plots of empirical standard error (empSE) of the meta-analytic slope-change (y-axis) versus slope-change heterogeneity (x-axis), when the ITS studies were analysed with OLS (A, C, E) and REML (B, D, F) using fixed-effect (red circles), DL+WT (green diamonds) and DL+HKSJ (brown crosses), REML+WT (purple diamonds) and REML+HKSJ (blue crosses) meta-analysis methods. Plots are presented separately by combinations of: the series length, 12 datapoints (A, B), 48 datapoints (C, D) or 100 datapoints (E, F); the number of included studies (rows) and the level of autocorrelation (columns). The solid red line depicts the nominal 95% coverage level. Simulation settings presented include a level-change of 1, level-change heterogeneity of 0, slope-change of 0.1 and fixed levels of autocorrelation.

#### 3.5 Ratio of model based standard errors to empirical standard errors for slope-change

##### 3.5.1 Estimation of level-change

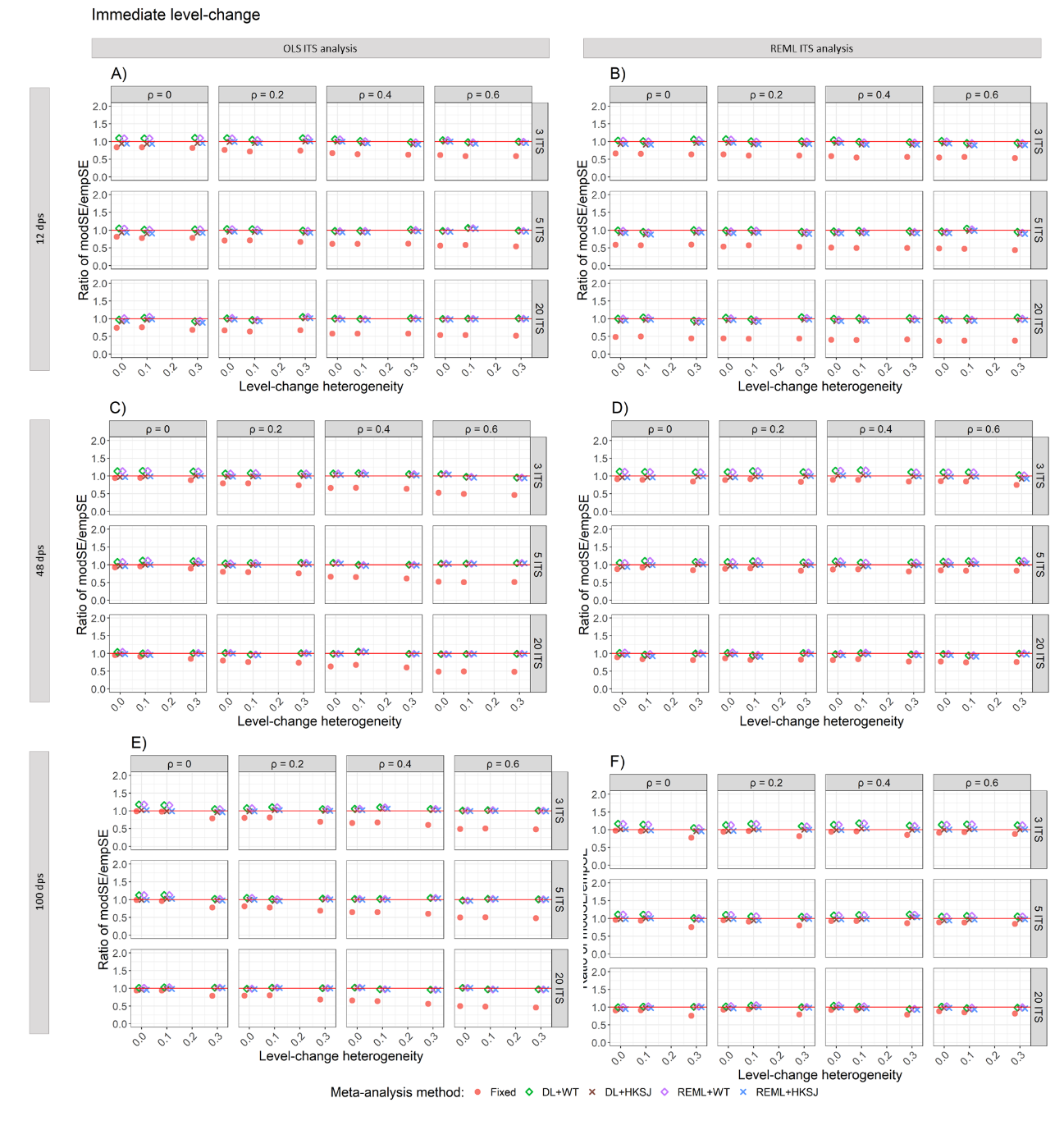

Appendix Figure S18. Plots of the ratio of model based standard error (modSE) to the empirical standard error (empSE)(y-axis) versus level-change heterogeneity (x-axis), when the ITS studies were analysed with OLS (A, C, E) and REML (B, D, F) using fixed-effect (red circles), DL+WT (green diamonds) and DL+HKSJ (brown crosses), REML+WT (purple diamonds) and REML+HKSJ (blue crosses) meta-analysis methods. Plots are presented separately by combinations of: the series length, 12 datapoints (A, B), 48 datapoints (C, D) or 100 datapoints (E, F); the number of included studies (rows) and the level of autocorrelation (columns). The solid red line depicts the nominal 95% coverage level. Simulation settings presented include a level-change of 1, slope-change of 0.1, slope-change heterogeneity of 0, and fixed levels of autocorrelation.

DL, DerSimonian and Laird. dps, datapoints. HKSJ, Hartung-Knapp / Sidik-Jonkman. ITS, interrupted time series. OLS, ordinary least squares. REML, restricted maximum likelihood. WT, Wald-type.

##### 3.5.2 Estimation of slope-change

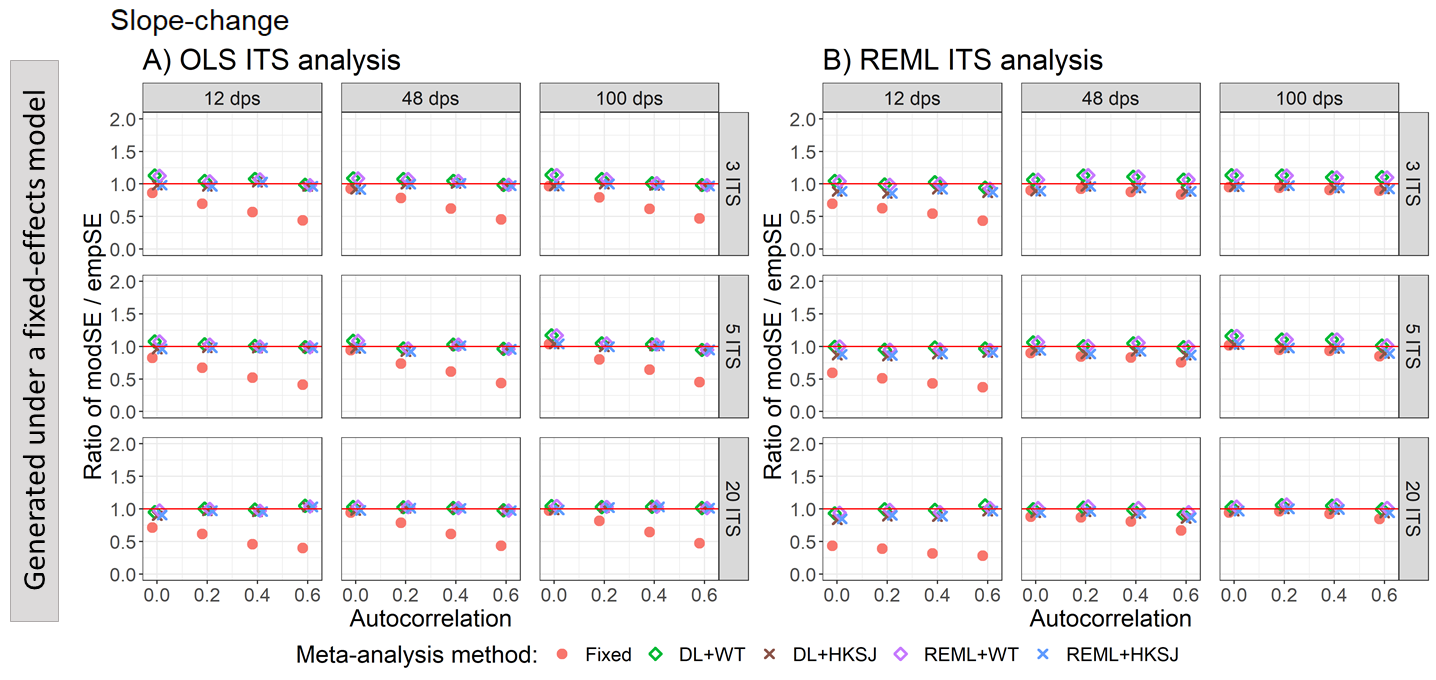

Appendix Figure S19. Plots of the ratio of model-based standard error (modSE) to the empirical standard error (empSE) of slope change (y-axis) when the data was generated under a fixed-effect model and the ITS studies were analysed with OLS (A) and REML (B) using fixed-effect (red circles), DL+WT (green diamonds) and REML+HKSJ (blue crosses) meta-analysis methods versus autocorrelation (x-axis). Plots are presented separately by the number of included studies (rows) and number of datapoints (columns). The solid red line depicts a ratio of one, where the model-based standard error and empirical standard error are equal and thus that the model-based standard error accurately estimates the true standard error. Simulation scenarios include a level-change of 1, level-change heterogeneity of 0, slope-change of 0.1, slope-change heterogeneity of 0, and fixed autocorrelation.

DL, DerSimonian and Laird. dps, datapoints. HKSJ, Hartung-Knapp / Sidik-Jonkman. ITS, interrupted time series. OLS, ordinary least squares. REML, restricted maximum likelihood. WT, Wald-type.

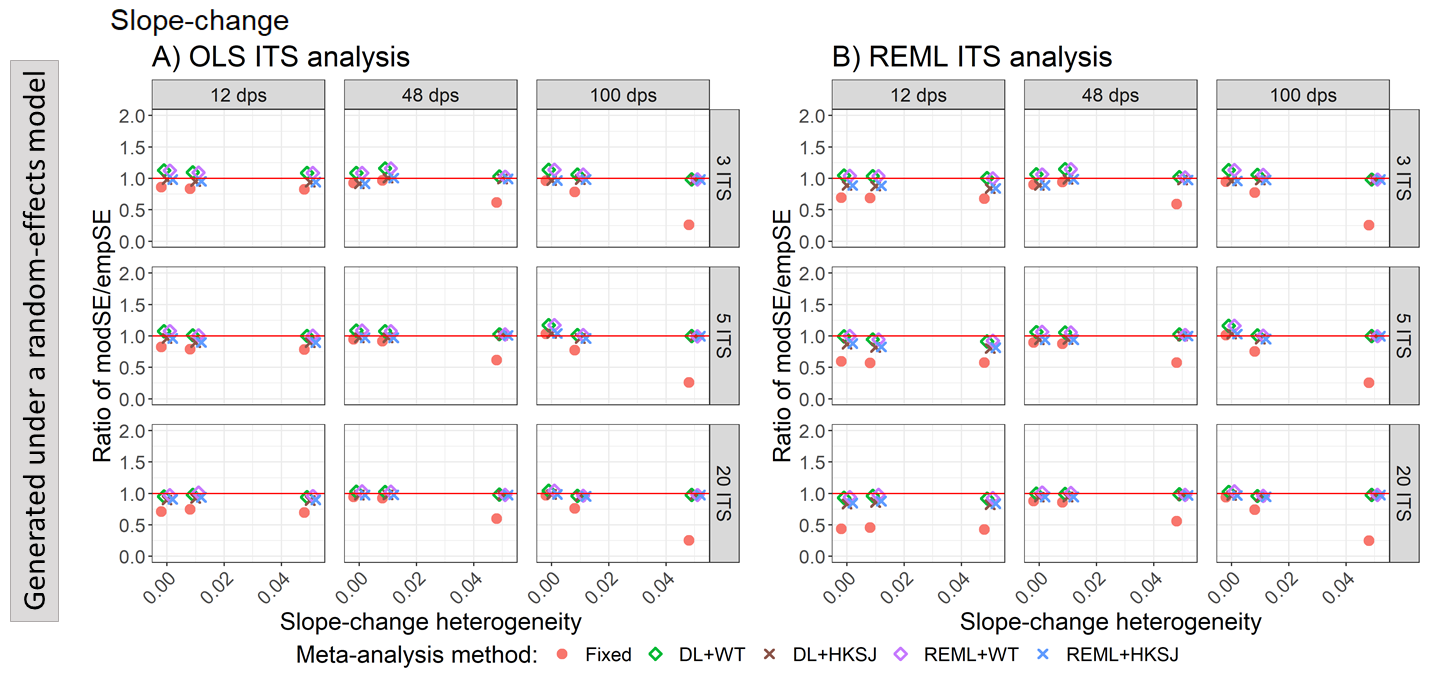

Appendix Figure S20. Plots of the ratio of model based standard error (modSE) to the empirical standard error (empSE) of slope-change (y-axis) when the ITS studies were analysed with OLS (A) and REML (B) using fixed-effect (red circles), DL+WT (green diamonds) and REML+HKSJ (blue crosses) meta-analysis methods versus slope-change heterogeneity (x-axis). Plots are presented separately by combinations of the number of included studies (rows) and number of datapoints (columns). The solid red line depicts a ratio of one, where the model-based standard error and empirical standard error are equal and thus that the model-based standard error accurately estimates the true standard error. Simulation scenarios include a level-change of 1, slope-change of 0.1, slope-change heterogeneity of 0, and autocorrelation of 0.

DL, DerSimonian and Laird. dps, datapoints. HKSJ, Hartung-Knapp / Sidik-Jonkman. ITS, interrupted time series. OLS, ordinary least squares. REML, restricted maximum likelihood. WT, Wald-type.

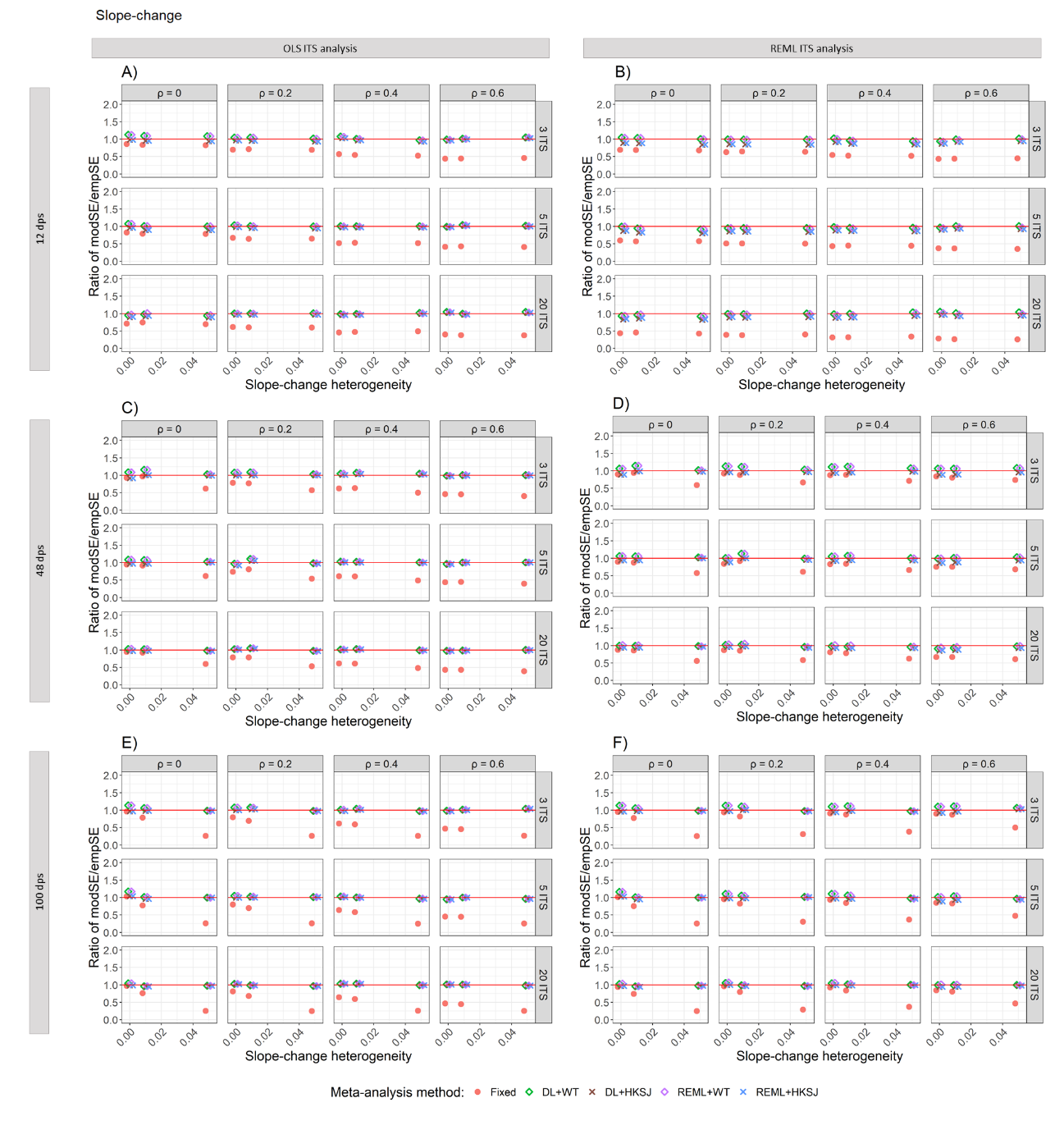

Appendix Figure S21. Plots of the ratio of the empirical standard error (empSE) to the model based standard error (modSE) of the meta-analytic slope-change (y-axis) versus slope-change heterogeneity (x-axis), when the ITS studies were analysed with OLS (A, C, E) and REML (B, D, F) using fixed-effect (red circles), DL+WT (green diamonds) and DL+HKSJ (brown crosses), REML+WT (purple diamonds) and REML+HKSJ (blue crosses) meta-analysis methods. Plots are presented separately by combinations of: the series length, 12 datapoints (A, B), 48 datapoints (C, D) or 100 datapoints (E, F); the number of included studies (rows) and the level of autocorrelation (columns). The solid red line depicts the nominal 95% coverage level. Simulation settings presented include a level-change of 1, level-change heterogeneity of 0, slope-change of 0.1 and fixed levels of autocorrelation.

DL, DerSimonian and Laird. dps, datapoints. HKSJ, Hartung-Knapp / Sidik-Jonkman. ITS, interrupted time series. OLS, ordinary least squares. REML, restricted maximum likelihood. WT, Wald-type.

#### 3.6 Statistical power

##### 3.6.1 Estimation of level-change

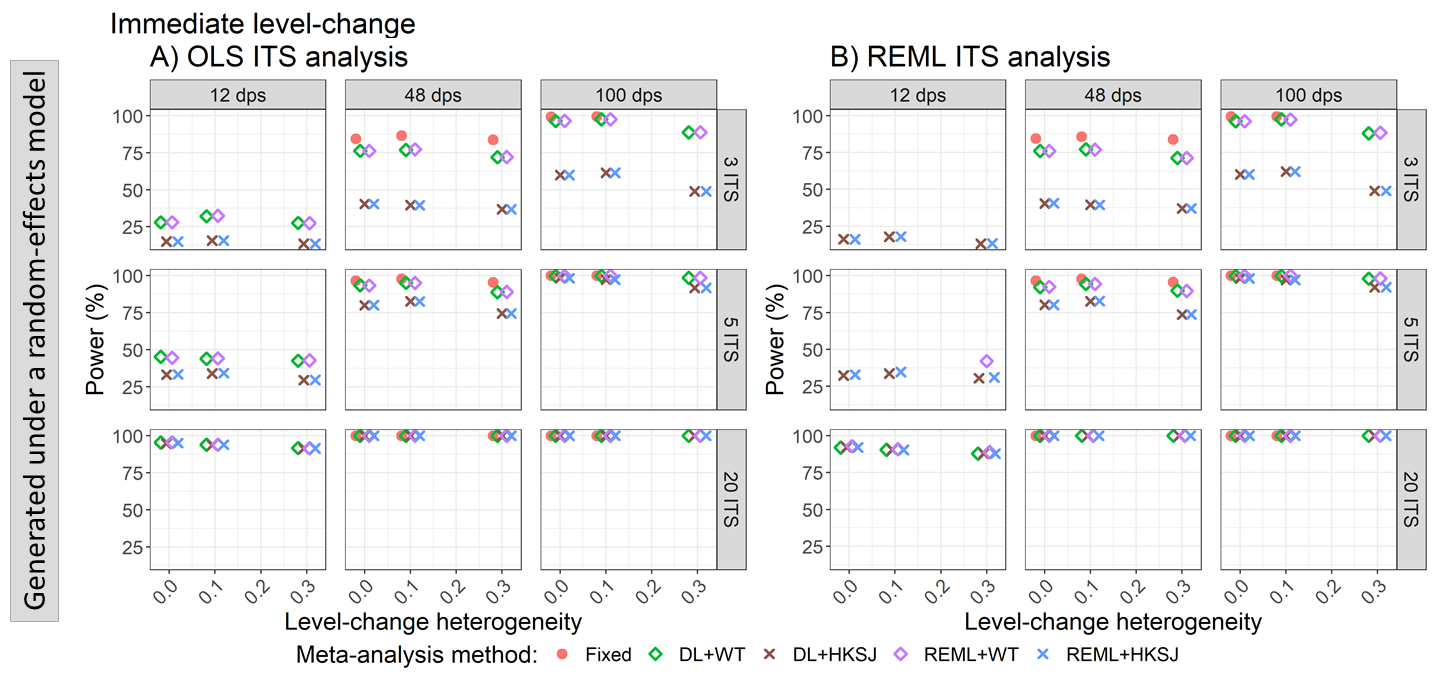

Appendix Figure S22. Plots of statistical power (the percentage of simulations that have a 95% confidence interval that did not include zero – only scenarios with a confidence interval coverage of greater than 90% are plotted) of the meta-analytic immediate level-change (y-axis) when the ITS studies were analysed with OLS (A) and REML (B) using the fixed effect (red circles), DL+WT (green diamonds) and REML+HKSJ (blue crosses) meta-analysis methods versus level-change heterogeneity (x-axis), by number of datapoints (horizontal facets) and number of included studies (vertical facets). Simulation settings include a level-change of 1, slope-change of 0.1, slope-change heterogeneity of 0, and fixed levels of autocorrelation.

DL, DerSimonian and Laird. dps, datapoints. HKSJ, Hartung-Knapp / Sidik-Jonkman. ITS, interrupted time series. OLS, ordinary least squares. REML, restricted maximum likelihood. WT, Wald-type.

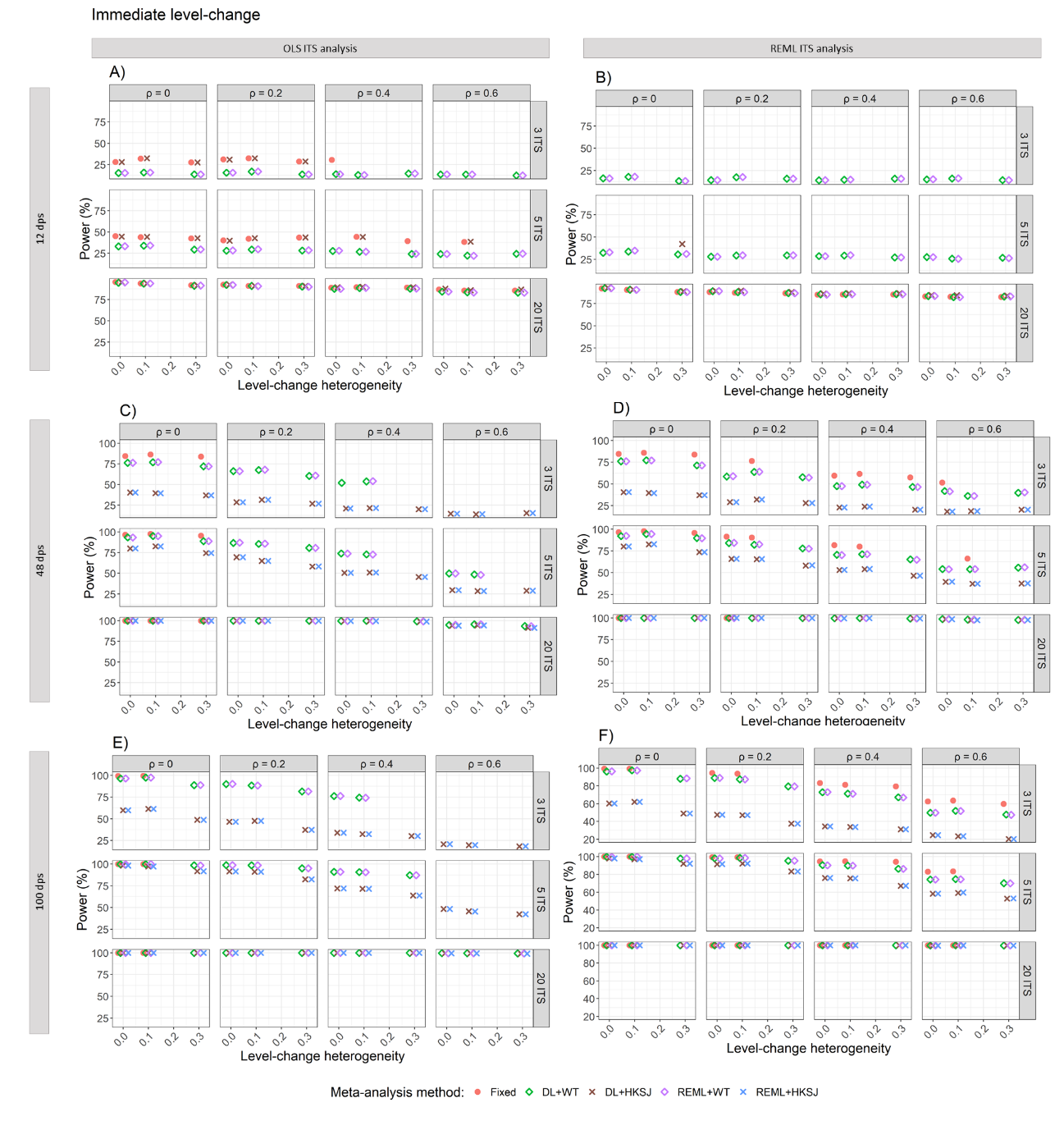

Appendix Figure S23. Plots of statistical power (the percentage of simulations that have a 95% confidence interval that did not include zero – only scenarios with a confidence interval coverage of greater than 90% are plotted) of the meta-analytic immediate level-change (y-axis) versus level-change heterogeneity (x-axis), when the ITS studies were analysed with OLS (A, C, E) and REML (B, D, F) using fixed-effect (red circles), DL+WT (green diamonds) and DL+HKSJ (brown crosses), REML+WT (purple diamonds) and REML+HKSJ (blue crosses) meta-analysis methods. Plots are presented separately by combinations of: the series length, 12 datapoints (A, B), 48 datapoints (C, D) or 100 datapoints (E, F); the number of included studies (rows) and the level of autocorrelation (columns). The solid red line depicts the nominal 95% coverage level. Simulation settings presented include a level-change of 1, slope-change of 0.1, slope-change heterogeneity of 0, and fixed levels of autocorrelation.

DL, DerSimonian and Laird. dps, datapoints. HKSJ, Hartung-Knapp / Sidik-Jonkman. ITS, interrupted time series. OLS, ordinary least squares. REML, restricted maximum likelihood. WT, Wald-type.

##### 3.6.2 Estimation of slope-change

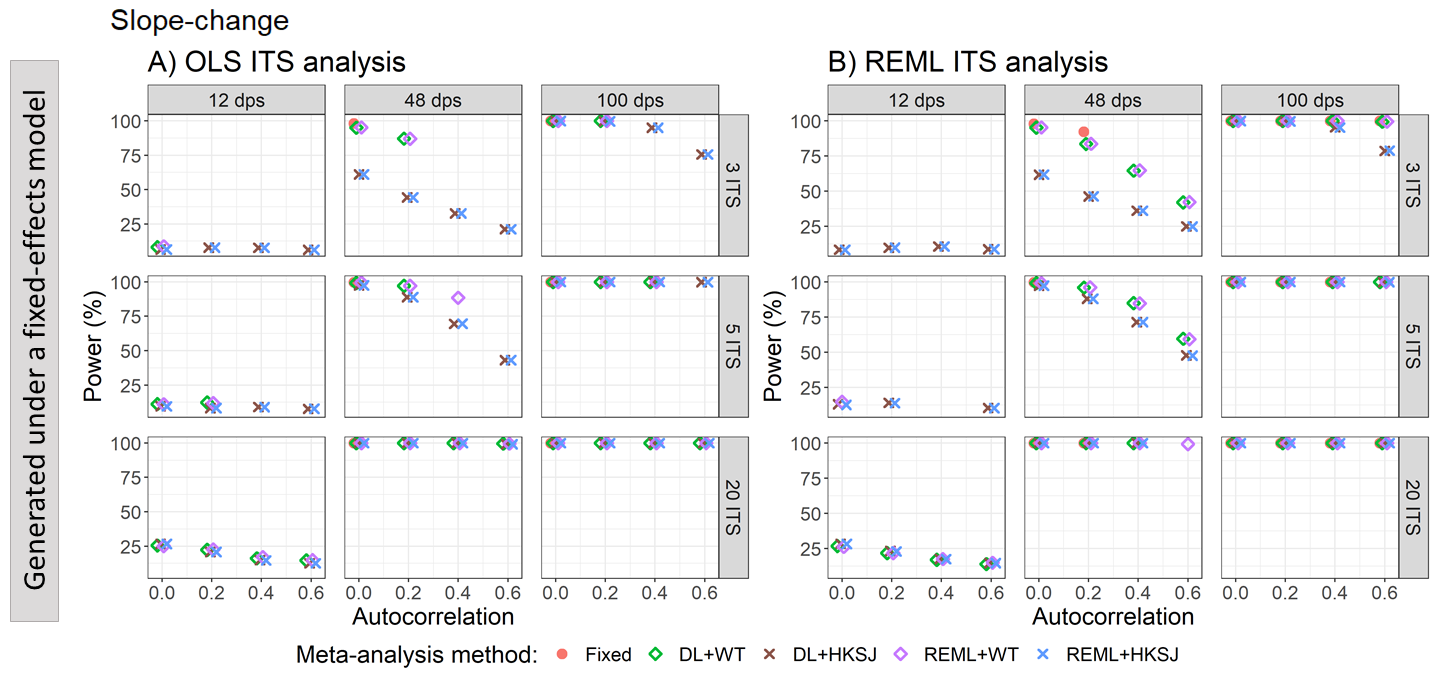

Appendix Figure S24. Plots of statistical power (the percentage of simulations that have a 95% confidence interval that did not include zero – only scenarios with a confidence interval coverage of greater than 90% are plotted) of the meta-analytic slope-change (y-axis) when the data was generated under a fixed-effect model and the ITS studies were analysed with OLS (A) and REML (B) using the fixed effect (red circles), DL+WT (green diamonds) and DL+HKSJ (brown crosses), REML+WT (purple diamonds) and REML+HKSJ (blue crosses) meta-analysis methods versus autocorrelation (x-axis). Plots are presented separately by combinations of the number of included studies (rows) and the number of datapoints (columns). Simulation scenarios include a level-change of 1, level-change heterogeneity of 0, slope-change of 0.1, slope-change heterogeneity of 0, and fixed levels of autocorrelation.

DL, DerSimonian and Laird. dps, datapoints. HKSJ, Hartung-Knapp / Sidik-Jonkman. ITS, interrupted time series. OLS, ordinary least squares. REML, restricted maximum likelihood. WT, Wald-type.

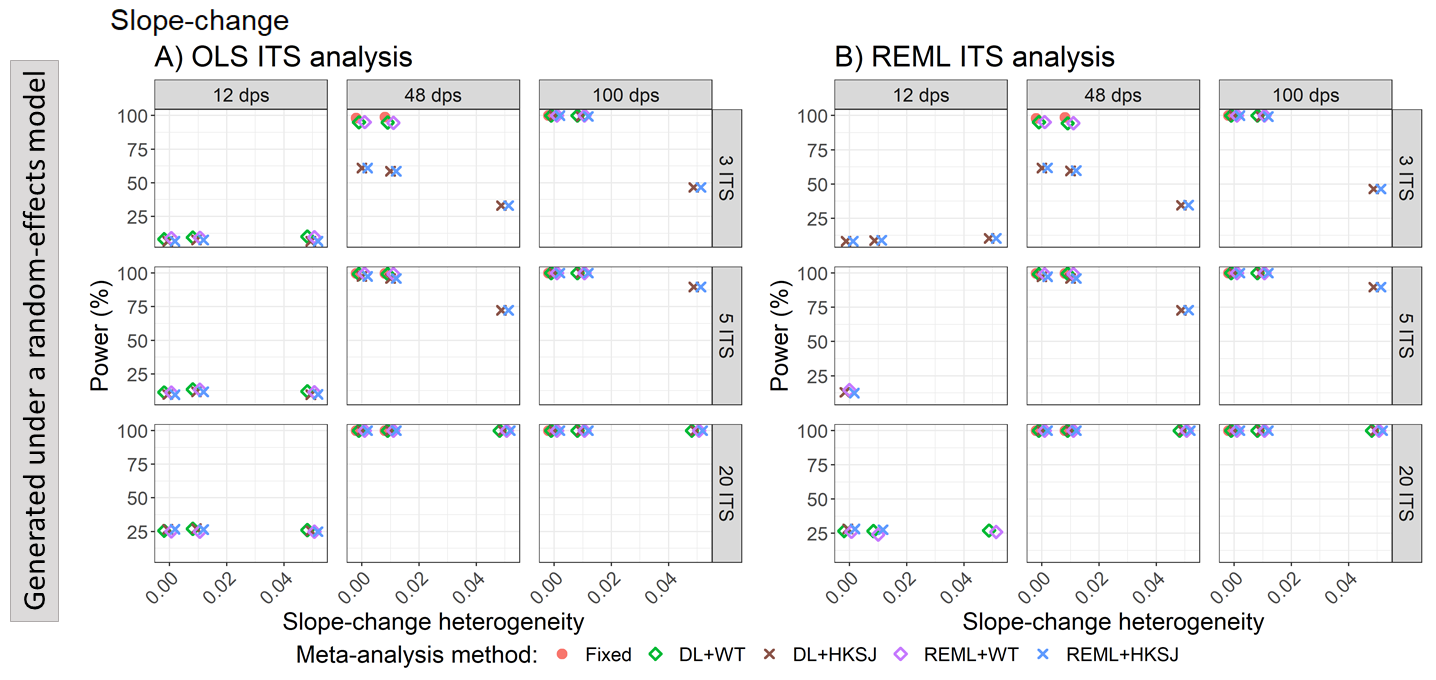

Appendix Figure S25. Plots of statistical power (the percentage of simulations that have a 95% confidence interval that did not include zero – only scenarios with a confidence interval coverage of greater than 90% are plotted) of the meta-analytic slope-change (y-axis) when the ITS are analysed with OLS (A) and REML (B) for the fixed effect (red circles), DL+WT (green diamonds) and DL+HKSJ (brown crosses), REML+WT (purple diamonds) and REML+HKSJ (blue crosses) meta-analysis methods versus slope-change heterogeneity (x-axis), by number of datapoints (horizontal facets) and number of included studies (vertical facets). Simulation settings include a level-change of 1, level-change heterogeneity of 0, slope-change of 0.1, and autocorrelation of 0.

DL, DerSimonian and Laird. dps, datapoints. HKSJ, Hartung-Knapp / Sidik-Jonkman. ITS, interrupted time series. OLS, ordinary least squares. REML, restricted maximum likelihood. WT, Wald-type.

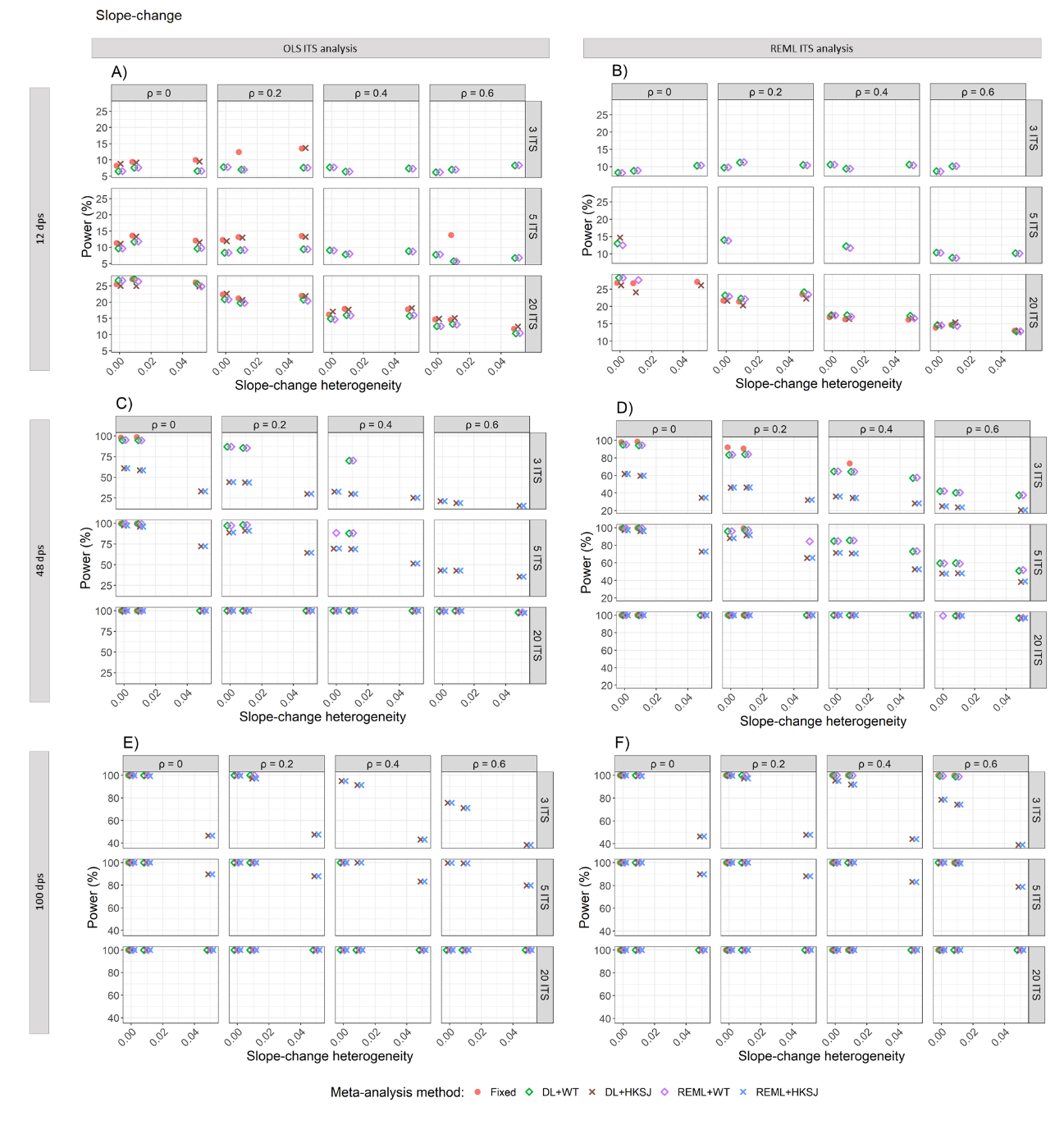

Appendix Figure S26. Plots of statistical power (the percentage of simulations that have a 95% confidence interval that did not include zero – only scenarios with a confidence interval coverage of greater than 90% are plotted) of the meta-analytic slope-change (y-axis) versus slope-change heterogeneity (x-axis), when the ITS studies were analysed with OLS (A, C, E) and REML (B, D, F) using fixed-effect (red circles), DL+WT (green diamonds) and DL+HKSJ (brown crosses), REML+WT (purple diamonds) and REML+HKSJ (blue crosses) meta-analysis methods. Plots are presented separately by combinations of: the series length, 12 datapoints (A, B), 48 datapoints (C, D) or 100 datapoints (E, F); the number of included studies (rows) and the level of autocorrelation (columns). The solid red line depicts the nominal 95% coverage level. Simulation settings presented include a level-change of 1, level-change heterogeneity of 0, slope-change of 0.1 and fixed levels of autocorrelation.

DL, DerSimonian and Laird. dps, datapoints. HKSJ, Hartung-Knapp / Sidik-Jonkman. ITS, interrupted time series. OLS, ordinary least squares. REML, restricted maximum likelihood. WT, Wald-type.

#### 3.7 Autocorrelation variation

Performance was not impacted by variability in autocorrelation with a meta-analysis, and this was not modified by the number of datapoints, number of studies or between-study variance in the level- or slope-change.

##### 3.7.1 Bias

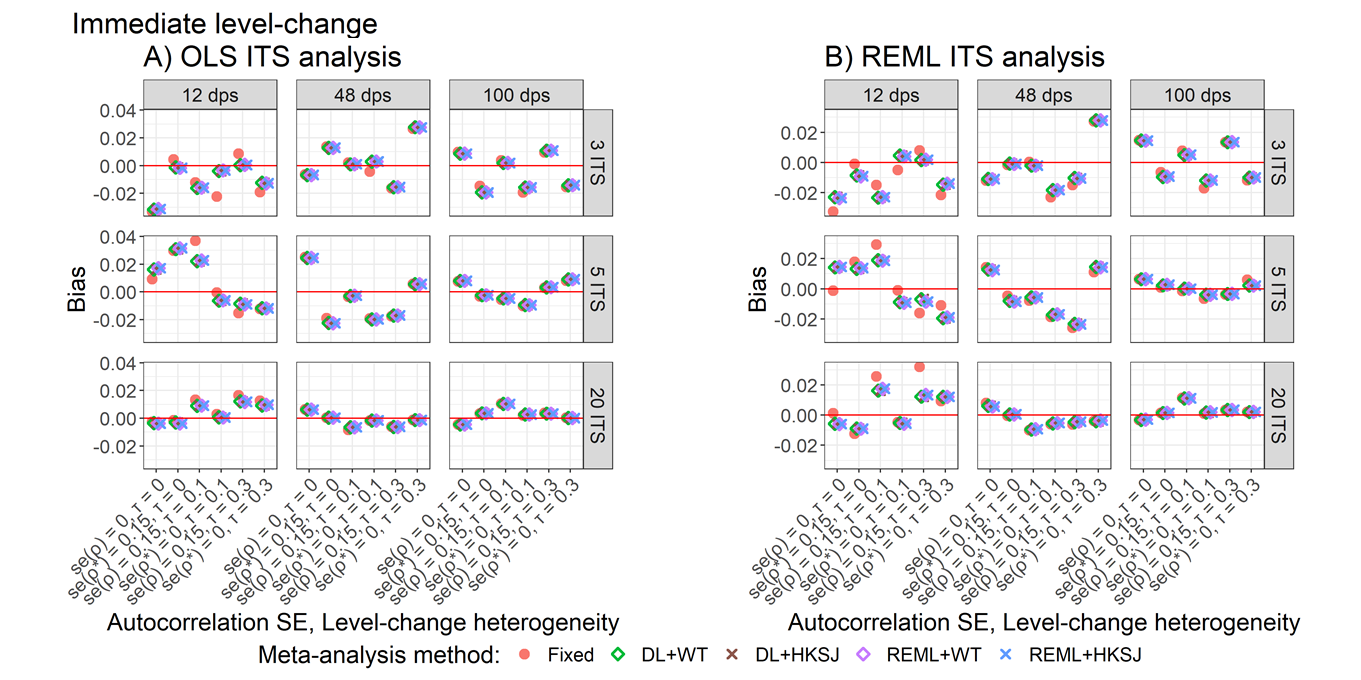

Appendix Figure S27. Plots of bias of immediate level-change (y-axis) when the ITS are analysed with OLS (A) and REML (B) using the fixed effect (red circles), DL+WT (green diamonds) and DL+HKSJ (brown crosses), REML+WT (purple diamonds) and REML+HKSJ (blue crosses) meta-analysis methods versus categories of autocorrelation variability and level-change heterogeneity combinations (x-axis). Plots are presented separately by combinations of the number of included studies (rows) and the number of datapoints (columns). Simulation scenarios include a level-change of 1, level-change heterogeneity of 0, slope-change of 0.1, autocorrelation of 0.4.

DL, DerSimonian and Laird. dps, datapoints. HKSJ, Hartung-Knapp / Sidik-Jonkman. ITS, interrupted time series. OLS, ordinary least squares. REML, restricted maximum likelihood. WT, Wald-type.

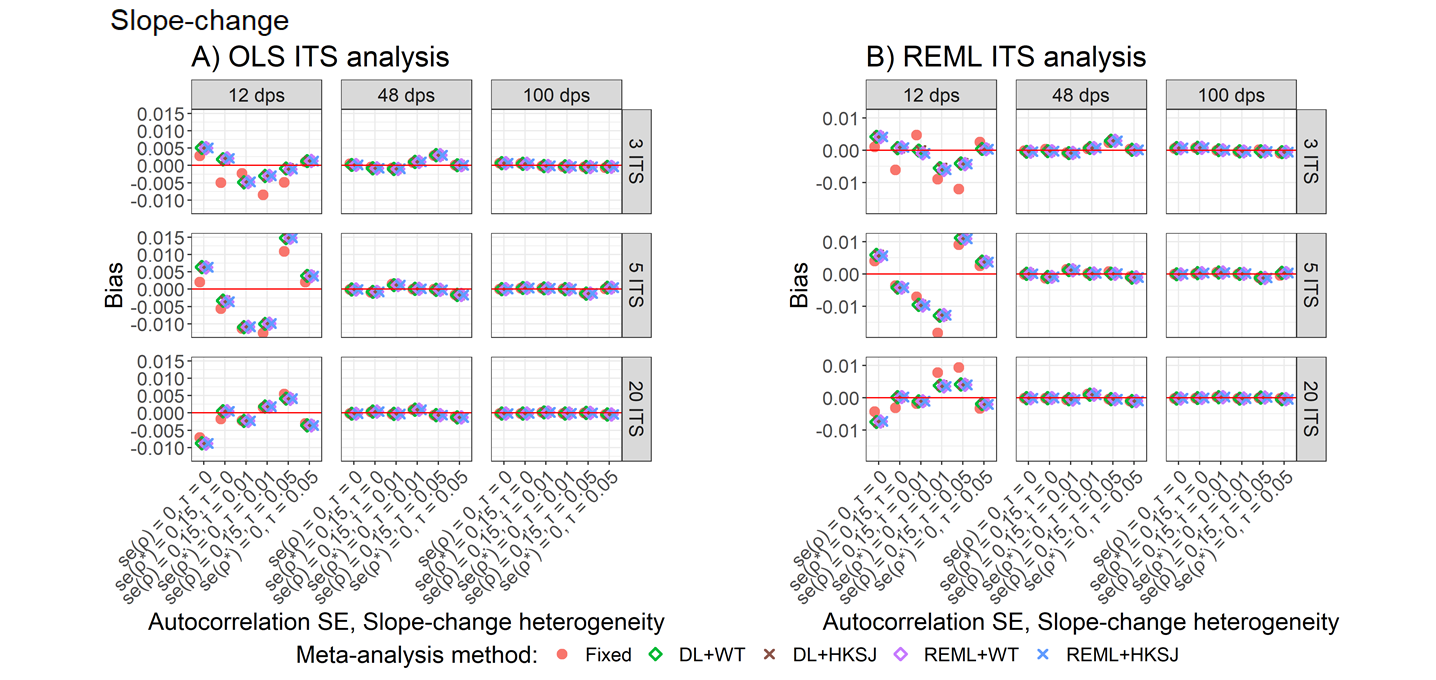

Appendix Figure S28. Plot of bias of slope-change (y-axis) when the ITS are analysed with OLS (A) and REML (B) for the fixed effect (red circles), DL+WT (green diamonds) and DL+HKSJ (brown crosses), REML+WT (purple diamonds) and REML+HKSJ (blue crosses) meta-analysis methods versus categories of autocorrelation variability and slope-change heterogeneity combinations (x-axis). Plots are presented separately by combinations of the number of included studies (rows) and the number of datapoints (columns). Simulation scenarios include a level-change of 1, level-change heterogeneity of 0, slope-change of 0.1, autocorrelation of 0.4.

DL, DerSimonian and Laird. dps, datapoints. HKSJ, Hartung-Knapp / Sidik-Jonkman. ITS, interrupted time series. OLS, ordinary least squares. REML, restricted maximum likelihood. WT, Wald-type.

##### 3.7.2 Coverage

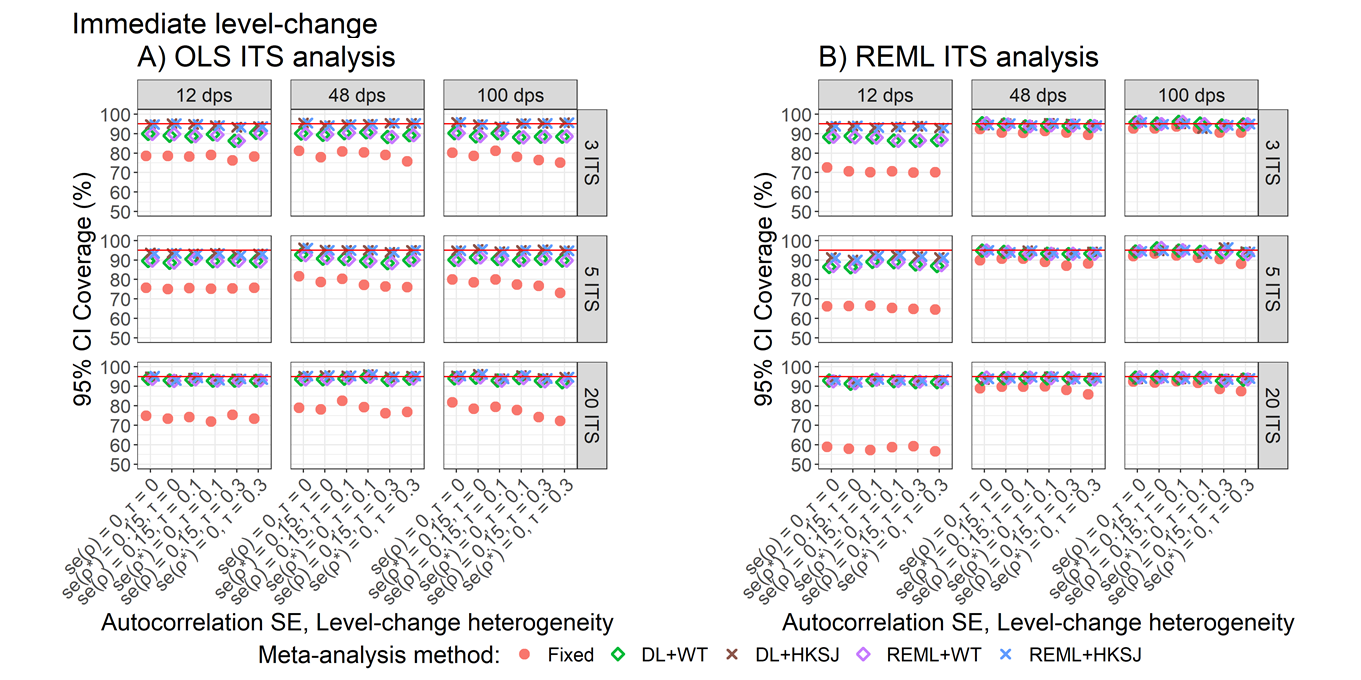

Appendix Figure S29. Plots of 95% confidence interval coverage of immediate level-change (y-axis) when the ITS are analysed with OLS (A) and REML (B) using the fixed effect (red circles), DL+WT (green diamonds) and DL+HKSJ (brown crosses), REML+WT (purple diamonds) and REML+HKSJ (blue crosses) meta-analysis methods versus categories of autocorrelation variability and level-change heterogeneity combinations (x-axis). Plots are presented separately by combinations of the number of included studies (rows) and the number of datapoints (columns). Simulation scenarios include a level-change of 1, level-change heterogeneity of 0, slope-change of 0.1, autocorrelation of 0.4.

DL, DerSimonian and Laird. dps, datapoints. HKSJ, Hartung-Knapp / Sidik-Jonkman. ITS, interrupted time series. OLS, ordinary least squares. REML, restricted maximum likelihood. WT, Wald-type.

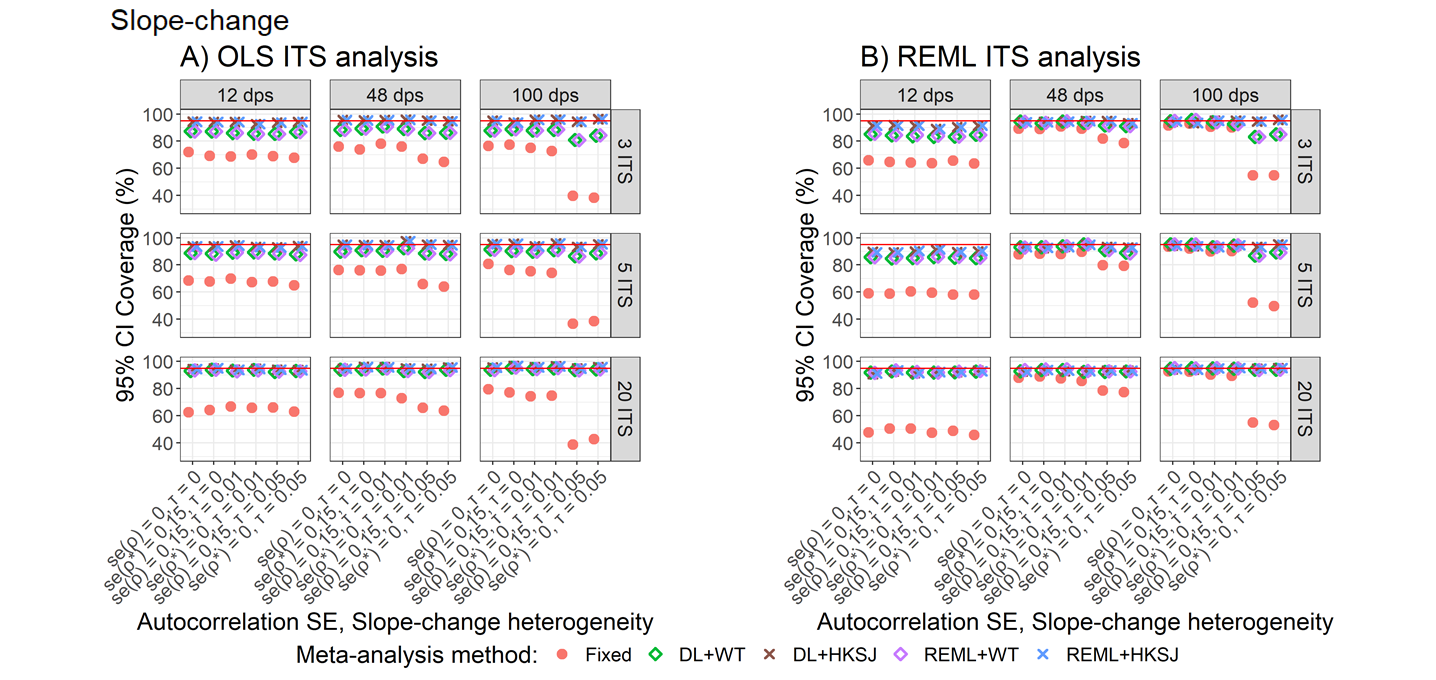

Appendix Figure S30. The 95% confidence interval coverage of slope-change (y-axis) when the ITS are analysed with OLS (A) and REML (B) for the fixed effect (red circles), DL+WT (green diamonds) and DL+HKSJ (brown crosses), REML+WT (purple diamonds) and REML+HKSJ (blue crosses) meta-analysis methods versus categories of autocorrelation variability and slope-change heterogeneity combinations (x-axis). Plots are presented separately by combinations of the number of included studies (rows) and the number of datapoints (columns). Simulation scenarios include a level-change of 1, level-change heterogeneity of 0, slope-change of 0.1, autocorrelation of 0.4.

DL, DerSimonian and Laird. dps, datapoints. HKSJ, Hartung-Knapp / Sidik-Jonkman. ITS, interrupted time series. OLS, ordinary least squares. REML, restricted maximum likelihood. WT, Wald-type.

##### 3.7.3 Empirical standard errors

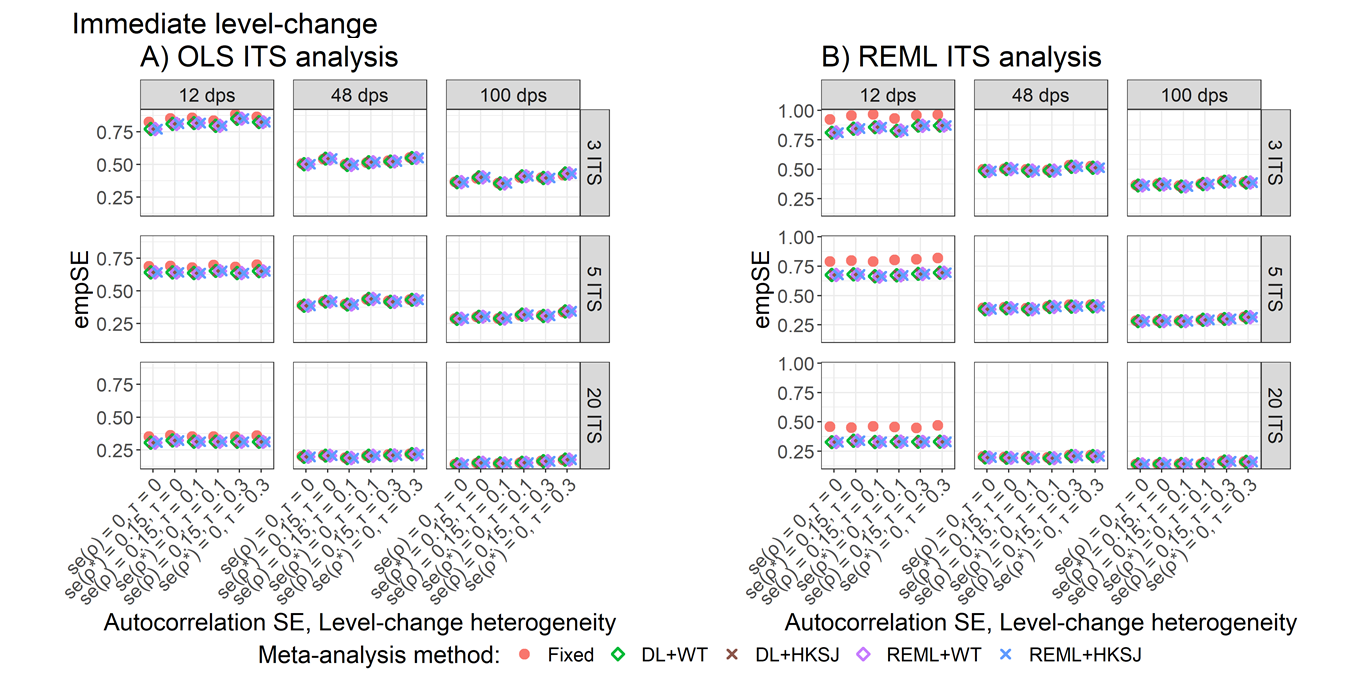

Appendix Figure S31. Plots of empirical standard error (empSE) of the immediate level-change (y-axis) when the ITS are analysed with OLS (A) and REML (B) for the fixed effect (red circles), DL+WT (green diamonds) and DL+HKSJ (brown crosses), REML+WT (purple diamonds) and REML+HKSJ (blue crosses) meta-analysis methods versus categories of autocorrelation variability and level-change heterogeneity combinations (x-axis). Plots are presented separately by combinations of the number of included studies (rows) and the number of datapoints (columns). Simulation scenarios include a level-change of 1, level-change heterogeneity of 0, slope-change of 0.1, autocorrelation of 0.4.

DL, DerSimonian and Laird. dps, datapoints. HKSJ, Hartung-Knapp / Sidik-Jonkman. ITS, interrupted time series. OLS, ordinary least squares. REML, restricted maximum likelihood. WT, Wald-type.

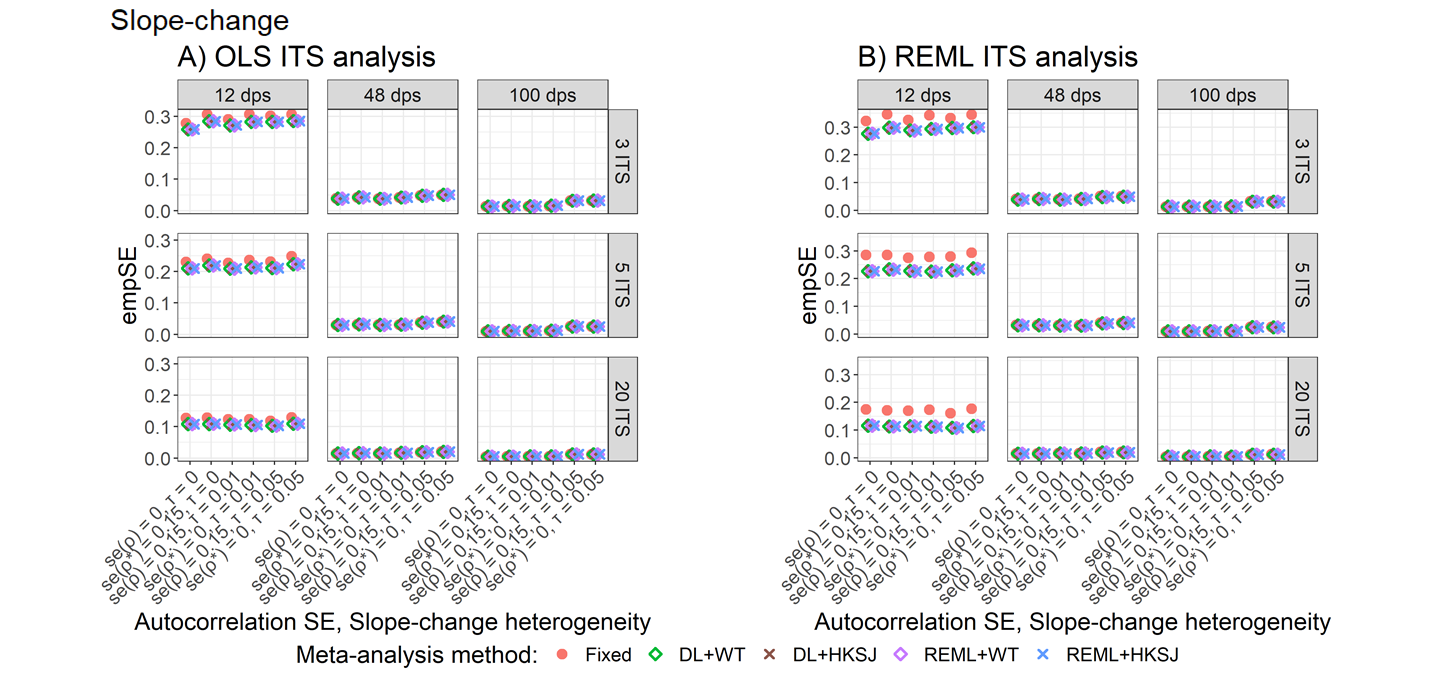

Appendix Figure S32. Plots of empirical standard error (empSE) of the slope-change (y-axis) when the ITS are analysed with OLS (A) and REML (B) for the fixed effect (red circles), DL+WT (green diamonds) and DL+HKSJ (brown crosses), REML+WT (purple diamonds) and REML+HKSJ (blue crosses) meta-analysis methods versus categories of autocorrelation variability and slope-change heterogeneity combinations (x-axis). Plots are presented separately by combinations of the number of included studies (rows) and the number of datapoints (columns). Simulation scenarios include a level-change of 1, level-change heterogeneity of 0, slope-change of 0.1, autocorrelation of 0.4.

DL, DerSimonian and Laird. dps, datapoints. HKSJ, Hartung-Knapp / Sidik-Jonkman. ITS, interrupted time series. OLS, ordinary least squares. REML, restricted maximum likelihood. WT, Wald-type.

##### 3.7.4 Ratio of model-based standard errors to empirical standard errors

The ratio of model-based standard errors to empirical standard errors were not impacted by variability in autocorrelation with a meta-analysis, and this was not modified by the number of datapoints, number of studies or between-study variance in the level- or slope-change.

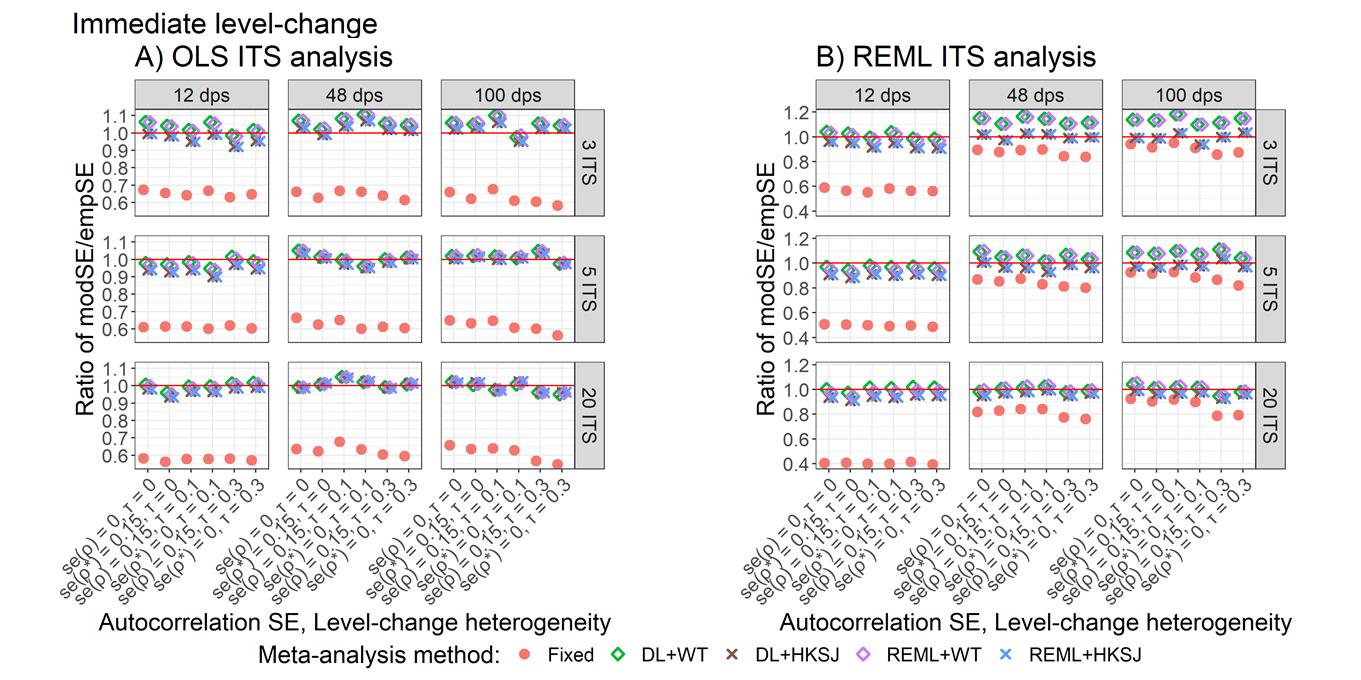

Appendix Figure S33. Plots of the ratio of the empirical standard error (empSE) to the model based standard error (modSE) of the meta-analytic immediate level-change (y-axis) when the ITS are analysed with OLS (A) and REML (B) for the fixed effect (red circles), DL+WT (green diamonds) and DL+HKSJ (brown crosses), REML+WT (purple diamonds) and REML+HKSJ (blue crosses) meta-analysis methods versus categories of autocorrelation variability and level-change heterogeneity combinations (x-axis). Plots are presented separately by combinations of the number of included studies (rows) and the number of datapoints (columns). Simulation scenarios include a level-change of 1, level-change heterogeneity of 0, slope-change of 0.1, autocorrelation of 0.4.

DL, DerSimonian and Laird. dps, datapoints. HKSJ, Hartung-Knapp / Sidik-Jonkman. ITS, interrupted time series. OLS, ordinary least squares. REML, restricted maximum likelihood. WT, Wald-type.

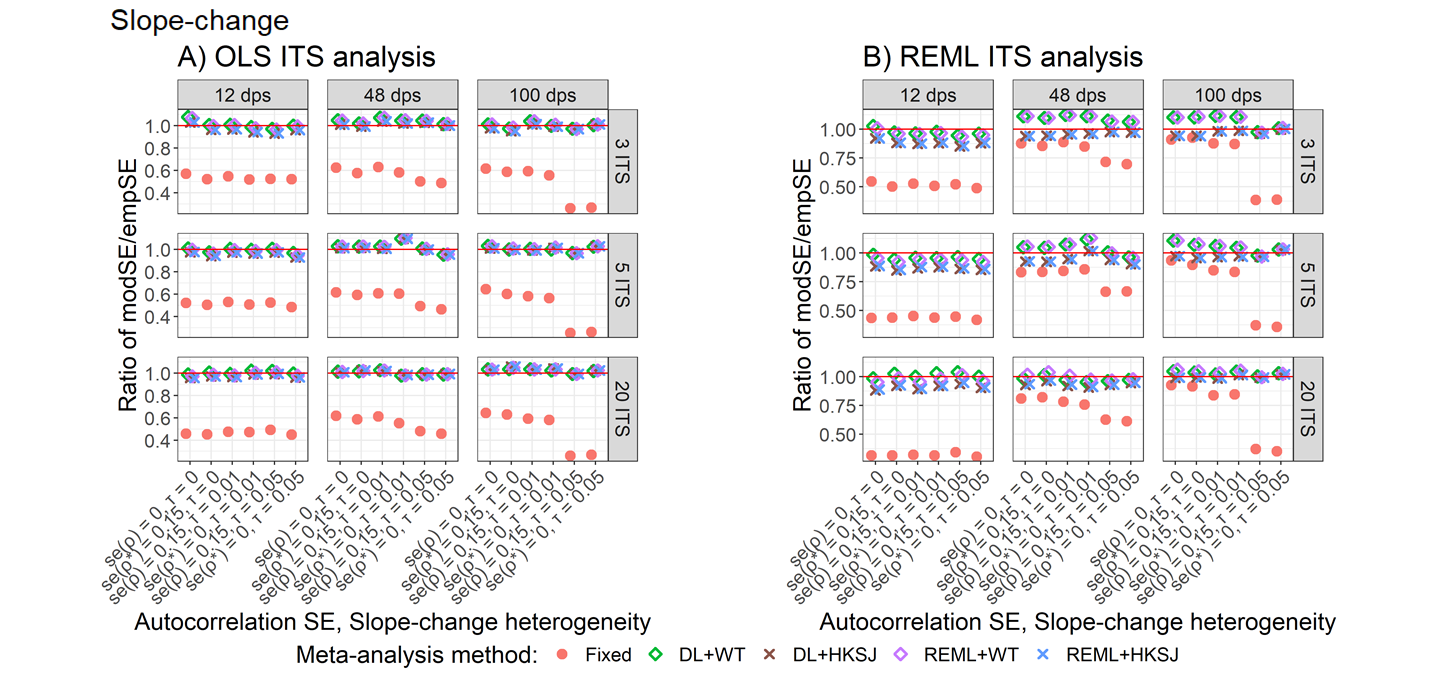

Appendix Figure S34. Plots of the ratio of the empirical standard error (empSE) to the model based standard error (modSE) of the meta-analytic slope-change (y-axis) when the ITS are analysed with OLS (A) and REML (B) for the fixed effect (red circles), DL+WT (green diamonds) and DL+HKSJ (brown crosses), REML+WT (purple diamonds) and REML+HKSJ (blue crosses) meta-analysis methods versus categories of autocorrelation variability and slope-change heterogeneity combinations (x-axis). Plots are presented separately by combinations of the number of included studies (rows) and the number of datapoints (columns). Simulation scenarios include a level-change of 1, level-change heterogeneity of 0, slope-change of 0.1, autocorrelation of 0.4.

DL, DerSimonian and Laird. dps, datapoints. HKSJ, Hartung-Knapp / Sidik-Jonkman. ITS, interrupted time series. OLS, ordinary least squares. REML, restricted maximum likelihood. WT, Wald-type.

##### 3.7.5 Statistical power

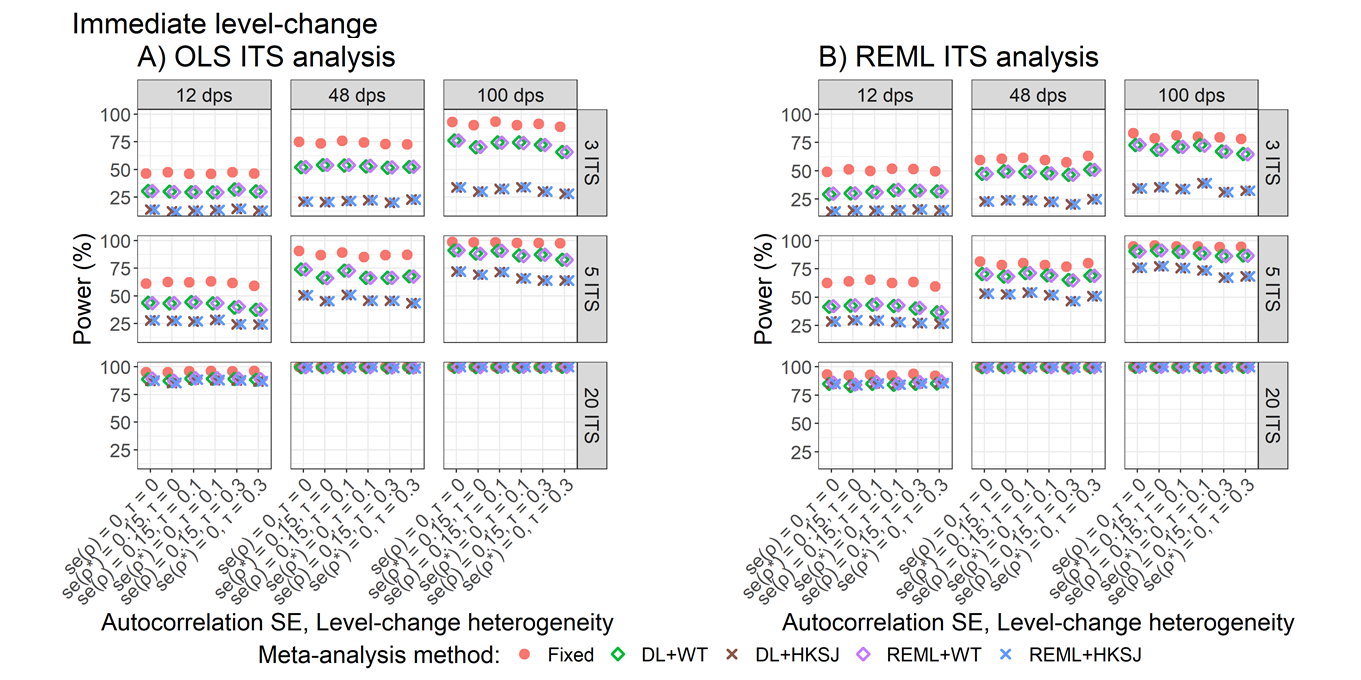

Appendix Figure S35. Plots of statistical power (the percentage of simulations that have a 95% confidence interval that did not include zero – only scenarios with a confidence interval coverage of greater than 90% are plotted) of the meta-analytic level-change (y-axis) when the ITS are analysed with OLS (A) and REML (B) for the fixed effect (red circles), DL+WT (green diamonds) and DL+HKSJ (brown crosses), REML+WT (purple diamonds) and REML+HKSJ (blue crosses) meta-analysis methods versus categories of autocorrelation variability and level-change heterogeneity combinations (x-axis). Plots are presented separately by combinations of the number of included studies (rows) and the number of datapoints (columns). Simulation scenarios include a level-change of 1, level-change heterogeneity of 0, slope-change of 0.1, autocorrelation of 0.4.

DL, DerSimonian and Laird. dps, datapoints. HKSJ, Hartung-Knapp / Sidik-Jonkman. ITS, interrupted time series. OLS, ordinary least squares. REML, restricted maximum likelihood. WT, Wald-type.

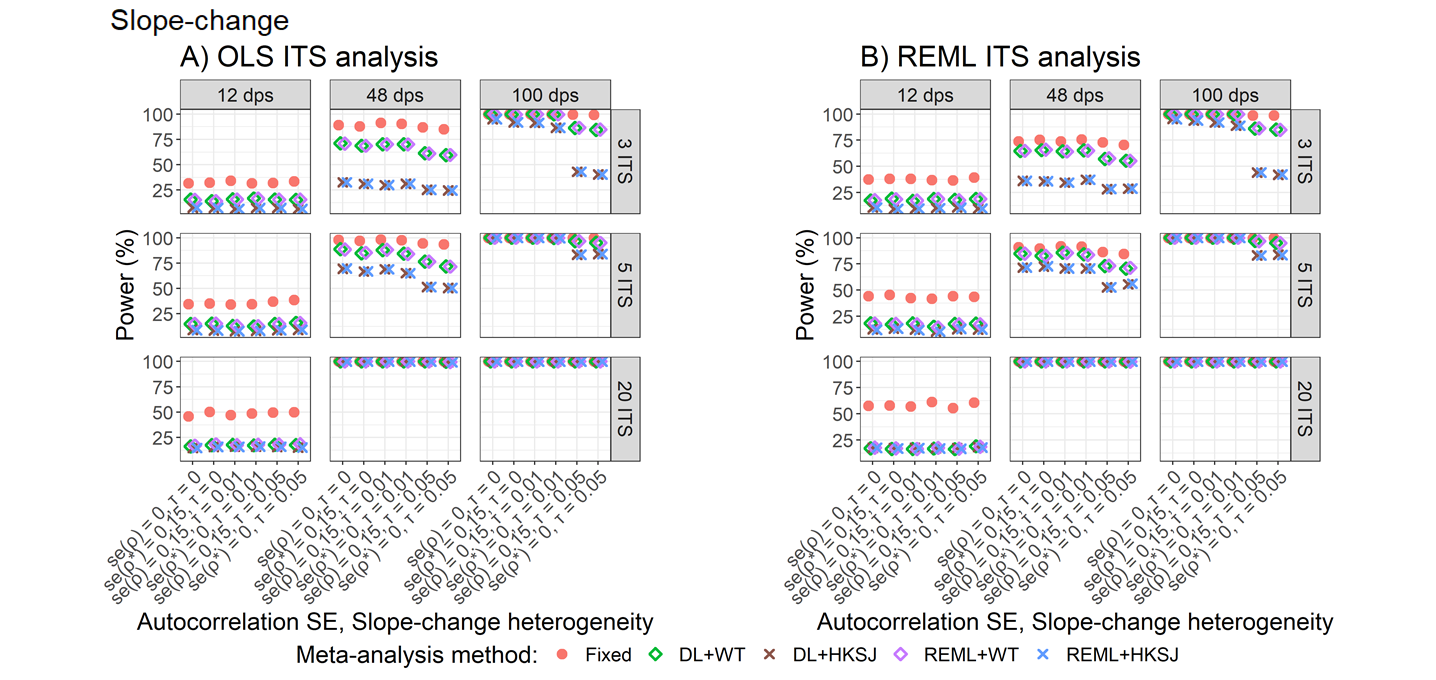

Appendix Figure S36. Plots of statistical power (the percentage of simulations that have a 95% confidence interval that did not include zero – only scenarios with a confidence interval coverage of greater than 90% are plotted) of the meta-analytic slope-change (y-axis) when the ITS are analysed with OLS (A) and REML (B) for the fixed effect (red circles), DL+WT (green diamonds) and DL+HKSJ (brown crosses), REML+WT (purple diamonds) and REML+HKSJ (blue crosses) meta-analysis methods versus categories of autocorrelation variability and slope-change heterogeneity combinations (x-axis). Plots are presented separately by combinations of the number of included studies (rows) and the number of datapoints (columns). Simulation scenarios include a level-change of 1, level-change heterogeneity of 0, slope-change of 0.1, autocorrelation of 0.4.

DL, DerSimonian and Laird. dps, datapoints. HKSJ, Hartung-Knapp / Sidik-Jonkman. ITS, interrupted time series. OLS, ordinary least squares. REML, restricted maximum likelihood. WT, Wald-type.

#### 3.8 Estimated heterogeneity

##### 3.8.1 Level-change heterogeneity

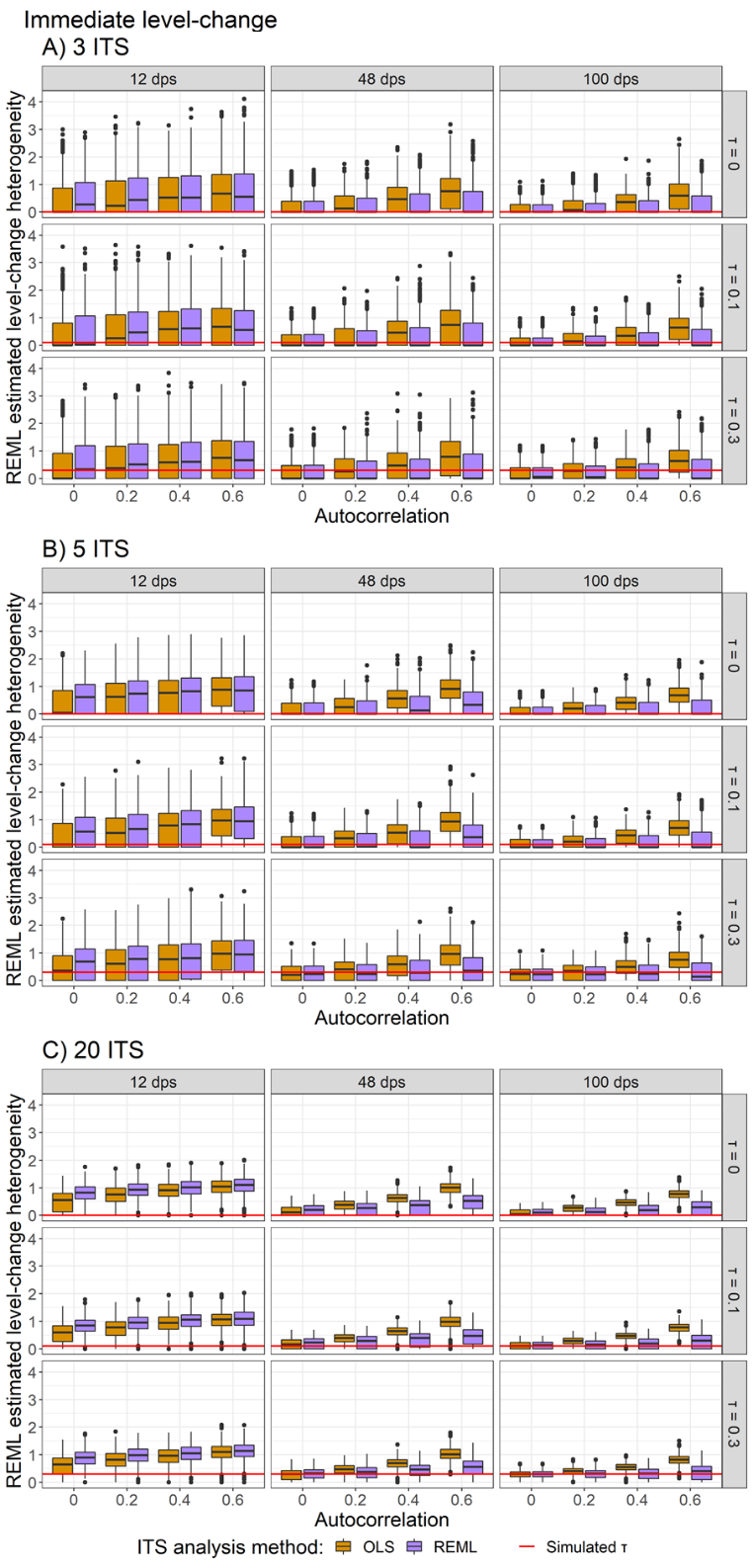

Appendix Figure S37. Plots of slope-change heterogeneity estimated using a random effects meta-analysis with the REML between-study variance estimator (y-axis) when the A) 3 ITS, B) 5 ITS and C) 20 ITS studies were analysed with OLS (orange) and REML (purple) ITS analysis methods. Plots are presented separately by combinations of the true slope-change heterogeneity (rows) and the number of datapoints (columns). The solid red lines indicate the true level-change heterogeneity. Simulation scenarios include a level-change of 1, level-change heterogeneity of 0, slope-change of 0.1, and fixed autocorrelation.

DL, DerSimonian and Laird. dps, datapoints. HKSJ, Hartung-Knapp / Sidik-Jonkman. ITS, interrupted time series. OLS, ordinary least squares. REML, restricted maximum likelihood. WT, Wald-type.

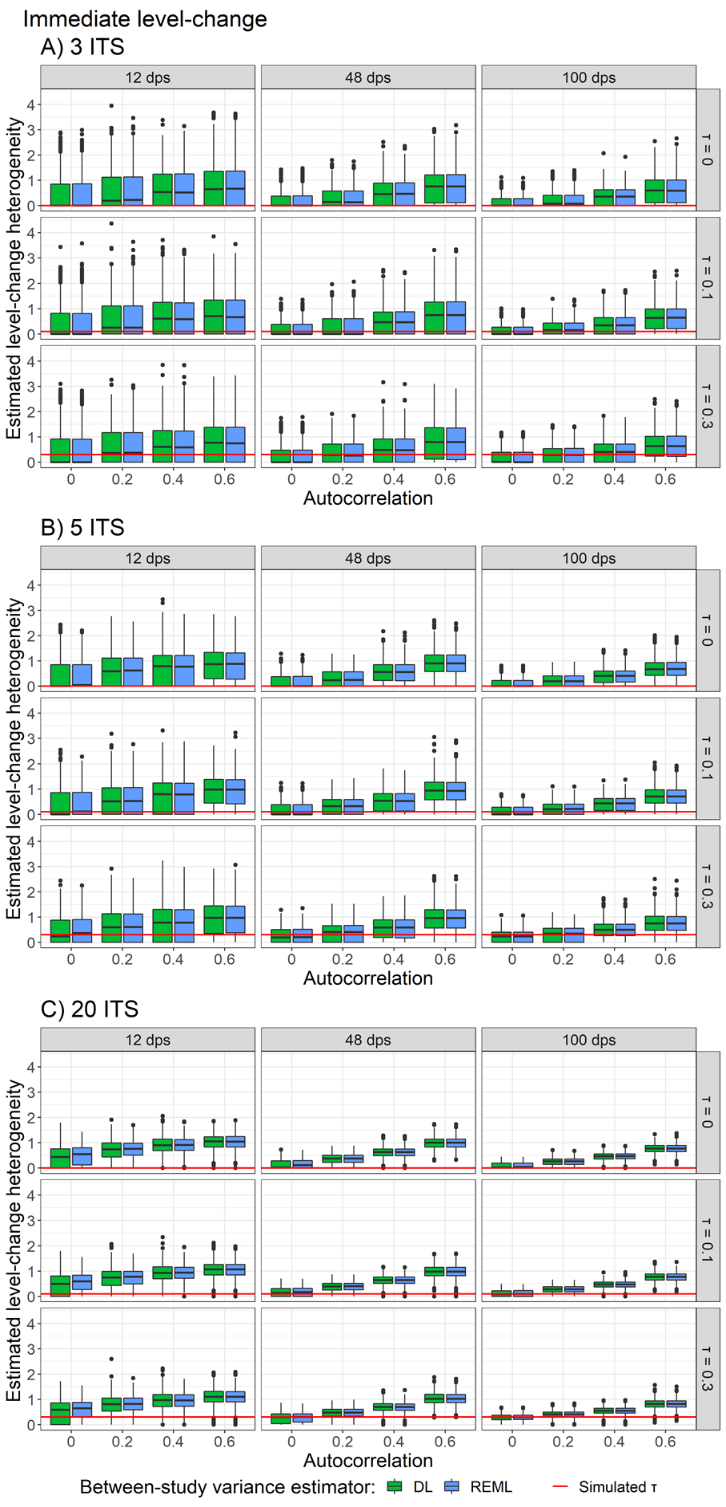

Appendix Figure S38. Plots of level-change heterogeneity estimated using a random effects meta-analysis with DL (green) and REML (purple) between-study variance estimators (y-axis) when the A) 3 ITS, B) 5 ITS and C) 20 ITS studies were analysed with OLS. Plots are presented separately by combinations of the true level-change heterogeneity (rows) and the number of datapoints (columns). The solid red lines indicate the true level-change heterogeneity. Simulation scenarios include a level-change of 1, slope-change of 0.1, slope-change heterogeneity of 0, and fixed autocorrelation.

DL, DerSimonian and Laird. dps, datapoints. HKSJ, Hartung-Knapp / Sidik-Jonkman. ITS, interrupted time series. OLS, ordinary least squares. REML, restricted maximum likelihood. WT, Wald-type.

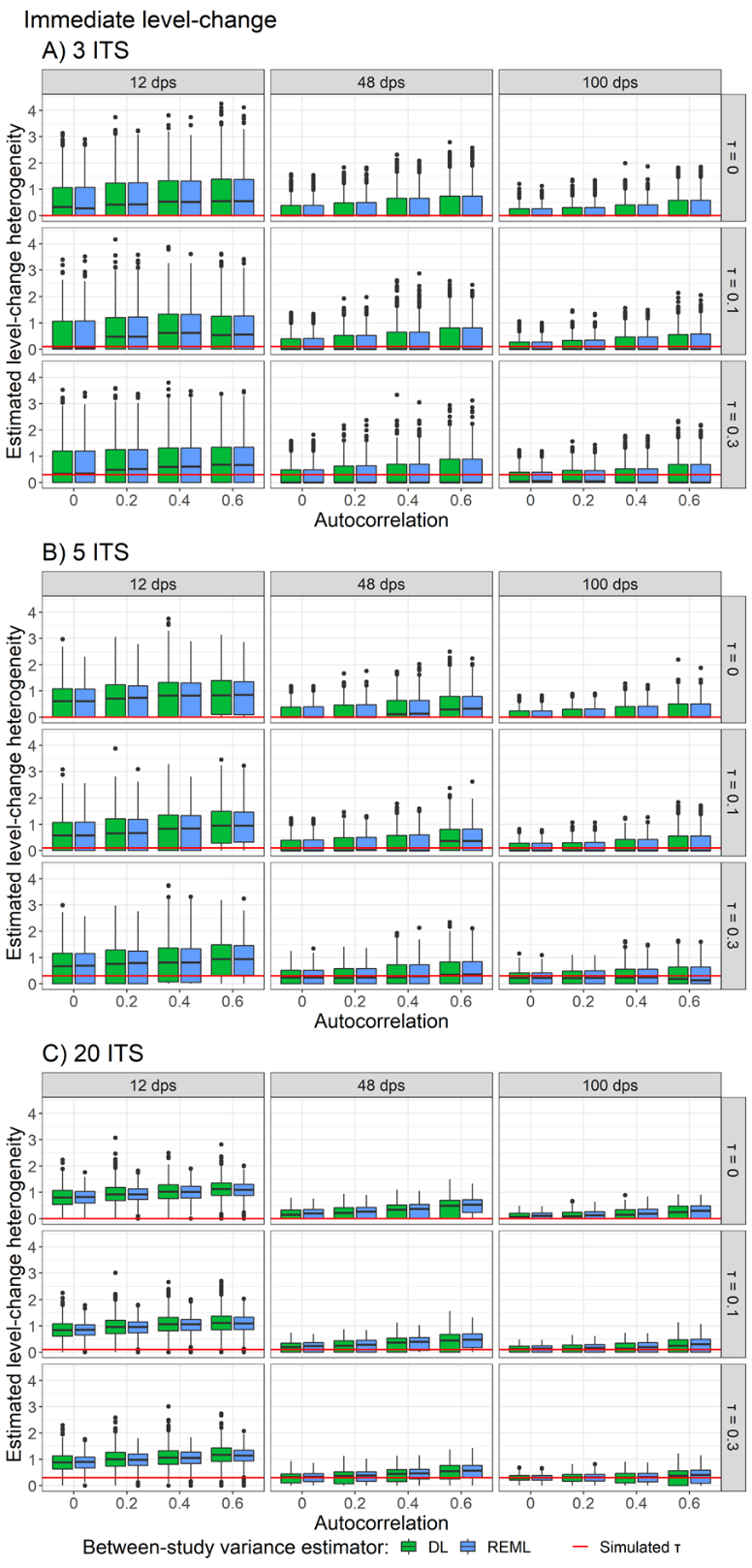

Appendix Figure S39. Plots of level-change heterogeneity estimated using a random effects meta-analysis with DL (green) and REML (purple) between-study variance estimators (y-axis) when the A) 3 ITS, B) 5 ITS and C) 20 ITS studies were analysed with REML. Plots are presented separately by combinations of the true level-change heterogeneity (rows) and the number of datapoints (columns). The solid red lines indicate the true level-change heterogeneity. Simulation scenarios include a level-change of 1, slope-change of 0.1, slope-change heterogeneity of 0, and fixed autocorrelation.

DL, DerSimonian and Laird. dps, datapoints. HKSJ, Hartung-Knapp / Sidik-Jonkman. ITS, interrupted time series. OLS, ordinary least squares. REML, restricted maximum likelihood. WT, Wald-type.

##### 3.8.2 Slope -change heterogeneity

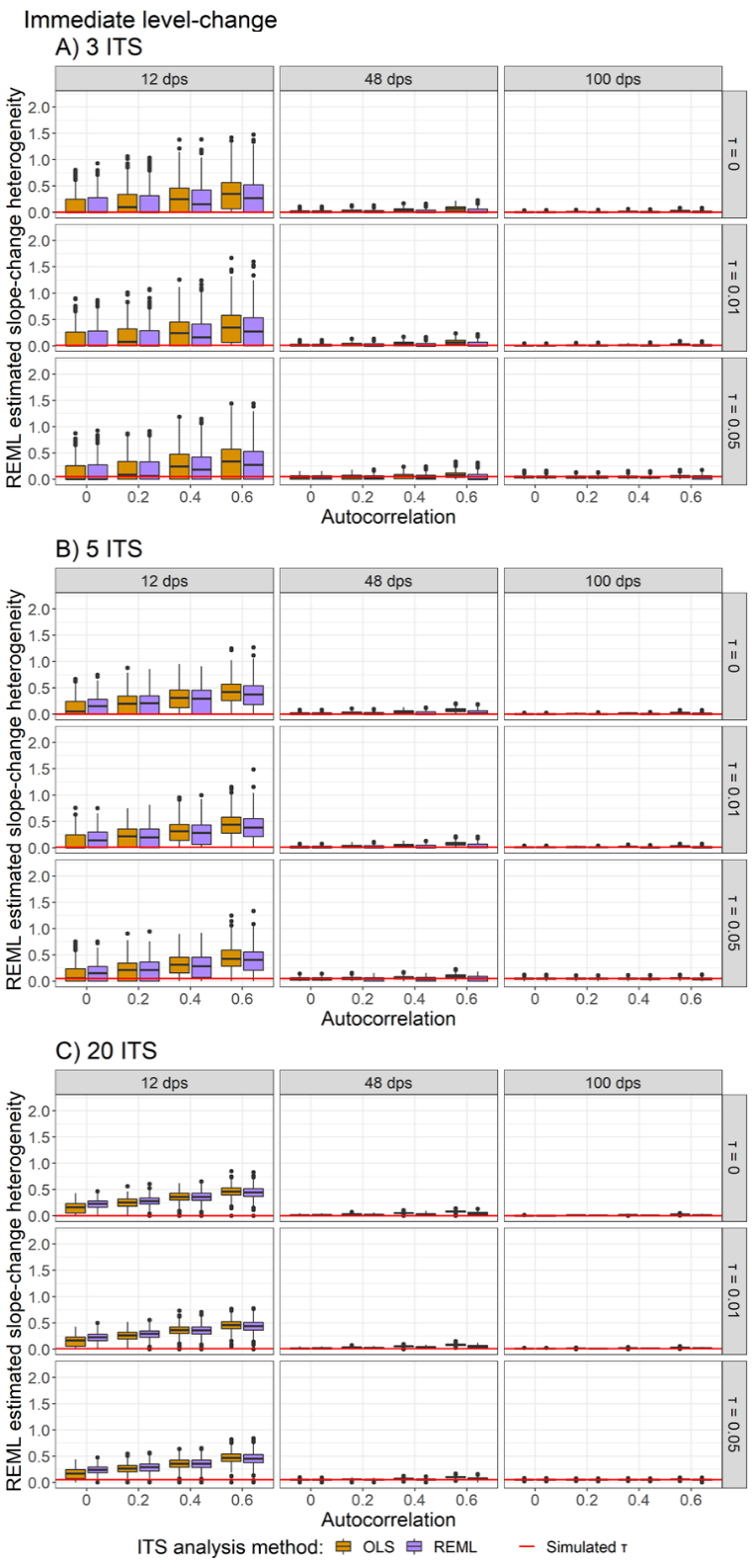

Appendix Figure S40. Plots of slope-change heterogeneity estimated using a random effects meta-analysis with the REML between-study variance estimator (y-axis) when the A) 3 ITS, B) 5 ITS and C) 20 ITS studies were analysed with OLS (orange) and REML (purple) ITS analysis methods. Plots are presented separately by combinations of the true slope-change heterogeneity (rows) and the number of datapoints (columns). The solid red lines indicate the true level-change heterogeneity. Simulation scenarios include a level-change of 1, level-change heterogeneity of 0, slope-change of 0.1, and fixed autocorrelation.

DL, DerSimonian and Laird. dps, datapoints. HKSJ, Hartung-Knapp / Sidik-Jonkman. ITS, interrupted time series. OLS, ordinary least squares. REML, restricted maximum likelihood. WT, Wald-type.

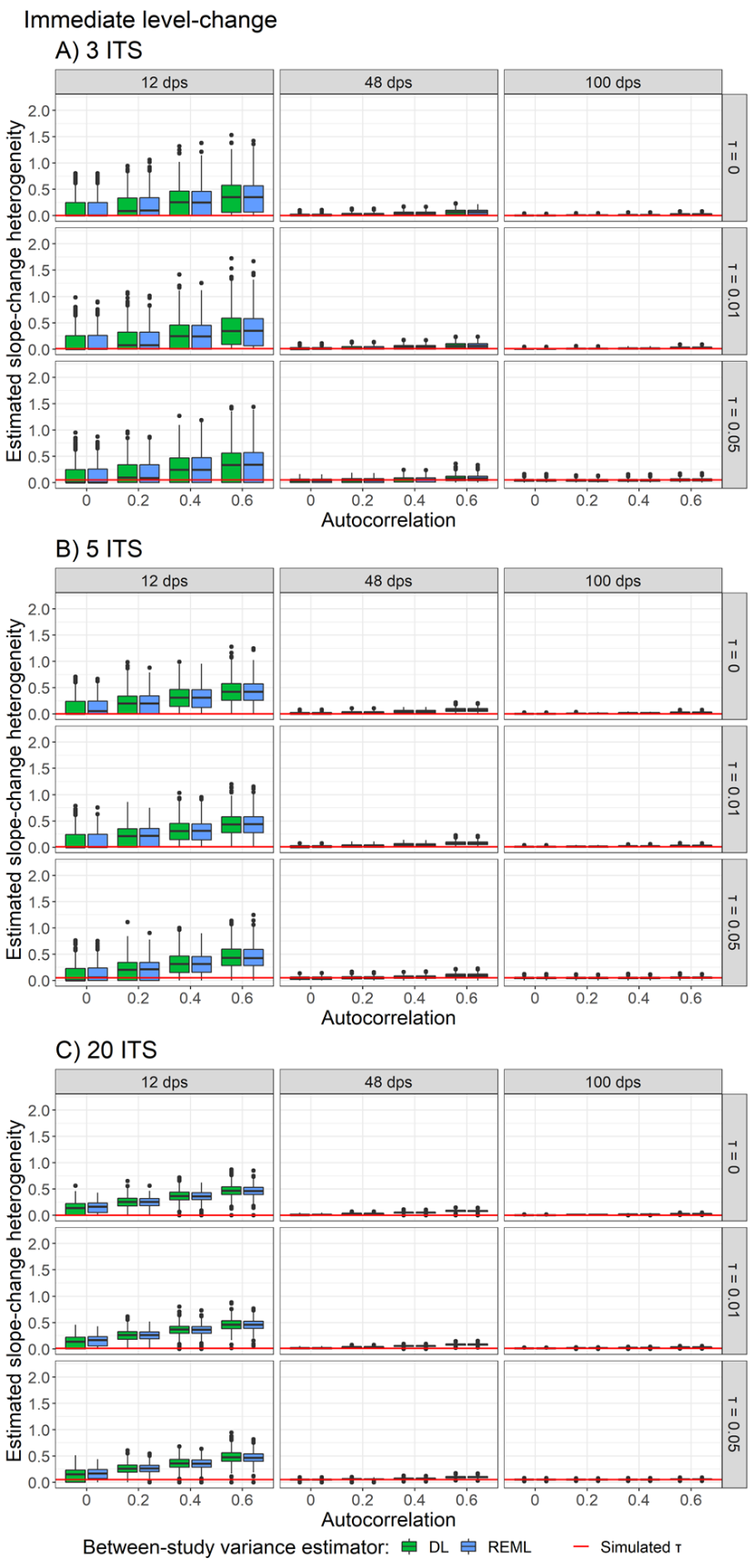

Appendix Figure S41. Plots of slope-change heterogeneity estimated using a random effects meta-analysis with DL (green) and REML (purple) between-study variance estimators (y-axis) when the A) 3 ITS, B) 5 ITS and C) 20 ITS studies were analysed with OLS. Plots are presented separately by combinations of the true slope-change heterogeneity (rows) and the number of datapoints (columns). The solid red lines indicate the true slope-change heterogeneity. Simulation scenarios include a level-change of 1, level-change heterogeneity of 0, slope-change of 0.1, and fixed autocorrelation.

DL, DerSimonian and Laird. dps, datapoints. HKSJ, Hartung-Knapp / Sidik-Jonkman. ITS, interrupted time series. OLS, ordinary least squares. REML, restricted maximum likelihood. WT, Wald-type.

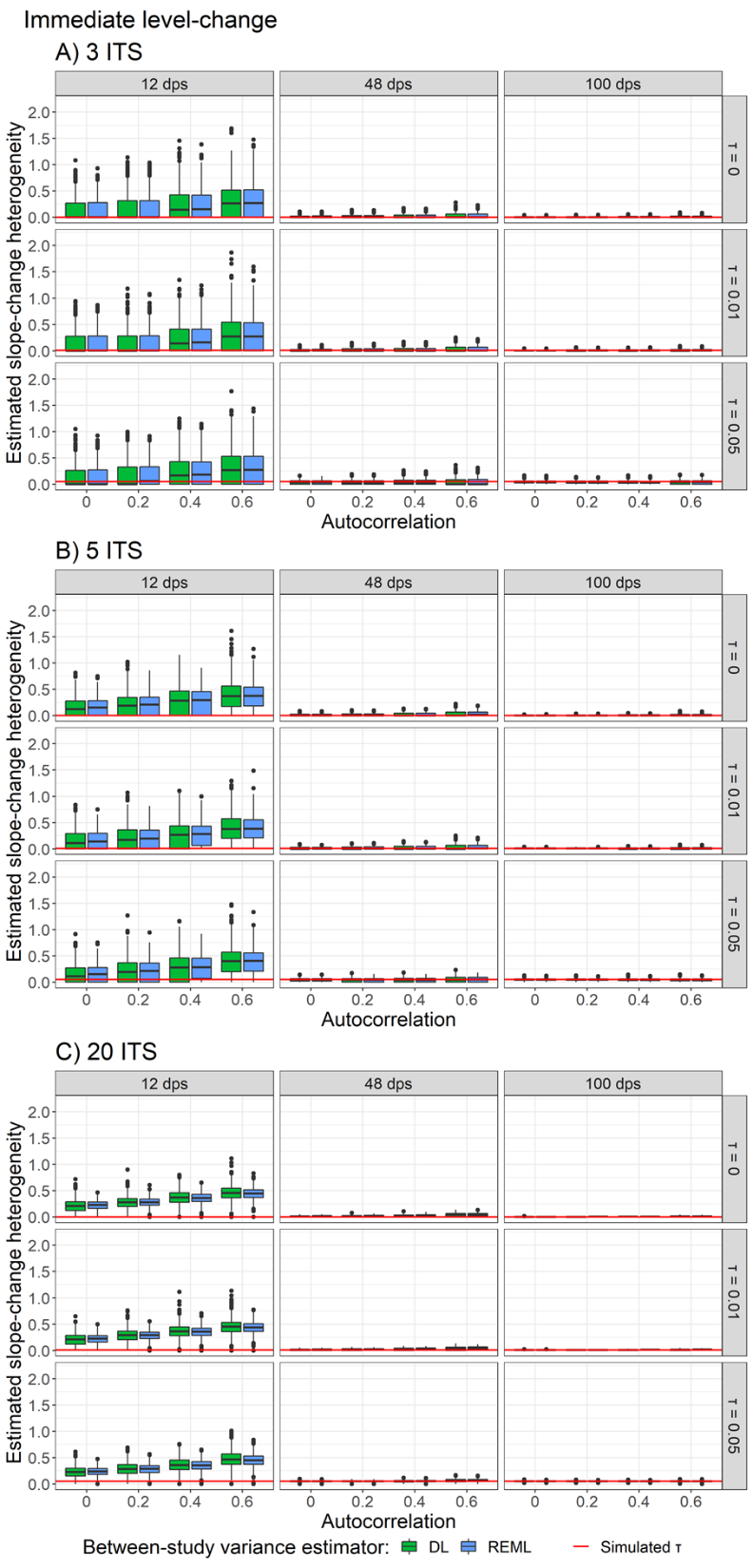

Appendix Figure S42. Plots of slope-change heterogeneity estimated using a random effects meta-analysis with DL (green) and REML (purple) between-study variance estimators (y-axis) when the A) 3 ITS, B) 5 ITS and C) 20 ITS studies were analysed with REML. Plots are presented separately by combinations of the true slope-change heterogeneity (rows) and the number of datapoints (columns). The solid red lines indicate the true slope-change heterogeneity. Simulation scenarios include a level-change of 1, level-change heterogeneity of 0, slope-change of 0.1, and fixed autocorrelation.

DL, DerSimonian and Laird. dps, datapoints. HKSJ, Hartung-Knapp / Sidik-Jonkman. ITS, interrupted time series. OLS, ordinary least squares. REML, restricted maximum likelihood. WT, Wald-type.

#### 3.9 Proportion of included studies analysed with REML vs PW

There was a higher rate of simulations with PW analyses among the 12 datapoint series scenarios, yet coverage, power, and empirical standard errors did not importantly differ from the simulations where REML was used.

##### 3.9.1 Estimation of level-change

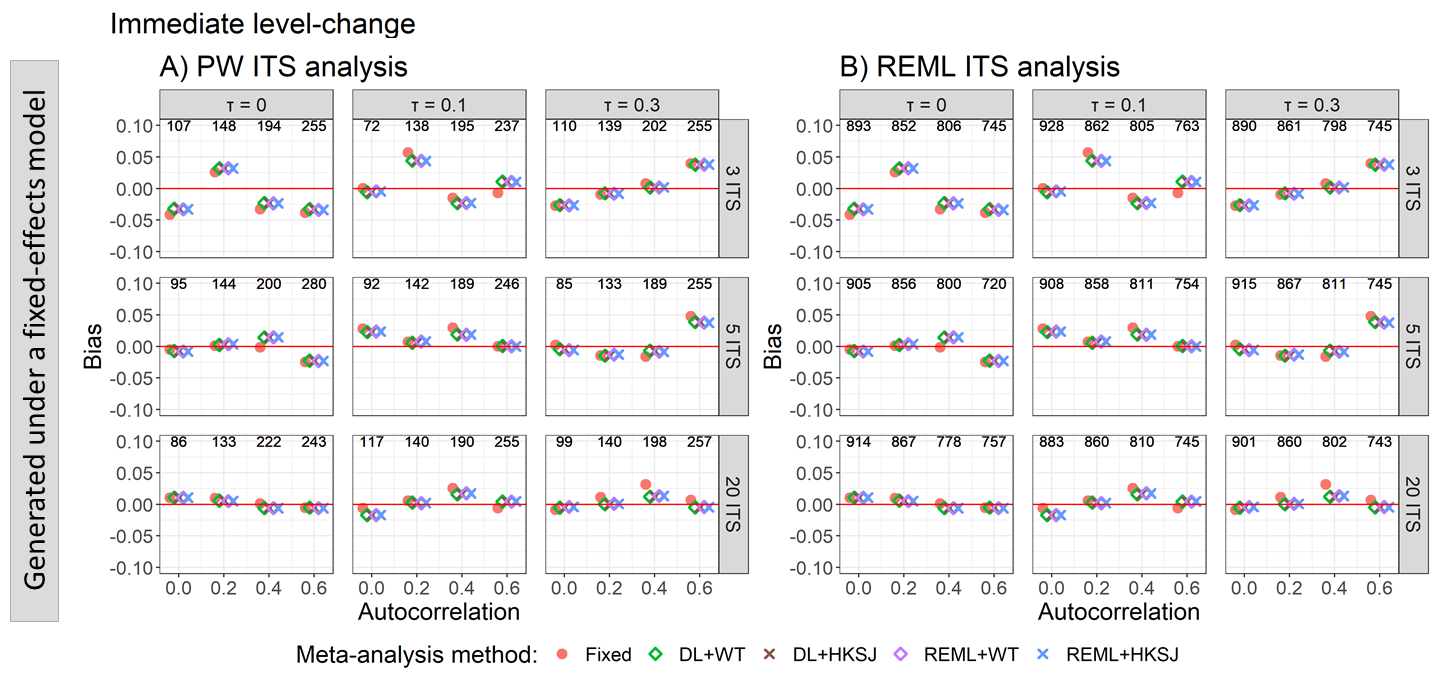

Appendix Figure S43. Plots of bias of the immediate level-change (y-axis) when the ITS are analysed with PW (A) and REML (B) for the fixed effect (red circles), DL+WT (green diamonds) and DL+HKSJ (brown crosses), REML+WT (purple diamonds) and REML+HKSJ (blue crosses) meta-analysis methods versus autocorrelation (x-axis). Plots are presented separately by combinations of the number of included studies (rows) and level-change heterogeneity (columns). Simulation scenarios include ITS with 12 datapoints, a level-change of 1, slope-change of 0, slope-change heterogeneity of 0, and fixed levels of autocorrelation.

DL, DerSimonian and Laird. dps, datapoints. HKSJ, Hartung-Knapp / Sidik-Jonkman. ITS, interrupted time series. PW, Prais-Winsten. REML, restricted maximum likelihood. WT, Wald-type.

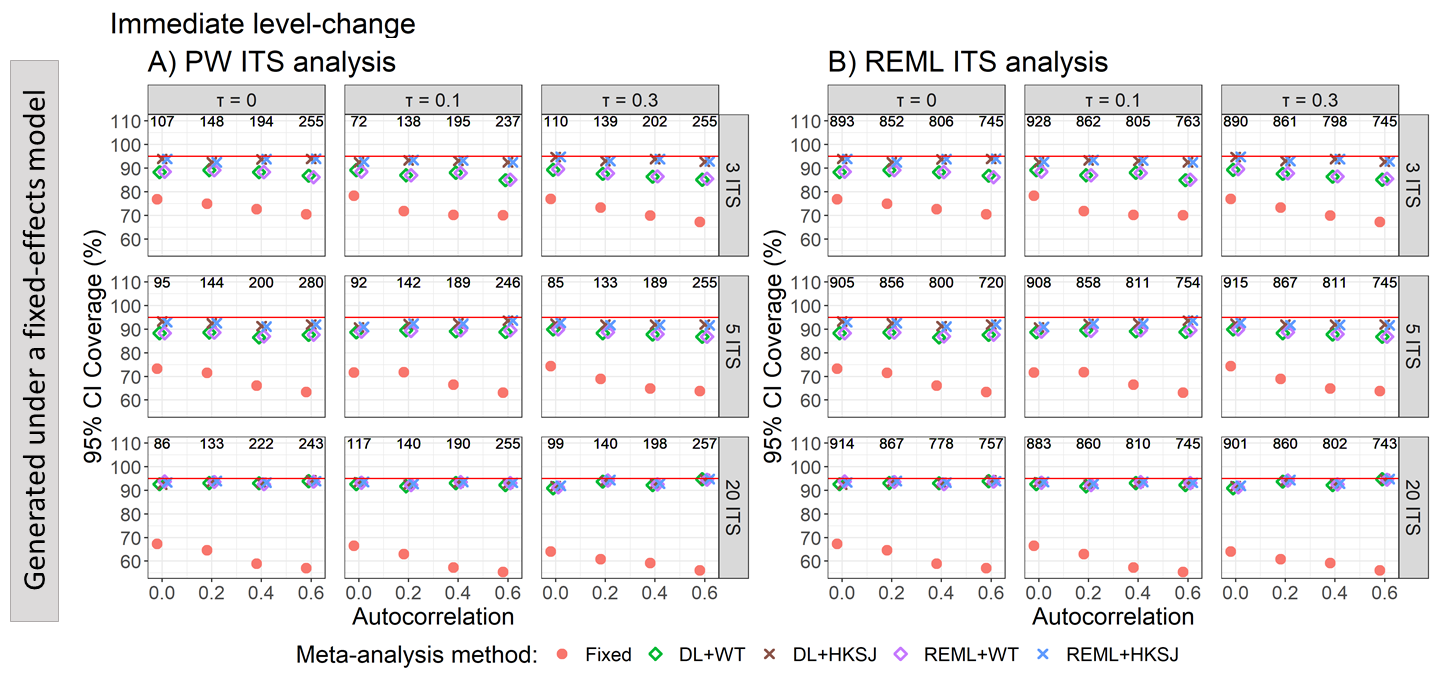

Appendix Figure S44. Plots of 95% confidence interval coverage of the immediate level-change (y-axis) when the ITS are analysed with PW (A) and REML (B) for the fixed effect (red circles), DL+WT (green diamonds) and DL+HKSJ (brown crosses), REML+WT (purple diamonds) and REML+HKSJ (blue crosses) meta-analysis methods versus autocorrelation (x-axis). Plots are presented separately by combinations of the number of included studies (rows) and level-change heterogeneity (columns). Simulation scenarios include ITS with 12 datapoints, a level-change of 1, slope-change of 0, slope-change heterogeneity of 0, and fixed levels of autocorrelation.

DL, DerSimonian and Laird. dps, datapoints. HKSJ, Hartung-Knapp / Sidik-Jonkman. ITS, interrupted time series. PW, Prais-Winsten. REML, restricted maximum likelihood. WT, Wald-type.

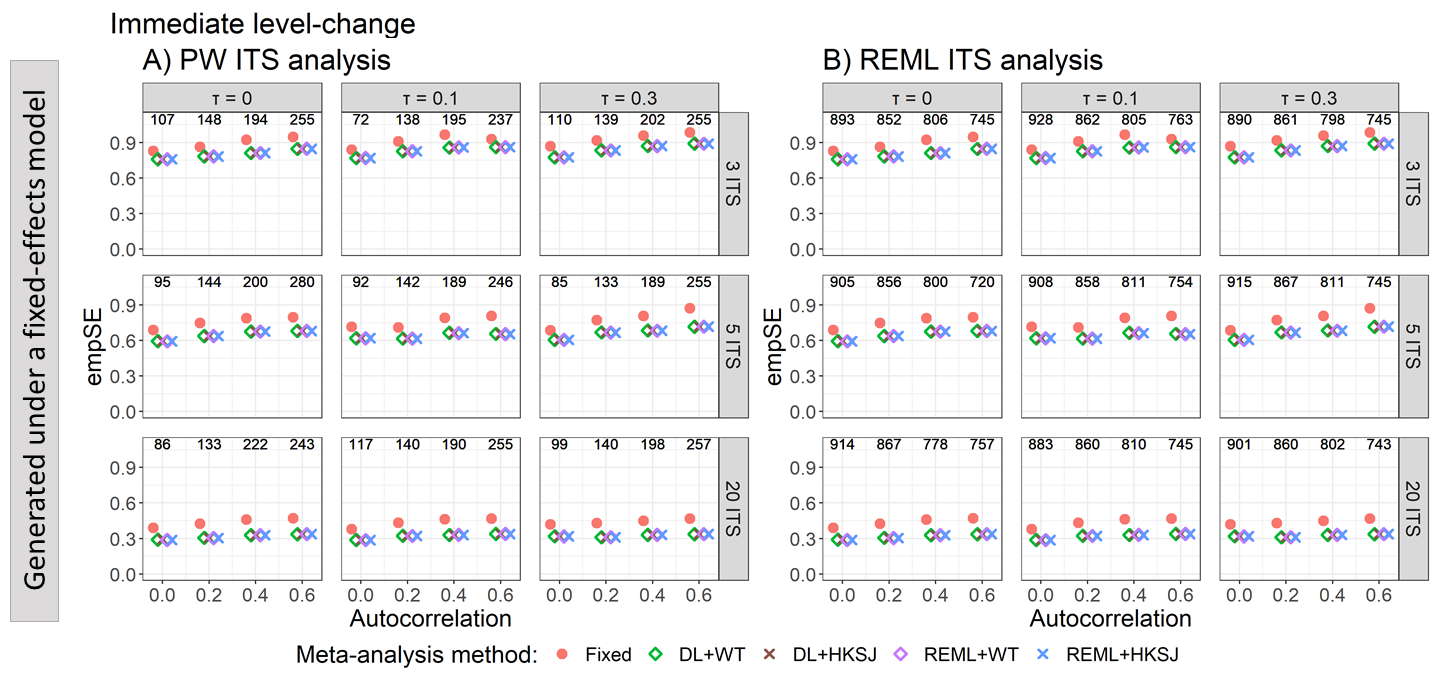

Appendix Figure S45. Plots of the empirical standard error of the immediate level-change (y-axis) when the ITS are analysed with PW (A) and REML (B) for the fixed effect (red circles), DL+WT (green diamonds) and DL+HKSJ (brown crosses), REML+WT (purple diamonds) and REML+HKSJ (blue crosses) meta-analysis methods versus autocorrelation (x-axis). Plots are presented separately by combinations of the number of included studies (rows) and level-change heterogeneity (columns). Simulation scenarios include ITS with 12 datapoints, a level-change of 1, slope-change of 0, slope-change heterogeneity of 0, and fixed levels of autocorrelation.

DL, DerSimonian and Laird. dps, datapoints. HKSJ, Hartung-Knapp / Sidik-Jonkman. ITS, interrupted time series. PW, Prais-Winsten. REML, restricted maximum likelihood. WT, Wald-type.

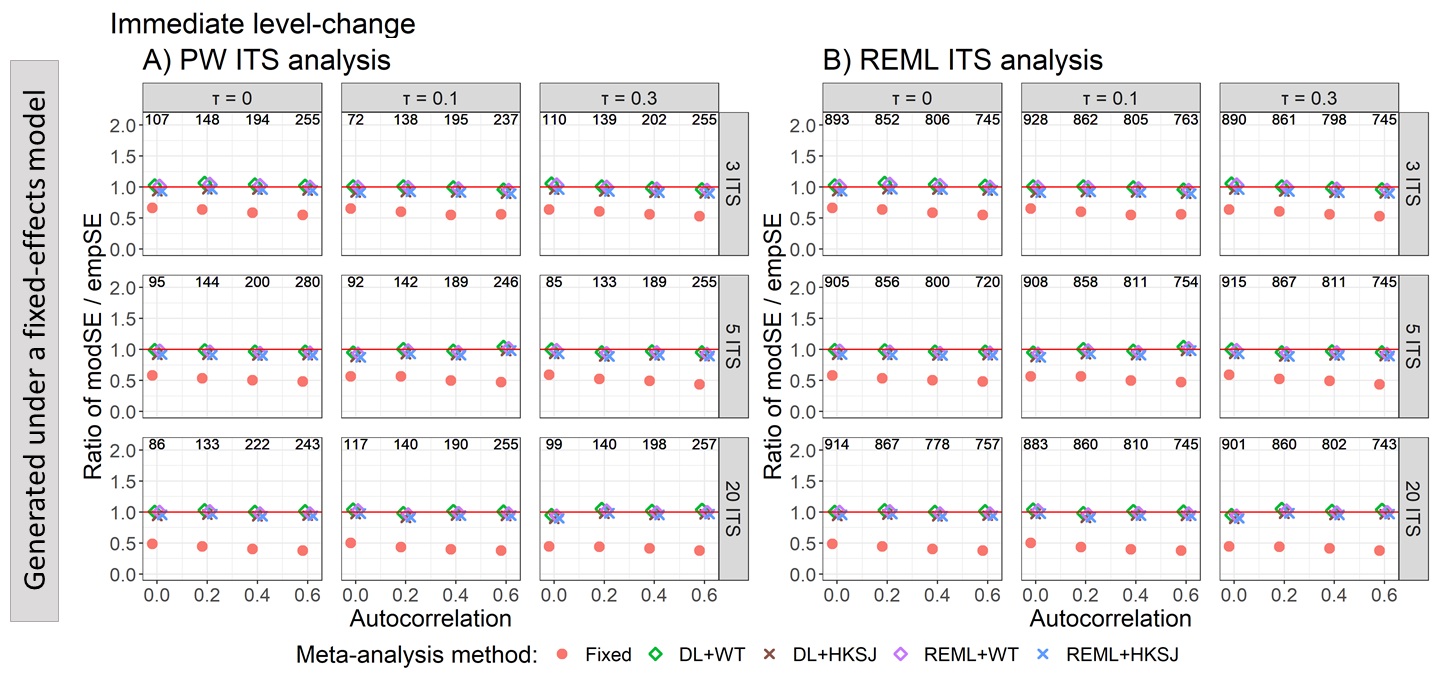

Appendix Figure S46. Plots of the ratio of model based standard error (modSE) to the empirical standard error (empSE) of the immediate level-change (y-axis) when the ITS are analysed with PW (A) and REML (B) for the fixed effect (red circles), DL+WT (green diamonds) and DL+HKSJ (brown crosses), REML+WT (purple diamonds) and REML+HKSJ (blue crosses) meta-analysis methods versus autocorrelation (x-axis). Plots are presented separately by combinations of the number of included studies (rows) and level-change heterogeneity (columns). Simulation scenarios include ITS with 12 datapoints, a level-change of 1, slope-change of 0, slope-change heterogeneity of 0, and fixed levels of autocorrelation.

DL, DerSimonian and Laird. dps, datapoints. HKSJ, Hartung-Knapp / Sidik-Jonkman. ITS, interrupted time series. PW, Prais-Winsten. REML, restricted maximum likelihood. WT, Wald-type.

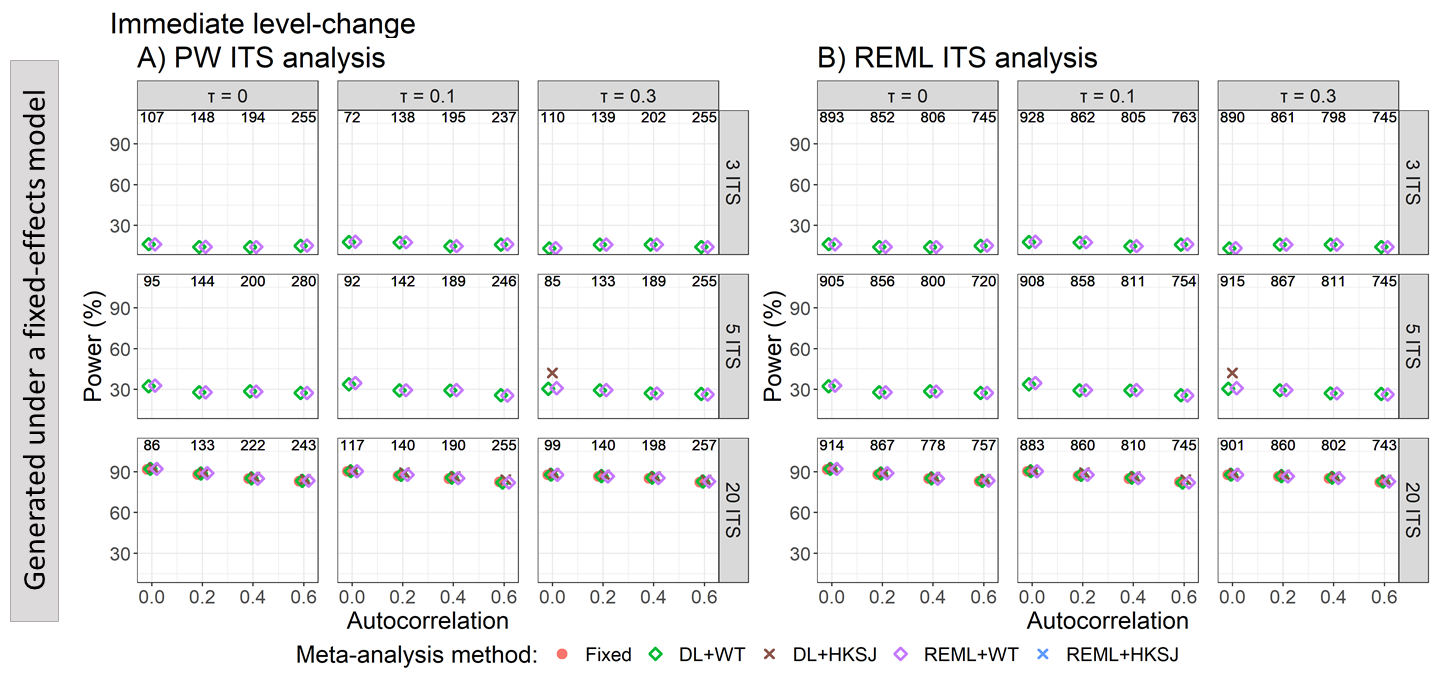

Appendix Figure S47. Plots of the statistical power (the percentage of simulations that have a 95% confidence interval that did not include zero – only scenarios with a confidence interval coverage of greater than 90% are plotted) of the immediate level-change (y-axis) when the ITS are analysed with PW (A) and REML (B) for the fixed effect (red circles), DL+WT (green diamonds) and DL+HKSJ (brown crosses), REML+WT (purple diamonds) and REML+HKSJ (blue crosses) meta-analysis methods versus autocorrelation (x-axis). Plots are presented separately by combinations of the number of included studies (rows) and level-change heterogeneity (columns). Simulation scenarios include ITS with 12 datapoints, a level-change of 1, slope-change of 0, slope-change heterogeneity of 0, and fixed levels of autocorrelation.

DL, DerSimonian and Laird. dps, datapoints. HKSJ, Hartung-Knapp / Sidik-Jonkman. ITS, interrupted time series. PW, Prais-Winsten. REML, restricted maximum likelihood. WT, Wald-type.

##### 3.9.2 Estimation of slope-change

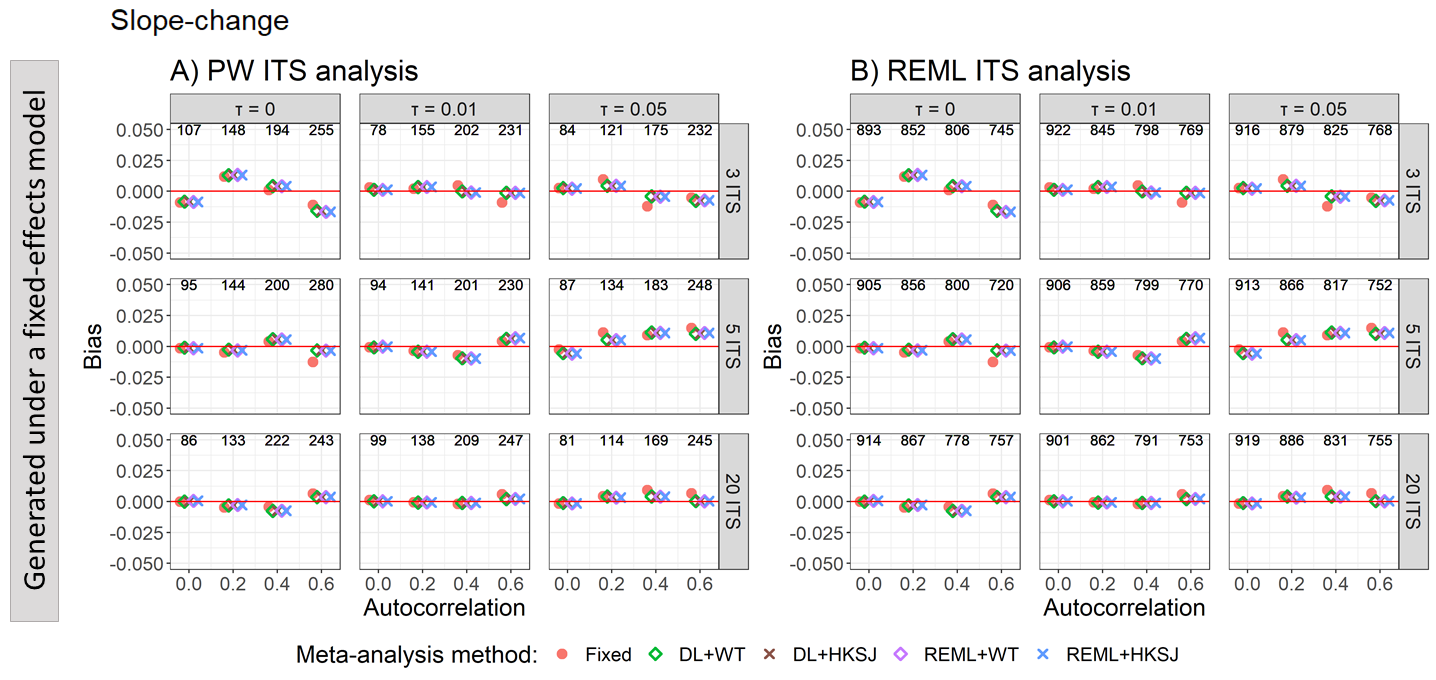

Appendix Figure S48. Plots of bias of the immediate level-change (y-axis) when the ITS are analysed with PW (A) and REML (B) for the fixed effect (red circles), DL+WT (green diamonds) and DL+HKSJ (brown crosses), REML+WT (purple diamonds) and REML+HKSJ (blue crosses) meta-analysis methods versus autocorrelation (x-axis). Plots are presented separately by combinations of the number of included studies (rows) and level-change heterogeneity (columns). Simulation scenarios include ITS with 12 datapoints, a level-change of 1, slope-change of 0, slope-change heterogeneity of 0, and fixed levels of autocorrelation.

DL, DerSimonian and Laird. dps, datapoints. HKSJ, Hartung-Knapp / Sidik-Jonkman. ITS, interrupted time series. PW, Prais-Winsten. REML, restricted maximum likelihood. WT, Wald-type.

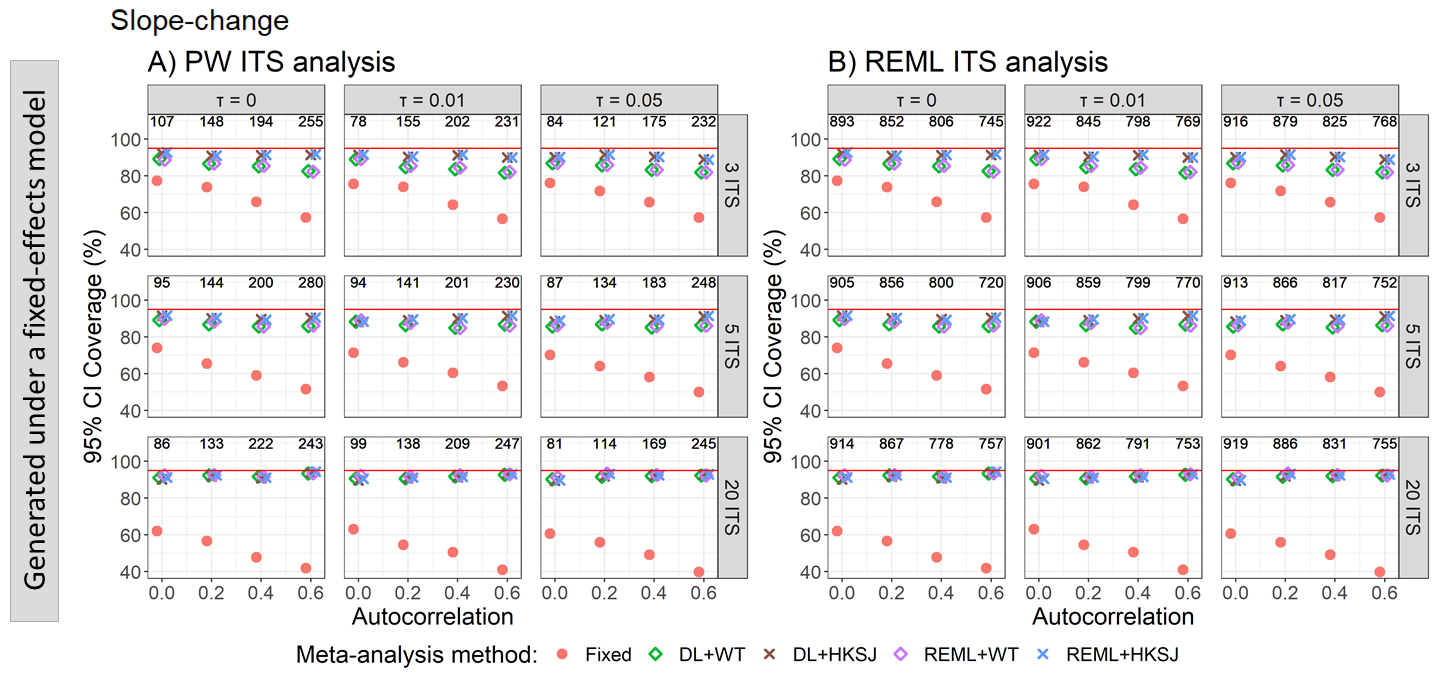

Appendix Figure S49. Plots of 95% confidence interval coverage of slope-change (y-axis) when the ITS are analysed with PW (A) and REML (B) for the fixed effect (red circles), DL+WT (green diamonds) and DL+HKSJ (brown crosses), REML+WT (purple diamonds) and REML+HKSJ (blue crosses) meta-analysis methods versus autocorrelation (x-axis). Plots are presented separately by combinations of the number of included studies (rows) and level-change heterogeneity (columns). Simulation scenarios include ITS with 12 datapoints, a level-change of 1, slope-change of 0, slope-change heterogeneity of 0, and fixed levels of autocorrelation.

DL, DerSimonian and Laird. dps, datapoints. HKSJ, Hartung-Knapp / Sidik-Jonkman. ITS, interrupted time series. PW, Prais-Winsten. REML, restricted maximum likelihood. WT, Wald-type.

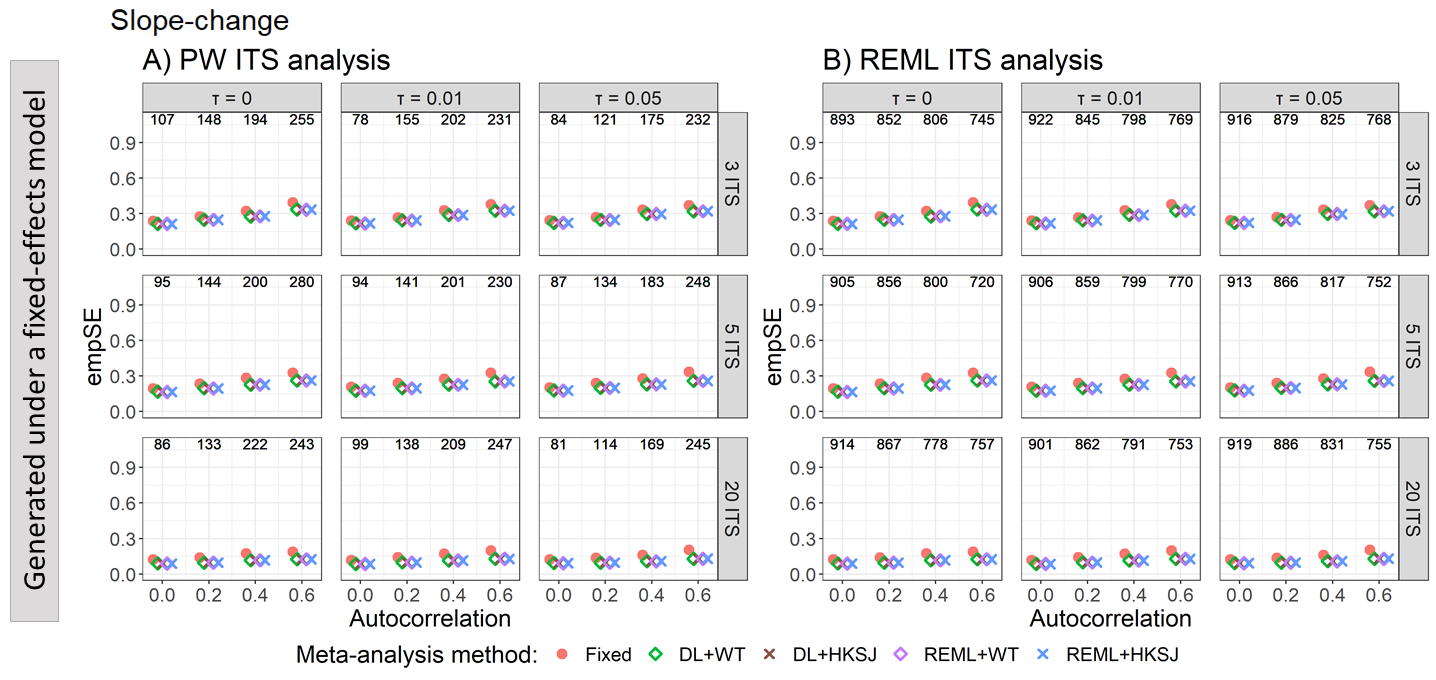

Appendix Figure S50. Plots of the empirical standard error of slope-change (y-axis) when the ITS are analysed with PW (A) and REML (B) for the fixed effect (red circles), DL+WT (green diamonds) and DL+HKSJ (brown crosses), REML+WT (purple diamonds) and REML+HKSJ (blue crosses) meta-analysis methods versus autocorrelation (x-axis). Plots are presented separately by combinations of the number of included studies (rows) and level-change heterogeneity (columns). Simulation scenarios include ITS with 12 datapoints, a level-change of 1, slope-change of 0, slope-change heterogeneity of 0, and fixed levels of autocorrelation.

DL, DerSimonian and Laird. dps, datapoints. HKSJ, Hartung-Knapp / Sidik-Jonkman. ITS, interrupted time series. PW, Prais-Winsten. REML, restricted maximum likelihood. WT, Wald-type.

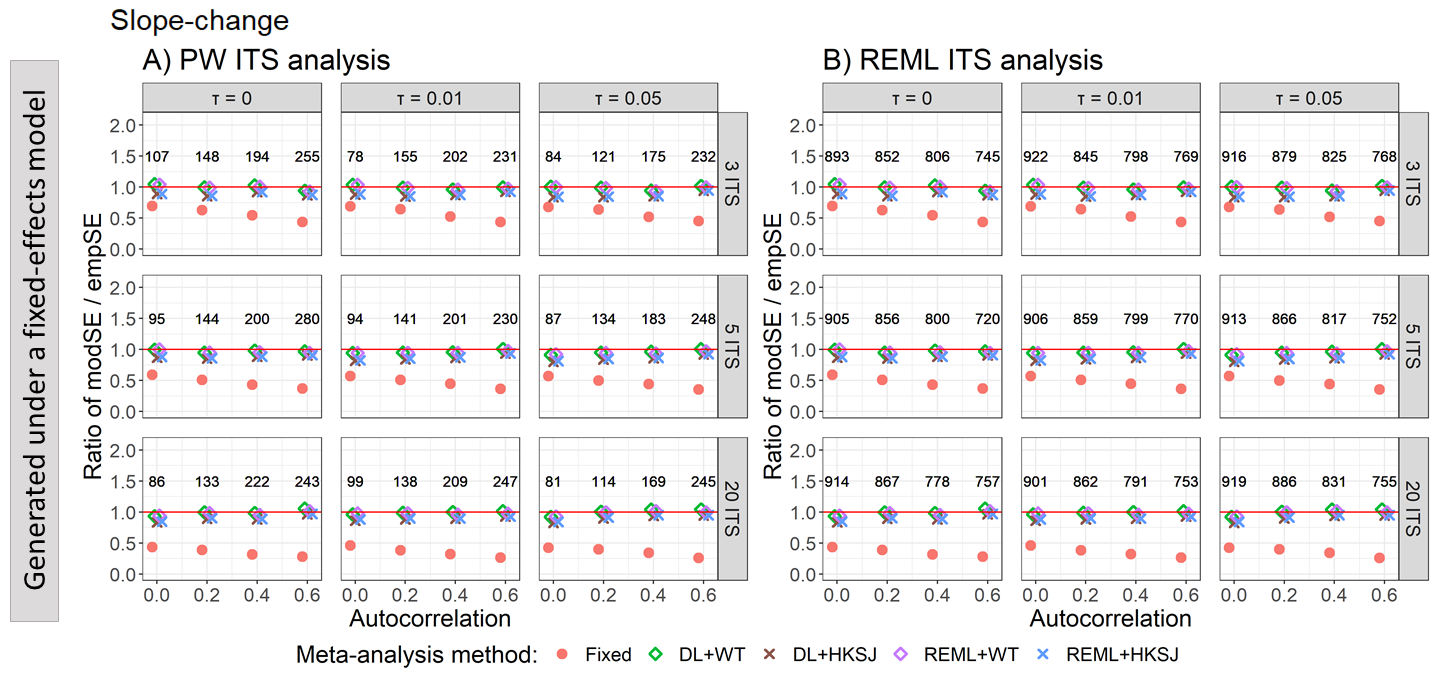

Appendix Figure S51. Plots of the ratio of model based standard error (modSE) to the empirical standard error (empSE) of slope-change (y-axis) when the ITS are analysed with PW (A) and REML (B) for the fixed effect (red circles), DL+WT (green diamonds) and DL+HKSJ (brown crosses), REML+WT (purple diamonds) and REML+HKSJ (blue crosses) meta-analysis methods versus autocorrelation (x-axis). Plots are presented separately by combinations of the number of included studies (rows) and level-change heterogeneity (columns). Simulation scenarios include ITS with 12 datapoints, a level-change of 1, slope-change of 0, slope-change heterogeneity of 0, and fixed levels of autocorrelation.

DL, DerSimonian and Laird. dps, datapoints. HKSJ, Hartung-Knapp / Sidik-Jonkman. ITS, interrupted time series. PW, Prais-Winsten. REML, restricted maximum likelihood. WT, Wald-type.

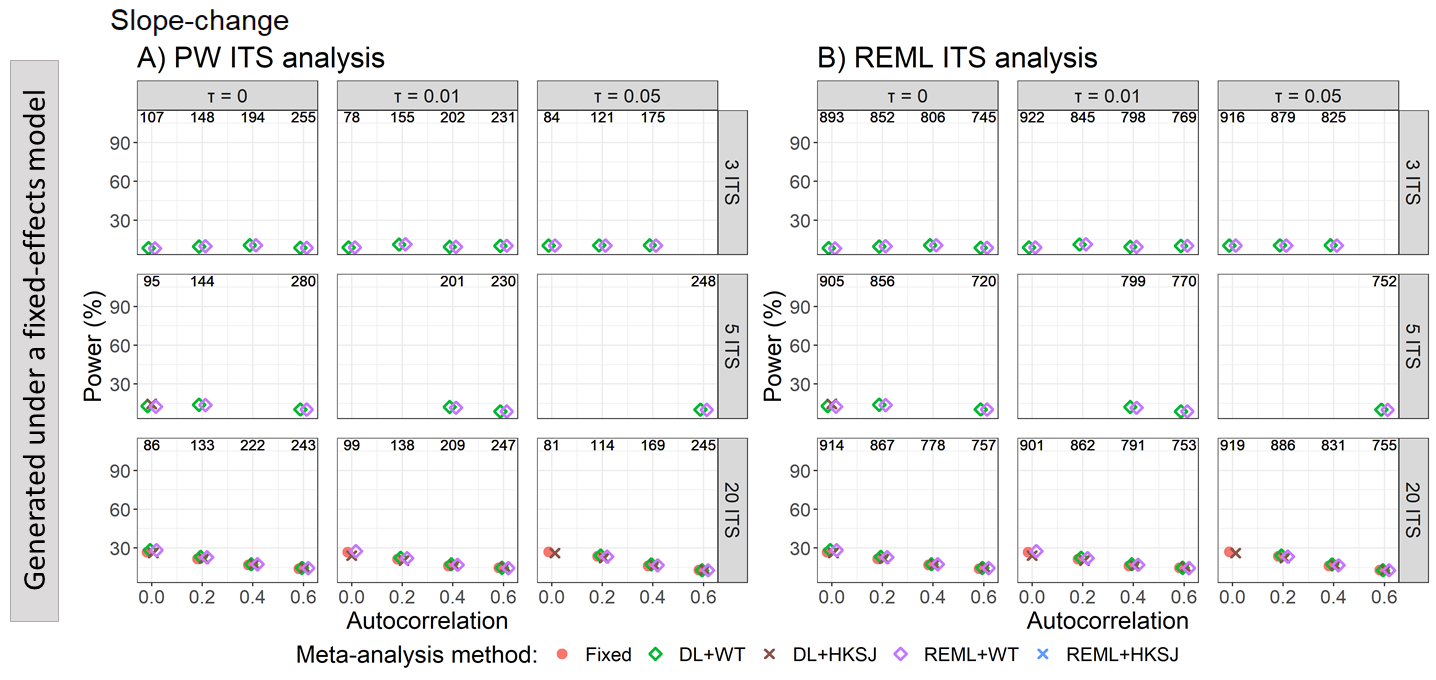

Appendix Figure S52. Plots of the statistical power (the percentage of simulations that have a 95% confidence interval that did not include zero – only scenarios with a confidence interval coverage of greater than 90% are plotted) of slope-change (y-axis) when the ITS are analysed with PW (A) and REML (B) for the fixed effect (red circles), DL+WT (green diamonds) and DL+HKSJ (brown crosses), REML+WT (purple diamonds) and REML+HKSJ (blue crosses) meta-analysis methods versus autocorrelation (x-axis). Plots are presented separately by combinations of the number of included studies (rows) and level-change heterogeneity (columns). Simulation scenarios include ITS with 12 datapoints, a level-change of 1, slope-change of 0, slope-change heterogeneity of 0, and fixed levels of autocorrelation.

DL, DerSimonian and Laird. dps, datapoints. HKSJ, Hartung-Knapp / Sidik-Jonkman. ITS, interrupted time series. PW, Prais-Winsten. REML, restricted maximum likelihood. WT, Wald-type.

### Appendix 4 – Results for alternative scenarios; other level- and slope-change combinations

There was no impact on the performance when level- and slope-change design parameters were varied.

#### 4.1 Bias

##### 4.1.1 Estimation of level-change – bias vs autocorrelation for scenarios with different level and slope-change combinations

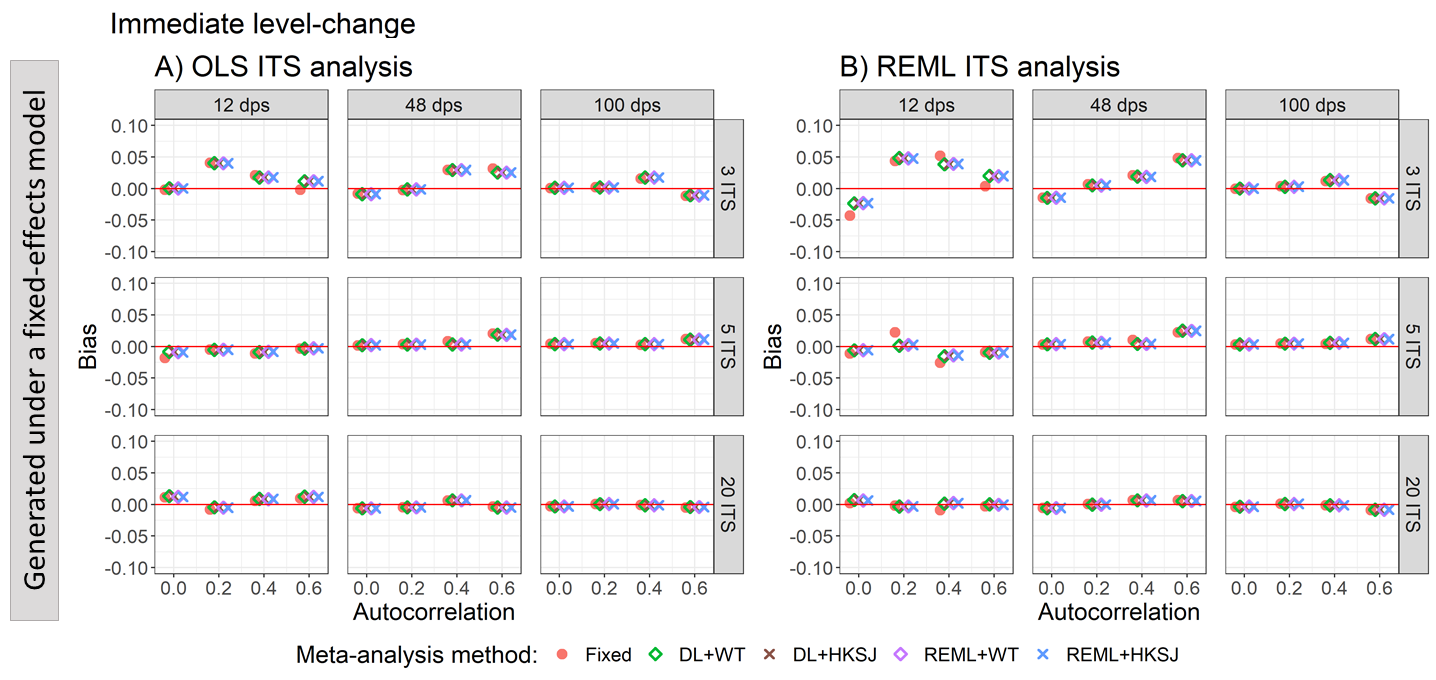

Appendix Figure S53. Plots of the bias of the immediate level-change (y-axis) when the data was generated under a fixed-effect model and the ITS studies were analysed with OLS (A) and REML (B) using fixed-effect (red circles), DL+WT (green diamonds) and DL+HKSJ (brown crosses), REML+WT (purple diamonds) and REML+HKSJ (blue crosses) meta-analysis methods versus autocorrelation (x-axis). Plots are presented separately by combinations of the number of included studies (rows) and the number of datapoints (columns). Simulation scenarios include a **level-change of 0**, level-change heterogeneity of 0, **slope-change of 0**, slope-change heterogeneity of 0, and fixed levels of autocorrelation.

DL, DerSimonian and Laird. dps, datapoints. HKSJ, Hartung-Knapp / Sidik-Jonkman. ITS, interrupted time series. OLS, ordinary least squares. REML, restricted maximum likelihood. WT, Wald-type.

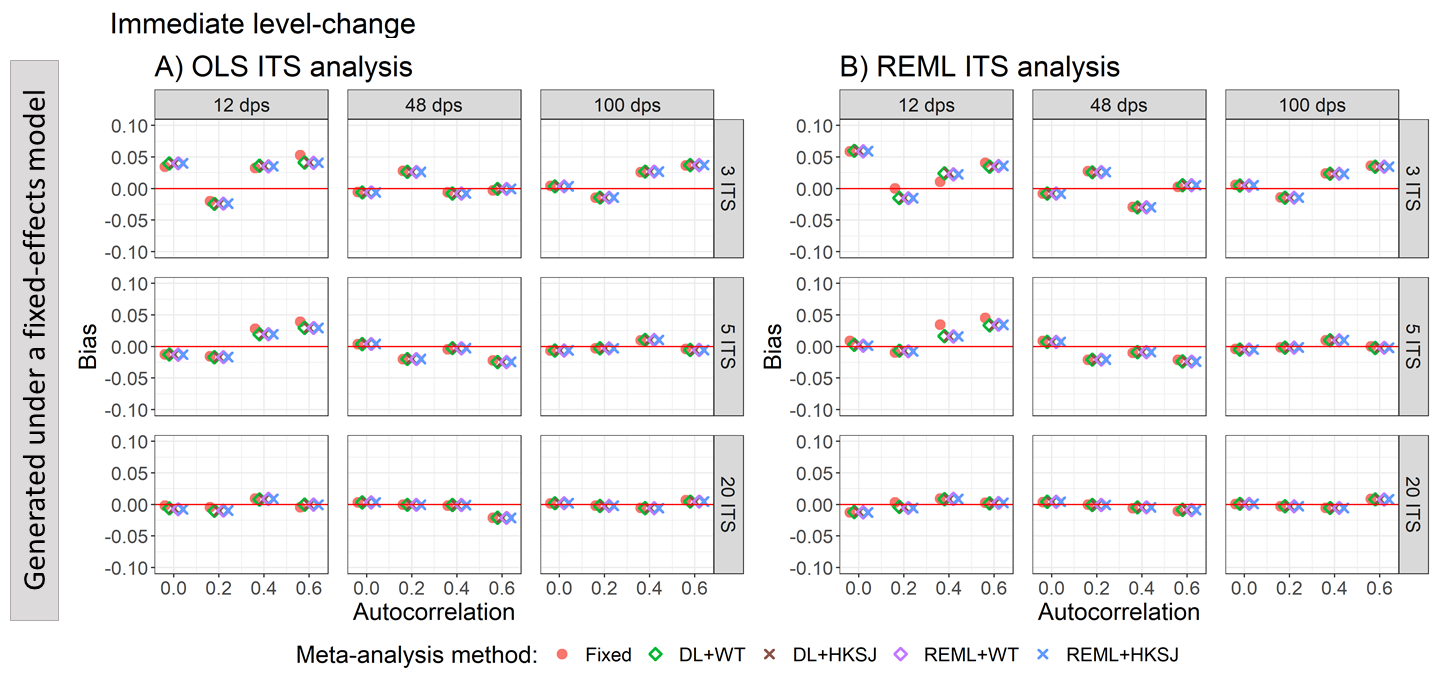

Appendix Figure S54. Plots of the bias of the immediate level-change (y-axis) when the data was generated under a fixed-effect model and the ITS studies were analysed with OLS (A) and REML (B) using fixed-effect (red circles), DL+WT (green diamonds) and DL+HKSJ (brown crosses), REML+WT (purple diamonds) and REML+HKSJ (blue crosses) meta-analysis methods versus autocorrelation (x-axis). Plots are presented separately by combinations of the number of included studies (rows) and the number of datapoints (columns). Simulation scenarios include a **level-change of 0**, level-change heterogeneity of 0, **slope-change of 0.1**, slope-change heterogeneity of 0, and fixed levels of autocorrelation.

DL, DerSimonian and Laird. dps, datapoints. HKSJ, Hartung-Knapp / Sidik-Jonkman. ITS, interrupted time series. OLS, ordinary least squares. REML, restricted maximum likelihood. WT, Wald-type.

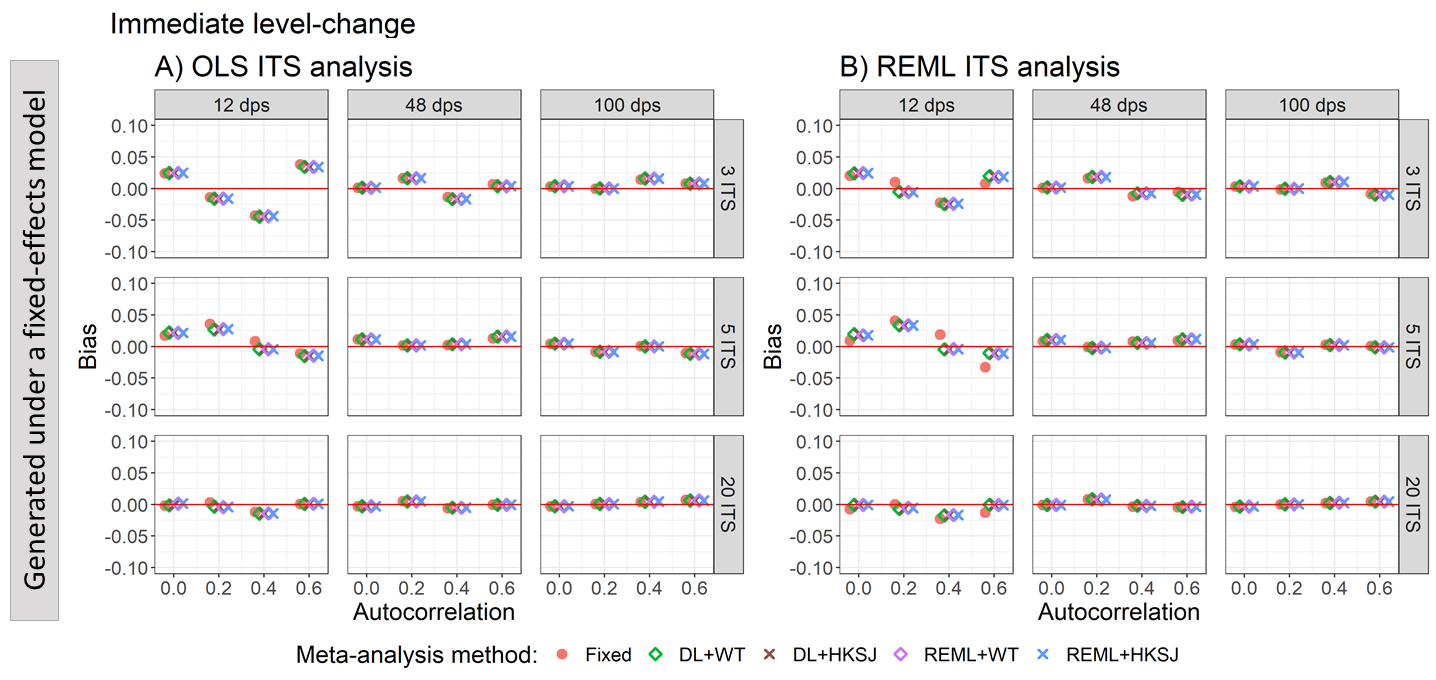

Appendix Figure S55. Plots of the bias of the immediate level-change (y-axis) when the data was generated under a fixed-effect model and the ITS studies were analysed with OLS (A) and REML (B) using fixed-effect (red circles), DL+WT (green diamonds) and DL+HKSJ (brown crosses), REML+WT (purple diamonds) and REML+HKSJ (blue crosses) meta-analysis methods versus autocorrelation (x-axis). Plots are presented separately by combinations of the number of included studies (rows) and the number of datapoints (columns). Simulation scenarios include a **level-change of** **1**, level-change heterogeneity of 0, **slope-change of 0**, slope-change heterogeneity of 0, and fixed levels of autocorrelation.

DL, DerSimonian and Laird. dps, datapoints. HKSJ, Hartung-Knapp / Sidik-Jonkman. ITS, interrupted time series. OLS, ordinary least squares. REML, restricted maximum likelihood. WT, Wald-type.

##### 4.1.2 Estimation of level-change – bias vs level-change heterogeneity for scenarios with different level and slope-change combinations

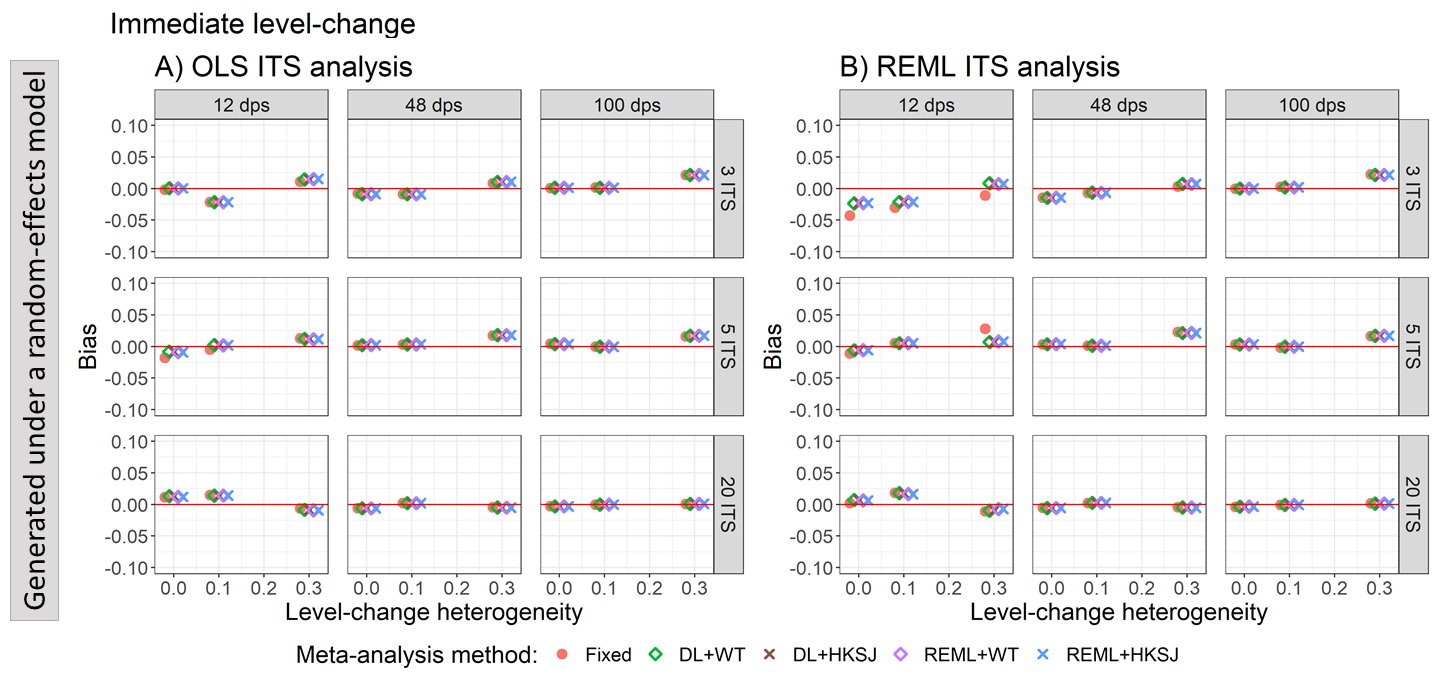

Appendix Figure S56. Plots of the bias of the immediate level-change (y-axis) when the ITS are analysed with OLS (A) and REML (B) using fixed-effect (red circles), DL+WT (green diamonds) and DL+HKSJ (brown crosses), REML+WT (purple diamonds) and REML+HKSJ (blue crosses) meta-analysis methods versus level-change heterogeneity (x-axis). Plots are presented separately by combinations of the number of included studies (rows) and the number of datapoints (columns). Simulation scenarios include a **level-change of 0,** **slope-change of 0**, slope-change heterogeneity of 0, and autocorrelation of zero.

DL, DerSimonian and Laird. dps, datapoints. HKSJ, Hartung-Knapp / Sidik-Jonkman. ITS, interrupted time series. OLS, ordinary least squares. REML, restricted maximum likelihood. WT, Wald-type.

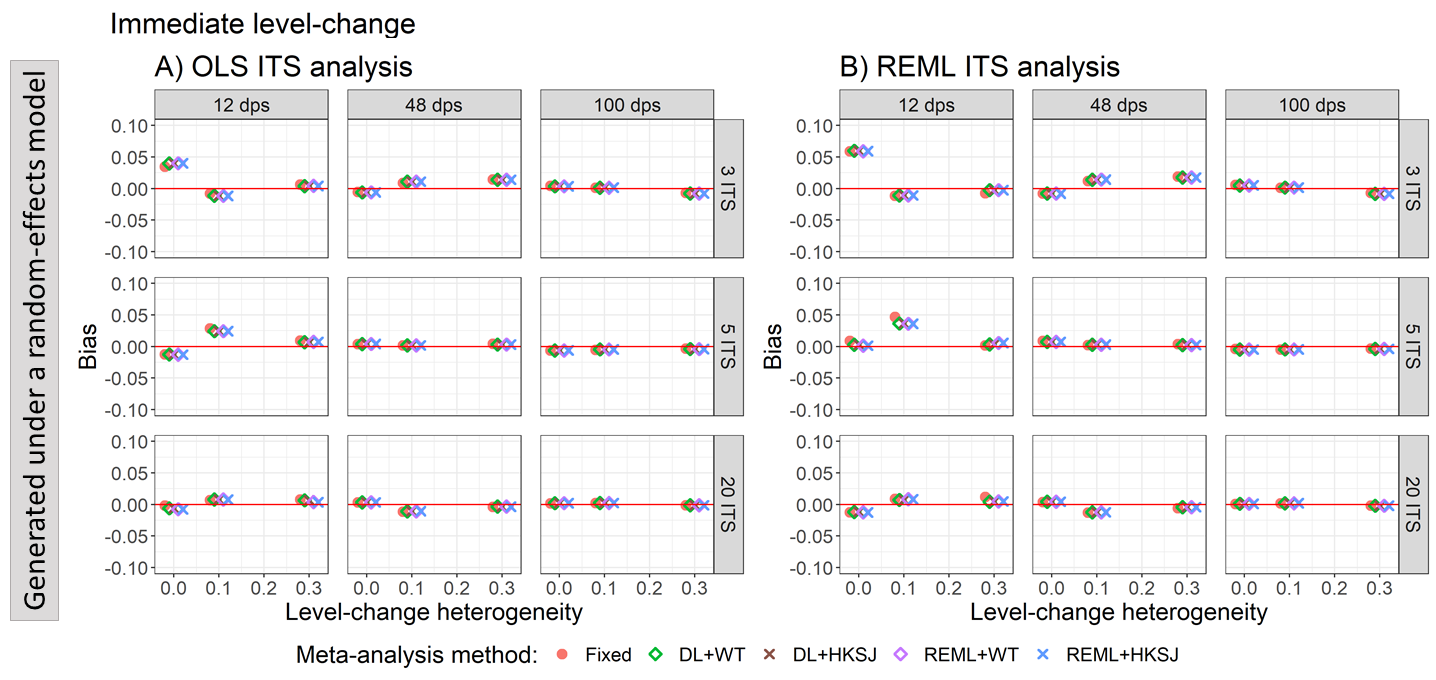

Appendix Figure S57. Plots of the bias of the immediate level-change (y-axis) when the ITS are analysed with OLS (A) and REML (B) using fixed-effect (red circles), DL+WT (green diamonds) and DL+HKSJ (brown crosses), REML+WT (purple diamonds) and REML+HKSJ (blue crosses) meta-analysis methods versus level-change heterogeneity (x-axis). Plots are presented separately by combinations of the number of included studies (rows) and the number of datapoints (columns). Simulation scenarios include a **level-change of 0,** **slope-change of 0.1**, slope-change heterogeneity of 0, and autocorrelation of zero.

DL, DerSimonian and Laird. dps, datapoints. HKSJ, Hartung-Knapp / Sidik-Jonkman. ITS, interrupted time series. OLS, ordinary least squares. REML, restricted maximum likelihood. WT, Wald-type.

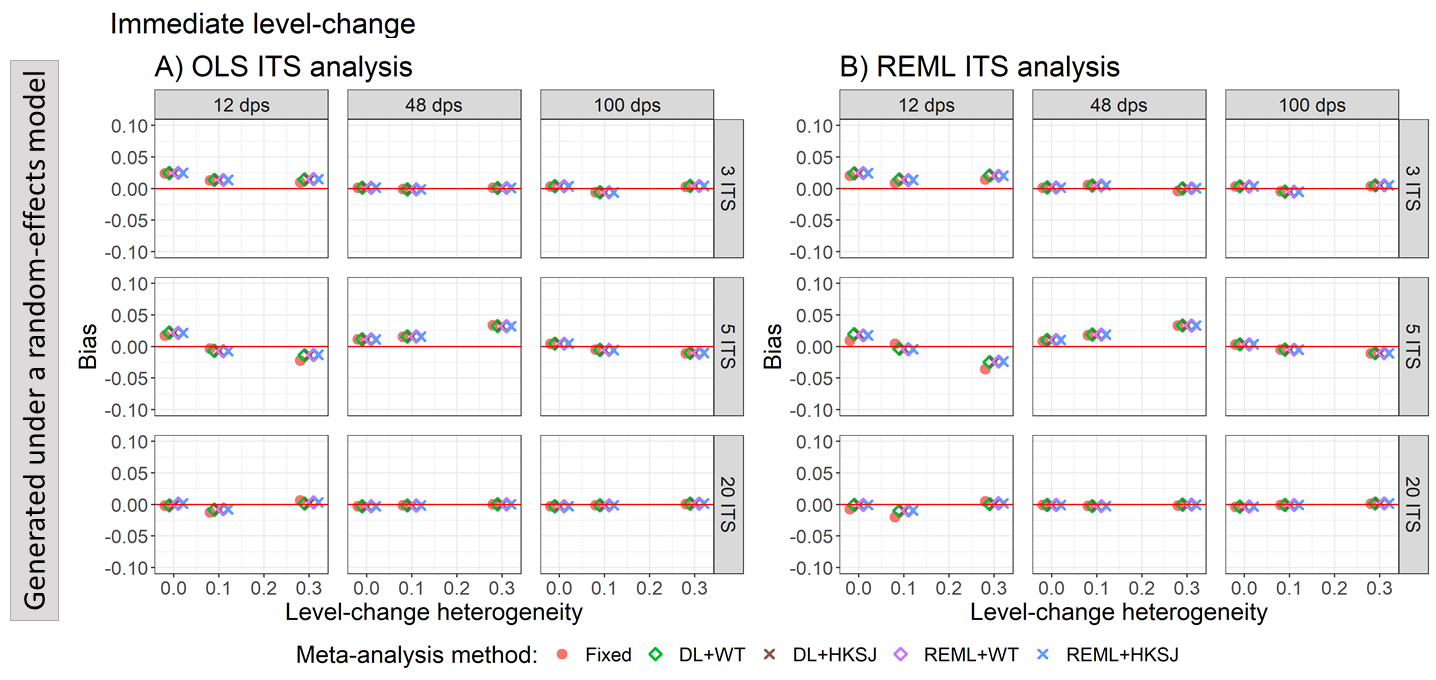

Appendix Figure S58. Plots of the bias of the immediate level-change (y-axis) when the ITS are analysed with OLS (A) and REML (B) using fixed-effect (red circles), DL+WT (green diamonds) and DL+HKSJ (brown crosses), REML+WT (purple diamonds) and REML+HKSJ (blue crosses) meta-analysis methods versus level-change heterogeneity (x-axis). Plots are presented separately by combinations of the number of included studies (rows) and the number of datapoints (columns). Simulation scenarios include a **level-change of 1,** **slope-change of 0**, slope-change heterogeneity of 0, and autocorrelation of zero.

DL, DerSimonian and Laird. dps, datapoints. HKSJ, Hartung-Knapp / Sidik-Jonkman. ITS, interrupted time series. OLS, ordinary least squares. REML, restricted maximum likelihood. WT, Wald-type.

##### 4.1.3 Estimation of slope-change – bias vs autocorrelation for scenarios with different level and slope-change combinations

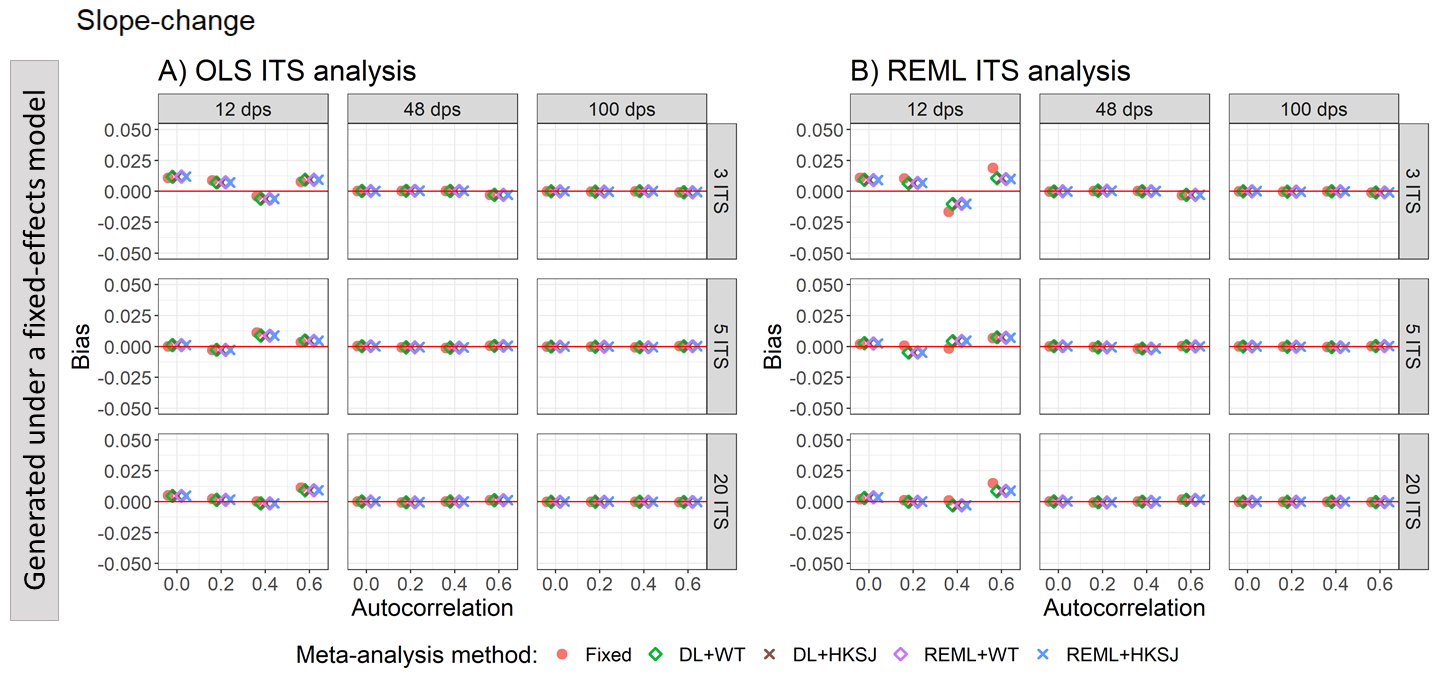

Appendix Figure S59. Plots of the bias of the slope-change (y-axis) when the data was generated under a fixed-effect model and the ITS studies were analysed with OLS (A) and REML (B) using fixed-effect (red circles), DL+WT (green diamonds) and DL+HKSJ (brown crosses), REML+WT (purple diamonds) and REML+HKSJ (blue crosses) meta-analysis methods versus autocorrelation (x-axis). Plots are presented separately by combinations of the number of included studies (rows) and the number of datapoints (columns). Simulation scenarios include a **level-change of 0**, level-change heterogeneity of 0, **slope-change of 0**, slope-change heterogeneity of 0, and fixed levels of autocorrelation.

DL, DerSimonian and Laird. dps, datapoints. HKSJ, Hartung-Knapp / Sidik-Jonkman. ITS, interrupted time series. OLS, ordinary least squares. REML, restricted maximum likelihood. WT, Wald-type.

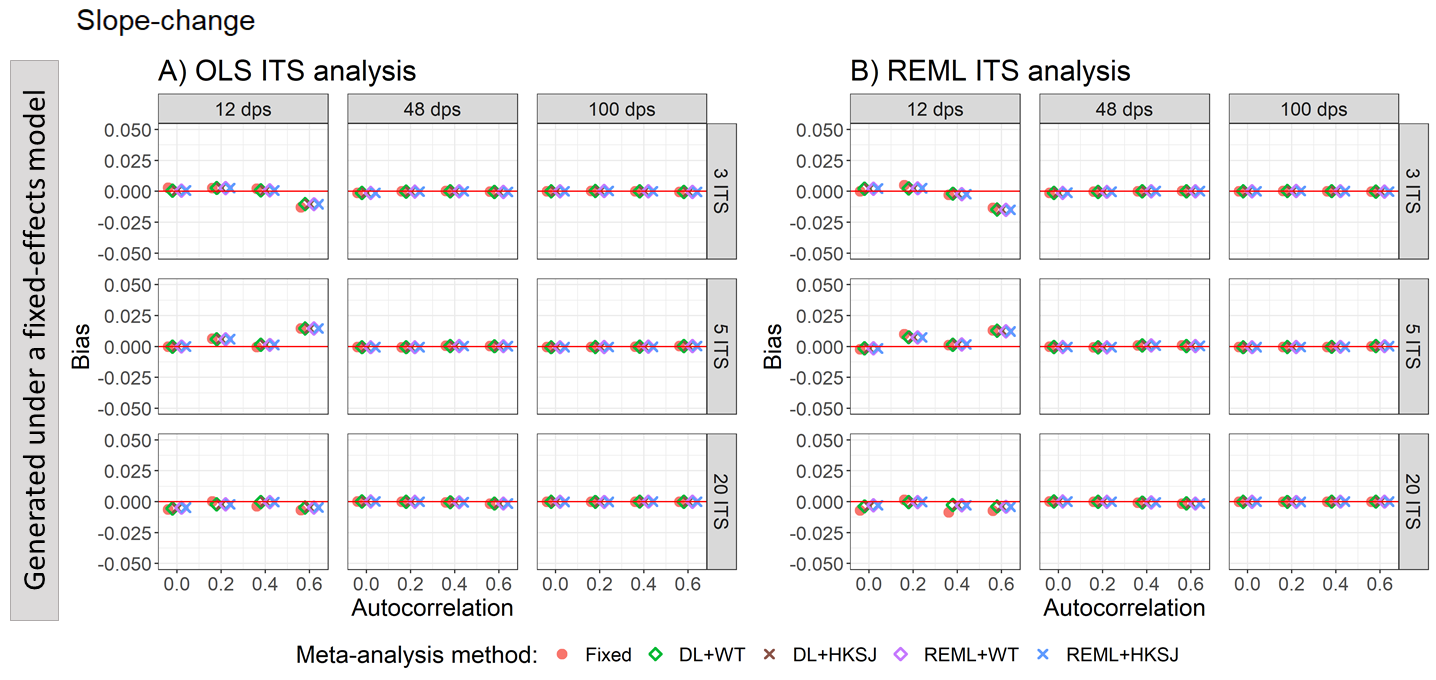

Appendix Figure S60. Plots of the bias of the slope-change (y-axis) when the data was generated under a fixed-effect model and the ITS studies were analysed with OLS (A) and REML (B) using fixed-effect (red circles), DL+WT (green diamonds) and DL+HKSJ (brown crosses), REML+WT (purple diamonds) and REML+HKSJ (blue crosses) meta-analysis methods versus autocorrelation (x-axis). Plots are presented separately by combinations of the number of included studies (rows) and the number of datapoints (columns). Simulation scenarios include a **level-change of 0**, level-change heterogeneity of 0, **slope-change of 0.1**, slope-change heterogeneity of 0, and fixed levels of autocorrelation.

DL, DerSimonian and Laird. dps, datapoints. HKSJ, Hartung-Knapp / Sidik-Jonkman. ITS, interrupted time series. OLS, ordinary least squares. REML, restricted maximum likelihood. WT, Wald-type.

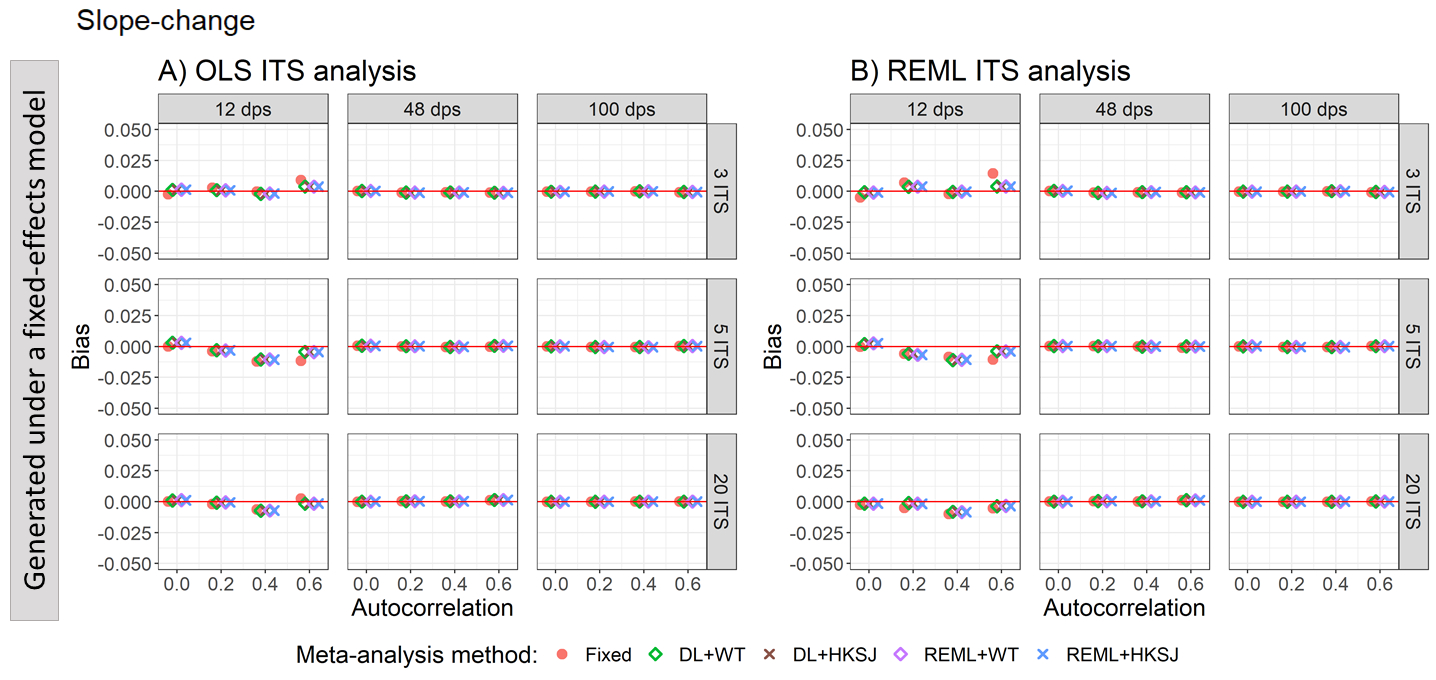

Appendix Figure S61. Plots of the bias of the slope-change (y-axis) when the data was generated under a fixed-effect model and the ITS studies were analysed with OLS (A) and REML (B) using fixed-effect (red circles), DL+WT (green diamonds) and DL+HKSJ (brown crosses), REML+WT (purple diamonds) and REML+HKSJ (blue crosses) meta-analysis methods versus autocorrelation (x-axis). Plots are presented separately by combinations of the number of included studies (rows) and the number of datapoints (columns). Simulation scenarios include a **level-change of 1**, level-change heterogeneity of 0, **slope-change of 0**, slope-change heterogeneity of 0, and fixed levels of autocorrelation.

DL, DerSimonian and Laird. dps, datapoints. HKSJ, Hartung-Knapp / Sidik-Jonkman. ITS, interrupted time series. OLS, ordinary least squares. REML, restricted maximum likelihood. WT, Wald-type.

##### 4.1.4 Estimation of slope-change – bias vs slope-change heterogeneity for scenarios with different level and slope-change combinations

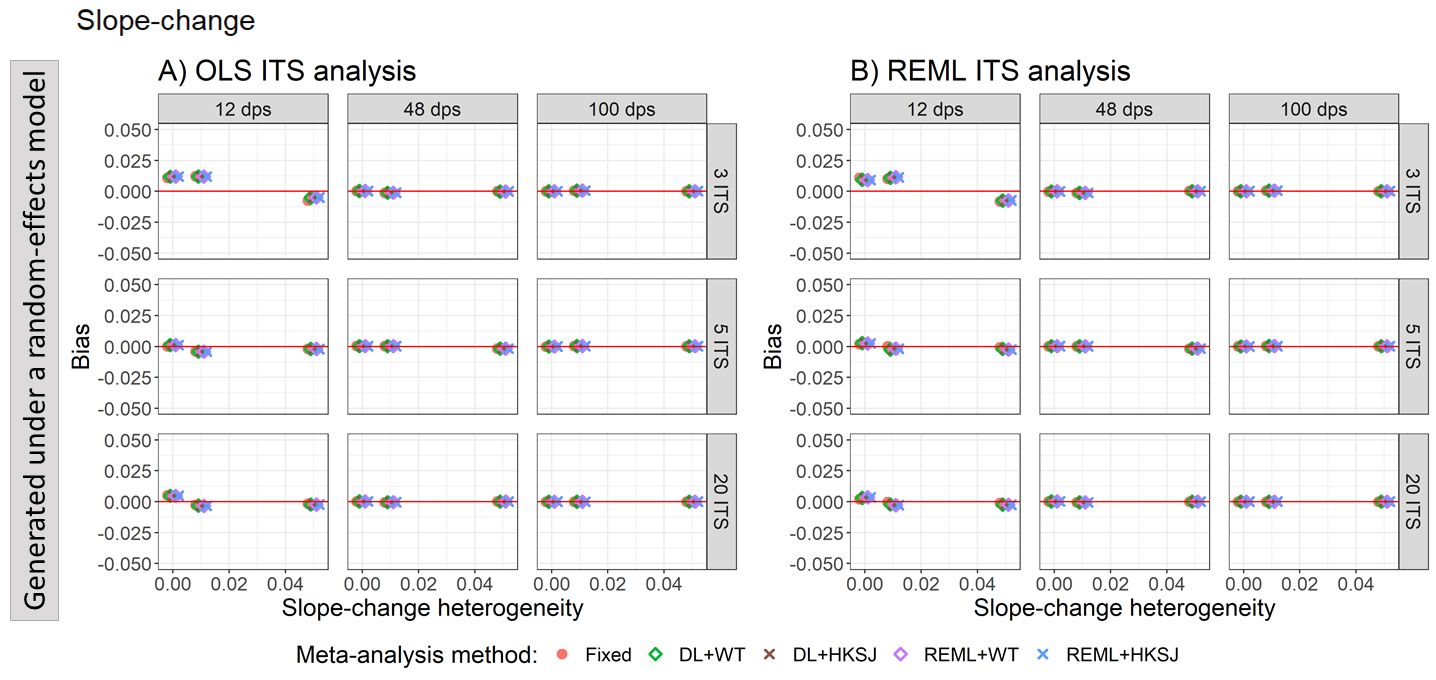

Appendix Figure S62. Plots of the bias of the slope-change (y-axis) when the ITS are analysed with OLS (A) and REML (B) using fixed-effect (red circles), DL+WT (green diamonds) and DL+HKSJ (brown crosses), REML+WT (purple diamonds) and REML+HKSJ (blue crosses) meta-analysis methods versus slope-change heterogeneity (x-axis). Plots are presented separately by combinations of the number of included studies (rows) and the number of datapoints (columns). Simulation scenarios include a **level-change of 0, slope-change of 0**, level-change heterogeneity of 0, and autocorrelation of zero.

DL, DerSimonian and Laird. dps, datapoints. HKSJ, Hartung-Knapp / Sidik-Jonkman. ITS, interrupted time series. OLS, ordinary least squares. REML, restricted maximum likelihood. WT, Wald-type.

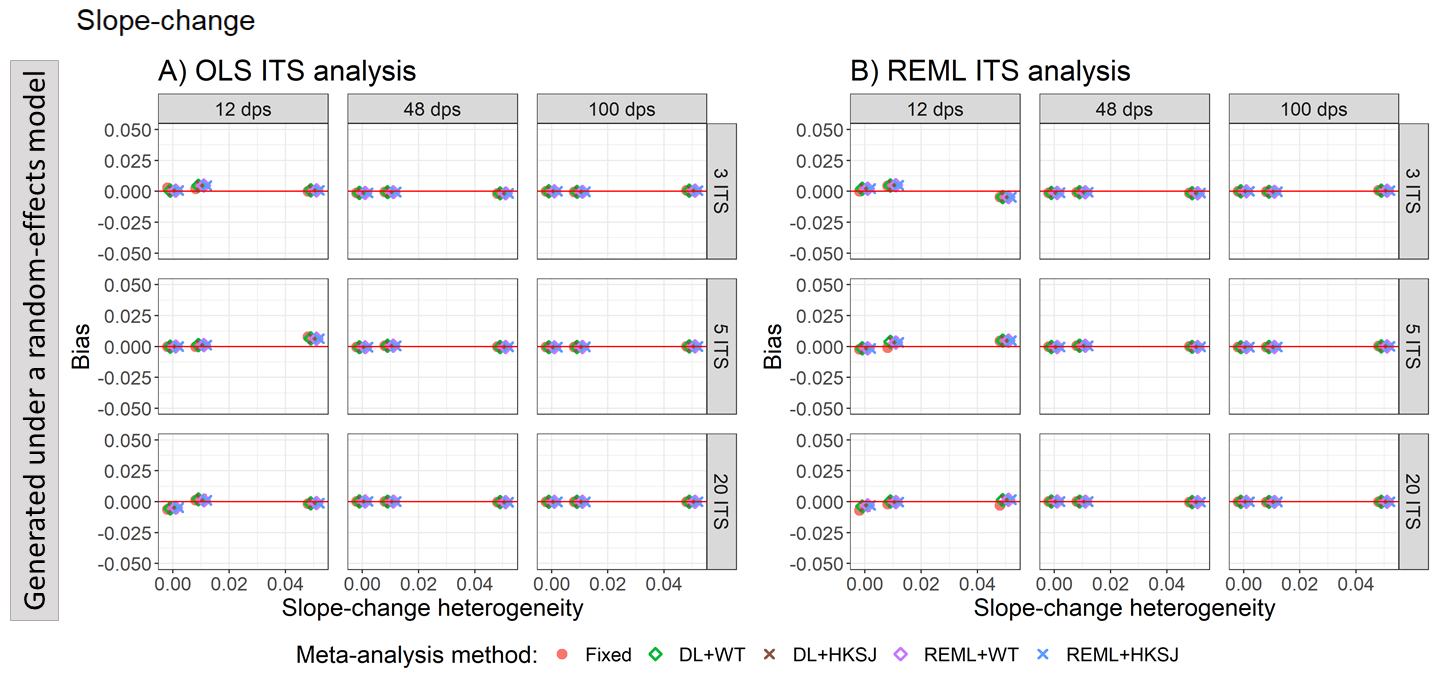

Appendix Figure S63. Plots of the bias of the slope-change (y-axis) when the ITS are analysed with OLS (A) and REML (B) using fixed-effect (red circles), DL+WT (green diamonds) and DL+HKSJ (brown crosses), REML+WT (purple diamonds) and REML+HKSJ (blue crosses) meta-analysis methods versus slope-change heterogeneity (x-axis). Plots are presented separately by combinations of the number of included studies (rows) and the number of datapoints (columns). Simulation scenarios include a **level-change of 0, slope-change of 0.1**, level-change heterogeneity of 0, and autocorrelation of zero.

DL, DerSimonian and Laird. dps, datapoints. HKSJ, Hartung-Knapp / Sidik-Jonkman. ITS, interrupted time series. OLS, ordinary least squares. REML, restricted maximum likelihood. WT, Wald-type.

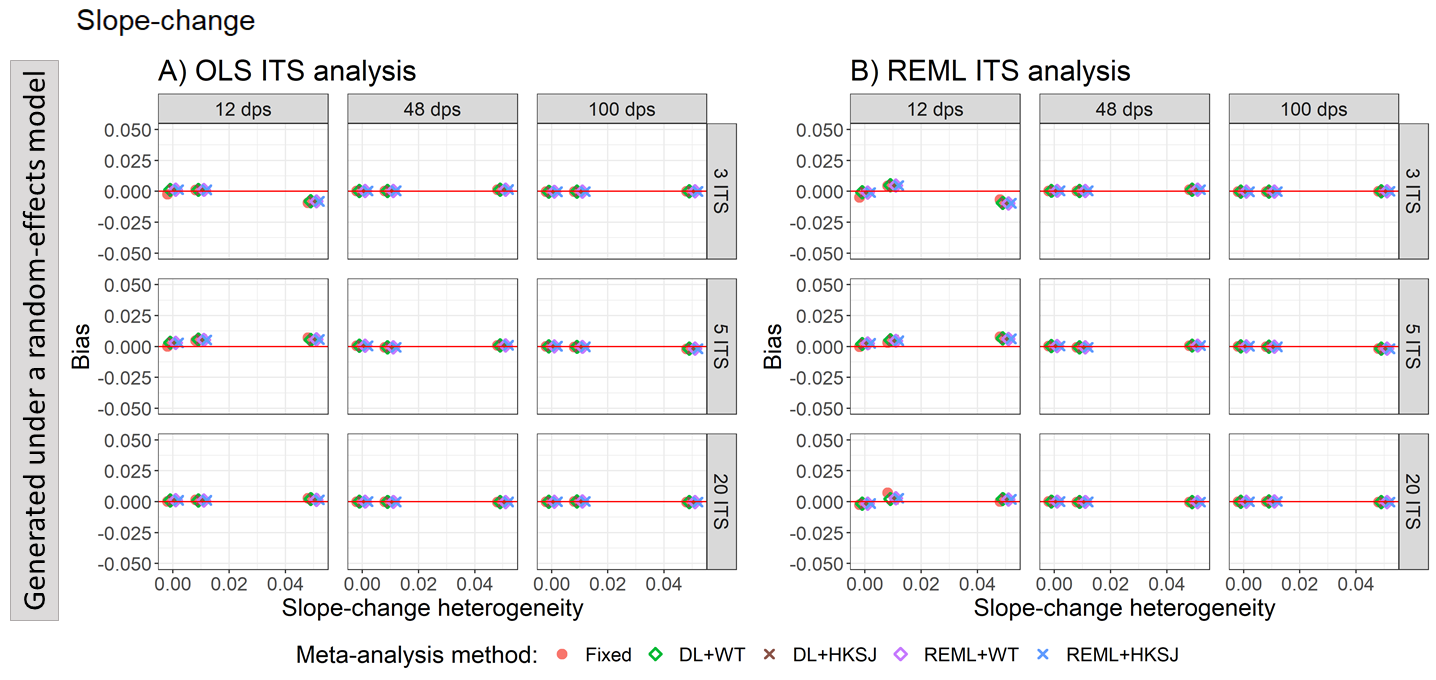

Appendix Figure S64. Plots of the bias of the slope-change (y-axis) when the ITS are analysed with OLS (A) and REML (B) using fixed-effect (red circles), DL+WT (green diamonds) and DL+HKSJ (brown crosses), REML+WT (purple diamonds) and REML+HKSJ (blue crosses) meta-analysis methods versus slope-change heterogeneity (x-axis). Plots are presented separately by combinations of the number of included studies (rows) and the number of datapoints (columns). Simulation scenarios include a **level-change of 1, slope-change of 0**, level-change heterogeneity of 0, and autocorrelation of zero.

DL, DerSimonian and Laird. dps, datapoints. HKSJ, Hartung-Knapp / Sidik-Jonkman. ITS, interrupted time series. OLS, ordinary least squares. REML, restricted maximum likelihood. WT, Wald-type.

#### 4.2 Coverage

##### 4.2.1 Estimation of level-change – coverage vs autocorrelation for scenarios with different level and slope-change combinations

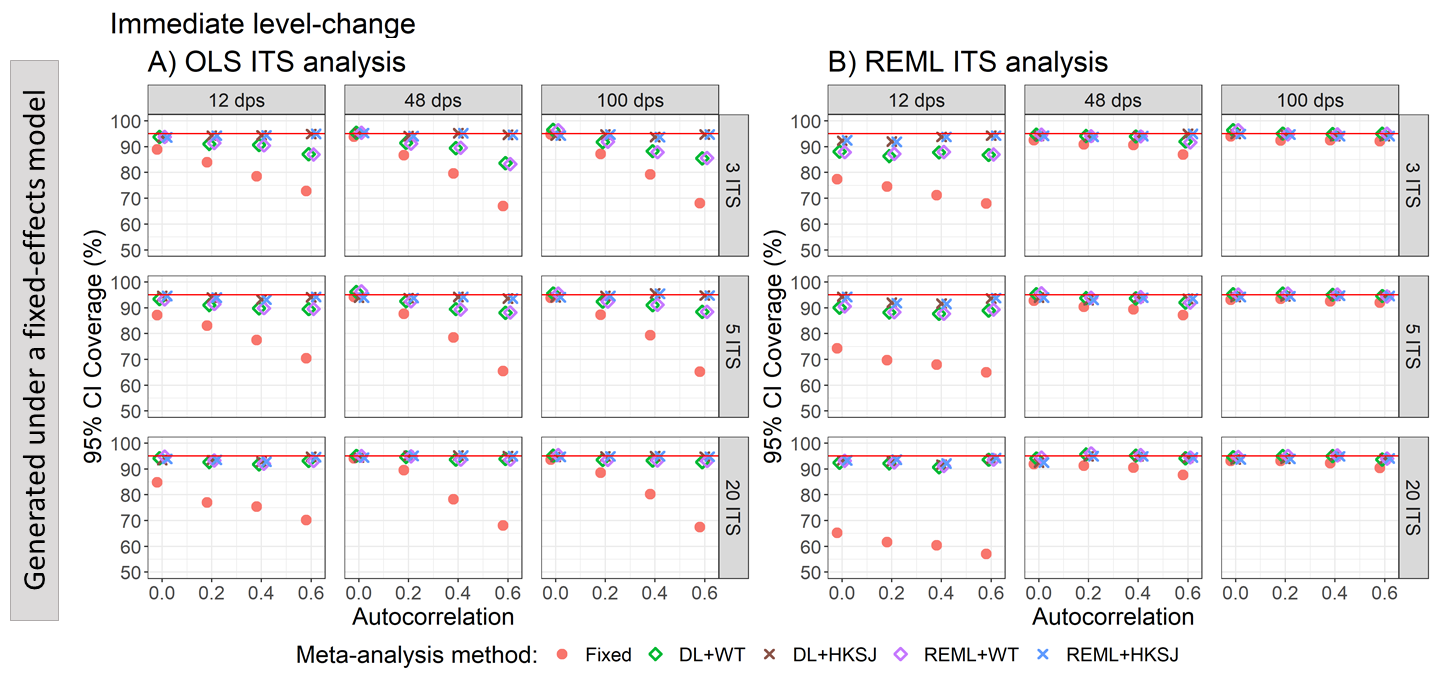

Appendix Figure S65. Plots of 95% confidence interval coverage of level-change (y-axis) when the data was generated under a fixed-effect model and the ITS studies were analysed with OLS (A) and REML (B) using fixed-effect (red circles), DL+WT (green diamonds) and DL+HKSJ (brown crosses), REML+WT (purple diamonds) and REML+HKSJ (blue crosses) meta-analysis methods versus autocorrelation (x-axis). Plots are presented separately by combinations of the number of included studies (rows) and the number of datapoints (columns). The solid red line depicts the nominal 95% coverage level. Simulation scenarios presented include a **level-change of 0,** level-change heterogeneity of 0**, slope-change of 0,** slope-change heterogeneity of 0, and fixed levels of autocorrelation.

DL, DerSimonian and Laird. dps, datapoints. HKSJ, Hartung-Knapp / Sidik-Jonkman. ITS, interrupted time series. OLS, ordinary least squares. REML, restricted maximum likelihood. WT, Wald-type.

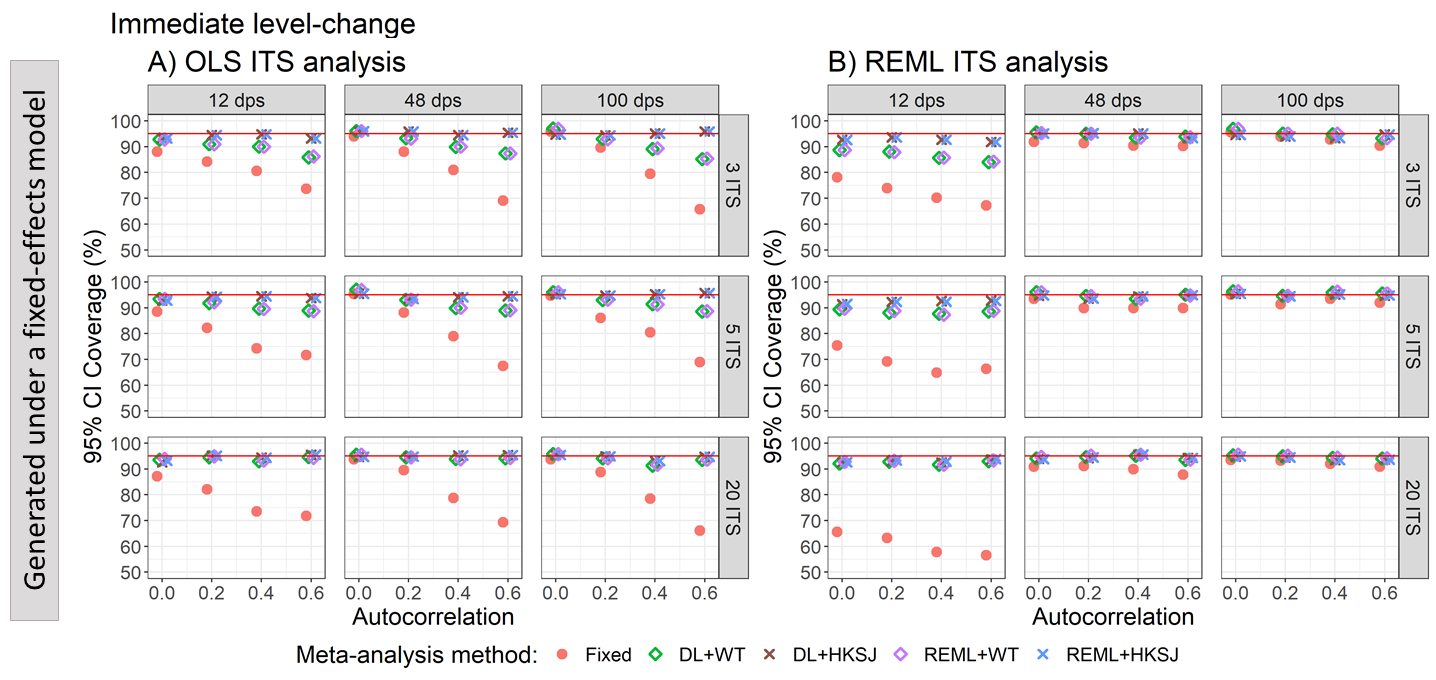

Appendix Figure S66. Plots of 95% confidence interval coverage of level-change (y-axis) when the data was generated under a fixed-effect model and the ITS studies were analysed with OLS (A) and REML (B) using fixed-effect (red circles), DL+WT (green diamonds) and DL+HKSJ (brown crosses), REML+WT (purple diamonds) and REML+HKSJ (blue crosses) meta-analysis methods versus autocorrelation (x-axis). Plots are presented separately by combinations of the number of included studies (rows) and the number of datapoints (columns). The solid red line depicts the nominal 95% coverage level. Simulation scenarios presented include a **level-change of 0,** level-change heterogeneity of 0**, slope-change of 0.1,** slope-change heterogeneity of 0, and fixed levels of autocorrelation.

DL, DerSimonian and Laird. dps, datapoints. HKSJ, Hartung-Knapp / Sidik-Jonkman. ITS, interrupted time series. OLS, ordinary least squares. REML, restricted maximum likelihood. WT, Wald-type.

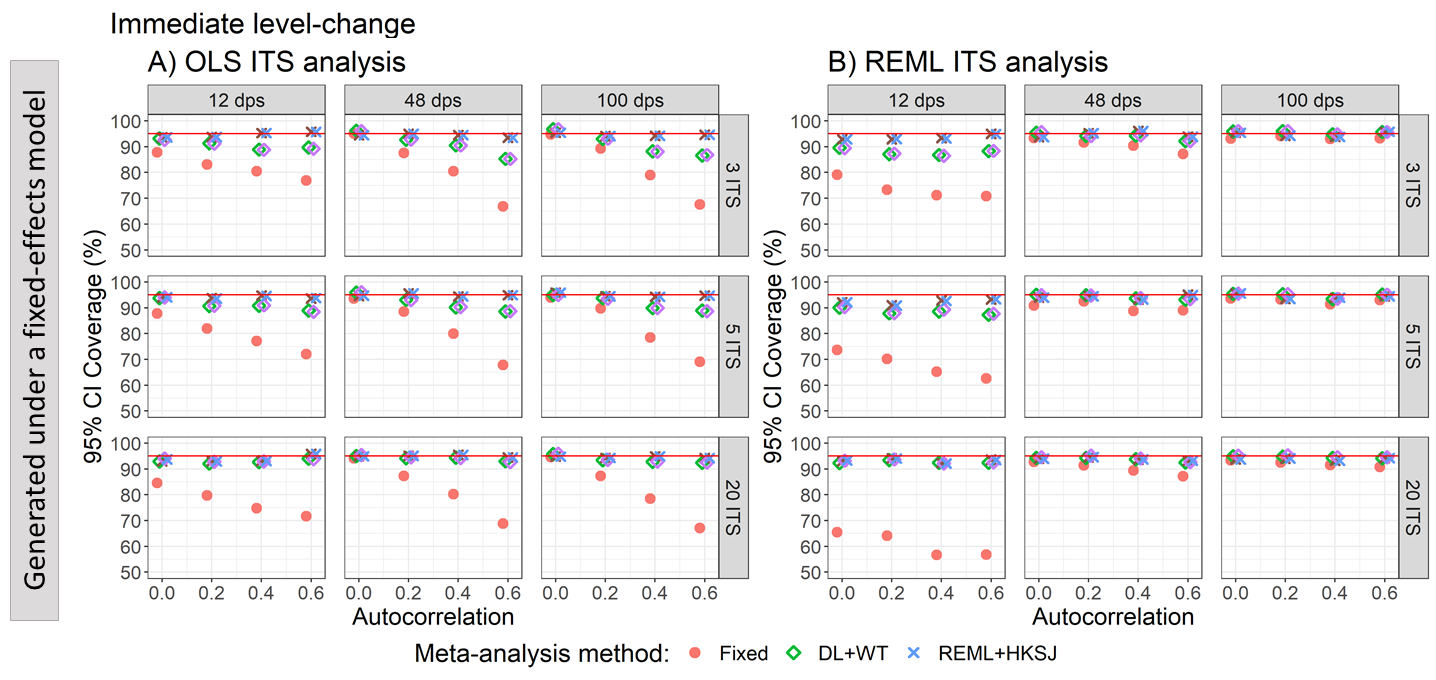

Appendix Figure S67. Plots of 95% confidence interval coverage of level-change (y-axis) when the data was generated under a fixed-effect model and the ITS studies were analysed with OLS (A) and REML (B) using fixed-effect (red circles), DL+WT (green diamonds) and DL+HKSJ (brown crosses), REML+WT (purple diamonds) and REML+HKSJ (blue crosses) meta-analysis methods versus autocorrelation (x-axis). Plots are presented separately by combinations of the number of included studies (rows) and the number of datapoints (columns). The solid red line depicts the nominal 95% coverage level. Simulation scenarios presented include a **level-change of 1,** level-change heterogeneity of 0**, slope-change of 0,** slope-change heterogeneity of 0, and fixed levels of autocorrelation.

DL, DerSimonian and Laird. dps, datapoints. HKSJ, Hartung-Knapp / Sidik-Jonkman. ITS, interrupted time series. OLS, ordinary least squares. REML, restricted maximum likelihood. WT, Wald-type.

##### 4.2.2 Estimation of level-change – coverage vs level-change heterogeneity for scenarios with different level and slope-change combinations

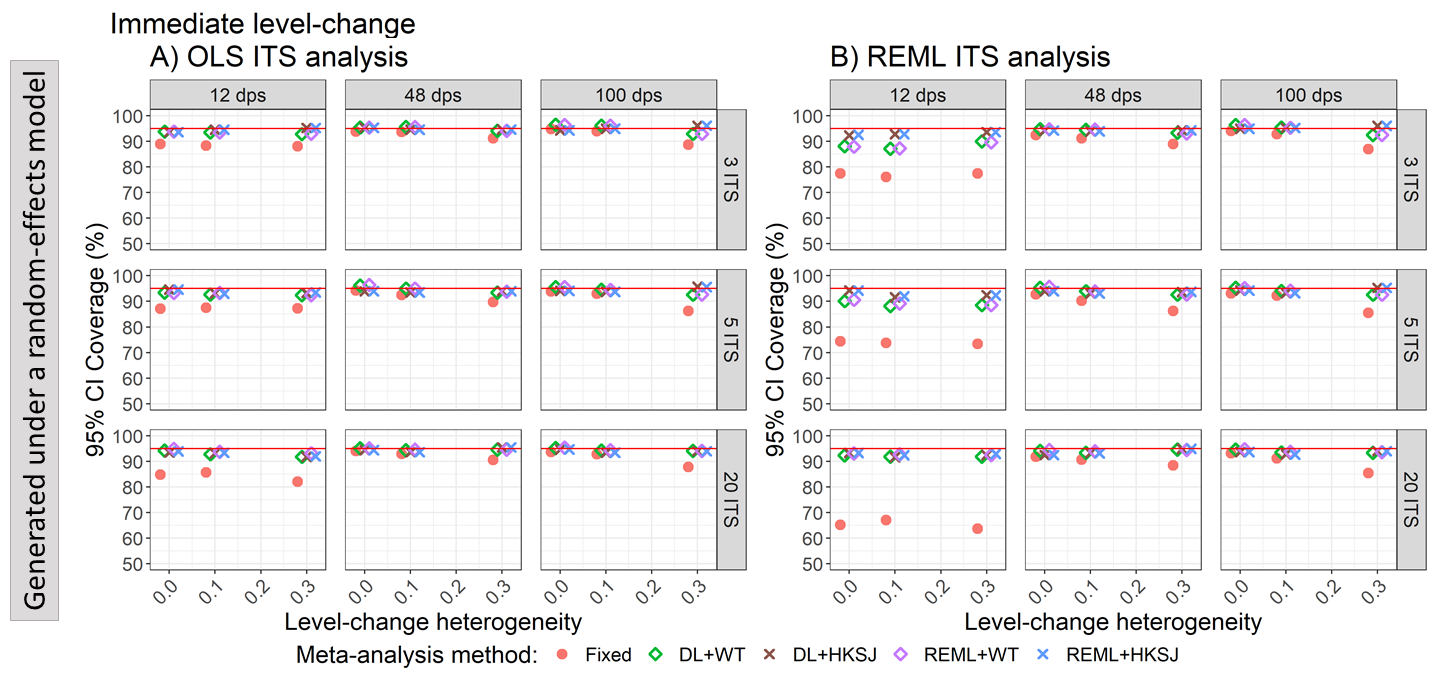

Appendix Figure S68. Plots of 95% confidence interval coverage of level-change (y-axis) when the ITS studies were analysed with OLS (A) and REML (B) using fixed-effect (red circles), DL+WT (green diamonds) and DL+HKSJ (brown crosses), REML+WT (purple diamonds) and REML+HKSJ (blue crosses) meta-analysis methods versus level-change heterogeneity (x-axis). Plots are presented separately by combinations of the number of included studies (rows) and number of datapoints (columns). The solid red line depicts the nominal 95% coverage level. Simulation scenarios include a **level-change of 0,** level-change heterogeneity of 0, **slope-change of 0,** and autocorrelation of 0.

DL, DerSimonian and Laird. dps, datapoints. HKSJ, Hartung-Knapp / Sidik-Jonkman. ITS, interrupted time series. OLS, ordinary least squares. REML, restricted maximum likelihood. WT, Wald-type.

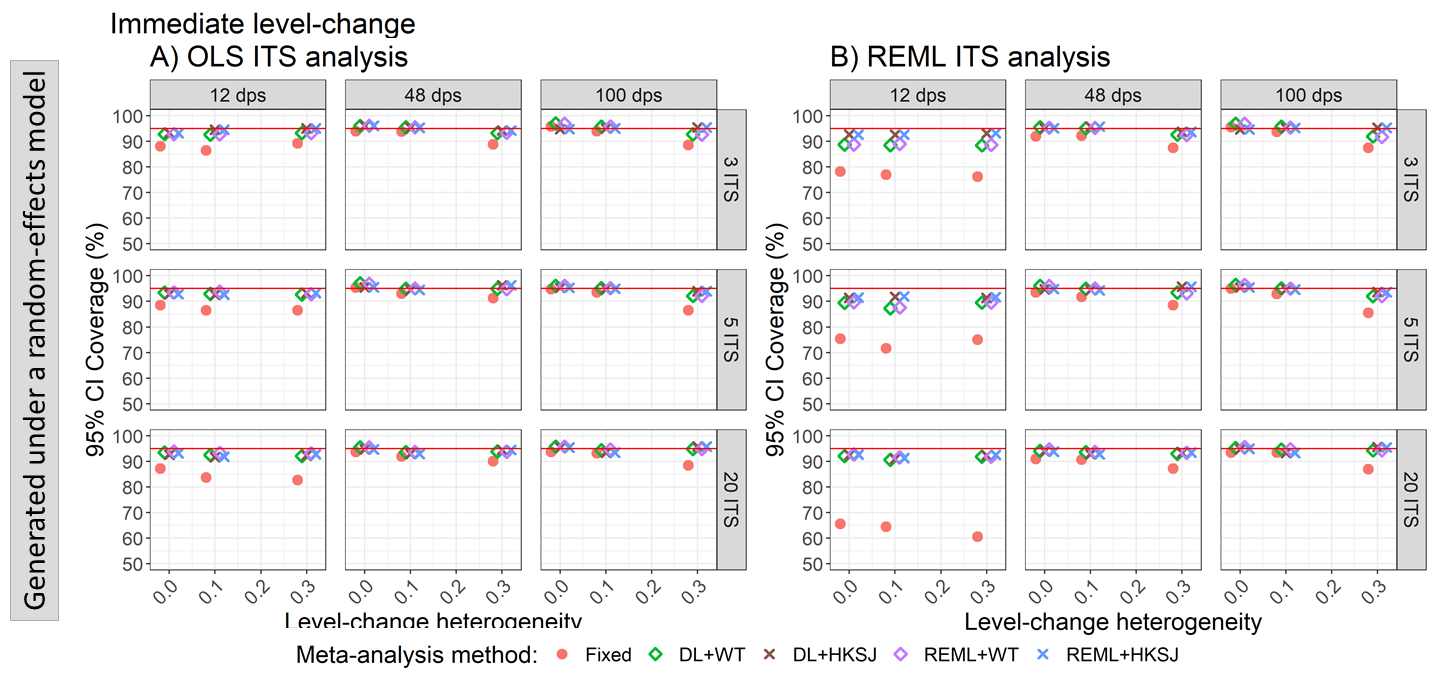

Appendix Figure S9. Plots of 95% confidence interval coverage of level-change (y-axis) when the ITS studies were analysed with OLS (A) and REML (B) using fixed-effect (red circles), DL+WT (green diamonds) and DL+HKSJ (brown crosses), REML+WT (purple diamonds) and REML+HKSJ (blue crosses) meta-analysis methods versus level-change heterogeneity (x-axis). Plots are presented separately by combinations of the number of included studies (rows) and number of datapoints (columns). The solid red line depicts the nominal 95% coverage level. Simulation scenarios include a **level-change of 0,** level-change heterogeneity of 0, **slope-change of 0.1,** and autocorrelation of 0.

DL, DerSimonian and Laird. dps, datapoints. HKSJ, Hartung-Knapp / Sidik-Jonkman. ITS, interrupted time series. OLS, ordinary least squares. REML, restricted maximum likelihood. WT, Wald-type.

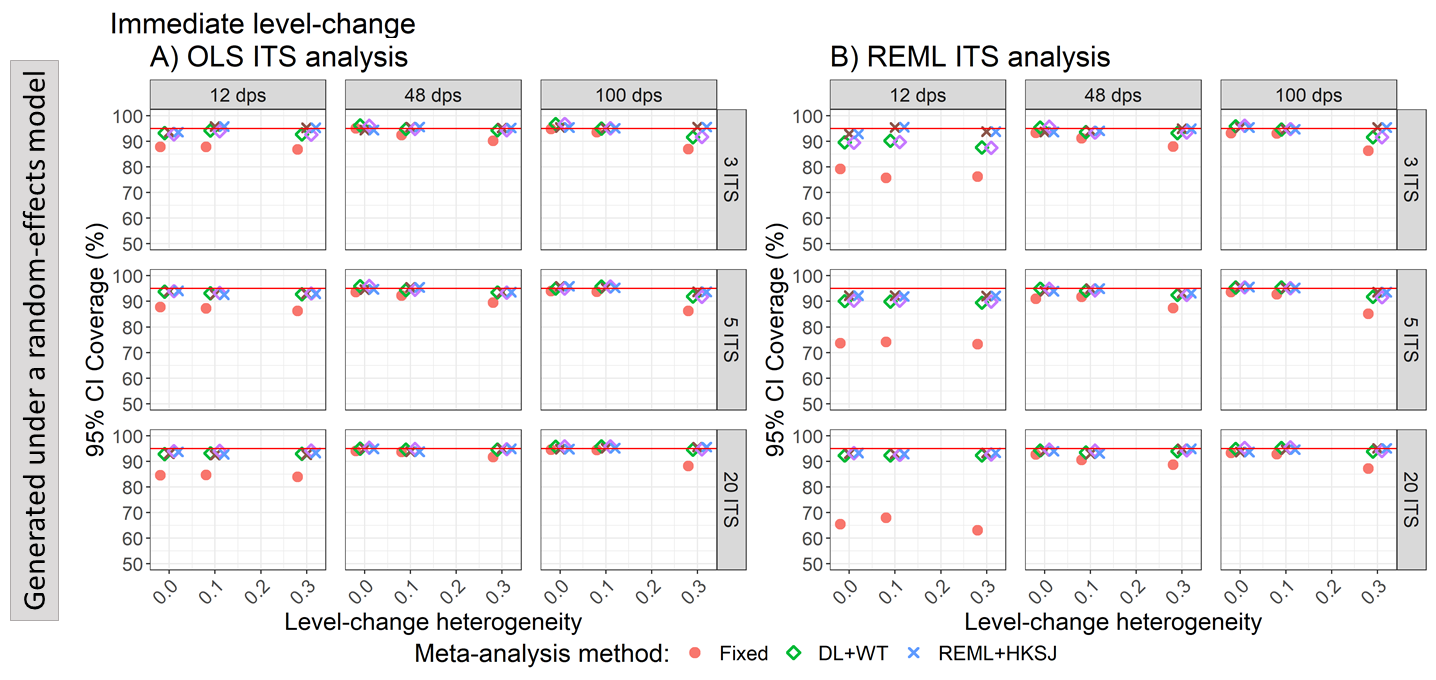

Appendix Figure S69. Plots of 95% confidence interval coverage of level-change (y-axis) when the ITS studies were analysed with OLS (A) and REML (B) using fixed-effect (red circles), DL+WT (green diamonds) and DL+HKSJ (brown crosses), REML+WT (purple diamonds) and REML+HKSJ (blue crosses) meta-analysis methods versus level-change heterogeneity (x-axis). Plots are presented separately by combinations of the number of included studies (rows) and number of datapoints (columns). The solid red line depicts the nominal 95% coverage level. Simulation scenarios include a **level-change of 1,** level-change heterogeneity of 0, **slope-change of 0,** and autocorrelation of 0.

DL, DerSimonian and Laird. dps, datapoints. HKSJ, Hartung-Knapp / Sidik-Jonkman. ITS, interrupted time series. OLS, ordinary least squares. REML, restricted maximum likelihood. WT, Wald-type.

##### 4.2.3 Estimation of slope-change – coverage vs autocorrelation for scenarios with different level and slope-change combinations

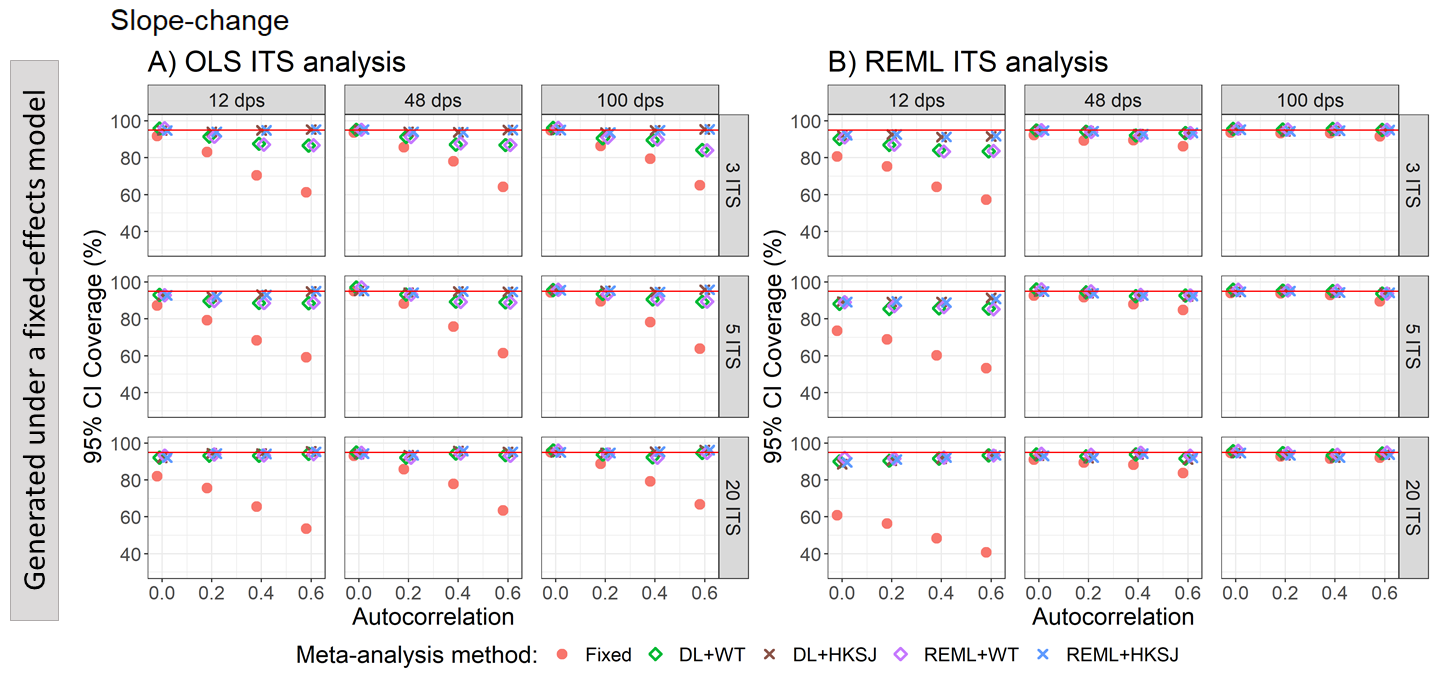

Appendix Figure S70. Plots of 95% confidence interval coverage of slope-change (y-axis) when the data was generated under a fixed-effect model and the ITS studies were analysed with OLS (A) and REML (B) using fixed-effect (red circles), DL+WT (green diamonds) and DL+HKSJ (brown crosses), REML+WT (purple diamonds) and REML+HKSJ (blue crosses) meta-analysis methods versus autocorrelation (x-axis). Plots are presented separately by combinations of the number of included studies (rows) and the number of datapoints (columns). The solid red line depicts the nominal 95% coverage level. Simulation scenarios presented include a **level-change of 0**, level-change heterogeneity of 0, **slope-change of 0**, slope-change heterogeneity of 0, and fixed levels of autocorrelation.

DL, DerSimonian and Laird. dps, datapoints. HKSJ, Hartung-Knapp / Sidik-Jonkman. ITS, interrupted time series. OLS, ordinary least squares. REML, restricted maximum likelihood. WT, Wald-type.

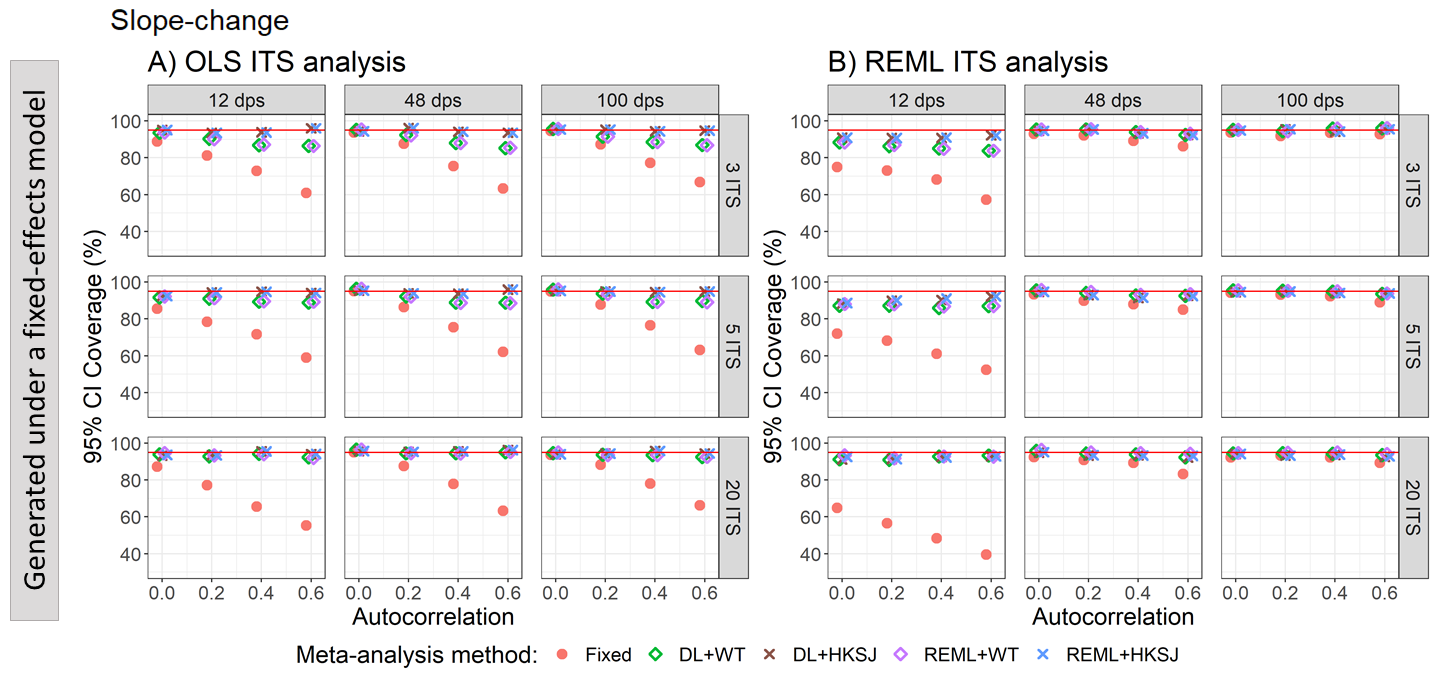

Appendix Figure S71. Plots of 95% confidence interval coverage of slope-change (y-axis) when the data was generated under a fixed-effect model and the ITS studies were analysed with OLS (A) and REML (B) using fixed-effect (red circles), DL+WT (green diamonds) and DL+HKSJ (brown crosses), REML+WT (purple diamonds) and REML+HKSJ (blue crosses) meta-analysis methods versus autocorrelation (x-axis). Plots are presented separately by combinations of the number of included studies (rows) and the number of datapoints (columns). The solid red line depicts the nominal 95% coverage level. Simulation scenarios presented include a **level-change of 0**, level-change heterogeneity of 0, **slope-change of 0.1**, slope-change heterogeneity of 0, and fixed levels of autocorrelation.

DL, DerSimonian and Laird. dps, datapoints. HKSJ, Hartung-Knapp / Sidik-Jonkman. ITS, interrupted time series. OLS, ordinary least squares. REML, restricted maximum likelihood. WT, Wald-type.

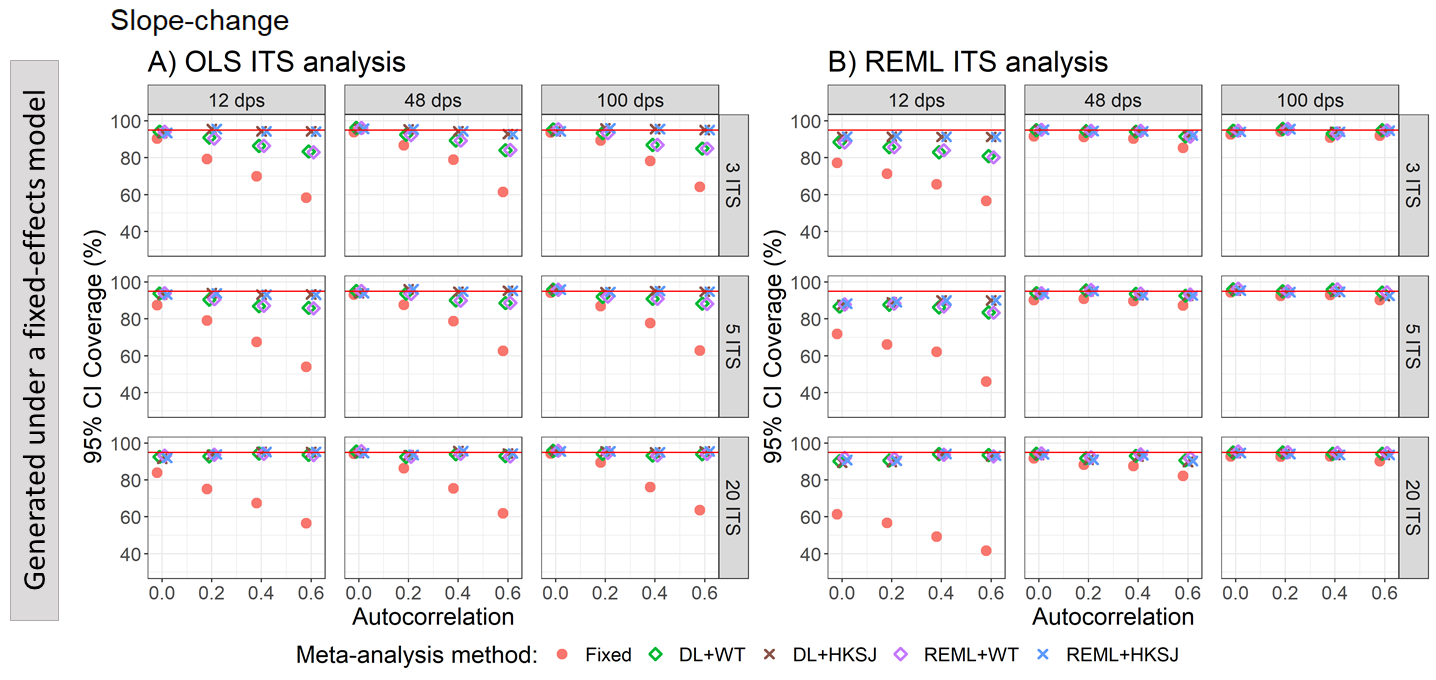

Appendix Figure S72. Plots of 95% confidence interval coverage of slope-change (y-axis) when the data was generated under a fixed-effect model and the ITS studies were analysed with OLS (A) and REML (B) using fixed-effect (red circles), DL+WT (green diamonds) and DL+HKSJ (brown crosses), REML+WT (purple diamonds) and REML+HKSJ (blue crosses) meta-analysis methods versus autocorrelation (x-axis). Plots are presented separately by combinations of the number of included studies (rows) and the number of datapoints (columns). The solid red line depicts the nominal 95% coverage level. Simulation scenarios presented include a **level-change of 1**, level-change heterogeneity of 0, **slope-change of 0**, slope-change heterogeneity of 0, and fixed levels of autocorrelation.

DL, DerSimonian and Laird. dps, datapoints. HKSJ, Hartung-Knapp / Sidik-Jonkman. ITS, interrupted time series. OLS, ordinary least squares. REML, restricted maximum likelihood. WT, Wald-type.

##### 4.2.4 Estimation of slope-change – coverage vs slope-change heterogeneity for scenarios with different level and slope-change combinations

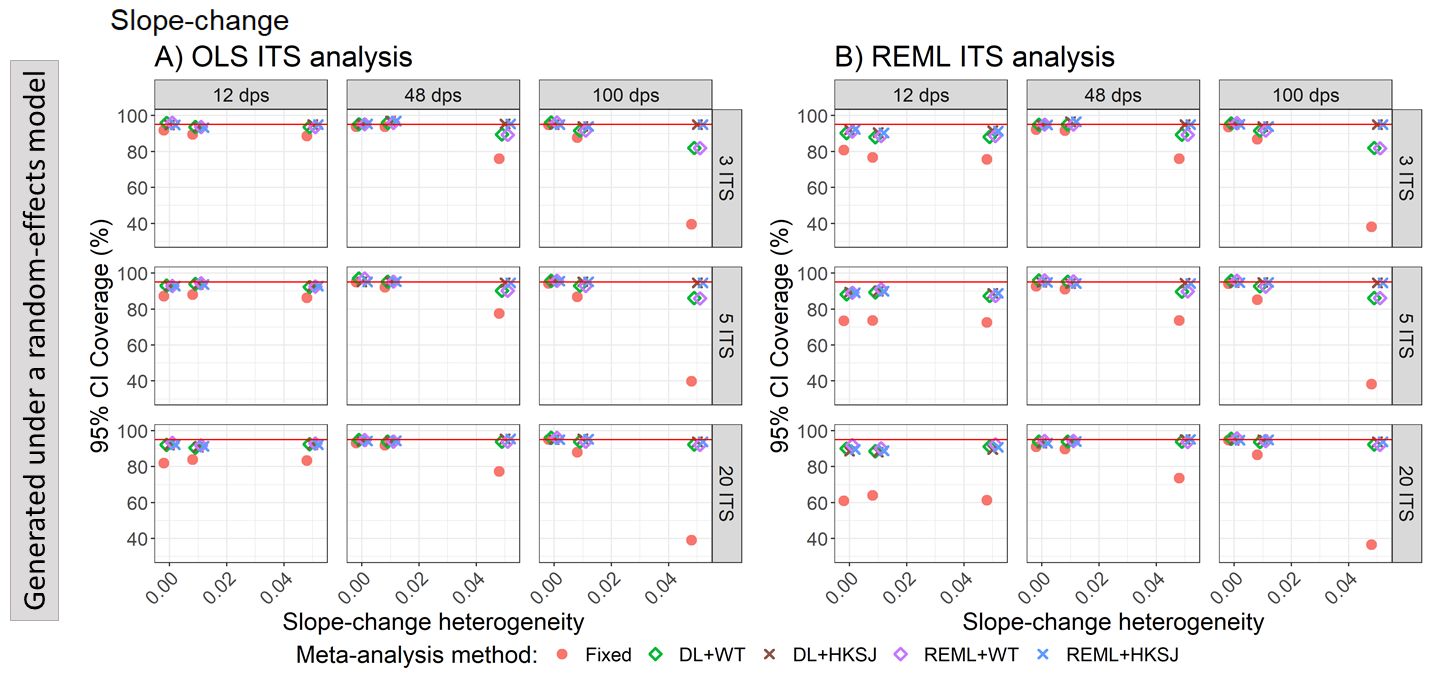

Appendix Figure S73. Plots of 95% confidence interval coverage of slope-change (y-axis) when the ITS studies were analysed with OLS (A) and REML (B) using fixed-effect (red circles), DL+WT (green diamonds) and DL+HKSJ (brown crosses), REML+WT (purple diamonds) and REML+HKSJ (blue crosses) meta-analysis methods versus slope-change heterogeneity (x-axis). Plots are presented separately by combinations of the number of included studies (rows) and number of datapoints (columns). The solid red line depicts the nominal 95% coverage level. Simulation scenarios include a **level-change of 0**, level-change heterogeneity of 0, **slope-change of 0**, and autocorrelation of 0.

DL, DerSimonian and Laird. dps, datapoints. HKSJ, Hartung-Knapp / Sidik-Jonkman. ITS, interrupted time series. OLS, ordinary least squares. REML, restricted maximum likelihood. WT, Wald-type.

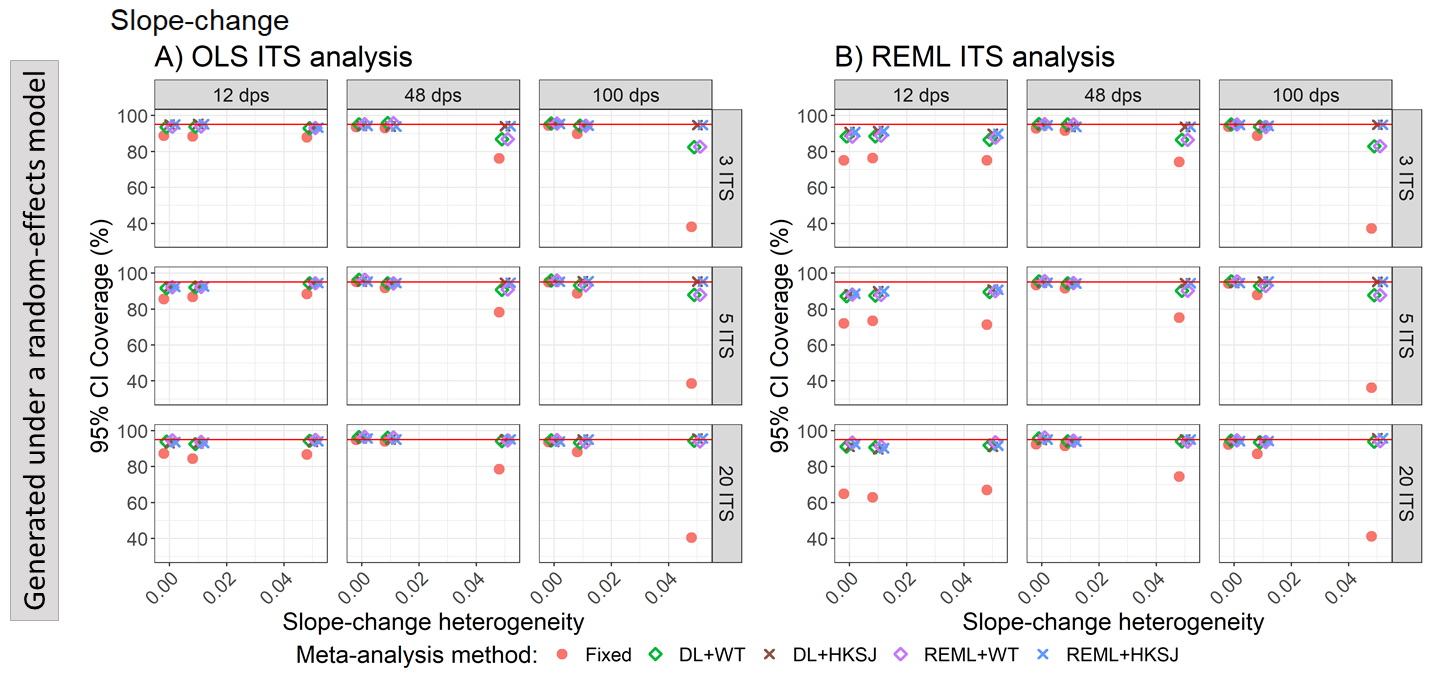

Appendix Figure S74. Plots of 95% confidence interval coverage of slope-change (y-axis) when the ITS studies were analysed with OLS (A) and REML (B) using fixed-effect (red circles), DL+WT (green diamonds) and DL+HKSJ (brown crosses), REML+WT (purple diamonds) and REML+HKSJ (blue crosses) meta-analysis methods versus slope-change heterogeneity (x-axis). Plots are presented separately by combinations of the number of included studies (rows) and number of datapoints (columns). The solid red line depicts the nominal 95% coverage level. Simulation scenarios include a **level-change of 0**, level-change heterogeneity of 0, **slope-change of 0.1**, and autocorrelation of 0.

DL, DerSimonian and Laird. dps, datapoints. HKSJ, Hartung-Knapp / Sidik-Jonkman. ITS, interrupted time series. OLS, ordinary least squares. REML, restricted maximum likelihood. WT, Wald-type.

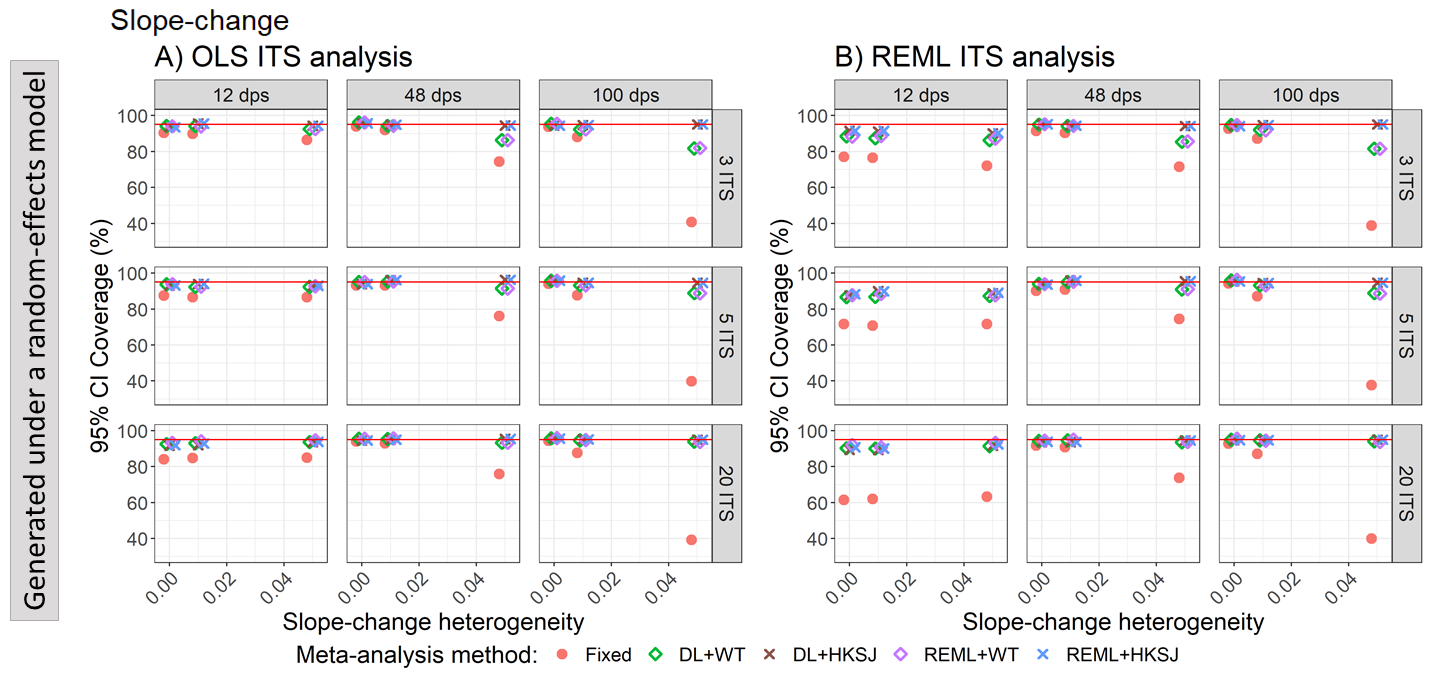

Appendix Figure S75. Plots of 95% confidence interval coverage of slope-change (y-axis) when the ITS studies were analysed with OLS (A) and REML (B) using fixed-effect (red circles), DL+WT (green diamonds) and DL+HKSJ (brown crosses), REML+WT (purple diamonds) and REML+HKSJ (blue crosses) meta-analysis methods versus slope-change heterogeneity (x-axis). Plots are presented separately by combinations of the number of included studies (rows) and number of datapoints (columns). The solid red line depicts the nominal 95% coverage level. Simulation scenarios include a **level-change of 1**, level-change heterogeneity of 0, **slope-change of 0**, and autocorrelation of 0.

DL, DerSimonian and Laird. dps, datapoints. HKSJ, Hartung-Knapp / Sidik-Jonkman. ITS, interrupted time series. OLS, ordinary least squares. REML, restricted maximum likelihood. WT, Wald-type.

#### 4.3 Empirical standard errors

##### 4.3.1 Estimation of level-change – empirical standard errors vs autocorrelation for scenarios with different level and slope-change combinations

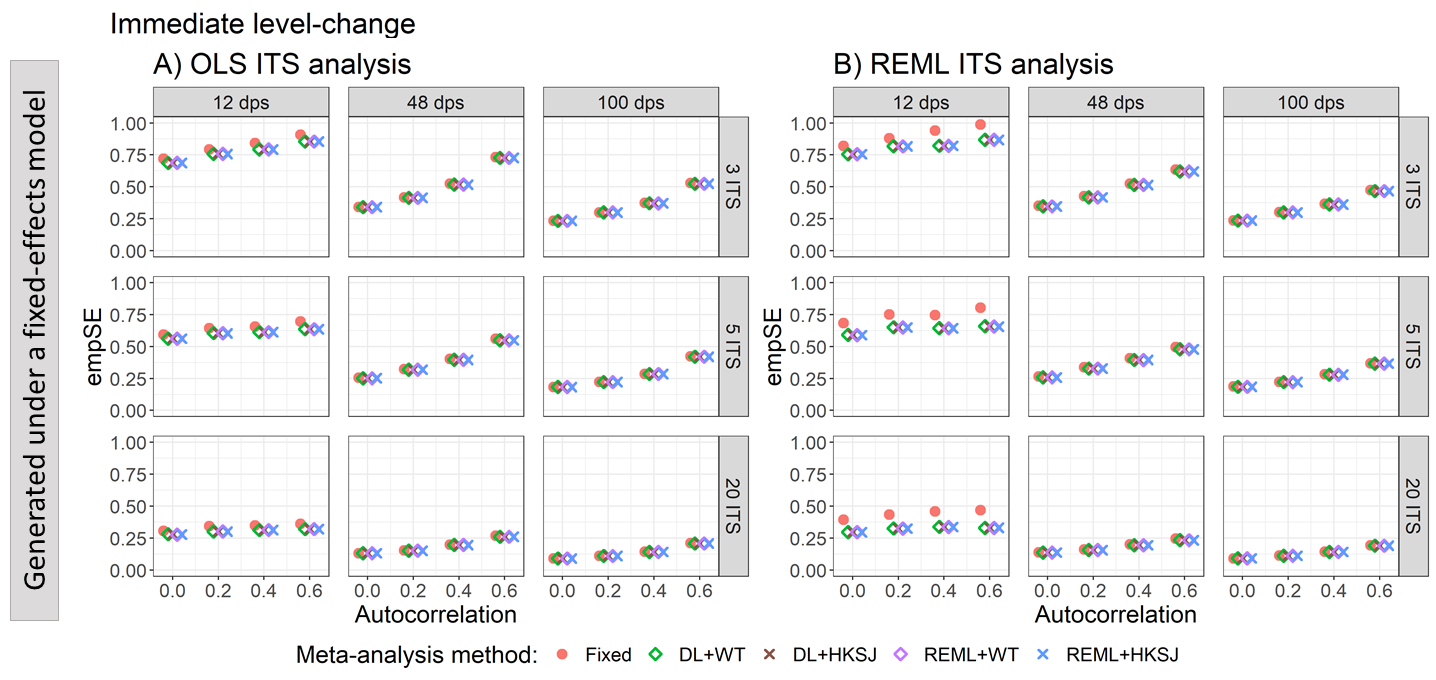

Appendix Figure S76. Plots of empirical standard error (empSE) of the immediate level-change (y-axis) when the data was generated under a fixed-effect model and the ITS studies were analysed with OLS (A) and REML (B) using fixed-effect (red circles), DL+WT (green diamonds) and DL+HKSJ (brown crosses), REML+WT (purple diamonds) and REML+HKSJ (blue crosses) meta-analysis methods versus autocorrelation (x-axis). Plots are presented separately by combinations of the number of included studies (rows) and the number of datapoints (columns). Simulation scenarios include a **level-change of 0,** level-change heterogeneity of 0, **slope-change of 0**, slope-change heterogeneity of 0, and fixed levels of autocorrelation.

DL, DerSimonian and Laird. dps, datapoints. HKSJ, Hartung-Knapp / Sidik-Jonkman. ITS, interrupted time series. OLS, ordinary least squares. REML, restricted maximum likelihood. WT, Wald-type.

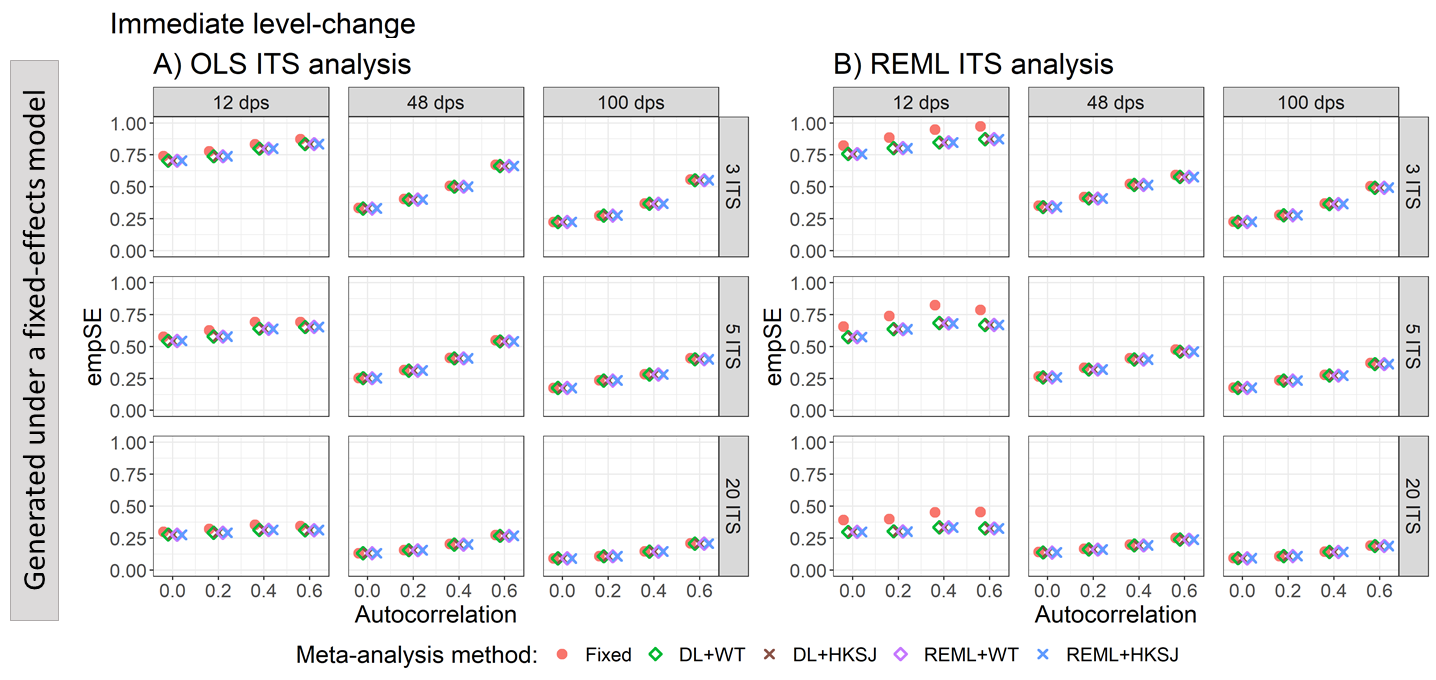

Appendix Figure S77. Plots of empirical standard error (empSE) of the immediate level-change (y-axis) when the data was generated under a fixed-effect model and the ITS studies were analysed with OLS (A) and REML (B) using fixed-effect (red circles), DL+WT (green diamonds) and DL+HKSJ (brown crosses), REML+WT (purple diamonds) and REML+HKSJ (blue crosses) meta-analysis methods versus autocorrelation (x-axis). Plots are presented separately by combinations of the number of included studies (rows) and the number of datapoints (columns). Simulation scenarios include a **level-change of 0,** level-change heterogeneity of 0, **slope-change of 0.1**, slope-change heterogeneity of 0, and fixed levels of autocorrelation.

DL, DerSimonian and Laird. dps, datapoints. HKSJ, Hartung-Knapp / Sidik-Jonkman. ITS, interrupted time series. OLS, ordinary least squares. REML, restricted maximum likelihood. WT, Wald-type.

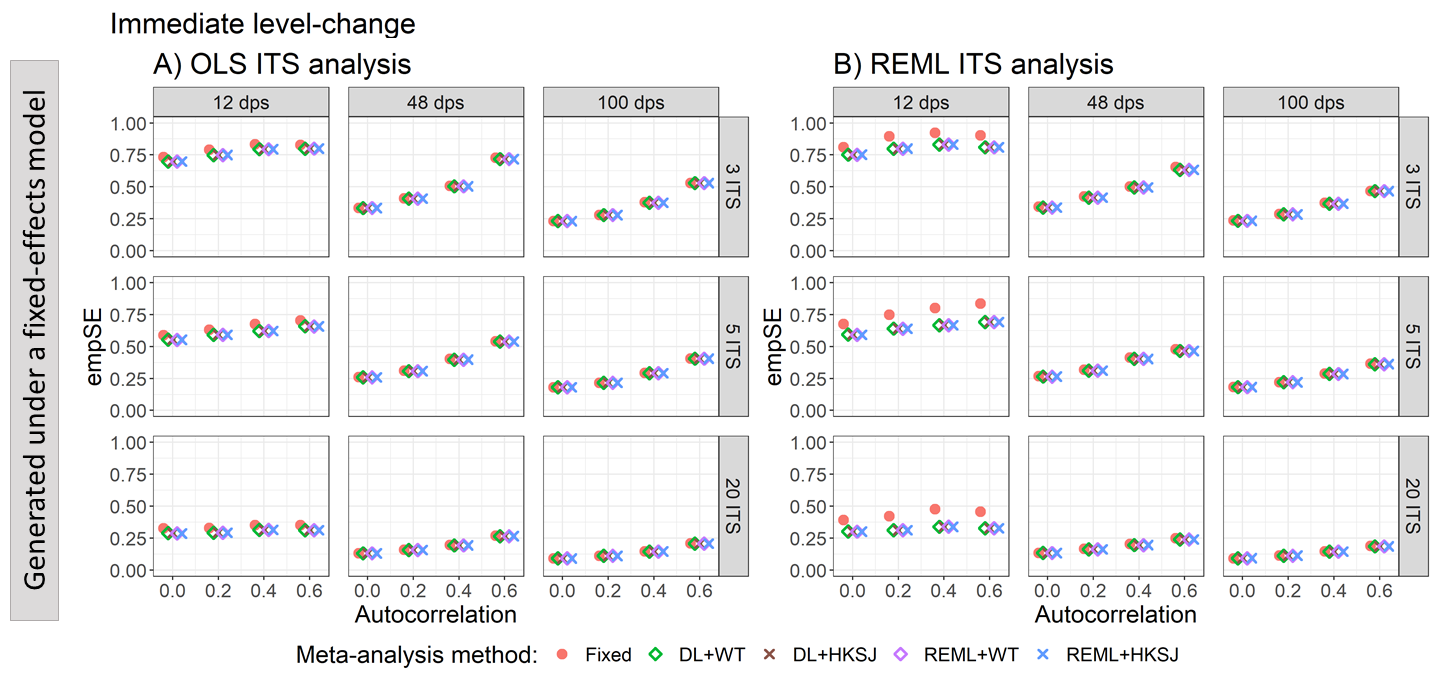

Appendix Figure S78. Plots of empirical standard error (empSE) of the immediate level-change (y-axis) when the data was generated under a fixed-effect model and the ITS studies were analysed with OLS (A) and REML (B) using fixed-effect (red circles), DL+WT (green diamonds) and DL+HKSJ (brown crosses), REML+WT (purple diamonds) and REML+HKSJ (blue crosses) meta-analysis methods versus autocorrelation (x-axis). Plots are presented separately by combinations of the number of included studies (rows) and the number of datapoints (columns). Simulation scenarios include a **level-change of 1,** level-change heterogeneity of 0, **slope-change of 0**, slope-change heterogeneity of 0, and fixed levels of autocorrelation.

DL, DerSimonian and Laird. dps, datapoints. HKSJ, Hartung-Knapp / Sidik-Jonkman. ITS, interrupted time series. OLS, ordinary least squares. REML, restricted maximum likelihood. WT, Wald-type.

##### 4.3.2 Estimation of level-change – empirical standard errors vs level-change heterogeneity for scenarios with different level and slope-change combinations

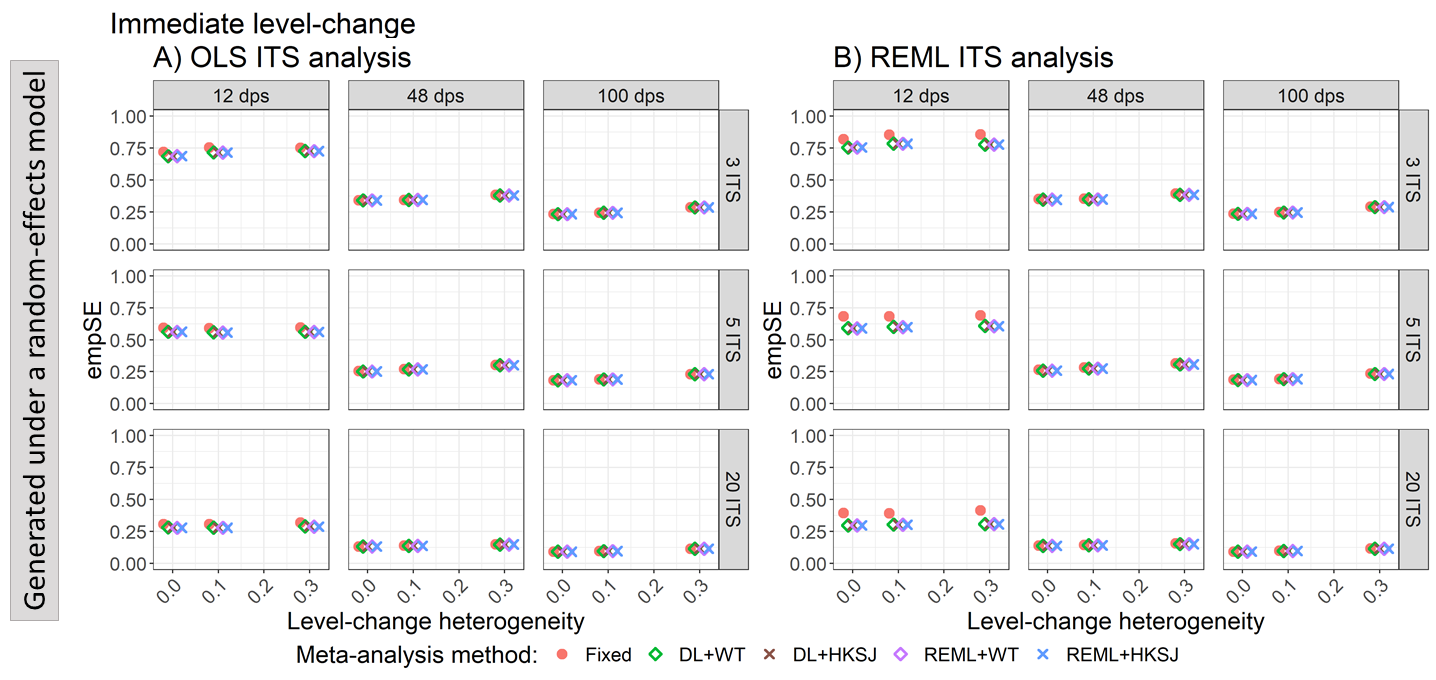

Appendix Figure S79. Plots of empirical standard error (empSE) of the immediate level-change (y-axis) when the ITS are analysed with OLS (A) and REML (B) using fixed-effect (red circles), DL+WT (green diamonds) and DL+HKSJ (brown crosses), REML+WT (purple diamonds) and REML+HKSJ (blue crosses) meta-analysis methods versus level-change heterogeneity (x-axis). Plots are presented separately by combinations of the number of included studies (rows) and the number of datapoints (columns). Simulation scenarios include a **level-change of 0, slope-change of 0,** slope-change heterogeneity of 0, and autocorrelation of 0.

DL, DerSimonian and Laird. dps, datapoints. HKSJ, Hartung-Knapp / Sidik-Jonkman. ITS, interrupted time series. OLS, ordinary least squares. REML, restricted maximum likelihood. WT, Wald-type.

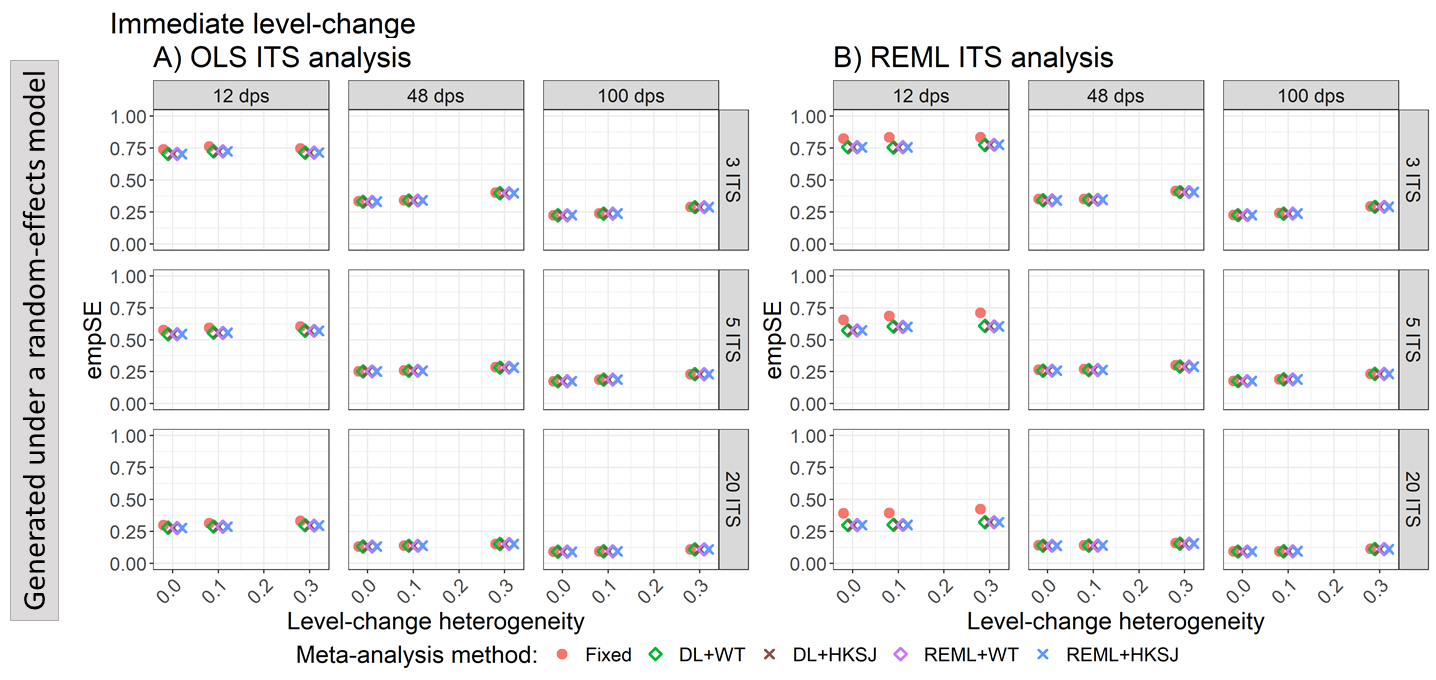

Appendix Figure S80. Plots of empirical standard error (empSE) of the immediate level-change (y-axis) when the ITS are analysed with OLS (A) and REML (B) using fixed-effect (red circles), DL+WT (green diamonds) and DL+HKSJ (brown crosses), REML+WT (purple diamonds) and REML+HKSJ (blue crosses) meta-analysis methods versus level-change heterogeneity (x-axis). Plots are presented separately by combinations of the number of included studies (rows) and the number of datapoints (columns). Simulation scenarios include a **level-change of 0, slope-change of 0.1,** slope-change heterogeneity of 0, and autocorrelation of 0.

DL, DerSimonian and Laird. dps, datapoints. HKSJ, Hartung-Knapp / Sidik-Jonkman. ITS, interrupted time series. OLS, ordinary least squares. REML, restricted maximum likelihood. WT, Wald-type.

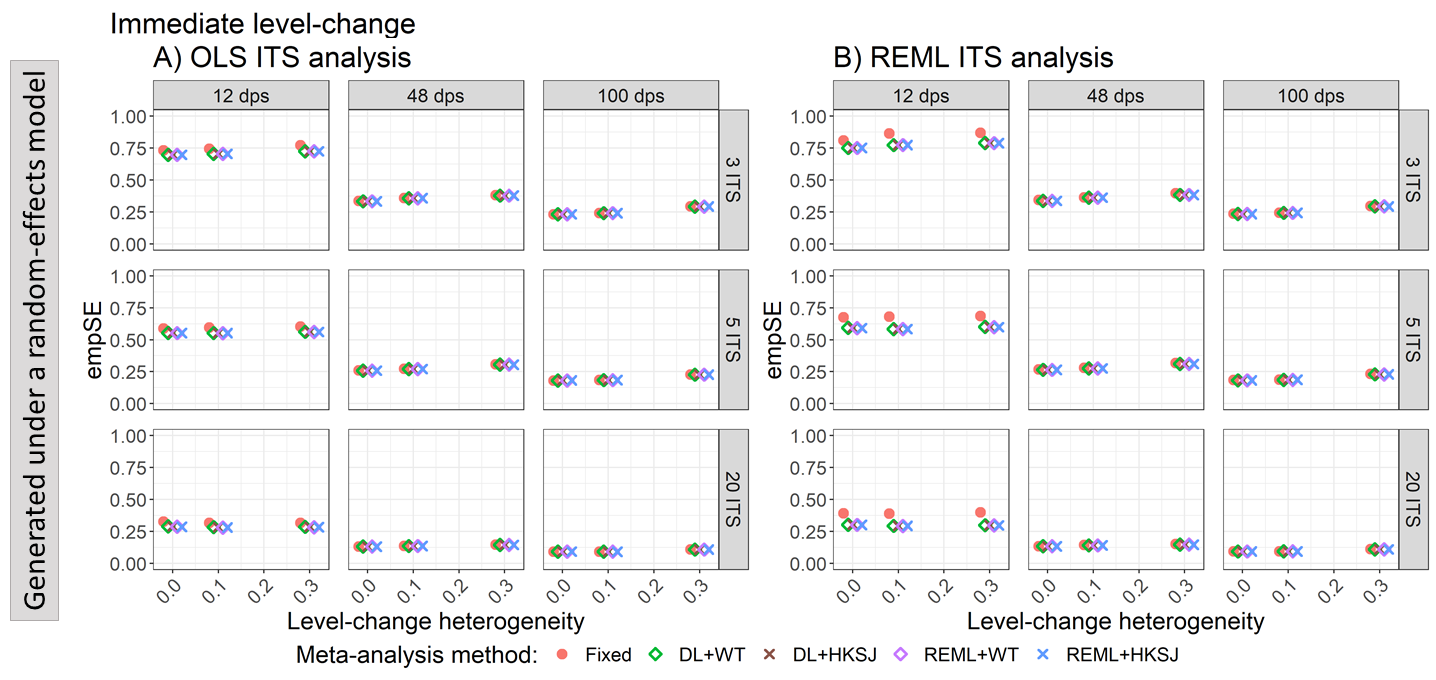

Appendix Figure S81. Plots of empirical standard error (empSE) of the immediate level-change (y-axis) when the ITS are analysed with OLS (A) and REML (B) using fixed-effect (red circles), DL+WT (green diamonds) and DL+HKSJ (brown crosses), REML+WT (purple diamonds) and REML+HKSJ (blue crosses) meta-analysis methods versus level-change heterogeneity (x-axis). Plots are presented separately by combinations of the number of included studies (rows) and the number of datapoints (columns). Simulation scenarios include a **level-change of 1, slope-change of 0,** slope-change heterogeneity of 0, and autocorrelation of 0.

DL, DerSimonian and Laird. dps, datapoints. HKSJ, Hartung-Knapp / Sidik-Jonkman. ITS, interrupted time series. OLS, ordinary least squares. REML, restricted maximum likelihood. WT, Wald-type.

##### 4.3.3 Estimation of slope-change – empirical standard errors vs autocorrelation for scenarios with different level and slope-change combinations

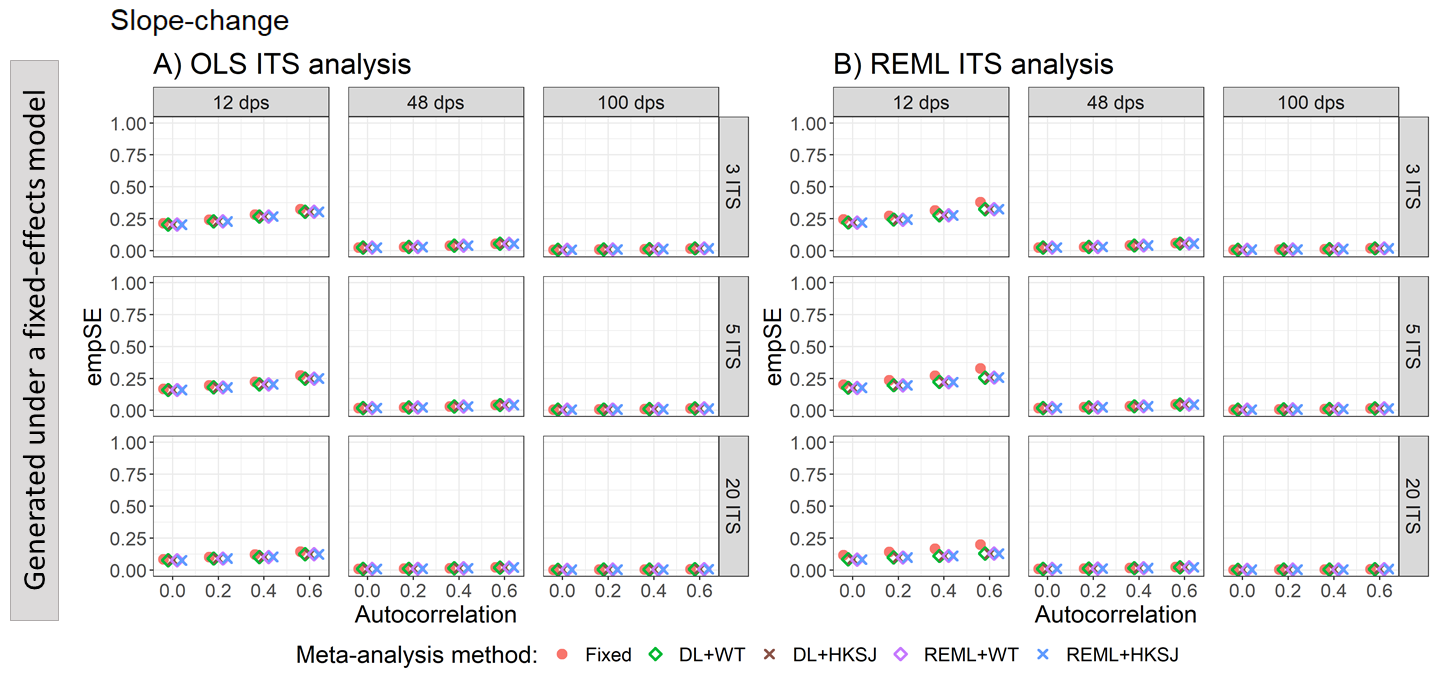

Appendix Figure S82. Plots of empirical standard error (empSE) of the immediate level-change (y-axis) when the data was generated under a fixed-effect model and the ITS studies were analysed with OLS (A) and REML (B) using fixed-effect (red circles), DL+WT (green diamonds) and DL+HKSJ (brown crosses), REML+WT (purple diamonds) and REML+HKSJ (blue crosses) meta-analysis methods versus autocorrelation (x-axis). Plots are presented separately by combinations of the number of included studies (rows) and the number of datapoints (columns). Simulation scenarios include a **level-change of 0,** level-change heterogeneity of 0, **slope-change of 0**, slope-change heterogeneity of 0, and fixed levels of autocorrelation.

DL, DerSimonian and Laird. dps, datapoints. HKSJ, Hartung-Knapp / Sidik-Jonkman. ITS, interrupted time series. OLS, ordinary least squares. REML, restricted maximum likelihood. WT, Wald-type.

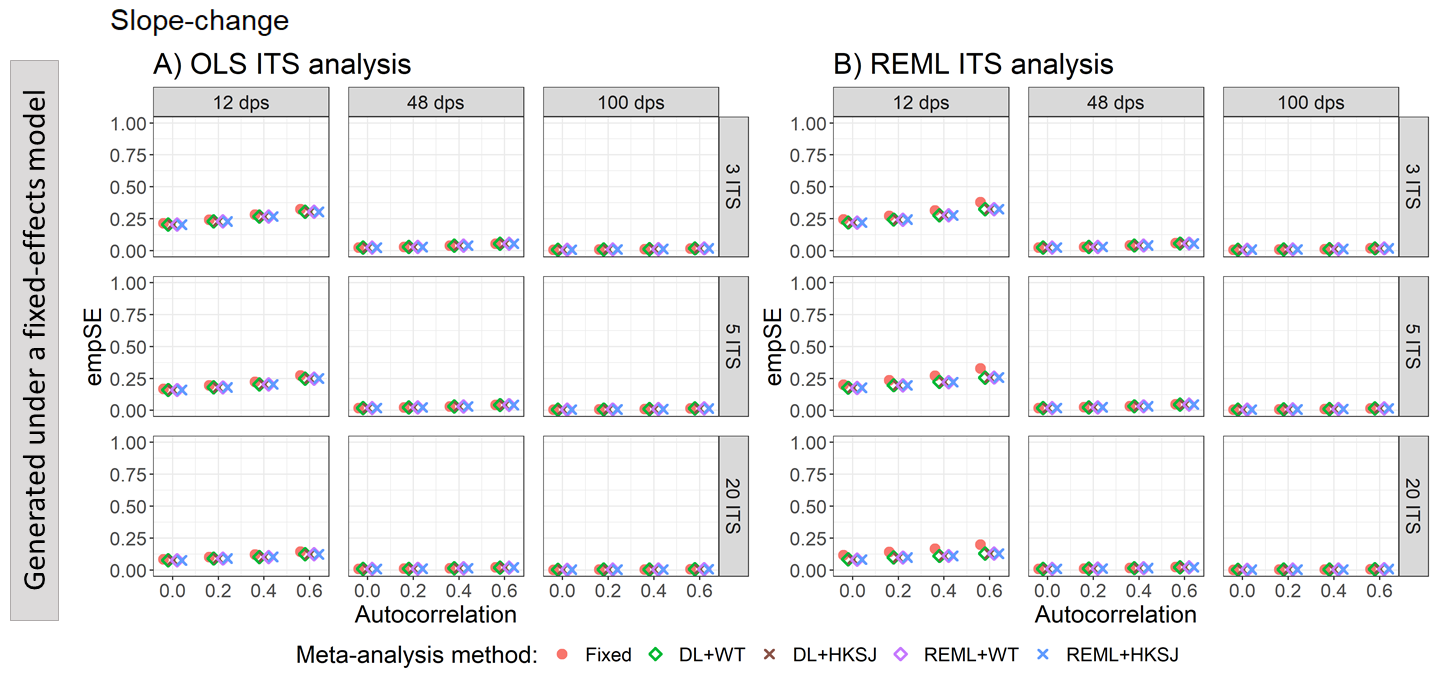

Appendix Figure S83. Plots of empirical standard error (empSE) of the immediate level-change (y-axis) when the data was generated under a fixed-effect model and the ITS studies were analysed with OLS (A) and REML (B) using fixed-effect (red circles), DL+WT (green diamonds) and DL+HKSJ (brown crosses), REML+WT (purple diamonds) and REML+HKSJ (blue crosses) meta-analysis methods versus autocorrelation (x-axis). Plots are presented separately by combinations of the number of included studies (rows) and the number of datapoints (columns). Simulation scenarios include a **level-change of 0,** level-change heterogeneity of 0, **slope-change of 0.1**, slope-change heterogeneity of 0, and fixed levels of autocorrelation.

DL, DerSimonian and Laird. dps, datapoints. HKSJ, Hartung-Knapp / Sidik-Jonkman. ITS, interrupted time series. OLS, ordinary least squares. REML, restricted maximum likelihood. WT, Wald-type.

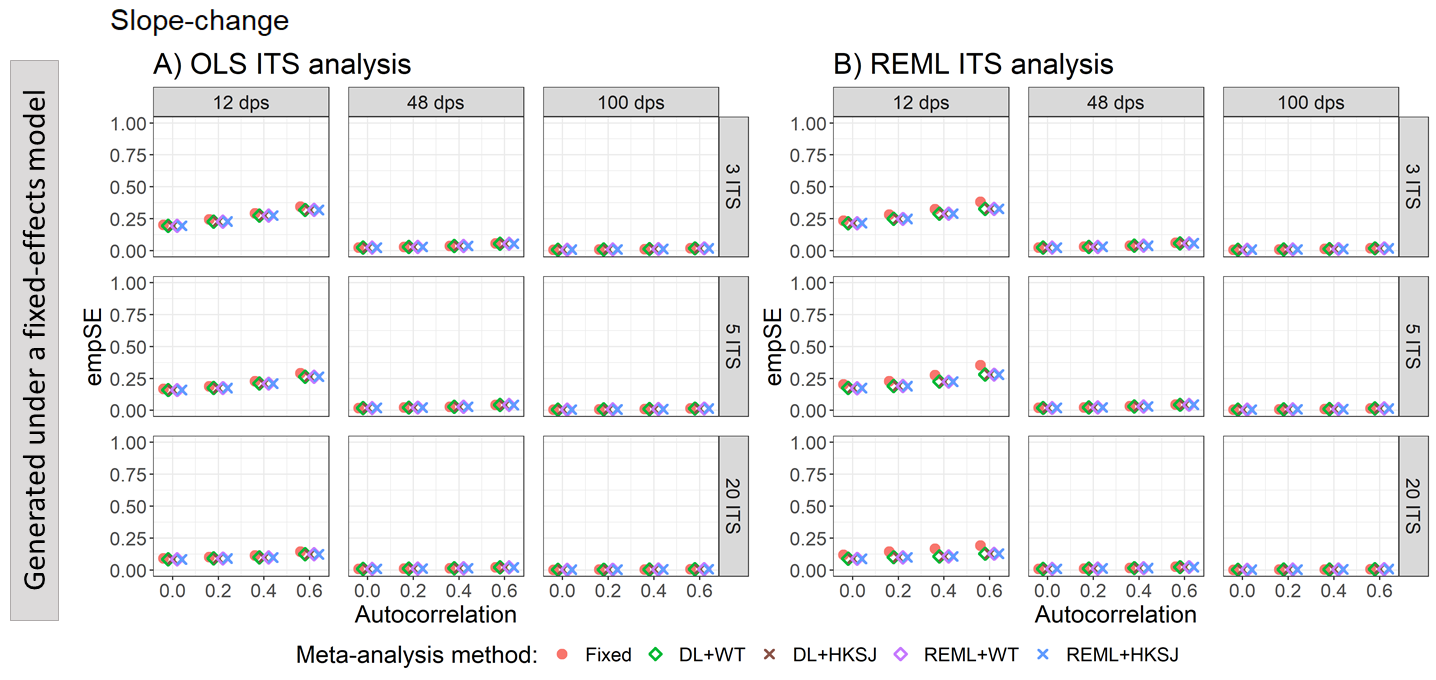

Appendix Figure S84. Plots of empirical standard error (empSE) of the immediate level-change (y-axis) when the data was generated under a fixed-effect model and the ITS studies were analysed with OLS (A) and REML (B) using fixed-effect (red circles), DL+WT (green diamonds) and DL+HKSJ (brown crosses), REML+WT (purple diamonds) and REML+HKSJ (blue crosses) meta-analysis methods versus autocorrelation (x-axis). Plots are presented separately by combinations of the number of included studies (rows) and the number of datapoints (columns). Simulation scenarios include a **level-change of 1,** level-change heterogeneity of 0, **slope-change of 0**, slope-change heterogeneity of 0, and fixed levels of autocorrelation.

DL, DerSimonian and Laird. dps, datapoints. HKSJ, Hartung-Knapp / Sidik-Jonkman. ITS, interrupted time series. OLS, ordinary least squares. REML, restricted maximum likelihood. WT, Wald-type.

##### 4.3.4 Estimation of slope-change – empirical standard errors vs slope-change heterogeneity for scenarios with different level and slope-change combinations

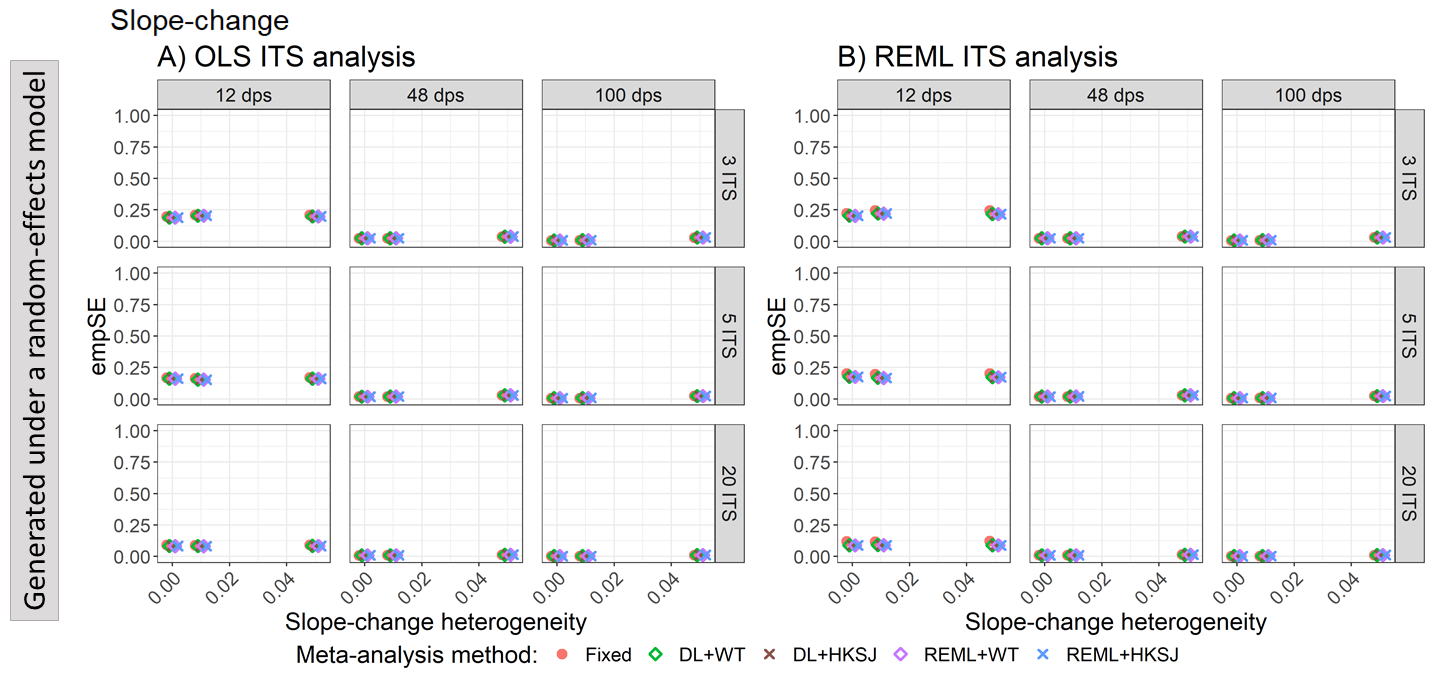

Appendix Figure S85. Plots of empirical standard error (empSE) of the immediate slope-change (y-axis) when the ITS are analysed with OLS (A) and REML (B) using fixed-effect (red circles), DL+WT (green diamonds) and DL+HKSJ (brown crosses), REML+WT (purple diamonds) and REML+HKSJ (blue crosses) meta-analysis methods versus slope-change heterogeneity (x-axis). Plots are presented separately by combinations of the number of included studies (rows) and the number of datapoints (columns). Simulation scenarios include a **level-change of 0**, level-change heterogeneity of 0, **slope-change of 0**, and fixed levels of autocorrelation.

DL, DerSimonian and Laird. dps, datapoints. HKSJ, Hartung-Knapp / Sidik-Jonkman. ITS, interrupted time series. OLS, ordinary least squares. REML, restricted maximum likelihood. WT, Wald-type.

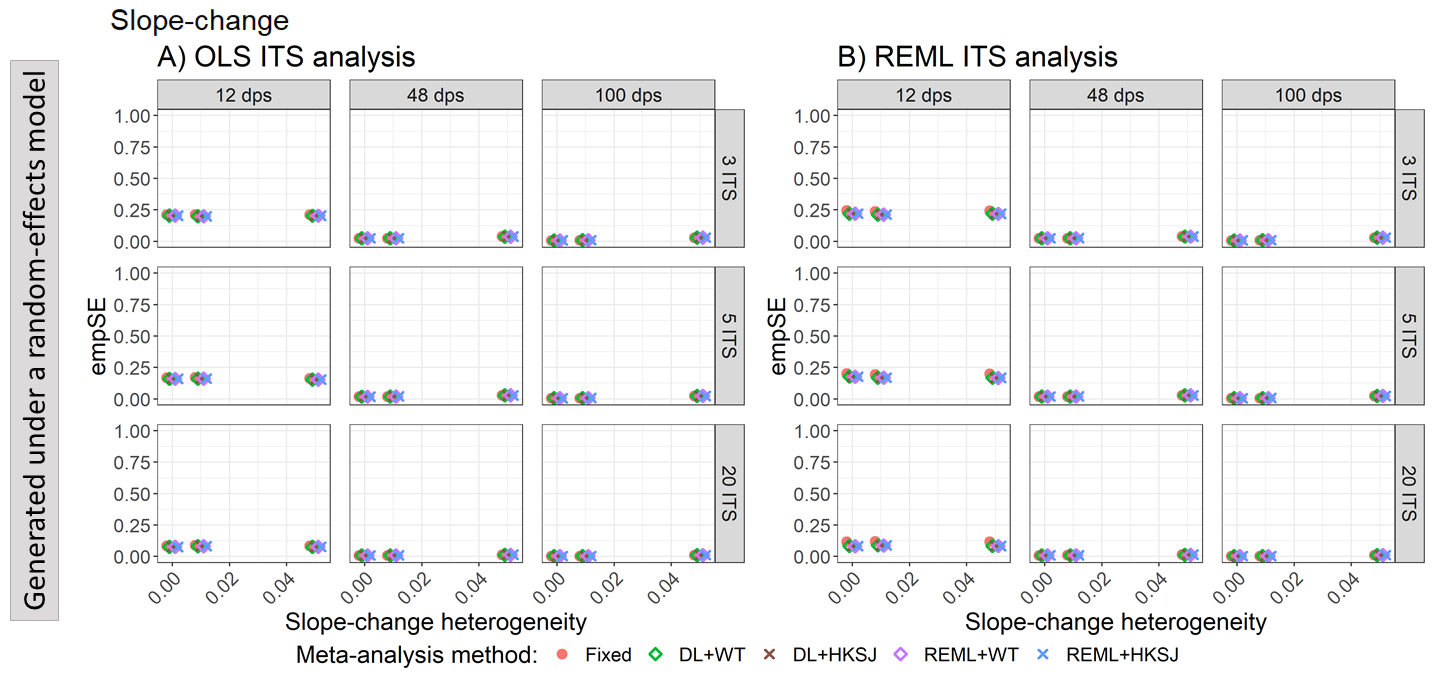

Appendix Figure S86. Plots of empirical standard error (empSE) of the immediate slope-change (y-axis) when the ITS are analysed with OLS (A) and REML (B) using fixed-effect (red circles), DL+WT (green diamonds) and DL+HKSJ (brown crosses), REML+WT (purple diamonds) and REML+HKSJ (blue crosses) meta-analysis methods versus slope-change heterogeneity (x-axis). Plots are presented separately by combinations of the number of included studies (rows) and the number of datapoints (columns). Simulation scenarios include a **level-change of 0**, level-change heterogeneity of 0, **slope-change of 0.1**, and fixed levels of autocorrelation.

DL, DerSimonian and Laird. dps, datapoints. HKSJ, Hartung-Knapp / Sidik-Jonkman. ITS, interrupted time series. OLS, ordinary least squares. REML, restricted maximum likelihood. WT, Wald-type.

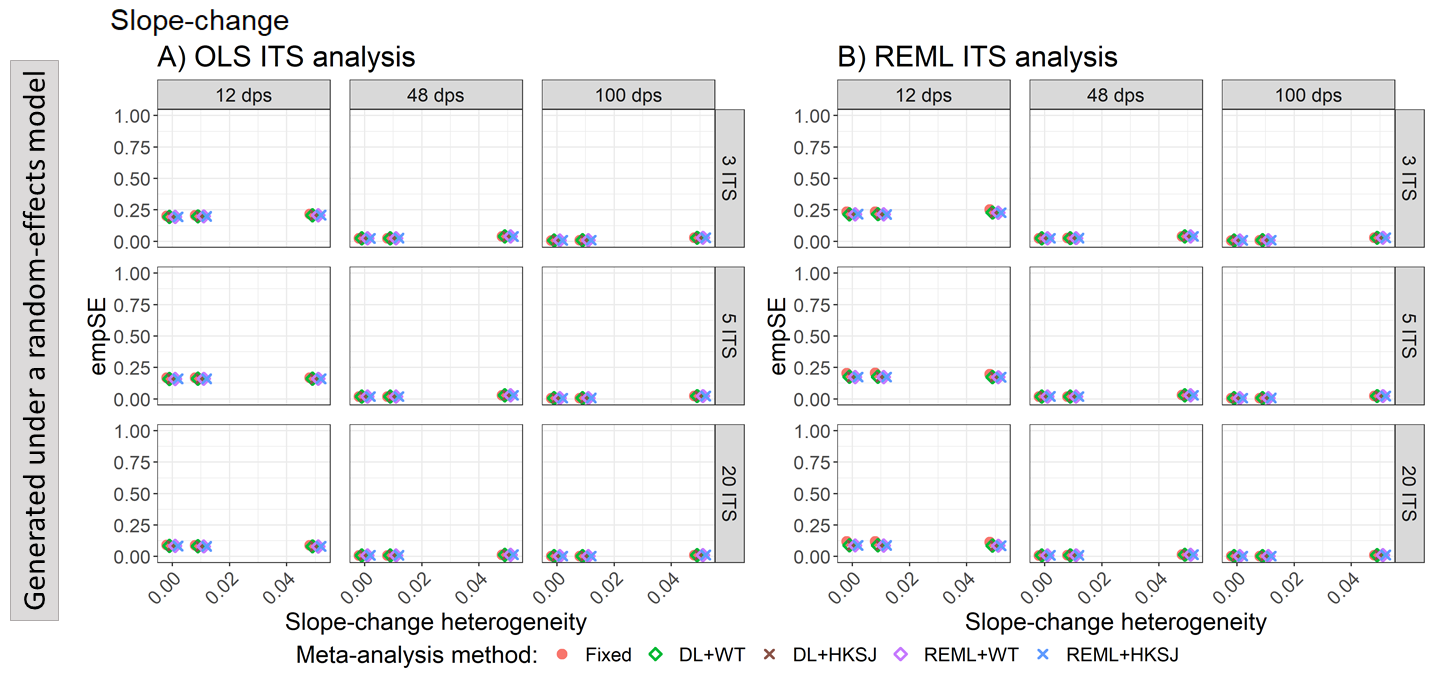

Appendix Figure S87. Plots of empirical standard error (empSE) of the immediate slope-change (y-axis) when the ITS are analysed with OLS (A) and REML (B) using fixed-effect (red circles), DL+WT (green diamonds) and DL+HKSJ (brown crosses), REML+WT (purple diamonds) and REML+HKSJ (blue crosses) meta-analysis methods versus slope-change heterogeneity (x-axis). Plots are presented separately by combinations of the number of included studies (rows) and the number of datapoints (columns). Simulation scenarios include a **level-change of 1**, level-change heterogeneity of 0, **slope-change of 0**, and fixed levels of autocorrelation.

DL, DerSimonian and Laird. dps, datapoints. HKSJ, Hartung-Knapp / Sidik-Jonkman. ITS, interrupted time series. OLS, ordinary least squares. REML, restricted maximum likelihood. WT, Wald-type.

#### 4.4 Ratio of model based standard errors to empirical standard errors

##### 4.4.1 Estimation of level-change – Ratio of model based standard errors to empirical standard errors vs autocorrelation for scenarios with different level and slope-change combinations

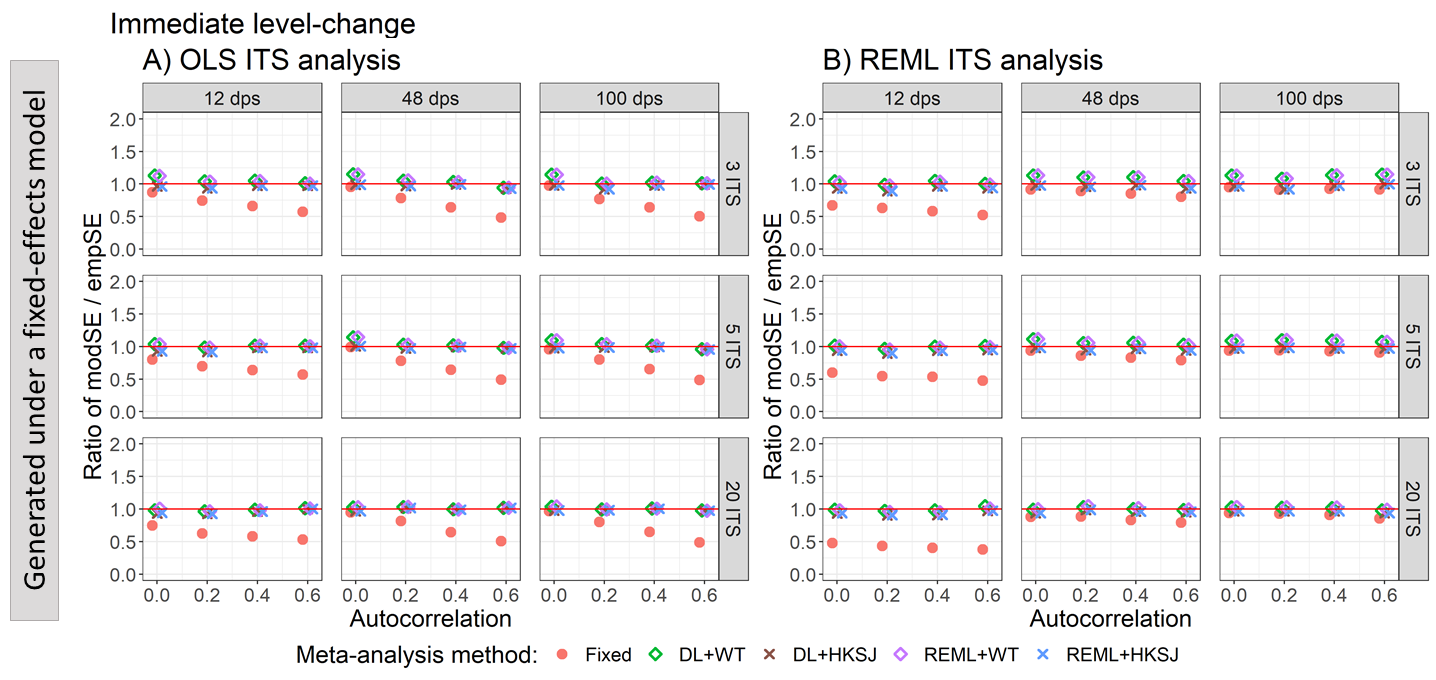

Appendix Figure S88. Plots of the ratio of model-based standard error (modSE) to the empirical standard error (empSE) of level-change (y-axis) when the data was generated under a fixed-effect model and the ITS studies were analysed with OLS (A) and REML (B) using fixed-effect (red circles), DL+WT (green diamonds) and REML+HKSJ (blue crosses) meta-analysis methods versus autocorrelation (x-axis). Plots are presented separately by the number of included studies (rows) and number of datapoints (columns). The solid red line depicts a ratio of one, where the model-based standard error and empirical standard error are equal and thus that the model-based standard error accurately estimates the true standard error. Simulation scenarios include a **level-change of 0**, level-change heterogeneity of 0, **slope-change of 0**, slope-change heterogeneity of 0, and fixed autocorrelation.

DL, DerSimonian and Laird. dps, datapoints. HKSJ, Hartung-Knapp / Sidik-Jonkman. ITS, interrupted time series. OLS, ordinary least squares. REML, restricted maximum likelihood. WT, Wald-type.

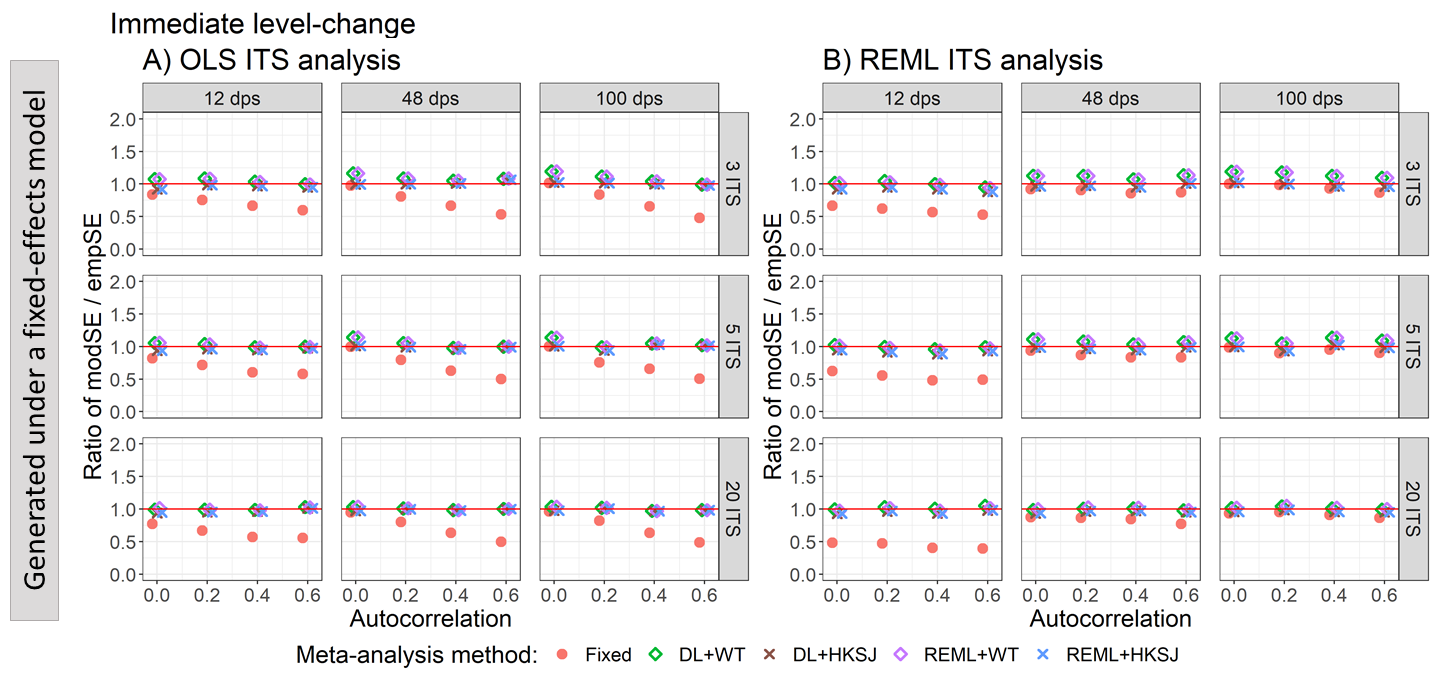

Appendix Figure S89. Plots of the ratio of model-based standard error (modSE) to the empirical standard error (empSE) of level-change (y-axis) when the data was generated under a fixed-effect model and the ITS studies were analysed with OLS (A) and REML (B) using fixed-effect (red circles), DL+WT (green diamonds) and REML+HKSJ (blue crosses) meta-analysis methods versus autocorrelation (x-axis). Plots are presented separately by the number of included studies (rows) and number of datapoints (columns). The solid red line depicts a ratio of one, where the model-based standard error and empirical standard error are equal and thus that the model-based standard error accurately estimates the true standard error. Simulation scenarios include a **level-change of 0**, level-change heterogeneity of 0, **slope-change of 0.1**, slope-change heterogeneity of 0, and fixed autocorrelation.

DL, DerSimonian and Laird. dps, datapoints. HKSJ, Hartung-Knapp / Sidik-Jonkman. ITS, interrupted time series. OLS, ordinary least squares. REML, restricted maximum likelihood. WT, Wald-type.

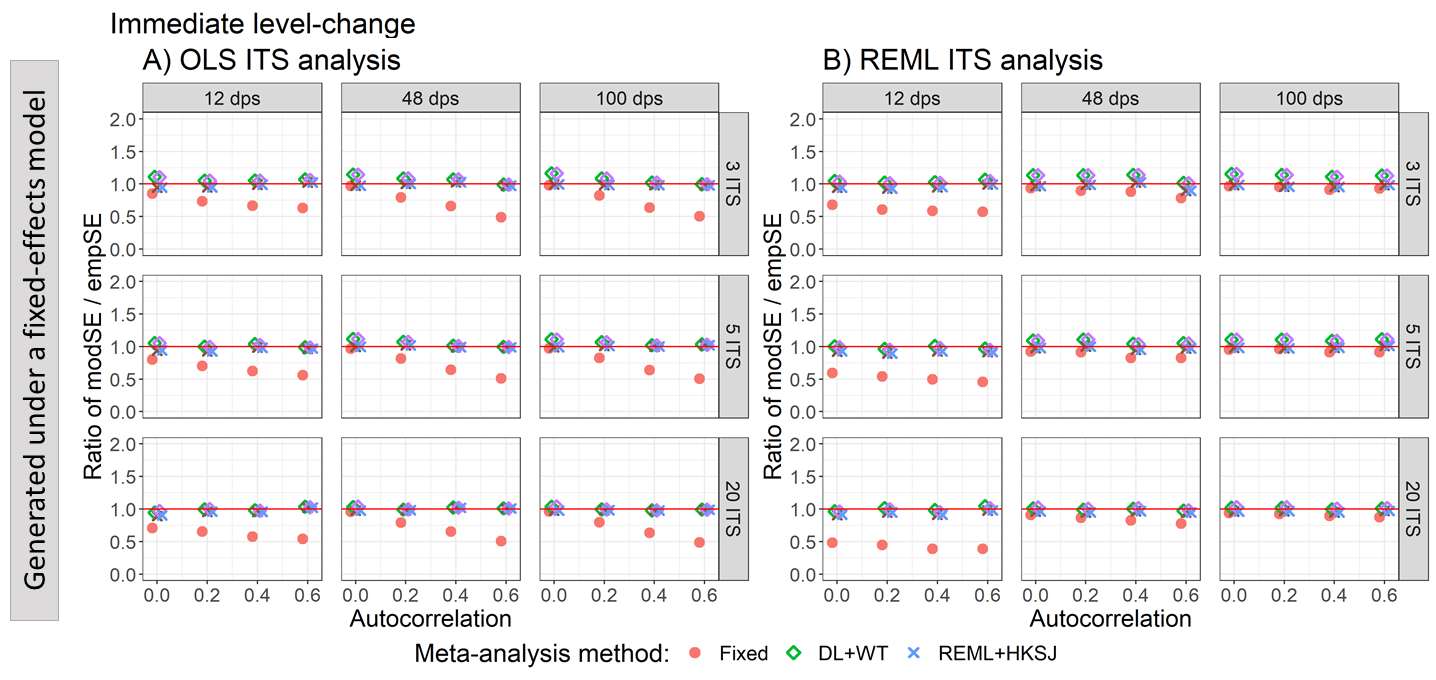

Appendix Figure S90. Plots of the ratio of model-based standard error (modSE) to the empirical standard error (empSE) of level-change (y-axis) when the data was generated under a fixed-effect model and the ITS studies were analysed with OLS (A) and REML (B) using fixed-effect (red circles), DL+WT (green diamonds) and REML+HKSJ (blue crosses) meta-analysis methods versus autocorrelation (x-axis). Plots are presented separately by the number of included studies (rows) and number of datapoints (columns). The solid red line depicts a ratio of one, where the model-based standard error and empirical standard error are equal and thus that the model-based standard error accurately estimates the true standard error. Simulation scenarios include a **level-change of 1**, level-change heterogeneity of 0, **slope-change of 0**, slope-change heterogeneity of 0, and fixed autocorrelation.

DL, DerSimonian and Laird. dps, datapoints. HKSJ, Hartung-Knapp / Sidik-Jonkman. ITS, interrupted time series. OLS, ordinary least squares. REML, restricted maximum likelihood. WT, Wald-type.

##### 4.4.2 Estimation of level-change – Ratio of model based standard errors to empirical standard errors vs level-change heterogeneity for scenarios with different level and slope-change combinations

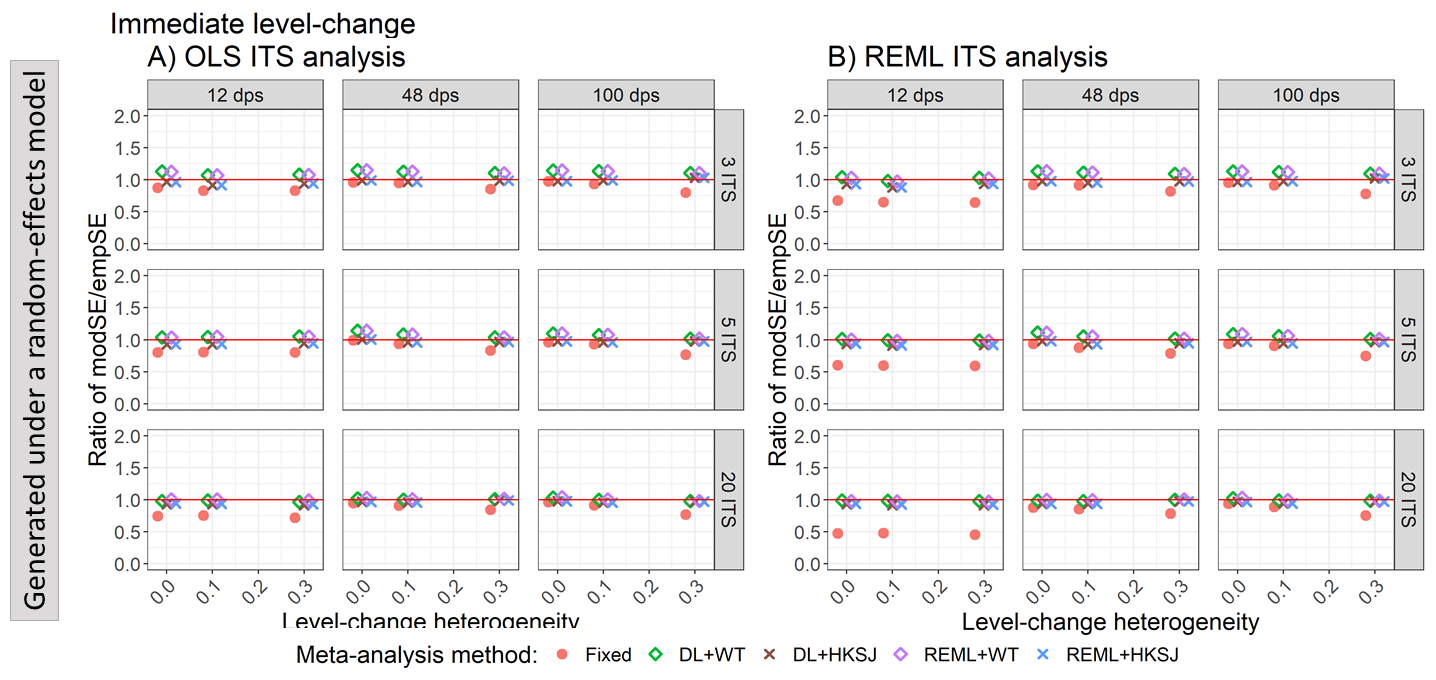

Appendix Figure S91. Plots of the ratio of model based standard error (modSE) to the empirical standard error (empSE) of level-change (y-axis) when the ITS studies were analysed with OLS (A) and REML (B) using fixed-effect (red circles), DL+WT (green diamonds) and REML+HKSJ (blue crosses) meta-analysis methods versus level-change heterogeneity (x-axis). Plots are presented separately by combinations of the number of included studies (rows) and number of datapoints (columns). The solid red line depicts a ratio of one, where the model-based standard error and empirical standard error are equal and thus that the model-based standard error accurately estimates the true standard error. Simulation scenarios include a **level-change of 0, slope-change of 0,** level-change heterogeneity of 0, and autocorrelation of 0.

DL, DerSimonian and Laird. dps, datapoints. HKSJ, Hartung-Knapp / Sidik-Jonkman. ITS, interrupted time series. OLS, ordinary least squares. REML, restricted maximum likelihood. WT, Wald-type.

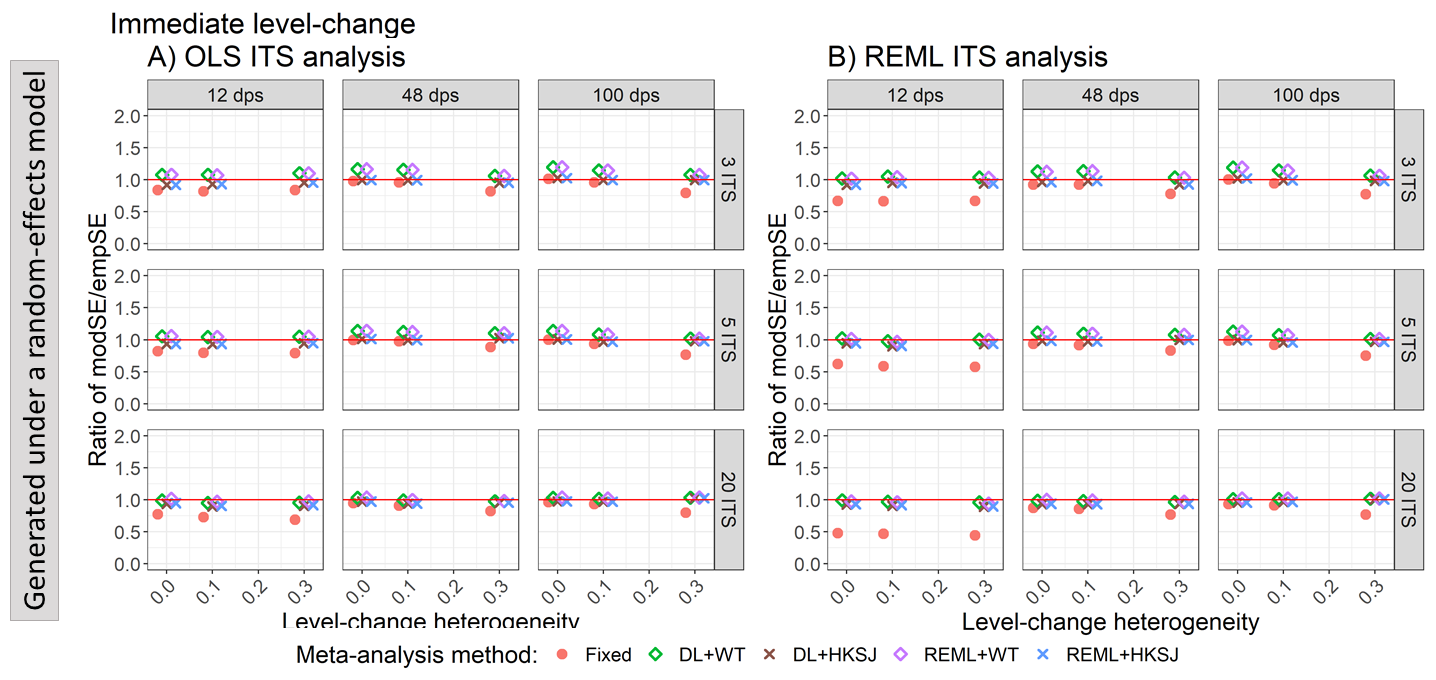

Appendix Figure S92. Plots of the ratio of model based standard error (modSE) to the empirical standard error (empSE) of level-change (y-axis) when the ITS studies were analysed with OLS (A) and REML (B) using fixed-effect (red circles), DL+WT (green diamonds) and REML+HKSJ (blue crosses) meta-analysis methods versus level-change heterogeneity (x-axis). Plots are presented separately by combinations of the number of included studies (rows) and number of datapoints (columns). The solid red line depicts a ratio of one, where the model-based standard error and empirical standard error are equal and thus that the model-based standard error accurately estimates the true standard error. Simulation scenarios include a **level-change of 0, slope-change of 0.1,** level-change heterogeneity of 0, and autocorrelation of 0.

DL, DerSimonian and Laird. dps, datapoints. HKSJ, Hartung-Knapp / Sidik-Jonkman. ITS, interrupted time series. OLS, ordinary least squares. REML, restricted maximum likelihood. WT, Wald-type.

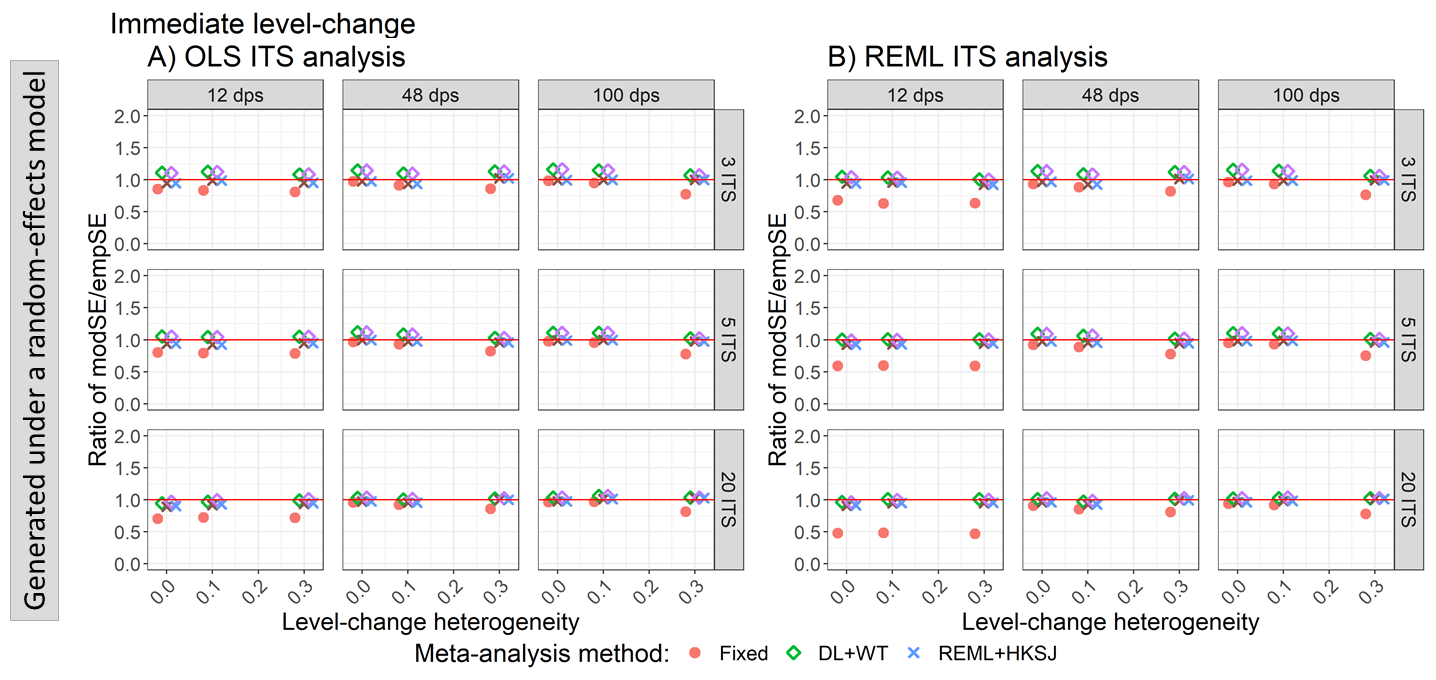

Appendix Figure S93. Plots of the ratio of model based standard error (modSE) to the empirical standard error (empSE) of level-change (y-axis) when the ITS studies were analysed with OLS (A) and REML (B) using fixed-effect (red circles), DL+WT (green diamonds) and REML+HKSJ (blue crosses) meta-analysis methods versus level-change heterogeneity (x-axis). Plots are presented separately by combinations of the number of included studies (rows) and number of datapoints (columns). The solid red line depicts a ratio of one, where the model-based standard error and empirical standard error are equal and thus that the model-based standard error accurately estimates the true standard error. Simulation scenarios include a **level-change of 1, slope-change of 0,** level-change heterogeneity of 0, and autocorrelation of 0.

DL, DerSimonian and Laird. dps, datapoints. HKSJ, Hartung-Knapp / Sidik-Jonkman. ITS, interrupted time series. OLS, ordinary least squares. REML, restricted maximum likelihood. WT, Wald-type.

##### 4.4.3 Estimation of slope -change – Ratio of model based standard errors to empirical standard errors vs autocorrelation for scenarios with different level and slope-change combinations

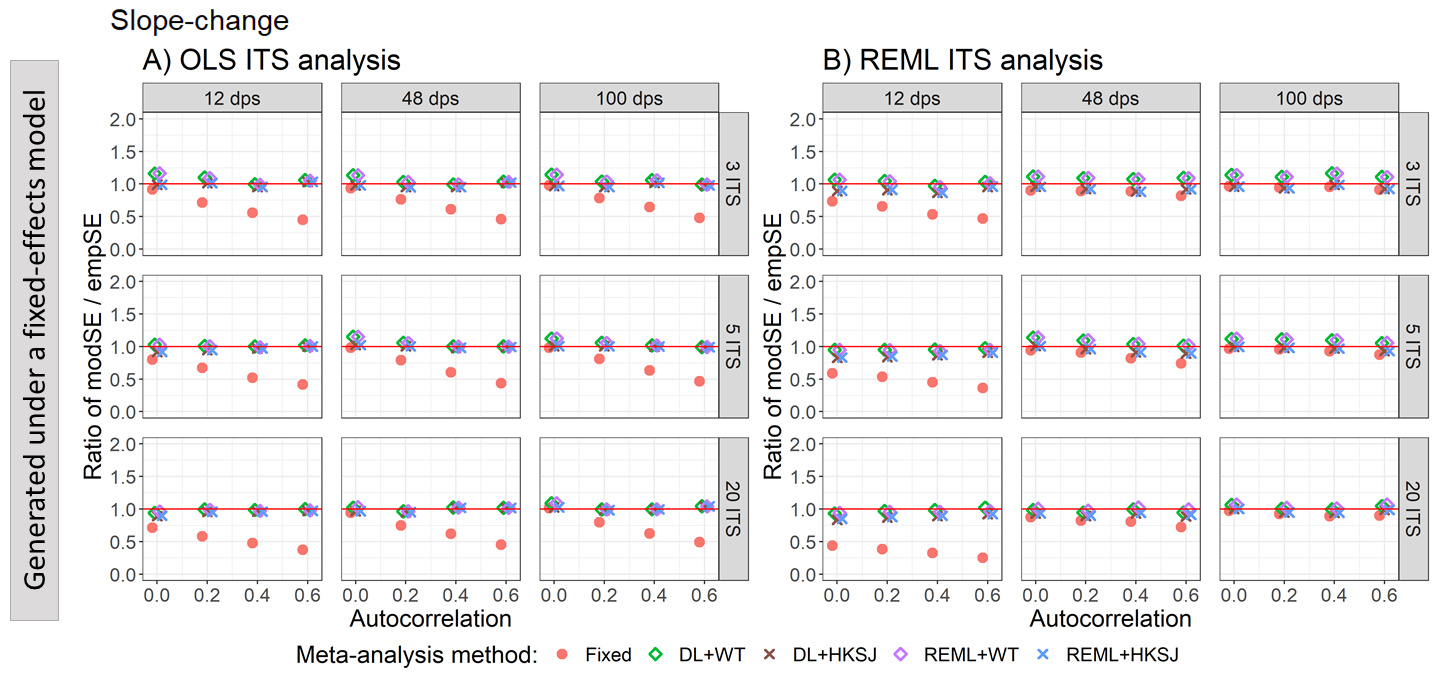

Appendix Figure S94. Plots of the ratio of model-based standard error (modSE) to the empirical standard error (empSE) of slope-change (y-axis) when the data was generated under a fixed-effect model and the ITS studies were analysed with OLS (A) and REML (B) using fixed-effect (red circles), DL+WT (green diamonds) and REML+HKSJ (blue crosses) meta-analysis methods versus autocorrelation (x-axis). Plots are presented separately by the number of included studies (rows) and number of datapoints (columns). The solid red line depicts a ratio of one, where the model-based standard error and empirical standard error are equal and thus that the model-based standard error accurately estimates the true standard error. Simulation scenarios include a **level-change of 0**, level-change heterogeneity of 0, **slope-change of 0**, slope-change heterogeneity of 0, and fixed autocorrelation.

DL, DerSimonian and Laird. dps, datapoints. HKSJ, Hartung-Knapp / Sidik-Jonkman. ITS, interrupted time series. OLS, ordinary least squares. REML, restricted maximum likelihood. WT, Wald-type.

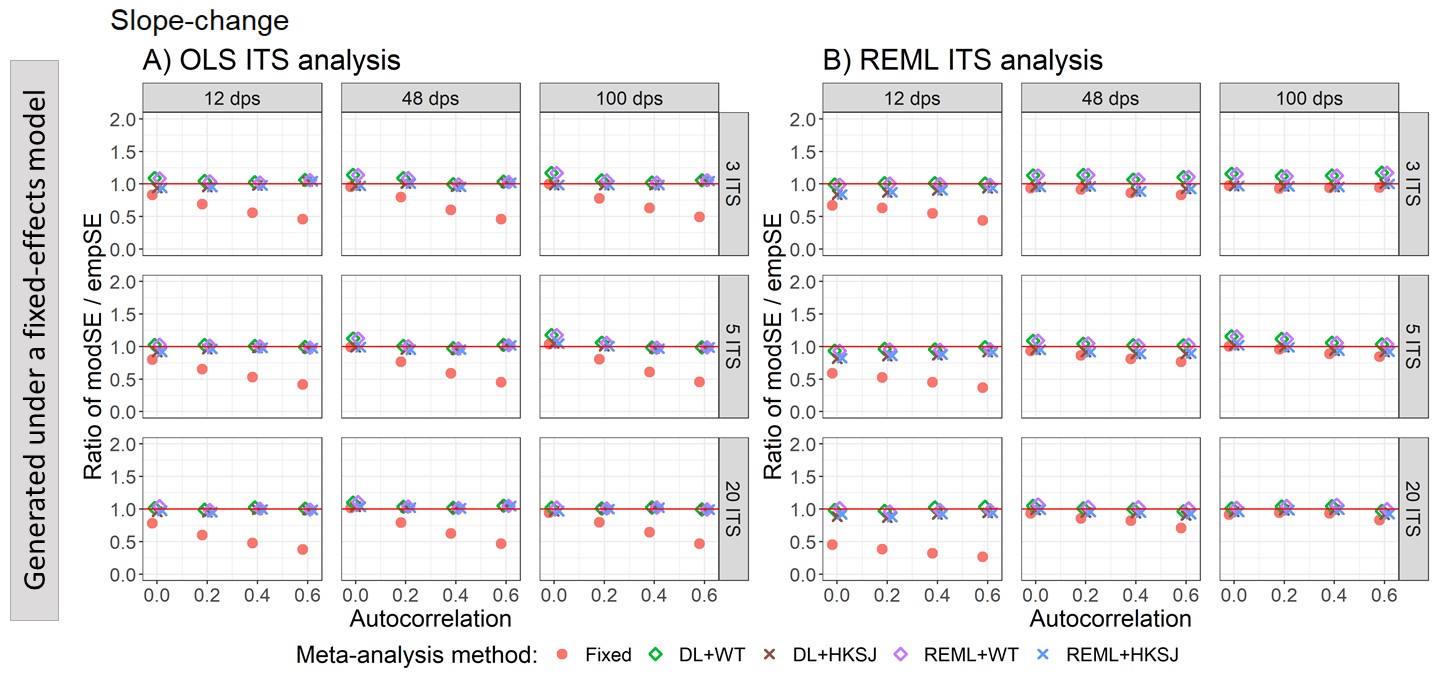

Appendix Figure S95. Plots of the ratio of model-based standard error (modSE) to the empirical standard error (empSE) of slope-change (y-axis) when the data was generated under a fixed-effect model and the ITS studies were analysed with OLS (A) and REML (B) using fixed-effect (red circles), DL+WT (green diamonds) and REML+HKSJ (blue crosses) meta-analysis methods versus autocorrelation (x-axis). Plots are presented separately by the number of included studies (rows) and number of datapoints (columns). The solid red line depicts a ratio of one, where the model-based standard error and empirical standard error are equal and thus that the model-based standard error accurately estimates the true standard error. Simulation scenarios include a **level-change of 0**, level-change heterogeneity of 0, **slope-change of 0.1**, slope-change heterogeneity of 0, and fixed autocorrelation.

DL, DerSimonian and Laird. dps, datapoints. HKSJ, Hartung-Knapp / Sidik-Jonkman. ITS, interrupted time series. OLS, ordinary least squares. REML, restricted maximum likelihood. WT, Wald-type.

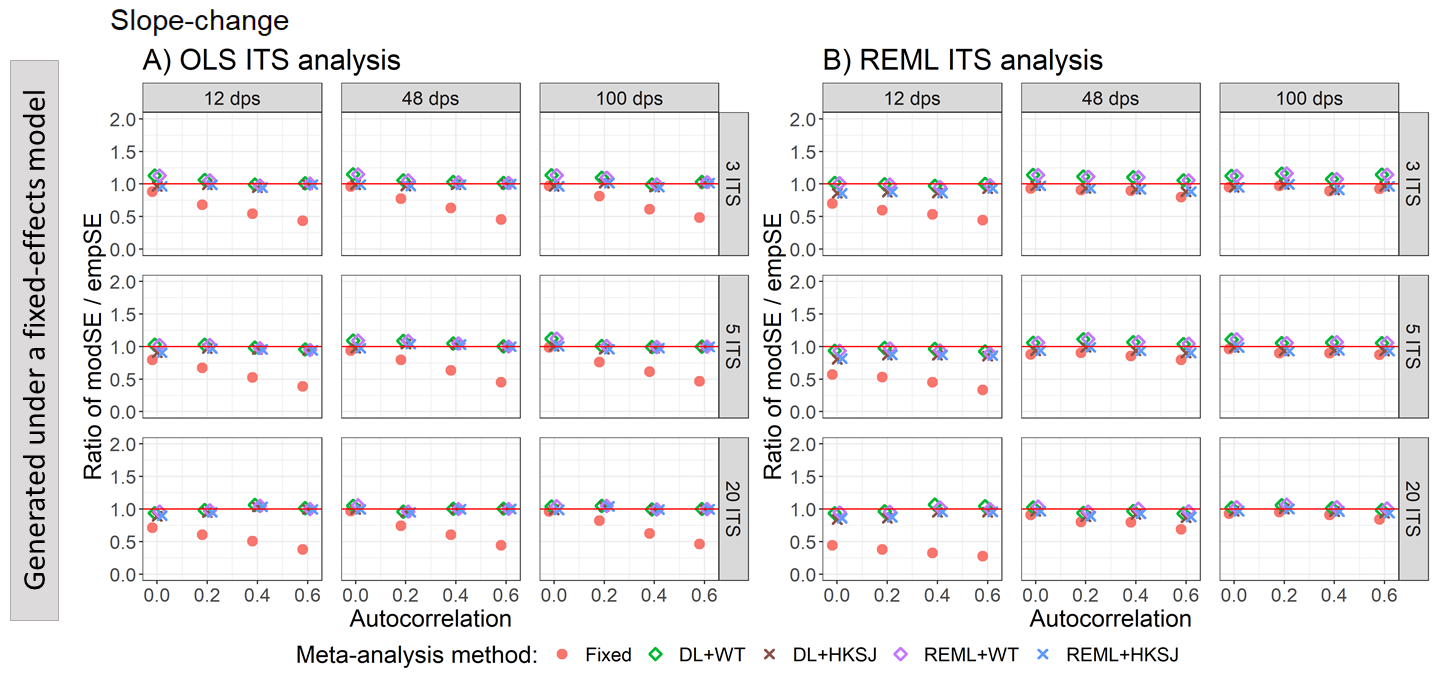

Appendix Figure S96. Plots of the ratio of model-based standard error (modSE) to the empirical standard error (empSE) of slope-change (y-axis) when the data was generated under a fixed-effect model and the ITS studies were analysed with OLS (A) and REML (B) using fixed-effect (red circles), DL+WT (green diamonds) and REML+HKSJ (blue crosses) meta-analysis methods versus autocorrelation (x-axis). Plots are presented separately by the number of included studies (rows) and number of datapoints (columns). The solid red line depicts a ratio of one, where the model-based standard error and empirical standard error are equal and thus that the model-based standard error accurately estimates the true standard error. Simulation scenarios include a **level-change of 1**, level-change heterogeneity of 0, **slope-change of 0**, slope-change heterogeneity of 0, and fixed autocorrelation.

DL, DerSimonian and Laird. dps, datapoints. HKSJ, Hartung-Knapp / Sidik-Jonkman. ITS, interrupted time series. OLS, ordinary least squares. REML, restricted maximum likelihood. WT, Wald-type.

##### 4.4.4 Estimation of slope -change – Ratio of model based standard errors to empirical standard errors vs slope-change heterogeneity for scenarios with different level and slope-change combinations

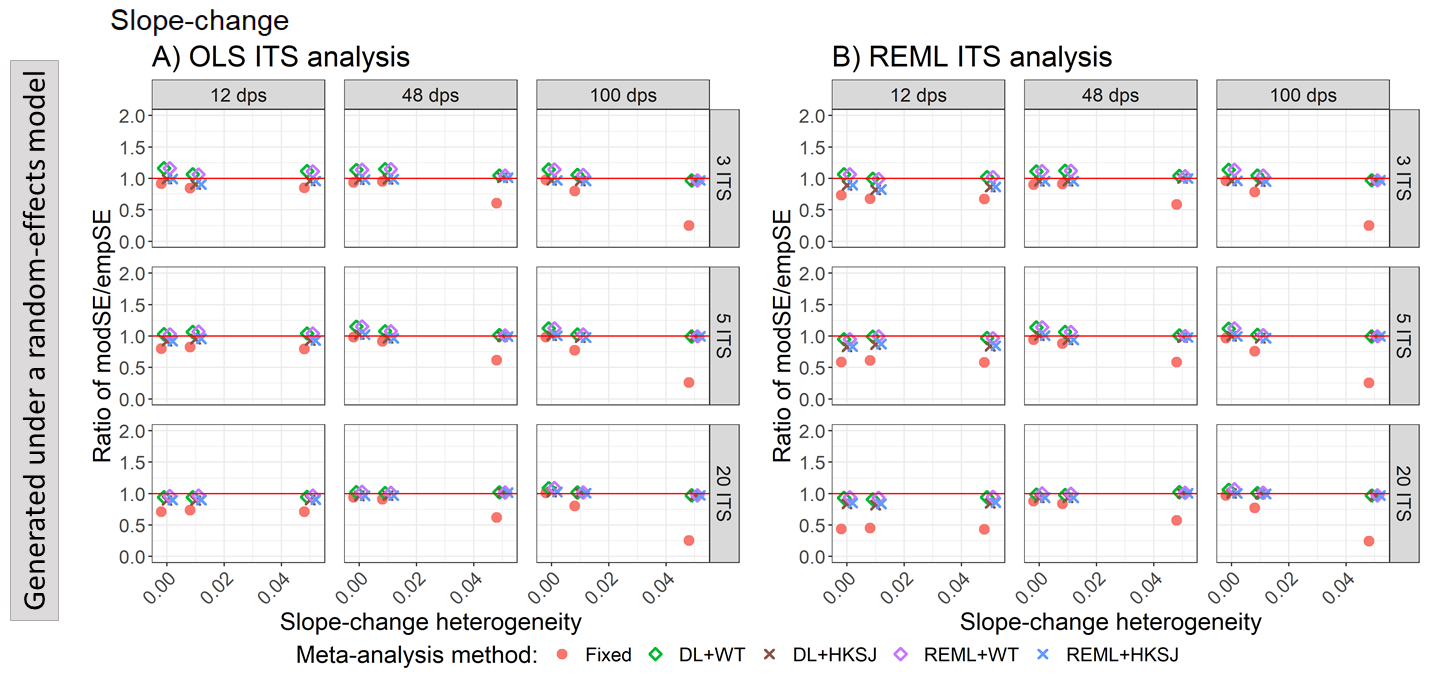

Appendix Figure S97. Plots of the ratio of model based standard error (modSE) to the empirical standard error (empSE) of slope-change (y-axis) when the ITS studies were analysed with OLS (A) and REML (B) using fixed-effect (red circles), DL+WT (green diamonds) and REML+HKSJ (blue crosses) meta-analysis methods versus slope-change heterogeneity (x-axis). Plots are presented separately by combinations of the number of included studies (rows) and number of datapoints (columns). The solid red line depicts a ratio of one, where the model-based standard error and empirical standard error are equal and thus that the model-based standard error accurately estimates the true standard error. Simulation scenarios include a **level-change of 0,** **slope-change of 0**, slope-change heterogeneity of 0, and autocorrelation of 0.

DL, DerSimonian and Laird. dps, datapoints. HKSJ, Hartung-Knapp / Sidik-Jonkman. ITS, interrupted time series. OLS, ordinary least squares. REML, restricted maximum likelihood. WT, Wald-type.

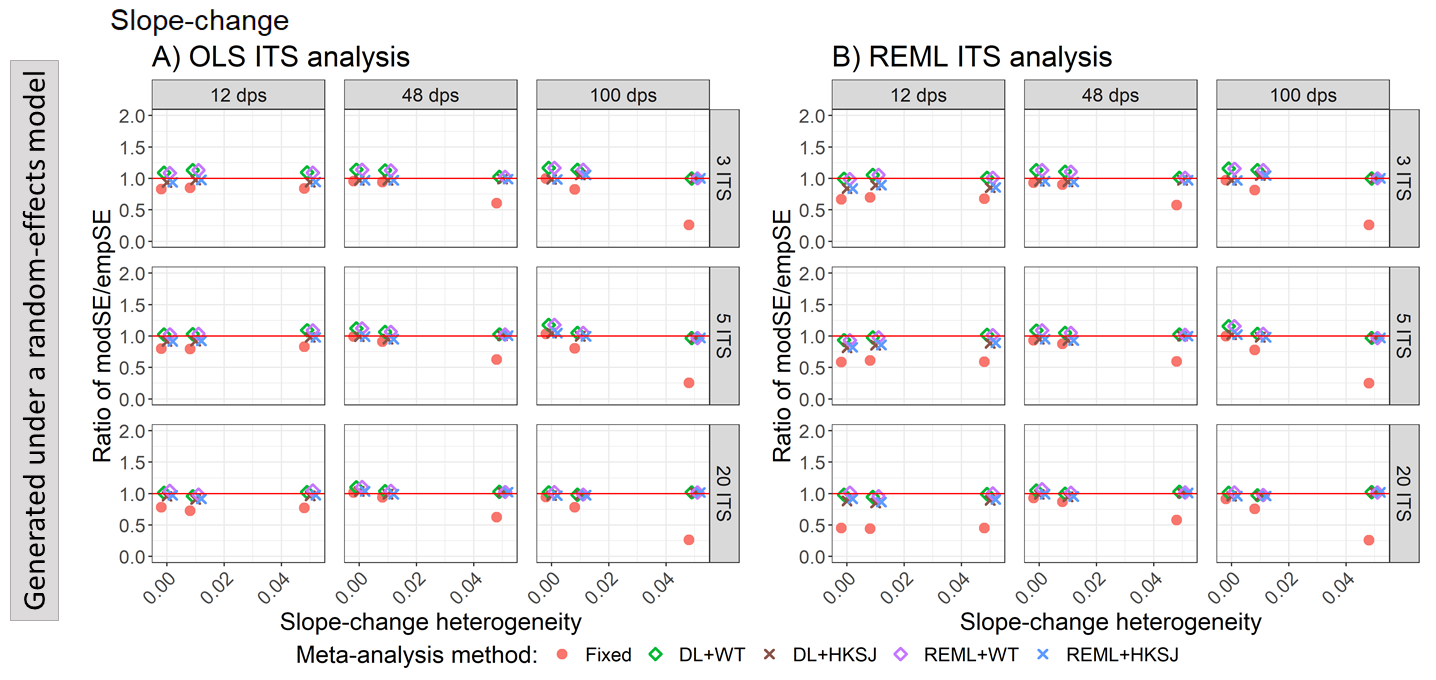

Appendix Figure S98. Plots of the ratio of model based standard error (modSE) to the empirical standard error (empSE) of slope-change (y-axis) when the ITS studies were analysed with OLS (A) and REML (B) using fixed-effect (red circles), DL+WT (green diamonds) and REML+HKSJ (blue crosses) meta-analysis methods versus slope-change heterogeneity (x-axis). Plots are presented separately by combinations of the number of included studies (rows) and number of datapoints (columns). The solid red line depicts a ratio of one, where the model-based standard error and empirical standard error are equal and thus that the model-based standard error accurately estimates the true standard error. Simulation scenarios include a **level-change of 0,** **slope-change of 0.1**, slope-change heterogeneity of 0, and autocorrelation of 0.

DL, DerSimonian and Laird. dps, datapoints. HKSJ, Hartung-Knapp / Sidik-Jonkman. ITS, interrupted time series. OLS, ordinary least squares. REML, restricted maximum likelihood. WT, Wald-type.

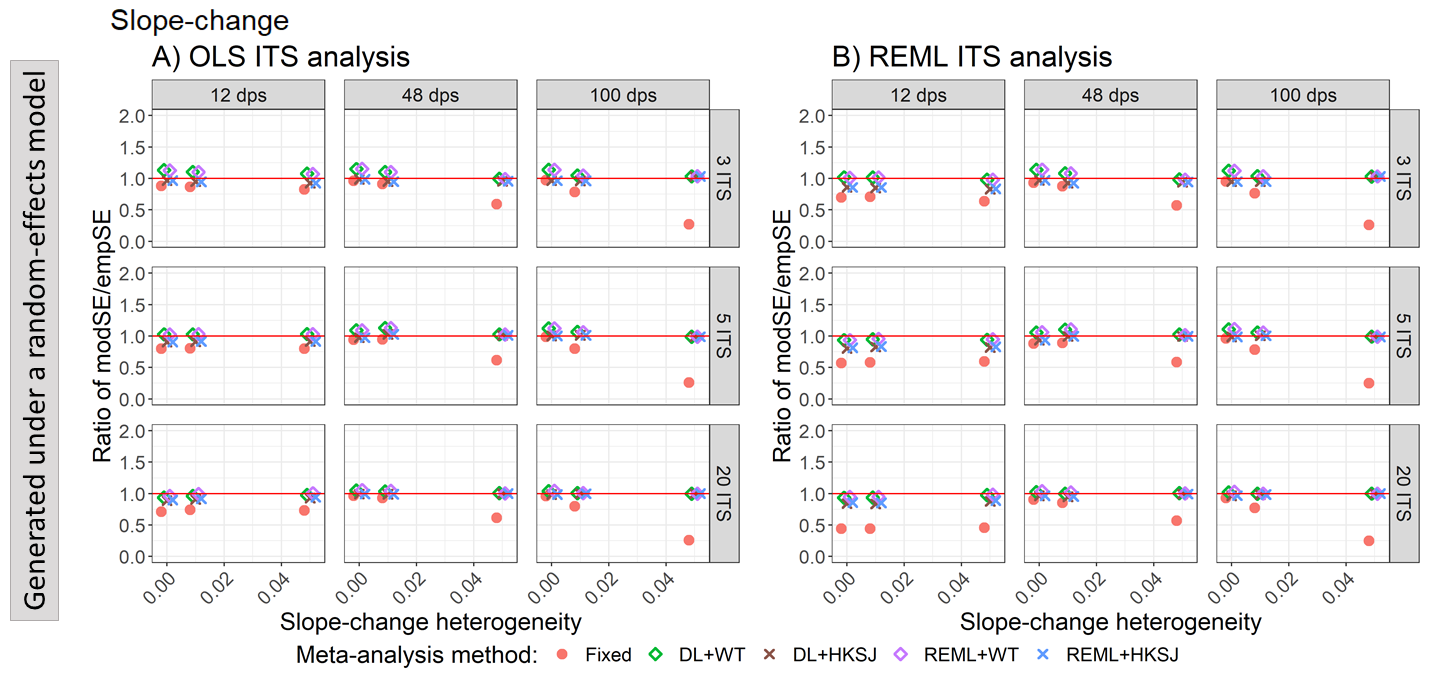

Appendix Figure S99. Plots of the ratio of model based standard error (modSE) to the empirical standard error (empSE) of slope-change (y-axis) when the ITS studies were analysed with OLS (A) and REML (B) using fixed-effect (red circles), DL+WT (green diamonds) and REML+HKSJ (blue crosses) meta-analysis methods versus slope-change heterogeneity (x-axis). Plots are presented separately by combinations of the number of included studies (rows) and number of datapoints (columns). The solid red line depicts a ratio of one, where the model-based standard error and empirical standard error are equal and thus that the model-based standard error accurately estimates the true standard error. Simulation scenarios include a **level-change of 1,** **slope-change of 0**, slope-change heterogeneity of 0, and autocorrelation of 0.

DL, DerSimonian and Laird. dps, datapoints. HKSJ, Hartung-Knapp / Sidik-Jonkman. ITS, interrupted time series. OLS, ordinary least squares. REML, restricted maximum likelihood. WT, Wald-type.

#### 4.5 Statistical power

##### 4.5.1 Estimation of level-change – statistical power vs autocorrelation for scenarios with different level and slope-change combinations

Appendix Figure S100. Plots of statistical power (the percentage of simulations that have a 95% confidence interval that did not include zero – only scenarios with a confidence interval coverage of greater than 90% are plotted) of the meta-analytic level-change (y-axis) when the data was generated under a fixed-effect model and the ITS studies were analysed with OLS (A) and REML (B) using the fixed effect (red circles), DL+WT (green diamonds) and DL+HKSJ (brown crosses), REML+WT (purple diamonds) and REML+HKSJ (blue crosses) meta-analysis methods versus autocorrelation (x-axis). Plots are presented separately by combinations of the number of included studies (rows) and the number of datapoints (columns). Simulation scenarios include a **level-change of 0**, level-change heterogeneity of 0, **slope-change of 0**, slope-change heterogeneity of 0, and fixed levels of autocorrelation.

DL, DerSimonian and Laird. dps, datapoints. HKSJ, Hartung-Knapp / Sidik-Jonkman. ITS, interrupted time series. OLS, ordinary least squares. REML, restricted maximum likelihood. WT, Wald-type.

Appendix Figure S101. Plots of statistical power (the percentage of simulations that have a 95% confidence interval that did not include zero – only scenarios with a confidence interval coverage of greater than 90% are plotted) of the meta-analytic level-change (y-axis) when the data was generated under a fixed-effect model and the ITS studies were analysed with OLS (A) and REML (B) using the fixed effect (red circles), DL+WT (green diamonds) and DL+HKSJ (brown crosses), REML+WT (purple diamonds) and REML+HKSJ (blue crosses) meta-analysis methods versus autocorrelation (x-axis). Plots are presented separately by combinations of the number of included studies (rows) and the number of datapoints (columns). Simulation scenarios include a **level-change of 0**, level-change heterogeneity of 0, **slope-change of 0.1**, slope-change heterogeneity of 0, and fixed levels of autocorrelation.

DL, DerSimonian and Laird. dps, datapoints. HKSJ, Hartung-Knapp / Sidik-Jonkman. ITS, interrupted time series. OLS, ordinary least squares. REML, restricted maximum likelihood. WT, Wald-type.

Appendix Figure S102. Plots of statistical power (the percentage of simulations that have a 95% confidence interval that did not include zero – only scenarios with a confidence interval coverage of greater than 90% are plotted) of the meta-analytic level-change (y-axis) when the data was generated under a fixed-effect model and the ITS studies were analysed with OLS (A) and REML (B) using the fixed effect (red circles), DL+WT (green diamonds) and DL+HKSJ (brown crosses), REML+WT (purple diamonds) and REML+HKSJ (blue crosses) meta-analysis methods versus autocorrelation (x-axis). Plots are presented separately by combinations of the number of included studies (rows) and the number of datapoints (columns). Simulation scenarios include a **level-change of 1**, level-change heterogeneity of 0, **slope-change of 0**, slope-change heterogeneity of 0, and fixed levels of autocorrelation.

DL, DerSimonian and Laird. dps, datapoints. HKSJ, Hartung-Knapp / Sidik-Jonkman. ITS, interrupted time series. OLS, ordinary least squares. REML, restricted maximum likelihood. WT, Wald-type.

##### 4.5.2 Estimation of level-change – statistical power vs level-change heterogeneity for scenarios with different level and slope-change combinations

Appendix Figure S103. Plots of statistical power (the percentage of simulations that have a 95% confidence interval that did not include zero – only scenarios with a confidence interval coverage of greater than 90% are plotted) of the meta-analytic immediate level-change (y-axis) when the ITS studies were analysed with OLS (A) and REML (B) using the fixed effect (red circles), DL+WT (green diamonds) and REML+HKSJ (blue crosses) meta-analysis methods versus level-change heterogeneity (x-axis), by number of datapoints (horizontal facets) and number of included studies (vertical facets). Simulation settings include a **level-change of 0, slope-change of 0**, slope-change heterogeneity of 0, and fixed levels of autocorrelation.

DL, DerSimonian and Laird. dps, datapoints. HKSJ, Hartung-Knapp / Sidik-Jonkman. ITS, interrupted time series. OLS, ordinary least squares. REML, restricted maximum likelihood. WT, Wald-type.

Appendix Figure S104. Plots of statistical power (the percentage of simulations that have a 95% confidence interval that did not include zero – only scenarios with a confidence interval coverage of greater than 90% are plotted) of the meta-analytic immediate level-change (y-axis) when the ITS studies were analysed with OLS (A) and REML (B) using the fixed effect (red circles), DL+WT (green diamonds) and REML+HKSJ (blue crosses) meta-analysis methods versus level-change heterogeneity (x-axis), by number of datapoints (horizontal facets) and number of included studies (vertical facets). Simulation settings include a **level-change of 0, slope-change of 0.1**, slope-change heterogeneity of 0, and fixed levels of autocorrelation.

DL, DerSimonian and Laird. dps, datapoints. HKSJ, Hartung-Knapp / Sidik-Jonkman. ITS, interrupted time series. OLS, ordinary least squares. REML, restricted maximum likelihood. WT, Wald-type.

Appendix Figure S105. Plots of statistical power (the percentage of simulations that have a 95% confidence interval that did not include zero – only scenarios with a confidence interval coverage of greater than 90% are plotted) of the meta-analytic immediate level-change (y-axis) when the ITS studies were analysed with OLS (A) and REML (B) using the fixed effect (red circles), DL+WT (green diamonds) and REML+HKSJ (blue crosses) meta-analysis methods versus level-change heterogeneity (x-axis), by number of datapoints (horizontal facets) and number of included studies (vertical facets). Simulation settings include a **level-change of 1, slope-change of 0**, slope-change heterogeneity of 0, and fixed levels of autocorrelation.

DL, DerSimonian and Laird. dps, datapoints. HKSJ, Hartung-Knapp / Sidik-Jonkman. ITS, interrupted time series. OLS, ordinary least squares. REML, restricted maximum likelihood. WT, Wald-type.

##### 4.5.3 Estimation of slope-change – statistical power vs autocorrelation for scenarios with different level and slope-change combinations

DL, DerSimonian and Laird. dps, datapoints. HKSJ, Hartung-Knapp / Sidik-Jonkman. ITS, interrupted time series. OLS, ordinary least squares. REML, restricted maximum likelihood. WT, Wald-type.

##### 4.5.4 Estimation of slope -change – statistical power vs slope-change heterogeneity for scenarios with different level and slope-change combinations

Appendix Figure S109. Plots of statistical power (the percentage of simulations that have a 95% confidence interval that did not include zero – only scenarios with a confidence interval coverage of greater than 90% are plotted) of the meta-analytic slope-change (y-axis) when the ITS are analysed with OLS (A) and REML (B) for the fixed effect (red circles), DL+WT (green diamonds) and DL+HKSJ (brown crosses), REML+WT (purple diamonds) and REML+HKSJ (blue crosses) meta-analysis methods versus slope-change heterogeneity (x-axis), by number of datapoints (horizontal facets) and number of included studies (vertical facets). Simulation settings include a **level-change of 0**, level-change heterogeneity of 0, **slope-change of 0**, and autocorrelation of 0.

DL, DerSimonian and Laird. dps, datapoints. HKSJ, Hartung-Knapp / Sidik-Jonkman. ITS, interrupted time series. OLS, ordinary least squares. REML, restricted maximum likelihood. WT, Wald-type.

Appendix Figure S110. Plots of statistical power (the percentage of simulations that have a 95% confidence interval that did not include zero – only scenarios with a confidence interval coverage of greater than 90% are plotted) of the meta-analytic slope-change (y-axis) when the ITS are analysed with OLS (A) and REML (B) for the fixed effect (red circles), DL+WT (green diamonds) and DL+HKSJ (brown crosses), REML+WT (purple diamonds) and REML+HKSJ (blue crosses) meta-analysis methods versus slope-change heterogeneity (x-axis), by number of datapoints (horizontal facets) and number of included studies (vertical facets). Simulation settings include a **level-change of 0**, level-change heterogeneity of 0, **slope-change of 0.1**, and autocorrelation of 0.

DL, DerSimonian and Laird. dps, datapoints. HKSJ, Hartung-Knapp / Sidik-Jonkman. ITS, interrupted time series. OLS, ordinary least squares. REML, restricted maximum likelihood. WT, Wald-type.

Appendix Figure S111. Plots of statistical power (the percentage of simulations that have a 95% confidence interval that did not include zero – only scenarios with a confidence interval coverage of greater than 90% are plotted) of the meta-analytic slope-change (y-axis) when the ITS are analysed with OLS (A) and REML (B) for the fixed effect (red circles), DL+WT (green diamonds) and DL+HKSJ (brown crosses), REML+WT (purple diamonds) and REML+HKSJ (blue crosses) meta-analysis methods versus slope-change heterogeneity (x-axis), by number of datapoints (horizontal facets) and number of included studies (vertical facets). Simulation settings include a **level-change of 1**, level-change heterogeneity of 0, **slope-change of 0**, and autocorrelation of 0.

DL, DerSimonian and Laird. dps, datapoints. HKSJ, Hartung-Knapp / Sidik-Jonkman. ITS, interrupted time series. OLS, ordinary least squares. REML, restricted maximum likelihood. WT, Wald-type.

#### 4.7 Autocorrelation variation

##### 4.7.1 Bias

Appendix Figure S112. Plots of bias of immediate level-change (y-axis) when the ITS are analysed with OLS (A) and REML (B) using the fixed effect (red circles), DL+WT (green diamonds) and DL+HKSJ (brown crosses), REML+WT (purple diamonds) and REML+HKSJ (blue crosses) meta-analysis methods versus categories of autocorrelation variability and level-change heterogeneity combinations (x-axis). Plots are presented separately by combinations of the number of included studies (rows) and the number of datapoints (columns). Simulation scenarios include a **level-change of 0**, level-change heterogeneity of 0, **slope-change of 0**, autocorrelation of 0.4.

DL, DerSimonian and Laird. dps, datapoints. HKSJ, Hartung-Knapp / Sidik-Jonkman. ITS, interrupted time series. OLS, ordinary least squares. REML, restricted maximum likelihood. WT, Wald-type.

Appendix Figure S113. Plots of bias of immediate level-change (y-axis) when the ITS are analysed with OLS (A) and REML (B) using the fixed effect (red circles), DL+WT (green diamonds) and DL+HKSJ (brown crosses), REML+WT (purple diamonds) and REML+HKSJ (blue crosses) meta-analysis methods versus categories of autocorrelation variability and level-change heterogeneity combinations (x-axis). Plots are presented separately by combinations of the number of included studies (rows) and the number of datapoints (columns). Simulation scenarios include a **level-change of 0**, level-change heterogeneity of 0, **slope-change of 0.1**, autocorrelation of 0.4.

DL, DerSimonian and Laird. dps, datapoints. HKSJ, Hartung-Knapp / Sidik-Jonkman. ITS, interrupted time series. OLS, ordinary least squares. REML, restricted maximum likelihood. WT, Wald-type.

Appendix Figure S114. Plots of bias of immediate level-change (y-axis) when the ITS are analysed with OLS (A) and REML (B) using the fixed effect (red circles), DL+WT (green diamonds) and DL+HKSJ (brown crosses), REML+WT (purple diamonds) and REML+HKSJ (blue crosses) meta-analysis methods versus categories of autocorrelation variability and level-change heterogeneity combinations (x-axis). Plots are presented separately by combinations of the number of included studies (rows) and the number of datapoints (columns). Simulation scenarios include a **level-change of 1**, level-change heterogeneity of 0, **slope-change of 0**, autocorrelation of 0.4.

DL, DerSimonian and Laird. dps, datapoints. HKSJ, Hartung-Knapp / Sidik-Jonkman. ITS, interrupted time series. OLS, ordinary least squares. REML, restricted maximum likelihood. WT, Wald-type.

Appendix Figure S115. Plot of bias of slope-change (y-axis) when the ITS are analysed with OLS (A) and REML (B) for the fixed effect (red circles), DL+WT (green diamonds) and DL+HKSJ (brown crosses), REML+WT (purple diamonds) and REML+HKSJ (blue crosses) meta-analysis methods versus categories of autocorrelation variability and slope-change heterogeneity combinations (x-axis). Plots are presented separately by combinations of the number of included studies (rows) and the number of datapoints (columns). Simulation scenarios include a **level-change of 0**, level-change heterogeneity of 0, **slope-change of 0**, autocorrelation of 0.4.

DL, DerSimonian and Laird. dps, datapoints. HKSJ, Hartung-Knapp / Sidik-Jonkman. ITS, interrupted time series. OLS, ordinary least squares. REML, restricted maximum likelihood. WT, Wald-type.

Appendix Figure S116. Plot of bias of slope-change (y-axis) when the ITS are analysed with OLS (A) and REML (B) for the fixed effect (red circles), DL+WT (green diamonds) and DL+HKSJ (brown crosses), REML+WT (purple diamonds) and REML+HKSJ (blue crosses) meta-analysis methods versus categories of autocorrelation variability and slope-change heterogeneity combinations (x-axis). Plots are presented separately by combinations of the number of included studies (rows) and the number of datapoints (columns). Simulation scenarios include a **level-change of 0**, level-change heterogeneity of 0, **slope-change of 0.1**, autocorrelation of 0.4.

DL, DerSimonian and Laird. dps, datapoints. HKSJ, Hartung-Knapp / Sidik-Jonkman. ITS, interrupted time series. OLS, ordinary least squares. REML, restricted maximum likelihood. WT, Wald-type.

Appendix Figure S117. Plot of bias of slope-change (y-axis) when the ITS are analysed with OLS (A) and REML (B) for the fixed effect (red circles), DL+WT (green diamonds) and DL+HKSJ (brown crosses), REML+WT (purple diamonds) and REML+HKSJ (blue crosses) meta-analysis methods versus categories of autocorrelation variability and slope-change heterogeneity combinations (x-axis). Plots are presented separately by combinations of the number of included studies (rows) and the number of datapoints (columns). Simulation scenarios include a **level-change of 1**, level-change heterogeneity of 0, **slope-change of 0**, autocorrelation of 0.4.

DL, DerSimonian and Laird. dps, datapoints. HKSJ, Hartung-Knapp / Sidik-Jonkman. ITS, interrupted time series. OLS, ordinary least squares. REML, restricted maximum likelihood. WT, Wald-type.

##### 4.7.2 Coverage

Appendix Figure S118. Plots of 95% confidence interval coverage of immediate level-change (y-axis) when the ITS are analysed with OLS (A) and REML (B) using the fixed effect (red circles), DL+WT (green diamonds) and DL+HKSJ (brown crosses), REML+WT (purple diamonds) and REML+HKSJ (blue crosses) meta-analysis methods versus categories of autocorrelation variability and level-change heterogeneity combinations (x-axis). Plots are presented separately by combinations of the number of included studies (rows) and the number of datapoints (columns). Simulation scenarios include a **level-change of 0**, level-change heterogeneity of 0, **slope-change of 0**, autocorrelation of 0.4.

DL, DerSimonian and Laird. dps, datapoints. HKSJ, Hartung-Knapp / Sidik-Jonkman. ITS, interrupted time series. OLS, ordinary least squares. REML, restricted maximum likelihood. WT, Wald-type.

Appendix Figure S119. Plots of 95% confidence interval coverage of immediate level-change (y-axis) when the ITS are analysed with OLS (A) and REML (B) using the fixed effect (red circles), DL+WT (green diamonds) and DL+HKSJ (brown crosses), REML+WT (purple diamonds) and REML+HKSJ (blue crosses) meta-analysis methods versus categories of autocorrelation variability and level-change heterogeneity combinations (x-axis). Plots are presented separately by combinations of the number of included studies (rows) and the number of datapoints (columns). Simulation scenarios include a **level-change of 0**, level-change heterogeneity of 0, **slope-change of 0.1**, autocorrelation of 0.4.

DL, DerSimonian and Laird. dps, datapoints. HKSJ, Hartung-Knapp / Sidik-Jonkman. ITS, interrupted time series. OLS, ordinary least squares. REML, restricted maximum likelihood. WT, Wald-type.

Appendix Figure S120. Plots of 95% confidence interval coverage of immediate level-change (y-axis) when the ITS are analysed with OLS (A) and REML (B) using the fixed effect (red circles), DL+WT (green diamonds) and DL+HKSJ (brown crosses), REML+WT (purple diamonds) and REML+HKSJ (blue crosses) meta-analysis methods versus categories of autocorrelation variability and level-change heterogeneity combinations (x-axis). Plots are presented separately by combinations of the number of included studies (rows) and the number of datapoints (columns). Simulation scenarios include a **level-change of 1**, level-change heterogeneity of 0, **slope-change of 0**, autocorrelation of 0.4

DL, DerSimonian and Laird. dps, datapoints. HKSJ, Hartung-Knapp / Sidik-Jonkman. ITS, interrupted time series. OLS, ordinary least squares. REML, restricted maximum likelihood. WT, Wald-type.

Appendix Figure S121. The 95% confidence interval coverage of slope-change (y-axis) when the ITS are analysed with OLS (A) and REML (B) for the fixed effect (red circles), DL+WT (green diamonds) and DL+HKSJ (brown crosses), REML+WT (purple diamonds) and REML+HKSJ (blue crosses) meta-analysis methods versus categories of autocorrelation variability and slope-change heterogeneity combinations (x-axis). Plots are presented separately by combinations of the number of included studies (rows) and the number of datapoints (columns). Simulation scenarios include a **level-change of 0**, level-change heterogeneity of 0, **slope-change of 0**, autocorrelation of 0.4.

DL, DerSimonian and Laird. dps, datapoints. HKSJ, Hartung-Knapp / Sidik-Jonkman. ITS, interrupted time series. OLS, ordinary least squares. REML, restricted maximum likelihood. WT, Wald-type.

Appendix Figure S122. The 95% confidence interval coverage of slope-change (y-axis) when the ITS are analysed with OLS (A) and REML (B) for the fixed effect (red circles), DL+WT (green diamonds) and DL+HKSJ (brown crosses), REML+WT (purple diamonds) and REML+HKSJ (blue crosses) meta-analysis methods versus categories of autocorrelation variability and slope-change heterogeneity combinations (x-axis). Plots are presented separately by combinations of the number of included studies (rows) and the number of datapoints (columns). Simulation scenarios include a **level-change of 0**, level-change heterogeneity of 0, **slope-change of 0.1**, autocorrelation of 0.4.

DL, DerSimonian and Laird. dps, datapoints. HKSJ, Hartung-Knapp / Sidik-Jonkman. ITS, interrupted time series. OLS, ordinary least squares. REML, restricted maximum likelihood. WT, Wald-type.

Appendix Figure S123. The 95% confidence interval coverage of slope-change (y-axis) when the ITS are analysed with OLS (A) and REML (B) for the fixed effect (red circles), DL+WT (green diamonds) and DL+HKSJ (brown crosses), REML+WT (purple diamonds) and REML+HKSJ (blue crosses) meta-analysis methods versus categories of autocorrelation variability and slope-change heterogeneity combinations (x-axis). Plots are presented separately by combinations of the number of included studies (rows) and the number of datapoints (columns). Simulation scenarios include a **level-change of 1**, level-change heterogeneity of 0, **slope-change of 0**, autocorrelation of 0.4.

DL, DerSimonian and Laird. dps, datapoints. HKSJ, Hartung-Knapp / Sidik-Jonkman. ITS, interrupted time series. OLS, ordinary least squares. REML, restricted maximum likelihood. WT, Wald-type.

##### 4.7.3 Empirical standard error

Appendix Figure S124. Plots of empirical standard error (empSE) of the immediate level-change (y-axis) when the ITS are analysed with OLS (A) and REML (B) for the fixed effect (red circles), DL+WT (green diamonds) and DL+HKSJ (brown crosses), REML+WT (purple diamonds) and REML+HKSJ (blue crosses) meta-analysis methods versus categories of autocorrelation variability and level-change heterogeneity combinations (x-axis). Plots are presented separately by combinations of the number of included studies (rows) and the number of datapoints (columns). Simulation scenarios include a **level-change of 0**, level-change heterogeneity of 0, **slope-change of 0**, autocorrelation of 0.4.

DL, DerSimonian and Laird. dps, datapoints. HKSJ, Hartung-Knapp / Sidik-Jonkman. ITS, interrupted time series. OLS, ordinary least squares. REML, restricted maximum likelihood. WT, Wald-type.

Appendix Figure S125. Plots of empirical standard error (empSE) of the immediate level-change (y-axis) when the ITS are analysed with OLS (A) and REML (B) for the fixed effect (red circles), DL+WT (green diamonds) and DL+HKSJ (brown crosses), REML+WT (purple diamonds) and REML+HKSJ (blue crosses) meta-analysis methods versus categories of autocorrelation variability and level-change heterogeneity combinations (x-axis). Plots are presented separately by combinations of the number of included studies (rows) and the number of datapoints (columns). Simulation scenarios include a **level-change of 0**, level-change heterogeneity of 0, **slope-change of 0.1**, autocorrelation of 0.4

DL, DerSimonian and Laird. dps, datapoints. HKSJ, Hartung-Knapp / Sidik-Jonkman. ITS, interrupted time series. OLS, ordinary least squares. REML, restricted maximum likelihood. WT, Wald-type.

Appendix Figure S126. Plots of empirical standard error (empSE) of the immediate level-change (y-axis) when the ITS are analysed with OLS (A) and REML (B) for the fixed effect (red circles), DL+WT (green diamonds) and DL+HKSJ (brown crosses), REML+WT (purple diamonds) and REML+HKSJ (blue crosses) meta-analysis methods versus categories of autocorrelation variability and level-change heterogeneity combinations (x-axis). Plots are presented separately by combinations of the number of included studies (rows) and the number of datapoints (columns). Simulation scenarios include a **level-change of 1**, level-change heterogeneity of 0, **slope-change of 0**, autocorrelation of 0.4

DL, DerSimonian and Laird. dps, datapoints. HKSJ, Hartung-Knapp / Sidik-Jonkman. ITS, interrupted time series. OLS, ordinary least squares. REML, restricted maximum likelihood. WT, Wald-type.

Appendix Figure S127. Plots of empirical standard error (empSE) of the slope-change (y-axis) when the ITS are analysed with OLS (A) and REML (B) for the fixed effect (red circles), DL+WT (green diamonds) and DL+HKSJ (brown crosses), REML+WT (purple diamonds) and REML+HKSJ (blue crosses) meta-analysis methods versus categories of autocorrelation variability and slope-change heterogeneity combinations (x-axis). Plots are presented separately by combinations of the number of included studies (rows) and the number of datapoints (columns). Simulation scenarios include a **level-change of 0**, level-change heterogeneity of 0, **slope-change of 0**, autocorrelation of 0.4

DL, DerSimonian and Laird. Dps, datapoints. HKSJ, Hartung-Knapp / Sidik-Jonkman. ITS, interrupted time series. OLS, ordinary least squares. REML, restricted maximum likelihood. WT, Wald-type.

Appendix Figure S128. Plots of empirical standard error (empSE) of the immediate level-change (y-axis) when the ITS are analysed with OLS (A) and REML (B) for the fixed effect (red circles), DL+WT (green diamonds) and DL+HKSJ (brown crosses), REML+WT (purple diamonds) and REML+HKSJ (blue crosses) meta-analysis methods versus categories of autocorrelation variability and slope-change heterogeneity combinations (x-axis). Plots are presented separately by combinations of the number of included studies (rows) and the number of datapoints (columns). Simulation scenarios include a **level-change of 0**, level-change heterogeneity of 0, **slope-change of 0.1**, autocorrelation of 0.4

DL, DerSimonian and Laird. dps, datapoints. HKSJ, Hartung-Knapp / Sidik-Jonkman. ITS, interrupted time series. OLS, ordinary least squares. REML, restricted maximum likelihood. WT, Wald-type.

Appendix Figure S129. Plots of empirical standard error (empSE) of the slope-change (y-axis) when the ITS are analysed with OLS (A) and REML (B) for the fixed effect (red circles), DL+WT (green diamonds) and DL+HKSJ (brown crosses), REML+WT (purple diamonds) and REML+HKSJ (blue crosses) meta-analysis methods versus categories of autocorrelation variability and slope-change heterogeneity combinations (x-axis). Plots are presented separately by combinations of the number of included studies (rows) and the number of datapoints (columns). Simulation scenarios include a **level-change of 1**, level-change heterogeneity of 0, **slope-change of 0**, autocorrelation of 0.4

DL, DerSimonian and Laird. dps, datapoints. HKSJ, Hartung-Knapp / Sidik-Jonkman. ITS, interrupted time series. OLS, ordinary least squares. REML, restricted maximum likelihood. WT, Wald-type.

##### 4.7.4 Ratio of model based standard errors to empirical standard errors

Appendix Figure S130. Plots of the ratio of the empirical standard error (empSE) to the model based standard error (modSE) of the meta-analytic immediate level-change (y-axis) when the ITS are analysed with OLS (A) and REML (B) for the fixed effect (red circles), DL+WT (green diamonds) and DL+HKSJ (brown crosses), REML+WT (purple diamonds) and REML+HKSJ (blue crosses) meta-analysis methods versus categories of autocorrelation variability and level-change heterogeneity combinations (x-axis). Plots are presented separately by combinations of the number of included studies (rows) and the number of datapoints (columns). Simulation scenarios include a **level-change of 0,** level-change heterogeneity of 0, **slope-change of 0**, autocorrelation of 0.4.

DL, DerSimonian and Laird. dps, datapoints. HKSJ, Hartung-Knapp / Sidik-Jonkman. ITS, interrupted time series. OLS, ordinary least squares. REML, restricted maximum likelihood. WT, Wald-type.

Appendix Figure S131. Plots of the ratio of the empirical standard error (empSE) to the model based standard error (modSE) of the meta-analytic immediate level-change (y-axis) when the ITS are analysed with OLS (A) and REML (B) for the fixed effect (red circles), DL+WT (green diamonds) and DL+HKSJ (brown crosses), REML+WT (purple diamonds) and REML+HKSJ (blue crosses) meta-analysis methods versus categories of autocorrelation variability and level-change heterogeneity combinations (x-axis). Plots are presented separately by combinations of the number of included studies (rows) and the number of datapoints (columns). Simulation scenarios include a **level-change of 0,** level-change heterogeneity of 0, **slope-change of 0.1**, autocorrelation of 0.4.

DL, DerSimonian and Laird. dps, datapoints. HKSJ, Hartung-Knapp / Sidik-Jonkman. ITS, interrupted time series. OLS, ordinary least squares. REML, restricted maximum likelihood. WT, Wald-type.

Appendix Figure S132. Plots of the ratio of the empirical standard error (empSE) to the model based standard error (modSE) of the meta-analytic immediate level-change (y-axis) when the ITS are analysed with OLS (A) and REML (B) for the fixed effect (red circles), DL+WT (green diamonds) and DL+HKSJ (brown crosses), REML+WT (purple diamonds) and REML+HKSJ (blue crosses) meta-analysis methods versus categories of autocorrelation variability and level-change heterogeneity combinations (x-axis). Plots are presented separately by combinations of the number of included studies (rows) and the number of datapoints (columns). Simulation scenarios include a **level-change of 1,** level-change heterogeneity of 0, **slope-change of 0**, autocorrelation of 0.4.

DL, DerSimonian and Laird. dps, datapoints. HKSJ, Hartung-Knapp / Sidik-Jonkman. ITS, interrupted time series. OLS, ordinary least squares. REML, restricted maximum likelihood. WT, Wald-type.

Appendix Figure S133. Plots of the ratio of the empirical standard error (empSE) to the model based standard error (modSE) of the meta-analytic slope-change (y-axis) when the ITS are analysed with OLS (A) and REML (B) for the fixed effect (red circles), DL+WT (green diamonds) and DL+HKSJ (brown crosses), REML+WT (purple diamonds) and REML+HKSJ (blue crosses) meta-analysis methods versus categories of autocorrelation variability and slope-change heterogeneity combinations (x-axis). Plots are presented separately by combinations of the number of included studies (rows) and the number of datapoints (columns). Simulation scenarios include a **level-change of 0,** level-change heterogeneity of 0, **slope-change of 0**, autocorrelation of 0.4.

DL, DerSimonian and Laird. dps, datapoints. HKSJ, Hartung-Knapp / Sidik-Jonkman. ITS, interrupted time series. OLS, ordinary least squares. REML, restricted maximum likelihood. WT, Wald-type.

Appendix Figure S134. Plots of the ratio of the empirical standard error (empSE) to the model based standard error (modSE) of the meta-analytic slope-change (y-axis) when the ITS are analysed with OLS (A) and REML (B) for the fixed effect (red circles), DL+WT (green diamonds) and DL+HKSJ (brown crosses), REML+WT (purple diamonds) and REML+HKSJ (blue crosses) meta-analysis methods versus categories of autocorrelation variability and slope-change heterogeneity combinations (x-axis). Plots are presented separately by combinations of the number of included studies (rows) and the number of datapoints (columns). Simulation scenarios include a **level-change of 0,** level-change heterogeneity of 0, **slope-change of 0.1**, autocorrelation of 0.4.

DL, DerSimonian and Laird. dps, datapoints. HKSJ, Hartung-Knapp / Sidik-Jonkman. ITS, interrupted time series. OLS, ordinary least squares. REML, restricted maximum likelihood. WT, Wald-type.

Appendix Figure S135. Plots of the ratio of the empirical standard error (empSE) to the model based standard error (modSE) of the meta-analytic slope-change (y-axis) when the ITS are analysed with OLS (A) and REML (B) for the fixed effect (red circles), DL+WT (green diamonds) and DL+HKSJ (brown crosses), REML+WT (purple diamonds) and REML+HKSJ (blue crosses) meta-analysis methods versus categories of autocorrelation variability and slope-change heterogeneity combinations (x-axis). Plots are presented separately by combinations of the number of included studies (rows) and the number of datapoints (columns). Simulation scenarios include a **level-change of 1,** level-change heterogeneity of 0, **slope-change of 0**, autocorrelation of 0.4.

DL, DerSimonian and Laird. dps, datapoints. HKSJ, Hartung-Knapp / Sidik-Jonkman. ITS, interrupted time series. OLS, ordinary least squares. REML, restricted maximum likelihood. WT, Wald-type.

##### 4.7.5 Statistical power

Appendix Figure S136. Plots of statistical power (the percentage of simulations that have a 95% confidence interval that did not include zero – only scenarios with a confidence interval coverage of greater than 90% are plotted) of the meta-analytic level-change (y-axis) when the ITS are analysed with OLS (A) and REML (B) for the fixed effect (red circles), DL+WT (green diamonds) and DL+HKSJ (brown crosses), REML+WT (purple diamonds) and REML+HKSJ (blue crosses) meta-analysis methods versus categories of autocorrelation variability and level-change heterogeneity combinations (x-axis). Plots are presented separately by combinations of the number of included studies (rows) and the number of datapoints (columns). Simulation scenarios include a **level-change of 0,** level-change heterogeneity of 0, **slope-change of 0**, autocorrelation of 0.4.

DL, DerSimonian and Laird. dps, datapoints. HKSJ, Hartung-Knapp / Sidik-Jonkman. ITS, interrupted time series. OLS, ordinary least squares. REML, restricted maximum likelihood. WT, Wald-type.

Appendix Figure S137. Plots of statistical power (the percentage of simulations that have a 95% confidence interval that did not include zero – only scenarios with a confidence interval coverage of greater than 90% are plotted) of the meta-analytic level-change (y-axis) when the ITS are analysed with OLS (A) and REML (B) for the fixed effect (red circles), DL+WT (green diamonds) and DL+HKSJ (brown crosses), REML+WT (purple diamonds) and REML+HKSJ (blue crosses) meta-analysis methods versus categories of autocorrelation variability and level-change heterogeneity combinations (x-axis). Plots are presented separately by combinations of the number of included studies (rows) and the number of datapoints (columns). Simulation scenarios include a **level-change of 0,** level-change heterogeneity of 0, **slope-change of 0.1**, autocorrelation of 0.4.

DL, DerSimonian and Laird. dps, datapoints. HKSJ, Hartung-Knapp / Sidik-Jonkman. ITS, interrupted time series. OLS, ordinary least squares. REML, restricted maximum likelihood. WT, Wald-type.

Appendix Figure S138. Plots of statistical power (the percentage of simulations that have a 95% confidence interval that did not include zero – only scenarios with a confidence interval coverage of greater than 90% are plotted) of the meta-analytic level-change (y-axis) when the ITS are analysed with OLS (A) and REML (B) for the fixed effect (red circles), DL+WT (green diamonds) and DL+HKSJ (brown crosses), REML+WT (purple diamonds) and REML+HKSJ (blue crosses) meta-analysis methods versus categories of autocorrelation variability and level-change heterogeneity combinations (x-axis). Plots are presented separately by combinations of the number of included studies (rows) and the number of datapoints (columns). Simulation scenarios include a **level-change of 1,** level-change heterogeneity of 0, **slope-change of 0**, autocorrelation of 0.4.

DL, DerSimonian and Laird. dps, datapoints. HKSJ, Hartung-Knapp / Sidik-Jonkman. ITS, interrupted time series. OLS, ordinary least squares. REML, restricted maximum likelihood. WT, Wald-type.

Appendix Figure S139. Plots of statistical power (the percentage of simulations that have a 95% confidence interval that did not include zero – only scenarios with a confidence interval coverage of greater than 90% are plotted) of the meta-analytic slope-change (y-axis) when the ITS are analysed with OLS (A) and REML (B) for the fixed effect (red circles), DL+WT (green diamonds) and DL+HKSJ (brown crosses), REML+WT (purple diamonds) and REML+HKSJ (blue crosses) meta-analysis methods versus categories of autocorrelation variability and slope-change heterogeneity combinations (x-axis). Plots are presented separately by combinations of the number of included studies (rows) and the number of datapoints (columns). Simulation scenarios include a **level-change of 0,** level-change heterogeneity of 0, **slope-change of 0**, autocorrelation of 0.4.

DL, DerSimonian and Laird. dps, datapoints. HKSJ, Hartung-Knapp / Sidik-Jonkman. ITS, interrupted time series. OLS, ordinary least squares. REML, restricted maximum likelihood. WT, Wald-type.

Appendix Figure S140. Plots of statistical power (the percentage of simulations that have a 95% confidence interval that did not include zero – only scenarios with a confidence interval coverage of greater than 90% are plotted) of the meta-analytic slope-change (y-axis) when the ITS are analysed with OLS (A) and REML (B) for the fixed effect (red circles), DL+WT (green diamonds) and DL+HKSJ (brown crosses), REML+WT (purple diamonds) and REML+HKSJ (blue crosses) meta-analysis methods versus categories of autocorrelation variability and slope-change heterogeneity combinations (x-axis). Plots are presented separately by combinations of the number of included studies (rows) and the number of datapoints (columns). Simulation scenarios include a **level-change of 0,** level-change heterogeneity of 0, **slope-change of 0.1**, autocorrelation of 0.4.

DL, DerSimonian and Laird. dps, datapoints. HKSJ, Hartung-Knapp / Sidik-Jonkman. ITS, interrupted time series. OLS, ordinary least squares. REML, restricted maximum likelihood. WT, Wald-type.

Appendix Figure S141. Plots of statistical power (the percentage of simulations that have a 95% confidence interval that did not include zero – only scenarios with a confidence interval coverage of greater than 90% are plotted) of the meta-analytic slope-change (y-axis) when the ITS are analysed with OLS (A) and REML (B) for the fixed effect (red circles), DL+WT (green diamonds) and DL+HKSJ (brown crosses), REML+WT (purple diamonds) and REML+HKSJ (blue crosses) meta-analysis methods versus categories of autocorrelation variability and slope-change heterogeneity combinations (x-axis). Plots are presented separately by combinations of the number of included studies (rows) and the number of datapoints (columns). Simulation scenarios include a **level-change of 1,** level-change heterogeneity of 0, **slope-change of 0**, autocorrelation of 0.4.

DL, DerSimonian and Laird. dps, datapoints. HKSJ, Hartung-Knapp / Sidik-Jonkman. ITS, interrupted time series. OLS, ordinary least squares. REML, restricted maximum likelihood. WT, Wald-type.

#### 4.8 Estimated heterogeneity

##### 4.8.1 Heterogeneity in level-change parameter

| Appendix Figure S142. Plots of level-change heterogeneity estimated using a random effects meta-analysis with the REML between-study variance estimator (y-axis) when the A) 3 ITS, B) 5 ITS and C) 20 ITS studies were analysed with OLS (orange) and REML (purple) ITS analysis methods. Plots are presented separately by combinations of the true level-change heterogeneity (rows) and the number of datapoints (columns). The solid red lines indicate the true level-change heterogeneity. Simulation scenarios include a **level-change of 0**, slope-change heterogeneity of 0, **slope-change of 0**, and fixed autocorrelation.  DL, DerSimonian and Laird. dps, datapoints. HKSJ, Hartung-Knapp / Sidik-Jonkman. ITS, interrupted time series. OLS, ordinary least squares. REML, restricted maximum likelihood. WT, Wald-type. | Appendix Figure S143. Plots of level-change heterogeneity estimated using a random effects meta-analysis with the REML between-study variance estimator (y-axis) when the A) 3 ITS, B) 5 ITS and C) 20 ITS studies were analysed with OLS (orange) and REML (purple) ITS analysis methods. Plots are presented separately by combinations of the true level-change heterogeneity (rows) and the number of datapoints (columns). The solid red lines indicate the true level-change heterogeneity. Simulation scenarios include a **level-change of 0**, slope-change heterogeneity of 0, **slope-change of 0.1**, and fixed autocorrelation.  DL, DerSimonian and Laird. dps, datapoints. HKSJ, Hartung-Knapp / Sidik-Jonkman. ITS, interrupted time series. OLS, ordinary least squares. REML, restricted maximum likelihood. WT, Wald-type. | Appendix Figure S144. Plots of level-change heterogeneity estimated using a random effects meta-analysis with the REML between-study variance estimator (y-axis) when the A) 3 ITS, B) 5 ITS and C) 20 ITS studies were analysed with OLS (orange) and REML (purple) ITS analysis methods. Plots are presented separately by combinations of the true level-change heterogeneity (rows) and the number of datapoints (columns). The solid red lines indicate the true level-change heterogeneity. Simulation scenarios include a **level-change of 1**, slope-change heterogeneity of 0, **slope-change of 0**, and fixed autocorrelation.  DL, DerSimonian and Laird. dps, datapoints. HKSJ, Hartung-Knapp / Sidik-Jonkman. ITS, interrupted time series. OLS, ordinary least squares. REML, restricted maximum likelihood. WT, Wald-type. |
| --- | --- | --- |
| Appendix Figure S145. Plots of level-change heterogeneity estimated using a random effects meta-analysis with DL (green) and REML (blue) between-study variance estimators (y-axis) when the A) 3 ITS, B) 5 ITS and C) 20 ITS studies were analysed with OLS. Plots are presented separately by combinations of the true level-change heterogeneity (rows) and the number of datapoints (columns). The solid red lines indicate the true level-change heterogeneity. Simulation scenarios **include a level-change of 0, slope-change of 0,** slope-change heterogeneity of 0, and fixed autocorrelation.  DL, DerSimonian and Laird. dps, datapoints. HKSJ, Hartung-Knapp / Sidik-Jonkman. ITS, interrupted time series. OLS, ordinary least squares. REML, restricted maximum likelihood. WT, Wald-type. | Appendix Figure S146. Plots of level-change heterogeneity estimated using a random effects meta-analysis with DL (green) and REML (blue) between-study variance estimators (y-axis) when the A) 3 ITS, B) 5 ITS and C) 20 ITS studies were analysed with OLS. Plots are presented separately by combinations of the true level-change heterogeneity (rows) and the number of datapoints (columns). The solid red lines indicate the true level-change heterogeneity. Simulation scenarios **include a level-change of 0, slope-change of 0.1,** slope-change heterogeneity of 0, and fixed autocorrelation.  DL, DerSimonian and Laird. dps, datapoints. HKSJ, Hartung-Knapp / Sidik-Jonkman. ITS, interrupted time series. OLS, ordinary least squares. REML, restricted maximum likelihood. WT, Wald-type. | Appendix Figure S147. Plots of level-change heterogeneity estimated using a random effects meta-analysis with DL (green) and REML (blue) between-study variance estimators (y-axis) when the A) 3 ITS, B) 5 ITS and C) 20 ITS studies were analysed with OLS. Plots are presented separately by combinations of the true level-change heterogeneity (rows) and the number of datapoints (columns). The solid red lines indicate the true level-change heterogeneity. Simulation scenarios **include a level-change of 1, slope-change of 0,** slope-change heterogeneity of 0, and fixed autocorrelation.  DL, DerSimonian and Laird. dps, datapoints. HKSJ, Hartung-Knapp / Sidik-Jonkman. ITS, interrupted time series. OLS, ordinary least squares. REML, restricted maximum likelihood. WT, Wald-type. |

| Appendix Figure S148. Plots of level-change heterogeneity estimated using a random effects meta-analysis with DL (green) and REML (blue) between-study variance estimators (y-axis) when the A) 3 ITS, B) 5 ITS and C) 20 ITS studies were analysed with REML. Plots are presented separately by combinations of the true level-change heterogeneity (rows) and the number of datapoints (columns). The solid red lines indicate the true level-change heterogeneity. Simulation scenarios **include a level-change of 0, slope-change of 0,** slope-change heterogeneity of 0, and fixed autocorrelation.  DL, DerSimonian and Laird. dps, datapoints. HKSJ, Hartung-Knapp / Sidik-Jonkman. ITS, interrupted time series. OLS, ordinary least squares. REML, restricted maximum likelihood. WT, Wald-type. | Appendix Figure S149. Plots of level-change heterogeneity estimated using a random effects meta-analysis with DL (green) and REML (blue) between-study variance estimators (y-axis) when the A) 3 ITS, B) 5 ITS and C) 20 ITS studies were analysed with REML. Plots are presented separately by combinations of the true level-change heterogeneity (rows) and the number of datapoints (columns). The solid red lines indicate the true level-change heterogeneity. Simulation scenarios **include a level-change of 0, slope-change of 0.1,** slope-change heterogeneity of 0, and fixed autocorrelation.  DL, DerSimonian and Laird. dps, datapoints. HKSJ, Hartung-Knapp / Sidik-Jonkman. ITS, interrupted time series. OLS, ordinary least squares. REML, restricted maximum likelihood. WT, Wald-type. | Appendix Figure S150. Plots of level-change heterogeneity estimated using a random effects meta-analysis with DL (green) and REML (blue) between-study variance estimators (y-axis) when the A) 3 ITS, B) 5 ITS and C) 20 ITS studies were analysed with REML. Plots are presented separately by combinations of the true level-change heterogeneity (rows) and the number of datapoints (columns). The solid red lines indicate the true level-change heterogeneity. Simulation scenarios **include a level-change of 1, slope-change of 0,** slope-change heterogeneity of 0, and fixed autocorrelation.  DL, DerSimonian and Laird. dps, datapoints. HKSJ, Hartung-Knapp / Sidik-Jonkman. ITS, interrupted time series. OLS, ordinary least squares. REML, restricted maximum likelihood. WT, Wald-type. |
| --- | --- | --- |

##### 4.8.2 Heterogeneity in slope-change parameter

| Appendix Figure S151. Plots of slope-change heterogeneity estimated using a random effects meta-analysis with the REML between-study variance estimator (y-axis) when the A) 3 ITS, B) 5 ITS and C) 20 ITS studies were analysed with OLS (orange) and REML (purple) ITS analysis methods. Plots are presented separately by combinations of the true slope-change heterogeneity (rows) and the number of datapoints (columns). The solid red lines indicate the true slope-change heterogeneity. Simulation scenarios include a **level-change of 0**, level-change heterogeneity of 0, **slope-change of 0**, and fixed autocorrelation.  DL, DerSimonian and Laird. dps, datapoints. HKSJ, Hartung-Knapp / Sidik-Jonkman. ITS, interrupted time series. OLS, ordinary least squares. REML, restricted maximum likelihood. WT, Wald-type. | Appendix Figure S152. Plots of slope-change heterogeneity estimated using a random effects meta-analysis with the REML between-study variance estimator (y-axis) when the A) 3 ITS, B) 5 ITS and C) 20 ITS studies were analysed with OLS (orange) and REML (purple) ITS analysis methods. Plots are presented separately by combinations of the true slope-change heterogeneity (rows) and the number of datapoints (columns). The solid red lines indicate the true slope-change heterogeneity. Simulation scenarios include a **level-change of 0**, level-change heterogeneity of 0, **slope-change of 0.1**, and fixed autocorrelation.  DL, DerSimonian and Laird. dps, datapoints. HKSJ, Hartung-Knapp / Sidik-Jonkman. ITS, interrupted time series. OLS, ordinary least squares. REML, restricted maximum likelihood. WT, Wald-type. | Appendix Figure S153. Plots of slope-change heterogeneity estimated using a random effects meta-analysis with the REML between-study variance estimator (y-axis) when the A) 3 ITS, B) 5 ITS and C) 20 ITS studies were analysed with OLS (orange) and REML (purple) ITS analysis methods. Plots are presented separately by combinations of the true slope-change heterogeneity (rows) and the number of datapoints (columns). The solid red lines indicate the true slope-change heterogeneity. Simulation scenarios include a **level-change of 1**, level-change heterogeneity of 0, **slope-change of 0**, and fixed autocorrelation.  DL, DerSimonian and Laird. dps, datapoints. HKSJ, Hartung-Knapp / Sidik-Jonkman. ITS, interrupted time series. OLS, ordinary least squares. REML, restricted maximum likelihood. WT, Wald-type. |
| --- | --- | --- |

| Appendix Figure S154. Plots of slope-change heterogeneity estimated using a random effects meta-analysis with DL (green) and REML (blue) between-study variance estimators (y-axis) when the A) 3 ITS, B) 5 ITS and C) 20 ITS studies were analysed with OLS. Plots are presented separately by combinations of the true level-change heterogeneity (rows) and the number of datapoints (columns). The solid red lines indicate the true slope-change heterogeneity. Simulation scenarios **include a level-change of 0, slope-change of 0,** level-change heterogeneity of 0, and fixed autocorrelation.  DL, DerSimonian and Laird. dps, datapoints. HKSJ, Hartung-Knapp / Sidik-Jonkman. ITS, interrupted time series. OLS, ordinary least squares. REML, restricted maximum likelihood. WT, Wald-type. | Appendix Figure S155. Plots of slope-change heterogeneity estimated using a random effects meta-analysis with DL (green) and REML (blue) between-study variance estimators (y-axis) when the A) 3 ITS, B) 5 ITS and C) 20 ITS studies were analysed with OLS. Plots are presented separately by combinations of the true level-change heterogeneity (rows) and the number of datapoints (columns). The solid red lines indicate the true slope-change heterogeneity. Simulation scenarios **include a level-change of 0, slope-change of 0.1,** level-change heterogeneity of 0, and fixed autocorrelation.  DL, DerSimonian and Laird. dps, datapoints. HKSJ, Hartung-Knapp / Sidik-Jonkman. ITS, interrupted time series. OLS, ordinary least squares. REML, restricted maximum likelihood. WT, Wald-type. | Appendix Figure S156. Plots of level-change heterogeneity estimated using a random effects meta-analysis with DL (green) and REML (blue) between-study variance estimators (y-axis) when the A) 3 ITS, B) 5 ITS and C) 20 ITS studies were analysed with OLS. Plots are presented separately by combinations of the true level-change heterogeneity (rows) and the number of datapoints (columns). The solid red lines indicate the true slope-change heterogeneity. Simulation scenarios **include a level-change of 1, slope-change of 0,** level-change heterogeneity of 0, and fixed autocorrelation.  DL, DerSimonian and Laird. dps, datapoints. HKSJ, Hartung-Knapp / Sidik-Jonkman. ITS, interrupted time series. OLS, ordinary least squares. REML, restricted maximum likelihood. WT, Wald-type. |
| --- | --- | --- |

| Appendix Figure S157. Plots of slope-change heterogeneity estimated using a random effects meta-analysis with the REML between-study variance estimator (y-axis) when the A) 3 ITS, B) 5 ITS and C) 20 ITS studies were analysed with OLS (orange) and REML (blue) ITS analysis methods. Plots are presented separately by combinations of the true slope-change heterogeneity (rows) and the number of datapoints (columns). The solid red lines indicate the true slope-change heterogeneity. Simulation scenarios include a **level-change of 0**, level-change heterogeneity of 0, **slope-change of 0**, and fixed autocorrelation.  DL, DerSimonian and Laird. dps, datapoints. HKSJ, Hartung-Knapp / Sidik-Jonkman. ITS, interrupted time series. OLS, ordinary least squares. REML, restricted maximum likelihood. WT, Wald-type. | Appendix Figure S158. Plots of slope-change heterogeneity estimated using a random effects meta-analysis with the REML between-study variance estimator (y-axis) when the A) 3 ITS, B) 5 ITS and C) 20 ITS studies were analysed with OLS (orange) and REML (blue) ITS analysis methods. Plots are presented separately by combinations of the true slope-change heterogeneity (rows) and the number of datapoints (columns). The solid red lines indicate the true slope-change heterogeneity. Simulation scenarios include a **level-change of 0**, level-change heterogeneity of 0, **slope-change of 0.1**, and fixed autocorrelation.  DL, DerSimonian and Laird. dps, datapoints. HKSJ, Hartung-Knapp / Sidik-Jonkman. ITS, interrupted time series. OLS, ordinary least squares. REML, restricted maximum likelihood. WT, Wald-type. | Appendix Figure S159. Plots of slope-change heterogeneity estimated using a random effects meta-analysis with the REML between-study variance estimator (y-axis) when the A) 3 ITS, B) 5 ITS and C) 20 ITS studies were analysed with OLS (orange) and REML (blue) ITS analysis methods. Plots are presented separately by combinations of the true slope-change heterogeneity (rows) and the number of datapoints (columns). The solid red lines indicate the true slope-change heterogeneity. Simulation scenarios include a **level-change of 1**, level-change heterogeneity of 0, **slope-change of 0**, and fixed autocorrelation.  DL, DerSimonian and Laird. dps, datapoints. HKSJ, Hartung-Knapp / Sidik-Jonkman. ITS, interrupted time series. OLS, ordinary least squares. REML, restricted maximum likelihood. WT, Wald-type. |
| --- | --- | --- |

#### 4.9 Proportion of included studies analysed with REML vs PW

##### 4.9.1 Bias

Appendix Figure S160. Plots of bias of the immediate level-change (y-axis) when the ITS are analysed with PW (A) and REML (B) for the fixed effect (red circles), DL+WT (green diamonds) and DL+HKSJ (brown crosses), REML+WT (purple diamonds) and REML+HKSJ (blue crosses) meta-analysis methods versus autocorrelation (x-axis). Plots are presented separately by combinations of the number of included studies (rows) and level-change heterogeneity (columns). Simulation scenarios include ITS with 12 datapoints, a **level-change of 0, slope-change of 0**, slope-change heterogeneity of 0, and fixed levels of autocorrelation.

DL, DerSimonian and Laird. dps, datapoints. HKSJ, Hartung-Knapp / Sidik-Jonkman. ITS, interrupted time series. PW, Prais-Winsten. REML, restricted maximum likelihood. WT, Wald-type.

Appendix Figure S161. Plots of bias of the immediate level-change (y-axis) when the ITS are analysed with PW (A) and REML (B) for the fixed effect (red circles), DL+WT (green diamonds) and DL+HKSJ (brown crosses), REML+WT (purple diamonds) and REML+HKSJ (blue crosses) meta-analysis methods versus autocorrelation (x-axis). Plots are presented separately by combinations of the number of included studies (rows) and level-change heterogeneity (columns). Simulation scenarios include ITS with 12 datapoints, a **level-change of 0, slope-change of 0.1**, slope-change heterogeneity of 0, and fixed levels of autocorrelation.

DL, DerSimonian and Laird. dps, datapoints. HKSJ, Hartung-Knapp / Sidik-Jonkman. ITS, interrupted time series. PW, Prais-Winsten. REML, restricted maximum likelihood. WT, Wald-type.

Appendix Figure S162. Plots of bias of the immediate level-change (y-axis) when the ITS are analysed with PW (A) and REML (B) for the fixed effect (red circles), DL+WT (green diamonds) and DL+HKSJ (brown crosses), REML+WT (purple diamonds) and REML+HKSJ (blue crosses) meta-analysis methods versus autocorrelation (x-axis). Plots are presented separately by combinations of the number of included studies (rows) and level-change heterogeneity (columns). Simulation scenarios include ITS with 12 datapoints, a **level-change of 1, slope-change of 0**, slope-change heterogeneity of 0, and fixed levels of autocorrelation.

DL, DerSimonian and Laird. dps, datapoints. HKSJ, Hartung-Knapp / Sidik-Jonkman. ITS, interrupted time series. PW, Prais-Winsten. REML, restricted maximum likelihood. WT, Wald-type.

Appendix Figure S163. Plots of bias of the immediate level-change (y-axis) when the ITS are analysed with PW (A) and REML (B) for the fixed effect (red circles), DL+WT (green diamonds) and DL+HKSJ (brown crosses), REML+WT (purple diamonds) and REML+HKSJ (blue crosses) meta-analysis methods versus autocorrelation (x-axis). Plots are presented separately by combinations of the number of included studies (rows) and level-change heterogeneity (columns). Simulation scenarios include ITS with 12 datapoints, a **level-change of 0, slope-change of 0**, slope-change heterogeneity of 0, and fixed levels of autocorrelation.

DL, DerSimonian and Laird. dps, datapoints. HKSJ, Hartung-Knapp / Sidik-Jonkman. ITS, interrupted time series. PW, Prais-Winsten. REML, restricted maximum likelihood. WT, Wald-type.

Appendix Figure S164. Plots of bias of the immediate level-change (y-axis) when the ITS are analysed with PW (A) and REML (B) for the fixed effect (red circles), DL+WT (green diamonds) and DL+HKSJ (brown crosses), REML+WT (purple diamonds) and REML+HKSJ (blue crosses) meta-analysis methods versus autocorrelation (x-axis). Plots are presented separately by combinations of the number of included studies (rows) and level-change heterogeneity (columns). Simulation scenarios include ITS with 12 datapoints, a **level-change of 0, slope-change of 0.1**, slope-change heterogeneity of 0, and fixed levels of autocorrelation.

DL, DerSimonian and Laird. dps, datapoints. HKSJ, Hartung-Knapp / Sidik-Jonkman. ITS, interrupted time series. PW, Prais-Winsten. REML, restricted maximum likelihood. WT, Wald-type.

Appendix Figure S165. Plots of bias of the immediate level-change (y-axis) when the ITS are analysed with PW (A) and REML (B) for the fixed effect (red circles), DL+WT (green diamonds) and DL+HKSJ (brown crosses), REML+WT (purple diamonds) and REML+HKSJ (blue crosses) meta-analysis methods versus autocorrelation (x-axis). Plots are presented separately by combinations of the number of included studies (rows) and level-change heterogeneity (columns). Simulation scenarios include ITS with 12 datapoints, a **level-change of 1, slope-change of 0**, slope-change heterogeneity of 0, and fixed levels of autocorrelation.

DL, DerSimonian and Laird. dps, datapoints. HKSJ, Hartung-Knapp / Sidik-Jonkman. ITS, interrupted time series. PW, Prais-Winsten. REML, restricted maximum likelihood. WT, Wald-type.

##### 4.9.2 Coverage

Appendix Figure S166. Plots of 95% confidence interval coverage of the immediate level-change (y-axis) when the ITS are analysed with PW (A) and REML (B) for the fixed effect (red circles), DL+WT (green diamonds) and DL+HKSJ (brown crosses), REML+WT (purple diamonds) and REML+HKSJ (blue crosses) meta-analysis methods versus autocorrelation (x-axis). Plots are presented separately by combinations of the number of included studies (rows) and level-change heterogeneity (columns). Simulation scenarios include ITS with 12 datapoints, a **level-change of 0, slope-change of 0**, slope-change heterogeneity of 0, and fixed levels of autocorrelation.

DL, DerSimonian and Laird. dps, datapoints. HKSJ, Hartung-Knapp / Sidik-Jonkman. ITS, interrupted time series. PW, Prais-Winsten. REML, restricted maximum likelihood. WT, Wald-type.

Appendix Figure S167. Plots of 95% confidence interval coverage of the immediate level-change (y-axis) when the ITS are analysed with PW (A) and REML (B) for the fixed effect (red circles), DL+WT (green diamonds) and DL+HKSJ (brown crosses), REML+WT (purple diamonds) and REML+HKSJ (blue crosses) meta-analysis methods versus autocorrelation (x-axis). Plots are presented separately by combinations of the number of included studies (rows) and level-change heterogeneity (columns). Simulation scenarios include ITS with 12 datapoints, a **level-change of 0, slope-change of 0.1**, slope-change heterogeneity of 0, and fixed levels of autocorrelation.

DL, DerSimonian and Laird. dps, datapoints. HKSJ, Hartung-Knapp / Sidik-Jonkman. ITS, interrupted time series. PW, Prais-Winsten. REML, restricted maximum likelihood. WT, Wald-type.

Appendix Figure S168. Plots of 95% confidence interval coverage of the immediate level-change (y-axis) when the ITS are analysed with PW (A) and REML (B) for the fixed effect (red circles), DL+WT (green diamonds) and DL+HKSJ (brown crosses), REML+WT (purple diamonds) and REML+HKSJ (blue crosses) meta-analysis methods versus autocorrelation (x-axis). Plots are presented separately by combinations of the number of included studies (rows) and level-change heterogeneity (columns). Simulation scenarios include ITS with 12 datapoints, a **level-change of 1, slope-change of 0**, slope-change heterogeneity of 0, and fixed levels of autocorrelation.

DL, DerSimonian and Laird. dps, datapoints. HKSJ, Hartung-Knapp / Sidik-Jonkman. ITS, interrupted time series. PW, Prais-Winsten. REML, restricted maximum likelihood. WT, Wald-type.

Appendix Figure S169. Plots of 95% confidence interval coverage of slope-change (y-axis) when the ITS are analysed with PW (A) and REML (B) for the fixed effect (red circles), DL+WT (green diamonds) and DL+HKSJ (brown crosses), REML+WT (purple diamonds) and REML+HKSJ (blue crosses) meta-analysis methods versus autocorrelation (x-axis). Plots are presented separately by combinations of the number of included studies (rows) and level-change heterogeneity (columns). Simulation scenarios include ITS with 12 datapoints, a **level-change of 0, slope-change of 0**, slope-change heterogeneity of 0, and fixed levels of autocorrelation.

DL, DerSimonian and Laird. dps, datapoints. HKSJ, Hartung-Knapp / Sidik-Jonkman. ITS, interrupted time series. PW, Prais-Winsten. REML, restricted maximum likelihood. WT, Wald-type.

Appendix Figure S170. Plots of 95% confidence interval coverage of slope-change (y-axis) when the ITS are analysed with PW (A) and REML (B) for the fixed effect (red circles), DL+WT (green diamonds) and DL+HKSJ (brown crosses), REML+WT (purple diamonds) and REML+HKSJ (blue crosses) meta-analysis methods versus autocorrelation (x-axis). Plots are presented separately by combinations of the number of included studies (rows) and level-change heterogeneity (columns). Simulation scenarios include ITS with 12 datapoints, a **level-change of 0, slope-change of 0.1**, slope-change heterogeneity of 0, and fixed levels of autocorrelation.

DL, DerSimonian and Laird. dps, datapoints. HKSJ, Hartung-Knapp / Sidik-Jonkman. ITS, interrupted time series. PW, Prais-Winsten. REML, restricted maximum likelihood. WT, Wald-type.

Appendix Figure S171. Plots of 95% confidence interval coverage of slope-change (y-axis) when the ITS are analysed with PW (A) and REML (B) for the fixed effect (red circles), DL+WT (green diamonds) and DL+HKSJ (brown crosses), REML+WT (purple diamonds) and REML+HKSJ (blue crosses) meta-analysis methods versus autocorrelation (x-axis). Plots are presented separately by combinations of the number of included studies (rows) and level-change heterogeneity (columns). Simulation scenarios include ITS with 12 datapoints, a **level-change of 1, slope-change of 0**, slope-change heterogeneity of 0, and fixed levels of autocorrelation.

DL, DerSimonian and Laird. dps, datapoints. HKSJ, Hartung-Knapp / Sidik-Jonkman. ITS, interrupted time series. PW, Prais-Winsten. REML, restricted maximum likelihood. WT, Wald-type.

##### 4.9.3 Empirical standard error

Appendix Figure S172. Plots of the empirical standard error of the immediate level-change (y-axis) when the ITS are analysed with PW (A) and REML (B) for the fixed effect (red circles), DL+WT (green diamonds) and DL+HKSJ (brown crosses), REML+WT (purple diamonds) and REML+HKSJ (blue crosses) meta-analysis methods versus autocorrelation (x-axis). Plots are presented separately by combinations of the number of included studies (rows) and level-change heterogeneity (columns). Simulation scenarios include ITS with 12 datapoints, a **level-change of 0, slope-change of 0**, slope-change heterogeneity of 0, and fixed levels of autocorrelation.

DL, DerSimonian and Laird. dps, datapoints. HKSJ, Hartung-Knapp / Sidik-Jonkman. ITS, interrupted time series. PW, Prais-Winsten. REML, restricted maximum likelihood. WT, Wald-type.

Appendix Figure S173. Plots of the empirical standard error of the immediate level-change (y-axis) when the ITS are analysed with PW (A) and REML (B) for the fixed effect (red circles), DL+WT (green diamonds) and DL+HKSJ (brown crosses), REML+WT (purple diamonds) and REML+HKSJ (blue crosses) meta-analysis methods versus autocorrelation (x-axis). Plots are presented separately by combinations of the number of included studies (rows) and level-change heterogeneity (columns). Simulation scenarios include ITS with 12 datapoints, a **level-change of 0, slope-change of 0.1**, slope-change heterogeneity of 0, and fixed levels of autocorrelation.

DL, DerSimonian and Laird. dps, datapoints. HKSJ, Hartung-Knapp / Sidik-Jonkman. ITS, interrupted time series. PW, Prais-Winsten. REML, restricted maximum likelihood. WT, Wald-type.

Appendix Figure S174. Plots of the empirical standard error of the immediate level-change (y-axis) when the ITS are analysed with PW (A) and REML (B) for the fixed effect (red circles), DL+WT (green diamonds) and DL+HKSJ (brown crosses), REML+WT (purple diamonds) and REML+HKSJ (blue crosses) meta-analysis methods versus autocorrelation (x-axis). Plots are presented separately by combinations of the number of included studies (rows) and level-change heterogeneity (columns). Simulation scenarios include ITS with 12 datapoints, a **level-change of 1, slope-change of 0**, slope-change heterogeneity of 0, and fixed levels of autocorrelation.

DL, DerSimonian and Laird. dps, datapoints. HKSJ, Hartung-Knapp / Sidik-Jonkman. ITS, interrupted time series. PW, Prais-Winsten. REML, restricted maximum likelihood. WT, Wald-type.

Appendix Figure S175. Plots of the empirical standard error of slope-change (y-axis) when the ITS are analysed with PW (A) and REML (B) for the fixed effect (red circles), DL+WT (green diamonds) and DL+HKSJ (brown crosses), REML+WT (purple diamonds) and REML+HKSJ (blue crosses) meta-analysis methods versus autocorrelation (x-axis). Plots are presented separately by combinations of the number of included studies (rows) and level-change heterogeneity (columns). S Simulation scenarios include ITS with 12 datapoints, a **level-change of 0, slope-change of 0**, slope-change heterogeneity of 0, and fixed levels of autocorrelation.

DL, DerSimonian and Laird. dps, datapoints. HKSJ, Hartung-Knapp / Sidik-Jonkman. ITS, interrupted time series. PW, Prais-Winsten. REML, restricted maximum likelihood. WT, Wald-type.

Appendix Figure S176. Plots of the empirical standard error of slope-change (y-axis) when the ITS are analysed with PW (A) and REML (B) for the fixed effect (red circles), DL+WT (green diamonds) and DL+HKSJ (brown crosses), REML+WT (purple diamonds) and REML+HKSJ (blue crosses) meta-analysis methods versus autocorrelation (x-axis). Plots are presented separately by combinations of the number of included studies (rows) and level-change heterogeneity (columns). Simulation scenarios include ITS with 12 datapoints, a **level-change of 0, slope-change of 0.1**, slope-change heterogeneity of 0, and fixed levels of autocorrelation.

DL, DerSimonian and Laird. dps, datapoints. HKSJ, Hartung-Knapp / Sidik-Jonkman. ITS, interrupted time series. PW, Prais-Winsten. REML, restricted maximum likelihood. WT, Wald-type.

Appendix Figure S177. Plots of the empirical standard error of slope-change (y-axis) when the ITS are analysed with PW (A) and REML (B) for the fixed effect (red circles), DL+WT (green diamonds) and DL+HKSJ (brown crosses), REML+WT (purple diamonds) and REML+HKSJ (blue crosses) meta-analysis methods versus autocorrelation (x-axis). Plots are presented separately by combinations of the number of included studies (rows) and level-change heterogeneity (columns). Simulation scenarios include ITS with 12 datapoints, a **level-change of 1, slope-change of 0**, slope-change heterogeneity of 0, and fixed levels of autocorrelation.

DL, DerSimonian and Laird. dps, datapoints. HKSJ, Hartung-Knapp / Sidik-Jonkman. ITS, interrupted time series. PW, Prais-Winsten. REML, restricted maximum likelihood. WT, Wald-type.

##### 4.9.4 Ratio of model based standard errors to empirical standard errors

Appendix Figure S178. Plots of the ratio of model based standard error (modSE) to the empirical standard error (empSE) of the immediate level-change (y-axis) when the ITS are analysed with PW (A) and REML (B) for the fixed effect (red circles), DL+WT (green diamonds) and DL+HKSJ (brown crosses), REML+WT (purple diamonds) and REML+HKSJ (blue crosses) meta-analysis methods versus autocorrelation (x-axis). Plots are presented separately by combinations of the number of included studies (rows) and level-change heterogeneity (columns). Simulation scenarios include ITS with 12 datapoints, a **level-change of 0, slope-change of 0**, slope-change heterogeneity of 0, and fixed levels of autocorrelation.

DL, DerSimonian and Laird. dps, datapoints. HKSJ, Hartung-Knapp / Sidik-Jonkman. ITS, interrupted time series. PW, Prais-Winsten. REML, restricted maximum likelihood. WT, Wald-type.

Appendix Figure S179. Plots of the ratio of model based standard error (modSE) to the empirical standard error (empSE) of the immediate level-change (y-axis) when the ITS are analysed with PW (A) and REML (B) for the fixed effect (red circles), DL+WT (green diamonds) and DL+HKSJ (brown crosses), REML+WT (purple diamonds) and REML+HKSJ (blue crosses) meta-analysis methods versus autocorrelation (x-axis). Plots are presented separately by combinations of the number of included studies (rows) and level-change heterogeneity (columns). Simulation scenarios include ITS with 12 datapoints, a **level-change of 0, slope-change of 0.1**, slope-change heterogeneity of 0, and fixed levels of autocorrelation.

DL, DerSimonian and Laird. dps, datapoints. HKSJ, Hartung-Knapp / Sidik-Jonkman. ITS, interrupted time series. PW, Prais-Winsten. REML, restricted maximum likelihood. WT, Wald-type.

Appendix Figure S180. Plots of the ratio of model based standard error (modSE) to the empirical standard error (empSE) of the immediate level-change (y-axis) when the ITS are analysed with PW (A) and REML (B) for the fixed effect (red circles), DL+WT (green diamonds) and DL+HKSJ (brown crosses), REML+WT (purple diamonds) and REML+HKSJ (blue crosses) meta-analysis methods versus autocorrelation (x-axis). Plots are presented separately by combinations of the number of included studies (rows) and level-change heterogeneity (columns). Simulation scenarios include ITS with 12 datapoints, a **level-change of 1, slope-change of 0**, slope-change heterogeneity of 0, and fixed levels of autocorrelation.

DL, DerSimonian and Laird. dps, datapoints. HKSJ, Hartung-Knapp / Sidik-Jonkman. ITS, interrupted time series. PW, Prais-Winsten. REML, restricted maximum likelihood. WT, Wald-type.

Appendix Figure S181. Plots of the ratio of model based standard error (modSE) to the empirical standard error (empSE) of slope-change (y-axis) when the ITS are analysed with PW (A) and REML (B) for the fixed effect (red circles), DL+WT (green diamonds) and DL+HKSJ (brown crosses), REML+WT (purple diamonds) and REML+HKSJ (blue crosses) meta-analysis methods versus autocorrelation (x-axis). Plots are presented separately by combinations of the number of included studies (rows) and level-change heterogeneity (columns). Simulation scenarios include ITS with 12 datapoints, a **level-change of 0, slope-change of 0**, slope-change heterogeneity of 0, and fixed levels of autocorrelation.

DL, DerSimonian and Laird. dps, datapoints. HKSJ, Hartung-Knapp / Sidik-Jonkman. ITS, interrupted time series. PW, Prais-Winsten. REML, restricted maximum likelihood. WT, Wald-type.

Appendix Figure S182. Plots of the ratio of model based standard error (modSE) to the empirical standard error (empSE) of slope-change (y-axis) when the ITS are analysed with PW (A) and REML (B) for the fixed effect (red circles), DL+WT (green diamonds) and DL+HKSJ (brown crosses), REML+WT (purple diamonds) and REML+HKSJ (blue crosses) meta-analysis methods versus autocorrelation (x-axis). Plots are presented separately by combinations of the number of included studies (rows) and level-change heterogeneity (columns). Simulation scenarios include ITS with 12 datapoints, a **level-change of 0, slope-change of 0.1**, slope-change heterogeneity of 0, and fixed levels of autocorrelation.

DL, DerSimonian and Laird. dps, datapoints. HKSJ, Hartung-Knapp / Sidik-Jonkman. ITS, interrupted time series. PW, Prais-Winsten. REML, restricted maximum likelihood. WT, Wald-type.

Appendix Figure S183. Plots of the ratio of model based standard error (modSE) to the empirical standard error (empSE) of slope-change (y-axis) when the ITS are analysed with PW (A) and REML (B) for the fixed effect (red circles), DL+WT (green diamonds) and DL+HKSJ (brown crosses), REML+WT (purple diamonds) and REML+HKSJ (blue crosses) meta-analysis methods versus autocorrelation (x-axis). Plots are presented separately by combinations of the number of included studies (rows) and level-change heterogeneity (columns). Simulation scenarios include ITS with 12 datapoints, a **level-change of 1, slope-change of 0**, slope-change heterogeneity of 0, and fixed levels of autocorrelation.

DL, DerSimonian and Laird. dps, datapoints. HKSJ, Hartung-Knapp / Sidik-Jonkman. ITS, interrupted time series. PW, Prais-Winsten. REML, restricted maximum likelihood. WT, Wald-type.

##### 4.9.5 Statistical power

Appendix Figure S184. Plots of the statistical power (the percentage of simulations that have a 95% confidence interval that did not include zero – only scenarios with a confidence interval coverage of greater than 90% are plotted) of the immediate level-change (y-axis) when the ITS are analysed with PW (A) and REML (B) for the fixed effect (red circles), DL+WT (green diamonds) and DL+HKSJ (brown crosses), REML+WT (purple diamonds) and REML+HKSJ (blue crosses) meta-analysis methods versus autocorrelation (x-axis). Plots are presented separately by combinations of the number of included studies (rows) and level-change heterogeneity (columns). Simulation scenarios include ITS with 12 datapoints, a **level-change of 0, slope-change of 0**, slope-change heterogeneity of 0, and fixed levels of autocorrelation.

DL, DerSimonian and Laird. dps, datapoints. HKSJ, Hartung-Knapp / Sidik-Jonkman. ITS, interrupted time series. PW, Prais-Winsten. REML, restricted maximum likelihood. WT, Wald-type.

Appendix Figure S185. Plots of the statistical power (the percentage of simulations that have a 95% confidence interval that did not include zero – only scenarios with a confidence interval coverage of greater than 90% are plotted) of the immediate level-change (y-axis) when the ITS are analysed with PW (A) and REML (B) for the fixed effect (red circles), DL+WT (green diamonds) and DL+HKSJ (brown crosses), REML+WT (purple diamonds) and REML+HKSJ (blue crosses) meta-analysis methods versus autocorrelation (x-axis). Plots are presented separately by combinations of the number of included studies (rows) and level-change heterogeneity (columns). Simulation scenarios include ITS with 12 datapoints, a **level-change of 0, slope-change of 0.1**, slope-change heterogeneity of 0, and fixed levels of autocorrelation.

DL, DerSimonian and Laird. dps, datapoints. HKSJ, Hartung-Knapp / Sidik-Jonkman. ITS, interrupted time series. PW, Prais-Winsten. REML, restricted maximum likelihood. WT, Wald-type.

Appendix Figure S186. Plots of the statistical power (the percentage of simulations that have a 95% confidence interval that did not include zero – only scenarios with a confidence interval coverage of greater than 90% are plotted) of the immediate level-change (y-axis) when the ITS are analysed with PW (A) and REML (B) for the fixed effect (red circles), DL+WT (green diamonds) and DL+HKSJ (brown crosses), REML+WT (purple diamonds) and REML+HKSJ (blue crosses) meta-analysis methods versus autocorrelation (x-axis). Plots are presented separately by combinations of the number of included studies (rows) and level-change heterogeneity (columns). Simulation scenarios include ITS with 12 datapoints, a **level-change of 1, slope-change of 0**, slope-change heterogeneity of 0, and fixed levels of autocorrelation.

DL, DerSimonian and Laird. dps, datapoints. HKSJ, Hartung-Knapp / Sidik-Jonkman. ITS, interrupted time series. PW, Prais-Winsten. REML, restricted maximum likelihood. WT, Wald-type.

Appendix Figure S187. Plots of the statistical power (the percentage of simulations that have a 95% confidence interval that did not include zero – only scenarios with a confidence interval coverage of greater than 90% are plotted) of slope-change (y-axis) when the ITS are analysed with PW (A) and REML (B) for the fixed effect (red circles), DL+WT (green diamonds) and DL+HKSJ (brown crosses), REML+WT (purple diamonds) and REML+HKSJ (blue crosses) meta-analysis methods versus autocorrelation (x-axis). Plots are presented separately by combinations of the number of included studies (rows) and level-change heterogeneity (columns). Simulation scenarios include ITS with 12 datapoints, a **level-change of 0, slope-change of 0**, slope-change heterogeneity of 0, and fixed levels of autocorrelation.

DL, DerSimonian and Laird. dps, datapoints. HKSJ, Hartung-Knapp / Sidik-Jonkman. ITS, interrupted time series. PW, Prais-Winsten. REML, restricted maximum likelihood. WT, Wald-type.

Appendix Figure S189. Plots of the statistical power (the percentage of simulations that have a 95% confidence interval that did not include zero – only scenarios with a confidence interval coverage of greater than 90% are plotted) of slope-change (y-axis) when the ITS are analysed with PW (A) and REML (B) for the fixed effect (red circles), DL+WT (green diamonds) and DL+HKSJ (brown crosses), REML+WT (purple diamonds) and REML+HKSJ (blue crosses) meta-analysis methods versus autocorrelation (x-axis). Plots are presented separately by combinations of the number of included studies (rows) and level-change heterogeneity (columns). Simulation scenarios include ITS with 12 datapoints, a **level-change of 0, slope-change of 0.1**, slope-change heterogeneity of 0, and fixed levels of autocorrelation.

DL, DerSimonian and Laird. dps, datapoints. HKSJ, Hartung-Knapp / Sidik-Jonkman. ITS, interrupted time series. PW, Prais-Winsten. REML, restricted maximum likelihood. WT, Wald-type.

Appendix Figure S190. Plots of the statistical power (the percentage of simulations that have a 95% confidence interval that did not include zero – only scenarios with a confidence interval coverage of greater than 90% are plotted) of slope-change (y-axis) when the ITS are analysed with PW (A) and REML (B) for the fixed effect (red circles), DL+WT (green diamonds) and DL+HKSJ (brown crosses), REML+WT (purple diamonds) and REML+HKSJ (blue crosses) meta-analysis methods versus autocorrelation (x-axis). Plots are presented separately by combinations of the number of included studies (rows) and level-change heterogeneity (columns). Simulation scenarios include ITS with 12 datapoints, a **level-change of 1, slope-change of 0**, slope-change heterogeneity of 0, and fixed levels of autocorrelation.

DL, DerSimonian and Laird. dps, datapoints. HKSJ, Hartung-Knapp / Sidik-Jonkman. ITS, interrupted time series. PW, Prais-Winsten. REML, restricted maximum likelihood. WT, Wald-type.
